# Assessing Acute-Care 30-Day Mortality Prediction Using Clinical Features in CHoRUS Clinical Care for AI and MIMIC-IV

**DOI:** 10.64898/2026.08.05.26359541

**Authors:** Ria Chaudhry, Zhe Sage Chen

**Author notes:** **Corresponding Author:** Zhe Sage Chen, PhD, One Park Avenue, Rm 8-226, New York, NY 10016, USA.

## Abstract

**Background:** Mortality prediction models often combine early electronic health record data, but the relative prognostic value of baseline vulnerability, physiological severity, treatment exposure, and procedure burden remains unclear.

**Objective:** To compare routinely available first-24-hour clinical domains for visit-level 30-day mortality prediction and assess whether domain-level patterns replicated in MIMIC-IV.

**Methods:** We used CHoRUS (version 1), an OMOP-formatted acute-care dataset, with independent domain-level replication in MIMIC-IV (version 3.1). CHoRUS included 22,098 visits among 5,892 unique patients, with 1,004 30-day mortality events and 4.5% mortality prevalence. MIMIC-IV included 23,000 acute-care visits among 10,006 unique patients, with 819 events and 3.6% prevalence. Across both datasets, 45,098 visits and 15,898 unique patients were analyzed. Predictors were restricted to the first 24 hours after visit start. Performance was evaluated using AUPRC, AUROC, Brier score, calibration, sensitivity at 90% specificity, highest-risk 10% analyses, decision-curve analysis, and SHAP summaries. Because 30-day mortality was infrequent, the classification task was class-imbalanced. Accordingly, AUPRC was interpreted relative to the prevalence-based no-skill baseline, rather than as an absolute measure alone.

**Results and Conclusion:** Physiological severity produced the largest improvement beyond baseline in CHoRUS, with median AUPRC 0.38 and median AUROC 0.86, and showed the same primary domain-level pattern in MIMIC-IV. Treatment exposure and procedure burden provided smaller gains. In CHoRUS, the best pairwise model combined baseline, physiological severity, and procedure burden features, with median AUPRC 0.41; the all-domain model was slightly lower, with median AUPRC 0.40 and median AUROC 0.86. In MIMIC-IV, the all-domain model had the highest median AUPRC, 0.25, only modestly above the best pairwise model. First-24-hour physiological severity features therefore provided the most consistent prognostic information across datasets, supporting parsimonious, clinically interpretable acute-care risk models centered on high-quality early physiological data.

## 1 Introduction

During the first 24 hours of acute care, clinicians must rapidly assess severity to guide monitoring, escalation, and resource use. Electronic health records (EHRs) capture vital signs, laboratory testing, medications, procedures, utilization, and comorbidity during this period, but the relative value of these early domains for mortality risk stratification remains unclear. Short-term mortality prediction is clinically meaningful because it may help identify encounters requiring closer review or escalation [1].

Traditional severity scores such as APACHE, SAPS, and SOFA show the importance of early physiological measurements and organ dysfunction for mortality assessment [2, 3, 4]. Yet health systems need to know not only whether risk can be predicted, but which early structured data domains are most informative. Prior EHR models often use large combined feature sets, limiting interpretability and making it difficult to separate baseline vulnerability, physiological severity, treatment exposure, and procedure burden. Earlier studies evaluated richer claims, registry, laboratory, or EHR data [5, 6, 7, 8], and other explainable machine-learning studies grouped mortality predictors into domains [9, 10, 11, 12]. However, few studies compare multiple early clinical domains within a reproducible acute-care framework. We addressed this gap using CHoRUS, an OMOP-CDM acute-care dataset [13, 14], with MIMIC-IV used for independent domain-level replication. We quantified the predictive value of four domains: baseline vulnerability, physiological severity, treatment exposure, and procedure burden. We then evaluated pairwise combinations and all-domain models to determine whether broader feature aggregation improved 30-day mortality prediction. To our best knowledge, our effort represents the first systematic investigation on this clinically important question: which routinely available first-24-hour clinical domain features provide the greatest benefit for encounter-level Acute-care 30-day mortality prediction.

## 2 Methods

### 2.1 Study Design, Data Sources and Ethics

This retrospective prognostic study evaluated the value of first-24-hour clinical domains for acute-care 30-day mortality prediction and followed TRIPOD+AI reporting principles [15]. We used deidentified secondary data from CHoRUS (version 1) and MIMIC-IV (version 3.1). CHoRUS is an NIH Bridge2AI controlled-access clinical data resource [16]. MIMIC-IV is a deidentified hospital dataset available through PhysioNet after required training and data-use agreement completion [17]. Because the analysis used deidentified secondary data, involved no patient contact, and did not attempt reidentification, additional informed consent was not obtained.

### 2.2 Prediction Time Point, Outcome and Study Population

The prediction anchor was 24 hours after visit start. Predictors were restricted to information available during the first 24 hours. The outcome was death at or after 24 hours and within 30 days after visit start. The 24-hour prediction window was selected because it represents an early acute-care decision point at which initial clinical information is available but meaningful opportunities for risk stratification and intervention remain, it is a commonly used acute-care outcome window [18, 19]. The cohort construction was aligned with the prediction time point and outcome definitions to maintain the clinical utility of the selected cohort.

The CHoRUS acute-care cohort included hospital, observation, inpatient, emergency department, and emergency-to-inpatient visits. Temporally invalid visits were removed, visits with death before the 24-hour landmark were excluded, and visits shorter than 24 hours were retained. The final CHoRUS cohort included 22,098 visits among 5,892 unique patients, with 1,004 30-day mortality events and 4.5% prevalence.

The MIMIC-IV replication cohort included observation, emergency, and urgent acute-care visits. Because the full eligible cohort exceeded local compute constraints, we used a reproducible patient-level stratified subsample of 23,000 visits among 10,006 unique patients. This cohort included 819 30-day mortality events and 3.6% prevalence. Across both datasets, the analysis included 45,098 visits and 15,898 unique patients.

Because 30-day mortality was uncommon, both datasets produced class-imbalanced binary classification tasks. Since, our primary performance metric, AUPRC depends strongly on positive-class prevalence, we interpreted AUPRC relative to the no-skill baseline, approximately equal to the mortality prevalence in each dataset.

### 2.3 Feature Construction

Baseline vulnerability included demographic, visit-type, prior-utilization, and comorbidity information available at or before the index visit. Prior utilization included prior visit count, has prior visit, and prior acute visit count. Comorbidity was summarized with the Charlson Comorbidity Index. The MIMIC-IV implementation used ICD-9 and ICD-10 mappings. All features were harmonized across the datasets. Categorical baseline variables (race, gender, ethnicity and visit-type) were one-hot encoded [20]. CHoRUS contained 21 final baseline features. MIMIC-IV contained 25 final baseline features. These baseline features were chosen to represent information available to clinicians early in the visit before any physiological measurements, procedures and/or treatments have been conducted. Additional baseline encoding details and category lists are provided in Supplementary eMethods 3 and eTable 1.

Three early clinical domains were evaluated: physiological severity, treatment exposure, and procedure burden. Each of these were selected to represent different aspects of an acute-care clinical evaluation. Physiological severity, defined as measures of physiology collected during the visit, used OMOP measurement-derived features in CHoRUS and laboratory-event features in MIMIC-IV. The following summary features were calculated: maximum, minimum, mean, standard deviation, count, and missingness. Treatment exposure, defined as drug based interventions during the visit, used medication indicators, counts, and aggregate exposure features. Procedure burden, defined as procedural interventions conducted during the study ranging from EEG procedures to surgical interventions, used procedure indicators, counts, and three aggregate features: `unique_procedure_count_24h`, `procedure_count_total_24h`, and `any_procedure_24h`. Detailed breakdown of all features used and summary features can be found in supplementary eMethods 3 and eTables 2-4.

### 2.4 Feature Matrices and Clinical Domain Staging

Baseline features were retained in every matrix. We evaluated baseline alone; three standalone matrices adding physiological severity, treatment exposure, or procedure burden; three pairwise complementary matrices; and one all-domain matrix. Overfitting is a common machine learning problem that occurs when using large feature matrices [21]. To reduce the risk of overfitting the following strategy was used.

For each clinical domain the 50 most frequently occurring variables were retained (for the CHoRUS dataset, the physiological severity domain contained 3 measures that were unusable resulting in 47 final physiological measurement variables being selected). Further, for each clinical domain, the 21 most frequently occurring candidate features were selected within each training fold and applied unchanged to the held-out fold. The same fold-specific 21 features for a domain were reused across every matrix containing that domain. CHoRUS standalone, complementary, and all-domain matrices contained 42, 63, and 84 final features, respectively. MIMIC-IV matrices contained 46, 67, and 88 final features. This allowed for the reduction of the feature matrix size, further, cross validation, regularized logistic regression, and a constrained tree based algorithm helped reduce the risk of overfitting. This method is clinically aligned as well, because it allows for the most abundant information to be evaluated. Detailed feature definitions and fold-level selected features are provided in Supplementary eMethods 3, eTables 1–4, and eTables 33–34.

### 2.5 Performance Metrics

Since 30-day mortality was an imbalanced binary outcome, we prioritized threshold-independent discrimination metrics, especially area under the precision-recall curve (AUPRC) and area under the receiver operating characteristic curve (AUROC). AUPRC and AUROC were chosen as the primary metrics because, clinicians need to know whether a model can rank patients so that those at highest risk are prioritized for closer monitoring, escalation, or review. AUROC summarizes how well the model separates patients who die from those who survive across possible thresholds. AUPRC is especially important when mortality is uncommon because it focuses on performance among the patients flagged as high risk, summarizing the tradeoff between capturing deaths and avoiding excessive false alerts [22, 23].

We also reported threshold-based classification metrics using each algorithm’s default class prediction rule, including accuracy, precision, recall/sensitivity, specificity, F1 score, calibration curves, decision curves, sensitivity at 90% specificity and top 10% risk analysis. These metrics show the practical consequences of using a model at a decision threshold, including how many high-risk patients are correctly identified, how many are missed, and how many false alerts clinicians would need to manage.

### 2.6 Model Development and Internal Validation

We evaluated logistic regression, random forest, gradient boosting, and LightGBM using the same modeling approach across feature matrices. Five-fold patient-level grouped cross-validation prevented patient overlap between training and validation folds [24]. The best algorithm for each matrix was selected by highest mean cross-validated AUPRC, with mean AUROC as the tie-breaker. Reported results are held-out fold medians across the five patient-grouped folds for the selected algorithm. A fixed-algorithm sensitivity analysis using gradient boosting for CHoRUS and logistic regression for MIMIC-IV was conducted. Algorithm-specific results for every feature matrix are reported in Supplementary eTables 7 and 8.

## 3 Results

### 3.1 Cohort and Patient Characteristics

CHoRUS included 22,098 visits among 5,892 unique patients, with 1,004 30-day mortality events and 4.5% prevalence. MIMIC-IV included 23,000 visits among 10,006 unique patients, with 819 events and 3.6% prevalence. Table 1 reports demographic characteristics, and Figure 1 summarizes cohort and feature-matrix construction.

**Table 1:**
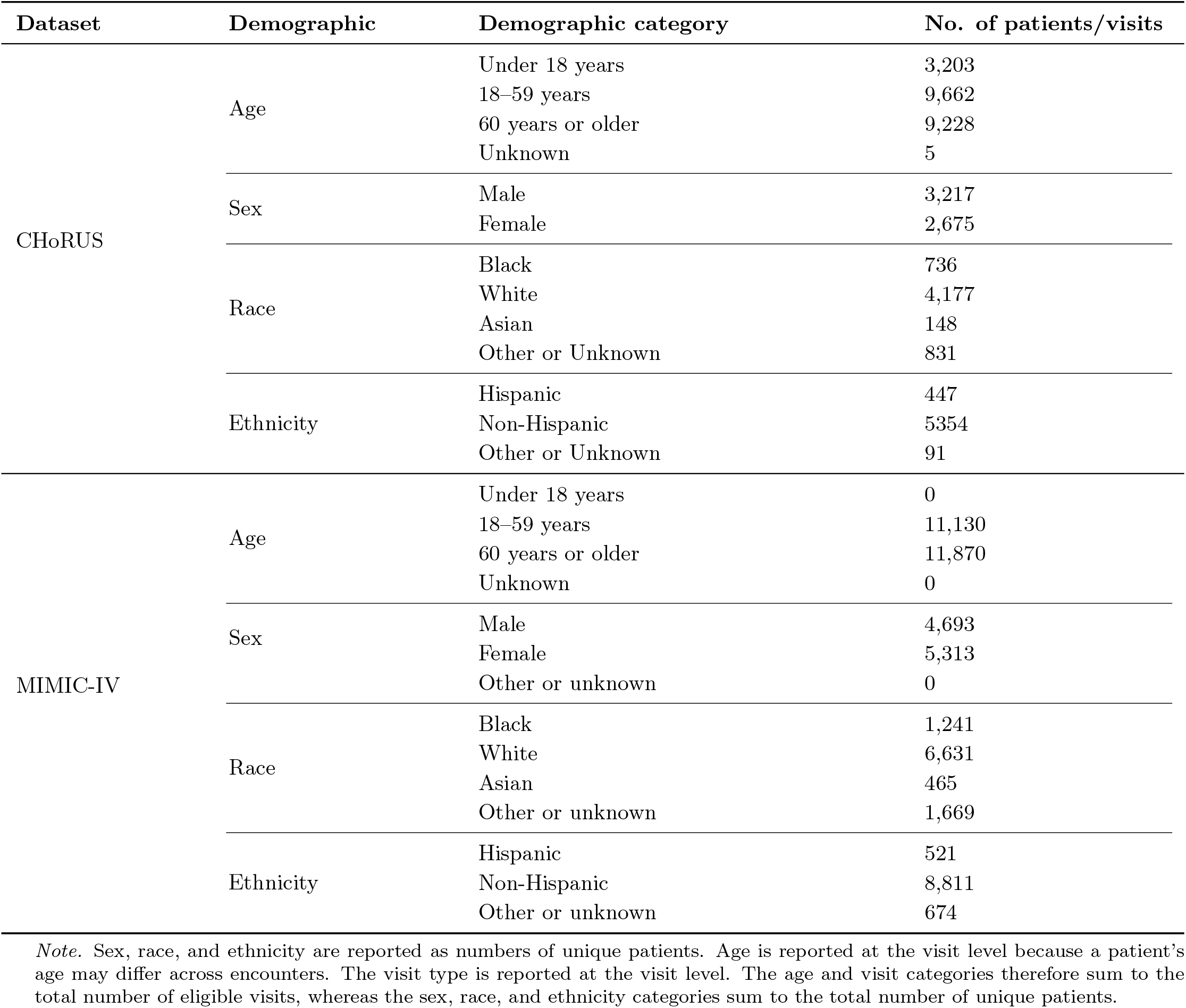
Cohort demographic characteristics.

**Table 2:**
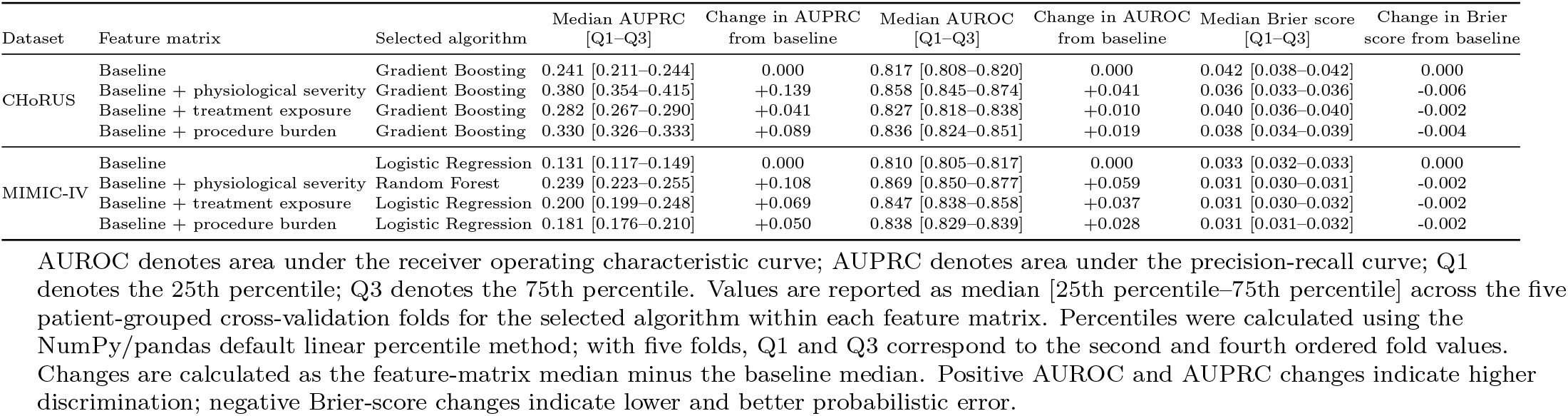
Standalone clinical-domain performance in CHoRUS and MIMIC-IV using selected algorithms.

**Figure 1:**
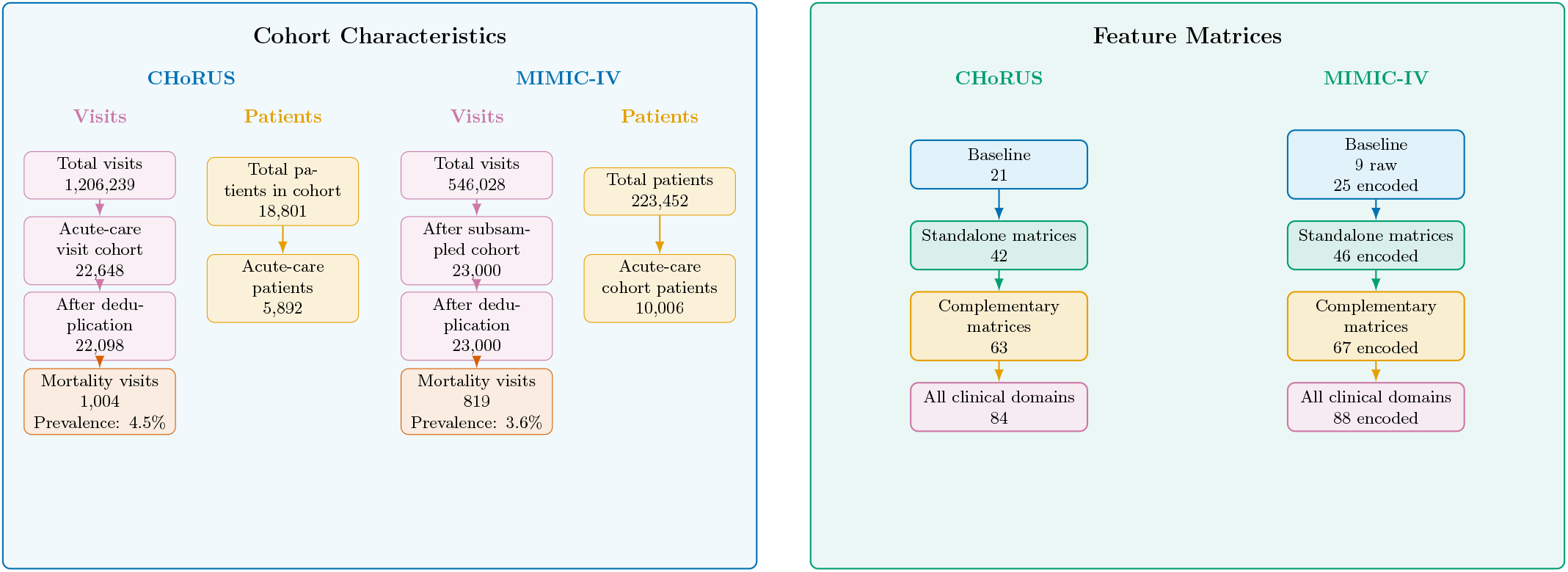
Cohort characteristics and feature matrix construction for CHoRUS and MIMIC-IV. Four-panel calibration curves using saved calibration-coordinate outputs. Panel A shows CHoRUS

**Figure 2:**
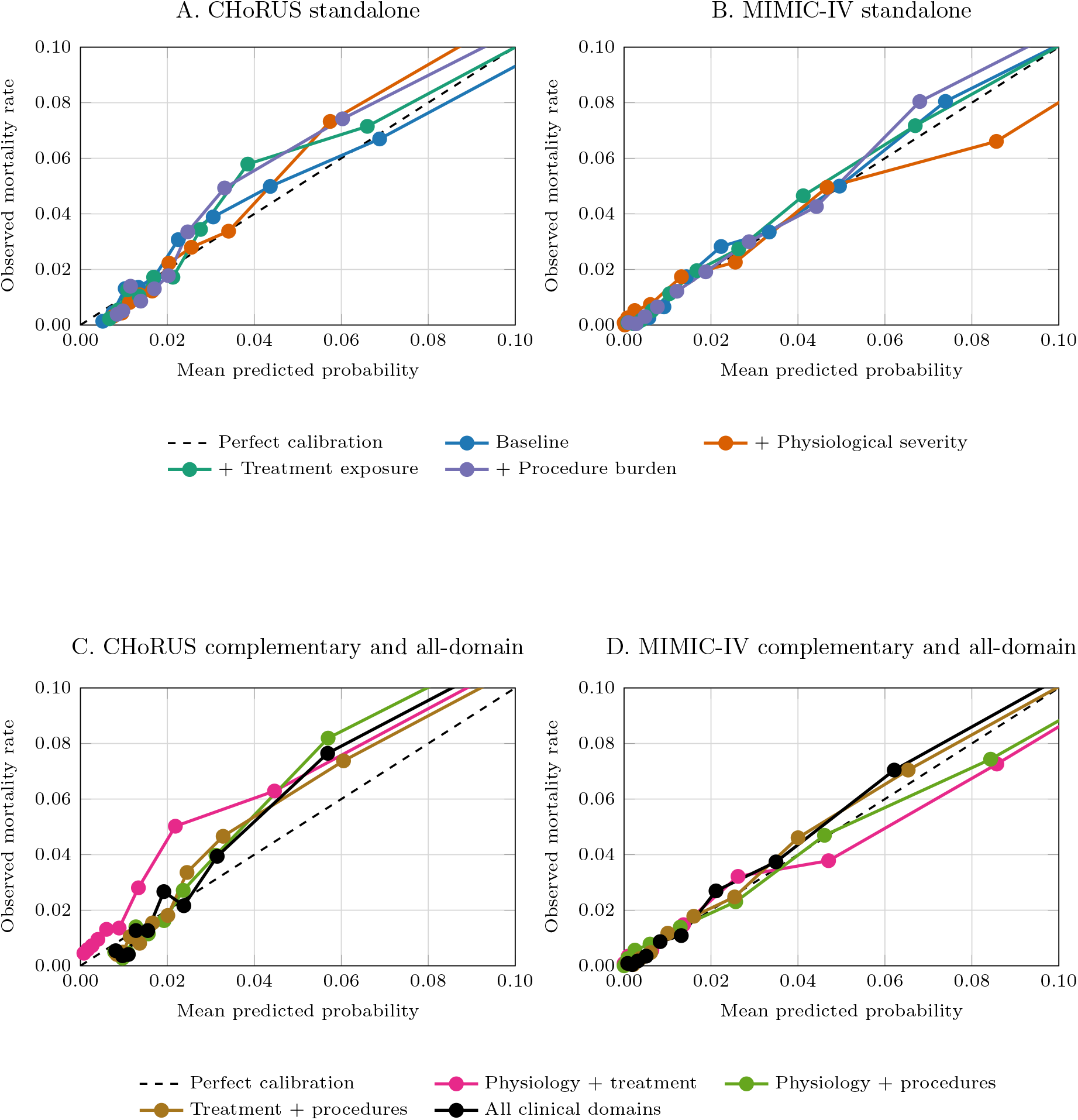
standalone feature matrices; Panel B shows MIMIC-IV standalone feature matrices; Panel C shows CHoRUS complementary and all-domain feature matrices; and Panel D shows MIMIC-IV complementary and all-domain feature matrices. The dashed diagonal represents perfect calibration. Both axes are restricted to probabilities from 0 to 0.10 to emphasize differences in calibration across the lower nine risk groups. The highest-risk group, which had mean predicted and observed probabilities above 0.10, is not displayed in this zoomed view.

### 3.2 Standalone Feature Matrices

Baseline features provided meaningful discrimination in both datasets. In CHoRUS, baseline achieved median AUPRC 0.24 (0.211-0.244), median AUROC 0.82 (0.808–0.820), and median Brier score 0.042 (0.038–0.042). In MIMIC-IV, baseline achieved median AUPRC 0.13 (0.117–0.149), median AUROC 0.81 (0.805–0.817), and median Brier score 0.033 (0.032–0.033).

Physiological severity produced the largest standalone gain beyond baseline in both datasets. In CHoRUS, it increased median AUPRC from 0.24 to 0.38, increased median AUROC from 0.82 to 0.86, and reduced Brier score from 0.042 to 0.036. Procedure burden was the best secondary standalone domain in CHoRUS, with median AUPRC 0.33, median AUROC 0.84, and median Brier score 0.038; treatment exposure produced smaller gains, with median AUPRC 0.28, median AUROC 0.83, and median Brier score 0.040. In MIMIC-IV, physiological severity again provided the largest gain, with AUPRC +0.108, AUROC +0.059, and Brier-score change -0.002. Treatment exposure was the strongest secondary domain, with AUPRC +0.069, AUROC +0.037, and Brier-score change -0.002, while procedure burden produced smaller gains.

Unless otherwise specified, reported performance values are five-fold cross-validation medians from the best-performing algorithm selected separately for each feature matrix; selected algorithms and complete algorithm-specific results are provided in the Supplement. Across algorithms, physiological severity provided the most consistent improvement beyond baseline. In the fixed-algorithm sensitivity analysis, the standalone clinical-domain ranking was unchanged in both datasets, supporting that the primary findings were not attributable to feature-matrix-specific algorithm selection (eTables 7–8).

### 3.3 Complementary Feature Matrices

Pairwise and all-domain models assessed whether clinical domains carried complementary information. Across both datasets, the best-performing pairwise models included physiological severity. In CHoRUS, baseline + physiological severity + procedure burden performed best, with median AUPRC 0.41, median AUROC 0.86, and median Brier score 0.035. Relative to baseline, this corresponded to AUPRC +0.163, AUROC +0.038, and Brier-score change -0.007; relative to baseline + physiological severity, the changes were AUPRC +0.024, AUROC -0.003, and Brier-score change -0.001. The CHoRUS all-domain model achieved median AUPRC 0.40, median AUROC 0.86, and median Brier score 0.035, slightly below the best pairwise model by AUPRC (-0.006) with the same displayed AUROC and Brier score.

In MIMIC-IV, the strongest pairwise model was metric-dependent: baseline + physiological severity + procedure burden had the highest pairwise median AUPRC (0.236), whereas baseline + physiological severity + treatment exposure had the highest pairwise median AUROC (0.872). The all-domain model had median AUPRC 0.25, median AUROC 0.87, and median Brier score 0.030, corresponding to AUPRC +0.011, AUROC +0.003, and no displayed Brier-score difference relative to the best AUPRC-based pairwise matrix. Figure 4 shows discrimination curves across the eight feature matrices.

**Figure 3:**
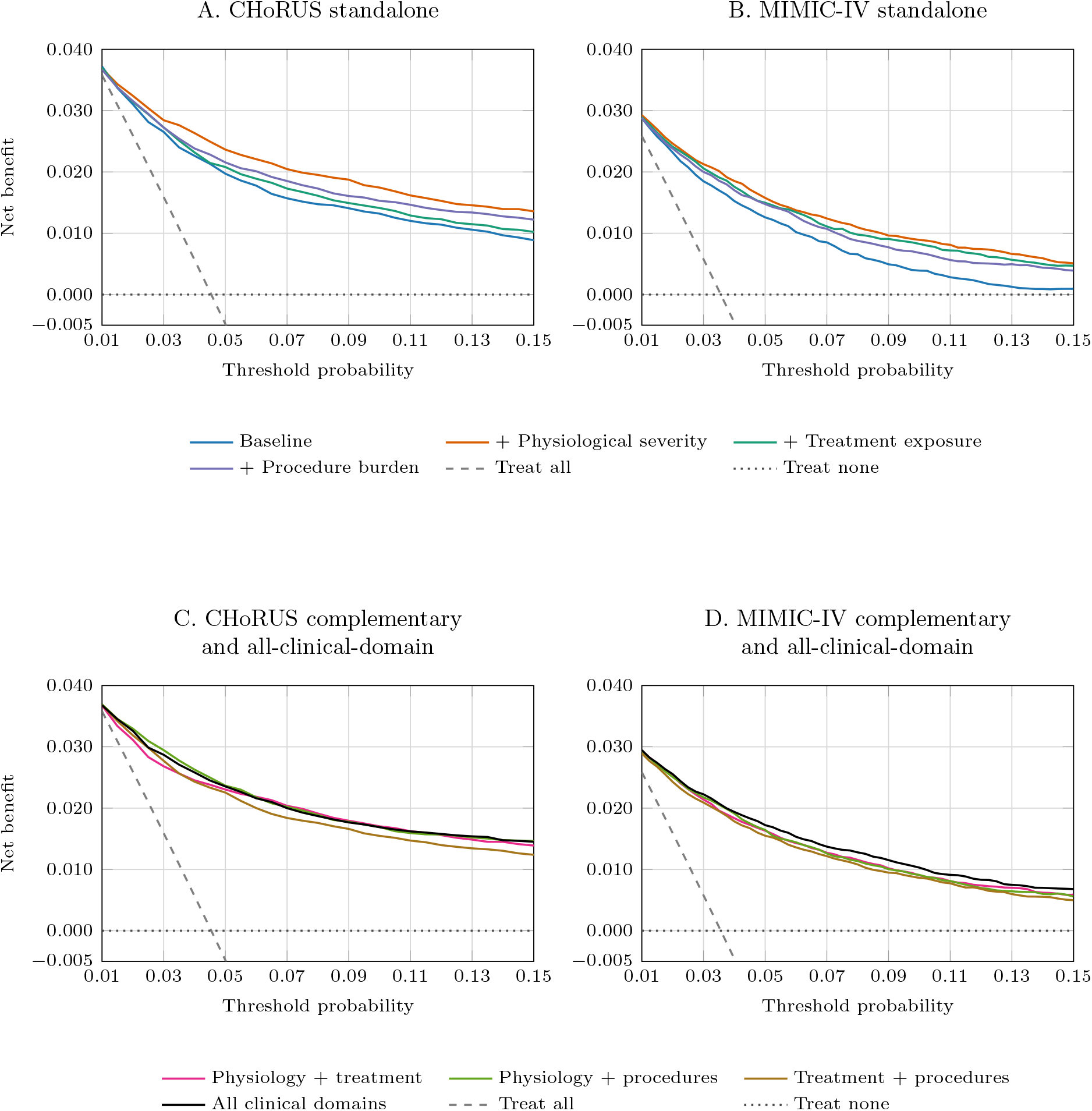
Four-panel decision-curve analysis using saved decision-curve coordinate outputs. Panel A shows CHoRUS standalone feature matrices; Panel B shows MIMIC-IV standalone feature matrices; Panel C shows CHoRUS complementary and all-clinical-domain feature matrices; Panel D shows MIMIC-IV complementary and all-clinical-domain feature matrices. Treat-all and treat-none reference strategies are shown in every panel: treat-none represents a strategy of flagging no encounters as high risk and has zero net benefit, whereas treat-all represents a strategy of flagging all encounters as high risk, with net benefit varying by threshold probability. The net-benefit axis is restricted to −0.005 to 0.040 to make differences among the model curves more visible; treat-all values below the displayed range are clipped.

**Figure 4:**
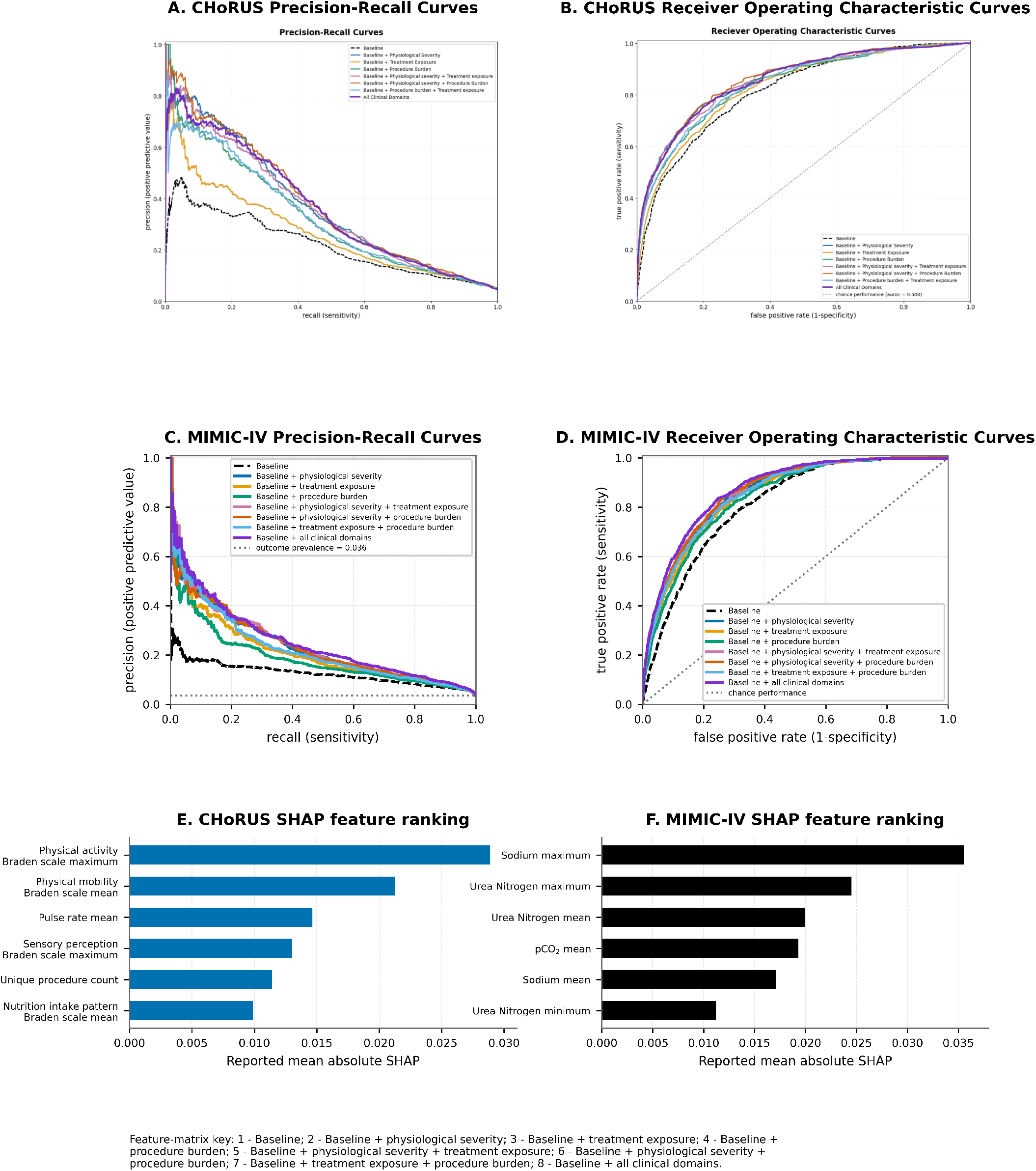
Discrimination and feature-importance results in CHoRUS and MIMIC-IV. Precision-recall and receiver operating characteristic curves compare the eight prespecified feature matrices using pooled out-of-fold predictions in CHoRUS (A and B) and the independently trained MIMIC-IV replication analysis (C and D). The horizontal reference line in each precision-recall panel represents outcome prevalence, and the diagonal reference line in each receiver operating characteristic panel represents chance discrimination. Panels E and F show the six highest-ranked features by reported mean absolute SHAP value in CHoRUS and MIMIC-IV, respectively. Identical colors denote the same feature matrices across datasets. AUPRC indicates area under the precision-recall curve; AUROC, area under the receiver operating characteristic curve; and SHAP, Shapley additive explanations.

### 3.4 Calibration

Within the displayed 0 to 0.10 predicted-probability range, most calibration points remained near the 45-degree reference line, although local underprediction and overprediction were present. In CHoRUS, baseline + physiological severity underestimated observed mortality in some intermediate risk groups, including mean predicted probability 0.057 with observed mortality 0.073. Baseline + physiological severity + treatment exposure showed more consistent underprediction in displayed CHoRUS risk groups, including predicted and observed probabilities of 0.045 and 0.063. In MIMIC-IV, deviations were generally smaller; for example, the baseline model produced predicted and observed probabilities of 0.050 and 0.050, while the all-domain model produced 0.062 and 0.070. In the highest-risk groups excluded from the zoomed figure, predicted mortality generally exceeded observed mortality. The full calibration plots are provided in Supplementary eFigure 1.

### 3.5 Clinical utility

Clinical utility was evaluated using operating points and decision-curve analysis.

#### 3.5.1 Sensitivity and Risk Analysis

At 90% specificity, physiological severity produced the clearest standalone sensitivity improvement. In CHoRUS, sensitivity increased from 0.507 for baseline to 0.609 for baseline + physiological severity, compared with 0.540 for treatment exposure and 0.562 for procedure burden; PPV increased from 19.4 to 22.5 deaths per 100 alerts. In MIMIC-IV, sensitivity increased from 0.411 for baseline to 0.557 for baseline + physiological severity, compared with 0.510 for treatment exposure and 0.482 for procedure burden; PPV increased from 13.2 to 17.1 deaths per 100 alerts.

In the highest-risk 10% analysis, CHoRUS baseline + physiological severity captured 57.0% of deaths, with top-10% PPV 25.9 and enrichment 5.7, compared with baseline 48.0%, top-10% PPV 21.8, and enrichment 4.8. In MIMIC-IV, baseline + physiological severity captured 51.8% of deaths, with top-10% PPV 18.4 and enrichment 5.2, compared with baseline 38.1%, top-10% PPV 13.6, and enrichment 3.8. Table 4 reports all operating-point values.

**Table 3:**
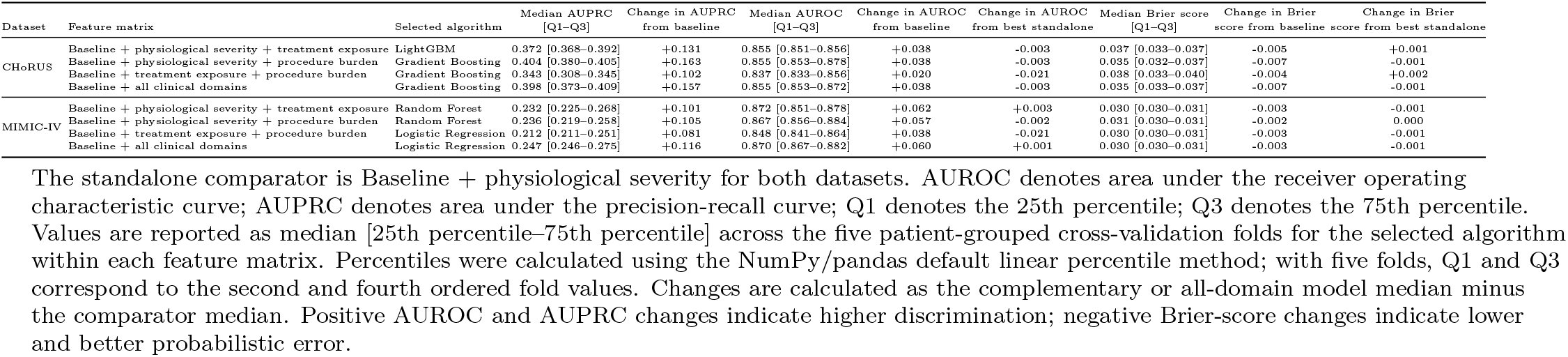
Complementary and all-domain performance in CHoRUS and MIMIC-IV using selected algorithms.

**Table 4:**
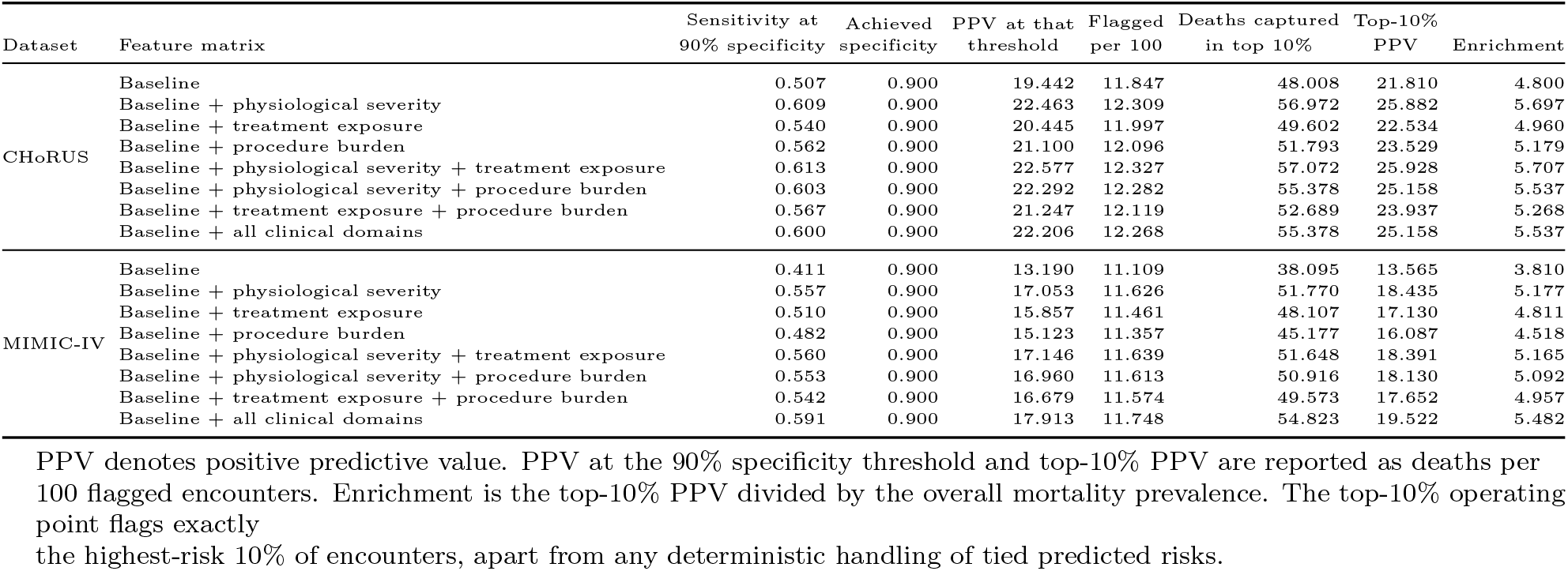
Sensitivity at 90% specificity and highest-risk 10% analyses in CHoRUS and MIMIC-IV.

| Dataset | Feature matrix | Sensitivity at 90% specificity | Achieved specificity | PPV at that threshold | Flagged per 100 | Deaths captured in top 10% | Top-10% PPV | Enrichment |
| --- | --- | --- | --- | --- | --- | --- | --- | --- |
| CHoRUS | Baseline | 0.507 | 0.900 | 19.442 | 11.847 | 48.008 | 21.810 | 4.800 |
|  | Baseline + physiological severity | 0.609 | 0.900 | 22.463 | 12.309 | 56.972 | 25.882 | 5.697 |
|  | Baseline + treatment exposure | 0.540 | 0.900 | 20.445 | 11.997 | 49.602 | 22.534 | 4.960 |
|  | Baseline + procedure burden | 0.562 | 0.900 | 21.100 | 12.096 | 51.793 | 23.529 | 5.179 |
|  | Baseline + physiological severity + treatment exposure | 0.613 | 0.900 | 22.577 | 12.327 | 57.072 | 25.928 | 5.707 |
|  | Baseline + physiological severity + procedure burden | 0.603 | 0.900 | 22.292 | 12.282 | 55.378 | 25.158 | 5.537 |
|  | Baseline + treatment exposure + procedure burden | 0.567 | 0.900 | 21.247 | 12.119 | 52.689 | 23.937 | 5.268 |
|  | Baseline + all clinical domains | 0.600 | 0.900 | 22.206 | 12.268 | 55.378 | 25.158 | 5.537 |
| MIMIC-IV | Baseline | 0.411 | 0.900 | 13.190 | 11.109 | 38.095 | 13.565 | 3.810 |
|  | Baseline + physiological severity | 0.557 | 0.900 | 17.053 | 11.626 | 51.770 | 18.435 | 5.177 |
|  | Baseline + treatment exposure | 0.510 | 0.900 | 15.857 | 11.461 | 48.107 | 17.130 | 4.811 |
|  | Baseline + procedure burden | 0.482 | 0.900 | 15.123 | 11.357 | 45.177 | 16.087 | 4.518 |
|  | Baseline + physiological severity + treatment exposure | 0.560 | 0.900 | 17.146 | 11.639 | 51.648 | 18.391 | 5.165 |
|  | Baseline + physiological severity + procedure burden | 0.553 | 0.900 | 16.960 | 11.613 | 50.916 | 18.130 | 5.092 |
|  | Baseline + treatment exposure + procedure burden | 0.542 | 0.900 | 16.679 | 11.574 | 49.573 | 17.652 | 4.957 |
|  | Baseline + all clinical domains | 0.591 | 0.900 | 17.913 | 11.748 | 54.823 | 19.522 | 5.482 |

PPV denotes positive predictive value. PPV at the 90% specificity threshold and top-10% PPV are reported as deaths per 100 flagged encounters. Enrichment is the top-10% PPV divided by the overall mortality prevalence. The top-10% operating point flags exactly the highest-risk 10% of encounters, apart from any deterministic handling of tied predicted risks.

#### 3.5.2 Decision Curve

In CHoRUS, baseline + physiological severity exceeded baseline net benefit from threshold 0.015 through 0.150, and baseline + physiological severity + procedure burden exceeded baseline over the same range. The all-domain CHoRUS model also exceeded baseline from 0.015 through 0.150, but was not uniformly above baseline + physiological severity. In MIMIC-IV, each standalone clinical-domain model exceeded baseline net benefit over 0.010–0.150, and the all-domain model exceeded both baseline and baseline + physiological severity over 0.010–0.150. The full decision-curve plots are provided in Supplementary eFigure 2.

### 3.6 SHAP Feature Importance results

SHAP was used to summarize model-specific feature attribution for the best-performing algorithm selected for each feature matrix by cross-validated AUPRC [25]. Because SHAP values came from separately trained models with different feature sets and sometimes different algorithms, absolute magnitudes were interpreted within each model and not compared directly across matrices or datasets.

In CHoRUS, recurring non-baseline predictors included Braden Scale physical activity, physical mobility, sensory perception, nutrition components, and pulse rate, concentrated in the physiological-severity domain. In MIMIC-IV, recurring non-baseline predictors included unique and total procedure counts, heparin-flush exposure, vial exposure, and 0.9% sodium chloride exposure or count features. Physiological-severity features, including red cell distribution width, sodium, hemoglobin, urea nitrogen, and white blood cell count, also recurred across feature matrices. Figure 4 shows discrimination and SHAP summaries.

### 3.7 Comparison between CHoRUS and MIMIC-IV

MIMIC-IV had lower mortality prevalence and more unique patients than CHoRUS. Absolute AUPRC values were higher in CHoRUS. The primary standalone clinical domain was the same in both datasets: baseline + physiological severity. The secondary standalone domain differed, with procedure burden stronger in CHoRUS and treatment exposure stronger in MIMIC-IV. For pairwise models, the strongest AUPRC-based models included physiological severity in both datasets. The all-domain model performed best in MIMIC-IV, whereas in CHoRUS the best pairwise model slightly outperformed the all-domain model.

## 4 Discussion

First-24-hour physiological severity features provided the largest and most consistent incremental prognostic value for acute-care 30-day mortality prediction. This was reflected by improvements in median AUPRC, AUROC, and Brier score beyond baseline vulnerability, and by the presence of physiological-severity features in all best-performing pairwise matrices. Treatment exposure and procedure burden produced smaller gains, and the main domain-level result replicated in MIMIC-IV. Threshold-dependent analyses and decision-curve analysis supported the same conclusion by showing improved risk stratification and greater estimated net benefit for models containing physiological-severity information.

The finding is clinically plausible. Baseline demographic, utilization, and comorbidity features estimate underlying vulnerability, whereas first-24-hour physiological measurements capture current physiological state and acute illness severity, including cardiovascular, respiratory, renal, metabolic, and hematologic dysfunction [26, 27, 28, 29]. This makes physiological-severity features directly relevant to early monitoring and escalation decisions. The operating-point results translated discrimination gains into clinically interpretable terms: models with physiological-severity features identified more deaths at 90% specificity and captured more mortality events in the highest-risk decile. These retrospective analyses show potential clinical utility, not proven improvement in patient outcomes.

The secondary domain differed by dataset. Procedure burden was stronger than treatment exposure in CHoRUS, whereas treatment exposure was stronger in MIMIC-IV. This suggests that these domains may depend more heavily on local documentation, clinical practice, coding standards, institutional protocols, and resource availability. SHAP summaries supported the importance of physiological-severity information, but the highest-ranking individual features differed across datasets. Thus, physiological severity may be more reproducible at the domain level than at the identical-feature level.

Combining all domains did not uniformly improve prediction. In CHoRUS, the best model combined baseline, physiological severity, and procedure burden; adding treatment exposure slightly lowered AUPRC while leaving displayed AUROC and Brier score unchanged. In MIMIC-IV, the all-domain model performed best, but only modestly exceeded the best pairwise model. These findings suggest that indiscriminate feature aggregation may not be necessary. Parsimonious models centered on high-quality physiological data may be easier to audit, maintain, calibrate, and implement than broader models that add less stable treatment and procedure signals.

This study extends prior mortality-prediction work showing that richer clinical data improve prognostic performance [5, 6, 7]. Rather than only asking whether more EHR data improve prediction, we compared the standalone and complementary value of clinically interpretable early domains within a consistent first-24-hour framework across two datasets. This provides a reusable approach for health systems to evaluate which local data streams should be prioritized for acute-care risk modeling, physiological measurement-quality improvement, and decision-support design. Strengths include the clinically interpretable domain framework, independent domain-level replication, patient-level grouped cross-validation, multiple algorithms, calibration, decision-curve analysis, operating-point analyses, and SHAP summaries.

This study has several limitations. These include different domain definitions across datasets, use of MIMIC-IV as replication rather than external validation of a locked model, imbalanced outcomes without class-balancing analyses, possible monitoring-intensity signals in count and missingness features, site-dependent treatment and procedure documentation, possible overlap between feature-domain and algorithm effects, absence of subgroup or fairness analyses, and the retrospective design.

The main clinical implication is that early physiological and laboratory measurements appear especially useful for identifying acute-care encounters at elevated short-term mortality risk. Baseline vulnerability remains informative, but current physiological state provides the most consistent added prognostic value. Treatment exposure and procedure burden may add context, but their value was smaller and less stable. For implementation, these findings support prioritizing reliable physiological measurement capture, unit standardization, missingness auditing, and calibration of physiologically centered risk models before expanding to broader treatment and procedure features.

## Data Availability

The CHoRUS data analyzed in this study are available through the controlled-access CHoRUS cloud environment. Access requires registration and authorization, and the data cannot be downloaded from the secure workspace. Information about requesting access is available at https://chorus4ai.org/dataset/.

MIMIC-IV is available to credentialed researchers through PhysioNet after completion of the required human-subjects training and acceptance of the PhysioNet Credentialed Health Data Use Agreement. This study used MIMIC-IV version 3.1, available at https://physionet.org/content/mimiciv/3.1/.

Patient-level data from either dataset cannot be redistributed by the authors.

## Code Availability

The reproducible pipeline is available at https://github.com/Ria-Chaudhry/Mortality_Prediction.

## Acknowledgement

Research reported in this publication was supported by the Office of the Director, National Institutes of Health Common Fund under award number OT2OD032701. The work is solely the responsibility of the authors and does not necessarily represent the official view of CHoRUS or the National Institutes of Health. We thank the CHoRUS Clinical Care for AI consortium and its contributing institutions for making the CHoRUS version 1 dataset available for this study.

We thank the MIT Laboratory for Computational Physiology and Beth Israel Deaconess Medical Center for creating and making available the MIMIC-IV database. This study used MIMIC-IV version 3.1, accessed through PhysioNet, following completion of the required training and data-use agreement.

## Author Information

Ria Chaudhry, Department of Psychological and Brain Sciences, University of Massachusetts Amherst, Amherst, MA 01003, USA.

Zhe Sage Chen, PhD, Department of Psychiatry and Department of Neuroscience, New York University Grossman School of Medicine, New York, NY 10016, USA.

## Author Contributions

**Concept and design:** Chaudhry, Chen.

**Acquisition, analysis, or interpretation of data:** Chaudhry, Chen.

**Drafting of the manuscript:** Chaudhry.

**Critical revision of the manuscript for important intellectual content:** Chaudhry, Chen.

**Statistical analysis:** Chaudhry.

**Administrative, technical, or material support:** Chen.

**Supervision:** Chen.

