## Supplementary Material for "Assessing Acute-Care 30-Day Mortality Prediction Using Clinical Features in CHoRUS Clinical Care for AI and MIMIC-IV"

**Supplementary contents.** Methods; demographic characteristics; feature definitions; model hyperparameters; performance metrics; confidence intervals; best-model clinical-domain comparisons; calibration curves; decision curves; sensitivity at 90% specificity; top-10% risk analysis; SHAP feature-importance tables; and features selected per fold.

**Performance Metrics** **10**

**Confidence Intervals** **21**

**Best-Model Clinical-Domain Comparisons** **22**

**Calibration Plots** **23**

**Decision-Curve Plots** **25**

**Sensitivity at 90% Specificity** **25**

**Top-10% Risk Analysis** **26**

**SHAP Feature-Importance Tables** **27**

**Features Selected per Fold** **62**

### Supplementary Methods

#### eMethods 1. Cohort Definitions and Visit-Type Mapping

CHoRUS cohort construction procedure is outlined here. First all visits were restricted to acute visit types. Acute visit types included: Hospital, Observation, Inpatient, Emergency room, Emergency and Inpatient visit. These were chosen from the visit type list to account for acute care visits. Then the cohort was deduplicated and temporally invalid visits were removed. Temporally invalid visits were defined by visit start date/time and visit end date/time. For a given visit, if the visit end date/time occurred before the visit start date/time, the visit was regarded as temporally invalid and removed from the cohort. This was done sequentially to preserve computational resources: the cohort was first limited by visit type, then deduplicated, and then temporally invalid visits were removed. There were no temporally invalid visits after deduplication; therefore, the final cohort count was 22,098 after deduplication. Further, all visits that included mortality occurring before the first 24 hours of visit start were removed. Visits shorter than 24 hours were retained and were not excluded solely on the basis of visit duration.

Multiple eligible acute-care visits from the same patient were retained because the unit of analysis was the encounter. To prevent information leakage across validation sets, data were partitioned at the patient level using five-fold grouped cross-validation. All encounters belonging to a given patient were assigned to the same fold, ensuring that no patient contributed visits to both the training and validation sets within the same cross-validation iteration. Performance metrics were calculated on the held-out encounters in each fold and summarized across the five folds

**eMethods 2. MIMIC-IV Patient-Level Subsampling**

We used the admissions, patients, lab events, prescriptions and procedures tables to construct our cohort. We used a reproducible patient-level stratified subsampling procedure for the independent replication analysis. Admissions were first grouped by patient identifier. Patients were then stratified according to whether they had at least one admission meeting the 30-day mortality outcome definition. Within each stratum, patients were randomly shuffled using a fixed random seed, and complete patient admission histories were retained until the sampled cohort reached 23,000 admissions. This approach preserved patient clusters, avoided sampling individual admissions from the same patient independently, and allowed subsequent patient-level cross-validation without train-test patient overlap. The same sampled cohort was used across all MIMIC-IV feature matrices.

**eMethods 3. Feature Construction**

**Baseline features.**

Baseline vulnerability was represented with demographic, visit-type, prior-utilization, and Charlson comorbidity predictors available at or before the index acute-care visit. Age at visit was calculated from the patient's year of birth and the index visit start date. Categorical variables were represented with one-hot encoded indicator columns when the saved model matrix stored encoded columns; otherwise, the fold-level feature-selection tables display the broad raw predictor labels while the categorical encoding is applied inside the modelling pipeline.

In CHoRUS, the fixed baseline model-ready matrix contained 21 columns: four numeric or binary variables retained directly (`age_at_visit`, `prior_visit_count`, `has_prior_visit`, and `prior_acute_visit_count`); sex indicators for female and male; race indicators for white, black, Asian, other, and unknown; ethnicity indicators for Hispanic, not Hispanic, other, and unknown; visit-type indicators for hospital, observation, inpatient visit, emergency room, and emergency-and-inpatient visit; and `Charlson_Comorbidity_Score`.

In MIMIC-IV, the current preprocessing code used nine raw baseline predictors: `age_at_visit`, `sex`, `race`, `ethnicity`, `visit_type`, `has_prior_visit`, `prior_visit_count`, `prior_acute_visit_count`, and `charlson_index`. Five numeric or binary variables were retained directly: `age_at_visit`, `has_prior_visit`, `prior_visit_count`, `prior_acute_visit_count`, and `charlson_index`. The one-hot encoded categories verified from the current encoded baseline output were sex `F` and `M`; race `AMERICAN_INDIAN_OR_ALASKA_NATIVE`, `ASIAN`, `BLACK`, `MULTIPLE`, `NATIVE_HAWAIIAN_OR_PACIFIC_ISLANDER`, `OTHER`, `UNKNOWN`, and `WHITE`; ethnicity `HISPANIC_OR_LATINO`, `NOT_HISPANIC_OR_LATINO`, and `UNKNOWN`; and visit type `AMBULATORY OBSERVATION`, `DIRECT EMER.`, `DIRECT OBSERVATION`, `EU OBSERVATION`, `EW EMER.`, `OBSERVATION ADMIT`, and `URGENT`. These 20 categorical indicators plus five retained numeric or binary variables yielded 25 transformed baseline columns in the current MIMIC-IV encoded baseline output. The MIMIC-IV features-selected-per-fold table displays the broad raw predictor labels for categorical predictors, so a row labelled `sex`, `race`, `ethnicity`, or `visit_type` does not mean that only one encoded column was used.

`prior_visit_count` was a numeric visit-level predictor representing the number of visits the patient had before the index visit. `has_prior_visit` was a binary predictor indicating whether the patient had at least one visit before the index visit; no separate `any_prior_visit` variable was present in the inspected MIMIC-IV code. `prior_acute_visit_count` was a numeric predictor representing the number of prior acute-care visits before the index visit. Prior acute-care utilization used the same visit-type definitions used to construct each acute-care study cohort; in the inspected MIMIC-IV code, acute-care admission types were `AMBULATORY OBSERVATION`, `DIRECT EMER.`, `DIRECT OBSERVATION`, `EU OBSERVATION`, `EW EMER.`, `OBSERVATION ADMIT`, and `URGENT`.

The Charlson Comorbidity Index (CCI) was used as an aggregate measure of pre-existing disease burden at the time of the acute-care visit. Broad Charlson condition categories were harmonized across datasets, but source vocabularies and operational mappings were dataset-specific. Diagnoses recorded before the prediction window were mapped to Charlson categories including myocardial infarction, congestive heart failure, peripheral vascular disease, cerebrovascular disease, dementia, chronic pulmonary disease, rheumatic or connective-tissue disease, peptic ulcer disease, mild or severe liver disease, diabetes with or without complications, hemiplegia or paraplegia, renal disease, malignant cancer, metastatic solid tumor, and HIV/AIDS. The inspected MIMIC-IV implementation used ICD-9 and ICD-10 diagnosis-code mappings, excluded current-admission diagnoses, and used a 365-day historical lookback before the index admission. The CHoRUS source in the project folder did not include an inspectable cohort-building script, so identical ICD-code implementation and lookback could not be verified from the files available for this revision. The non-age-adjusted CCI was used because age was included separately as a baseline predictor.

**Baseline + physiological severity: first-24-hour measurement features.**

Measurement features were extracted from the OMOP measurement table. For each eligible visit, we included measurements with timestamps greater than or equal to the visit start datetime and less than 24 hours after visit start. This ensured that the predictors were restricted to the prespecified early prediction window and did not use information recorded later in the hospitalization. We selected the 50 most frequently occurring measurement concepts prior to model training. Selected measurement concepts included heart rate, respiratory rate, oxygen saturation, blood pressure measurements, Glasgow Coma Scale measures, hemoglobin, creatinine, sodium, potassium, chloride, blood urea nitrogen, and related clinical measurements.

For each selected measurement concept, non-null numeric values recorded during the first 24 hours were summarized at the visit level using the mean, minimum, maximum, standard deviation, and count of recorded values. These summaries were then transformed into a wide feature matrix with one row per visit_occurrence_id. For example, a single measurement concept could contribute separate features representing its mean value, minimum value, maximum value, variability, and number of observations during the first 24 hours. To preserve information about whether a measurement was obtained, missingness indicators were also created for each measurement concept. After missingness indicators were created, missing numeric summary and count features were imputed as 0.

Fifty measurement concepts were initially selected. Three concepts contained null or unusable values and were removed, leaving 47 final measurement concepts for analysis. Six summary features were generated per final concept: mean, minimum, maximum, standard deviation, count, and missingness. The resulting physiological-severity candidate matrix therefore contained 47 concepts $\times$ 6 summaries = 282 features. To standardize the number of features contributed by each clinical domain, feature selection was performed separately within each cross-validation training fold. Within each fold, the measurement-derived candidates were ranked according to their occurrence frequency in the training partition, and the 21 most frequently occurring features were retained. The selected feature set was then applied unchanged to the corresponding validation partition. Because selection was repeated independently within each training fold, the specific measurement-derived features could differ across folds, while every physiological-severity matrix consistently contributed 21 features. This procedure reduced information leakage and ensured that comparisons across clinical domains were not driven by differences in feature-set size.

**Baseline + treatment exposure: first-24-hour medication features.**

Medication features were extracted from the OMOP drug exposure table. For each eligible acute-care visit, medication exposures were included if the drug exposure start datetime occurred at or after the visit start date/time and before 24 hours after visit start. This used the same first-24-hour prediction window as measurement features and ensured that medication features reflected information available during the first 24 hours of the encounter.

To reduce sparsity and avoid creating a high-dimensional medication feature matrix, we first identified the most common medication concepts recorded during the first 24 hours across the analytic cohort. The 50 most frequently occurring drug concept IDs were selected for Baseline + treatment exposure feature construction. For each selected medication concept, two visit-level features were created: a binary exposure indicator and an exposure count. The binary exposure feature indicated whether the medication was recorded at least once during the first 24 hours of the visit, while the count feature represented the number of recorded exposures for that medication during the same time window.

Medication features were then transformed into a wide visit-level feature matrix with one row per visit_occurrence_id. Visits without a selected medication exposure were assigned a value of 0 for the corresponding medication exposure and count columns. Four aggregate treatment-exposure features were included: `unique_drug_count_24h`, `repeat_drug_exposure_count_24h`, `time_to_first_drug_hours`, and `any_drug_24h`.

This process produced 104 medication-derived candidate features. To standardize the number of features contributed by each clinical domain, feature selection was performed separately within each cross-validation training fold. Within each fold, the medication-derived candidates were ranked according to their occurrence frequency in the training partition, and the 21 most frequently occurring features were retained. The selected feature set was then applied unchanged to the corresponding validation partition. Because selection was repeated independently within each training fold, the specific medication-derived features could differ across folds, while every treatment exposure matrix consistently contributed 21 features.

**Baseline + procedure burden: first-24-hour procedure-derived features.**

Procedure features were extracted from the OMOP procedure_occurrence table. For each acute-care visit, procedure occurrence information was included if the procedure occurrence start date/time occurred at or after visit start date/time and before 24 hours after the visit start. This feature matrix used the same first-24-hour prediction window as the physiological-severity and treatment-exposure feature matrices.

To reduce sparsity and avoid creating a high-dimensional procedure feature matrix, we first identified the most common procedure concepts recorded during the first 24 hours across the analytic cohort. The 50 most frequently occurring procedure concept IDs were selected for Baseline + procedure burden feature construction. For each selected procedure concept, two visit-level features were created: a binary procedure presence indicator and a procedure count. The binary feature indicated whether the procedure was recorded at least once during the first 24 hours of the visit, while the count feature represented the number of recorded occurrences for that procedure during the same time window. In addition, three nonredundant aggregate procedure-burden features were included: `unique_procedure_count_24h`, the number of unique procedure concepts recorded within the first 24 hours; `procedure_count_total_24h`, the total number of procedure records recorded within the first 24 hours; and `any_procedure_24h`, a binary indicator coded as 1 if at least one procedure was recorded during the first 24 hours of the index visit and 0 otherwise. Procedure missingness was not included as a selected aggregate procedure-burden feature. In the inspected MIMIC-IV procedure script, `procedure_missing_24h` was retained in the saved procedure matrix but removed from feature-selection candidates because it was the exact inverse of `any_procedure_24h`.

Procedure features were then transformed into a wide visit-level feature matrix with one row per visit_occurrence_id. Visits without a selected procedure occurrence were assigned a value of 0 for the corresponding procedure indicator and count columns.

This process produced 103 procedure-derived candidate features. To standardize the number of features contributed by each clinical domain, feature selection was performed separately within each cross-validation training fold. Within each fold, the procedure-derived candidates were ranked according to their occurrence frequency in the training partition, and the 21 most frequently occurring features were retained. The selected feature set was then applied unchanged to the corresponding validation partition. Because selection was repeated independently within each training fold, the specific procedure-derived features could differ across folds, while every procedure-burden matrix consistently contributed 21 features.

### Supplementary Tables

**eTable 1. Baseline predictors**

| No. | Model feature |
|---|---|
| 1 | `age_at_visit` |
| 2 | `prior_visit_count` |

| No. | Model feature |
|---|---|
| 3 | has_prior_visit |
| 4 | prior_acute_visit_count |
| 5 | sex_female |
| 6 | sex_male |
| 7 | race_white |
| 8 | race_black |
| 9 | race_asian |
| 10 | race_other |
| 11 | race_unknown |
| 12 | ethnicity_hispanic |
| 13 | ethnicity_not_hispanic |
| 14 | ethnicity_other |
| 15 | ethnicity_unknown |
| 16 | visit_type_hospital |
| 17 | visit_type_observation |
| 18 | visit_type_inpatient_visit |
| 19 | visit_type_emergency_room |
| 20 | visit_type_emergency_and_inpatient_visit |
| 21 | Charlson_Comorbidity_Score |

*Note.* Feature names are reported as used in the CHoRUS analytic model matrix. The CHoRUS baseline matrix contained 21 model-ready features. The MIMIC-IV baseline analysis used nine raw baseline predictors and, in the current encoded baseline output, five direct numeric or binary predictors plus 20 one-hot indicators, yielding 25 transformed baseline columns.

### eTable 2. Measurement concepts

| Concept ID | Concept name |
|---|---|
| 3661605 | Age adjusted minimum alveolar concentration |
| 4134573 | Bispectral index |
| 3020891 | Body temperature |
| 4302666 | Body temperature |
| 3014576 | Chloride [Moles/volume] in Serum or Plasma |
| 3016723 | Creatinine [Mass/volume] in Serum or Plasma |
| 3012888 | Diastolic blood pressure |
| 4353938 | End tidal carbon dioxide concentration |
| 4137697 | Expired isoflurane concentration |
| 4140731 | Expired sevoflurane concentration |
| 3037347 | Friction and shear Braden scale |

| Concept ID | Concept name |
|---|---|
| 4083352 | Glasgow Coma Scale motor response subscore |
| 4084912 | Glasgow Coma Scale verbal response subscore |
| 4084277 | Glasgow Coma Score eye opening subscore |
| 4296538 | Glasgow coma scale |
| 3027018 | Heart rate |
| 4239408 | Heart rate |
| 4140152 | Inspired isoflurane concentration |
| 4353936 | Inspired oxygen concentration |
| 4137519 | Inspired sevoflurane concentration |
| 4354253 | Invasive diastolic arterial pressure |
| 4108290 | Invasive mean arterial pressure |
| 4353843 | Invasive systolic arterial pressure |
| 3009744 | MCHC [Mass/volume] by Automated count |
| 3037022 | Moisture exposure Braden scale |
| 4068414 | Non-invasive diastolic arterial pressure |
| 4108289 | Non-invasive mean arterial pressure |
| 4354252 | Non-invasive systolic arterial pressure |
| 3035816 | Nutrition intake pattern Braden scale |
| 4218834 | Oral temperature |
| 40762499 | Oxygen saturation in Arterial blood by Pulse oximetry |
| 4020553 | Oxygen saturation measurement |
| 4101694 | Peak inspiratory pressure |
| 4196147 | Peripheral oxygen saturation |
| 3037318 | Physical activity Braden scale |
| 3035206 | Physical mobility Braden scale |
| 3023103 | Potassium [Moles/volume] in Serum or Plasma |
| 4098046 | Pulse oximetry |
| 4301868 | Pulse rate |
| 4064625 | QRS complex feature |
| 3024171 | Respiratory rate |
| 4313591 | Respiratory rate |
| 3036098 | Sensory perception Braden scale |
| 3019550 | Sodium [Moles/volume] in Serum or Plasma |
| 3004249 | Systolic blood pressure |
| 3013682 | Urea nitrogen [Mass/volume] in Serum or Plasma |
| 4108137 | Ventilator delivered tidal volume |

*Note.* The table lists the 47 OMOP concept IDs used to derive first-24-hour measurement features. Derived summary features are described in the eMethods.

**eTable 3. Medication concepts**

| Concept ID | Concept name |
|---|---|
| 35782395 | 10 ML Phenylephrine 0.1 MG/ML Injectable Solution |
| 19127213 | 10 ML sodium chloride 9 MG/ML Prefilled Syringe |
| 19079322 | 100 ML potassium chloride 0.1 MEQ/ML Injection |
| 40220388 | 100 ML propofol 10 MG/ML Injection [Diprivan] |
| 40221385 | 100 ML sodium chloride 9 MG/ML Injection |
| 42479436 | 1000 ML Sodium Chloride 9 MG/ML Injectable Solution |
| 40220357 | 1000 ML sodium chloride 9 MG/ML Injection |
| 35605482 | 2 ML ondansetron 2 MG/ML Injection |
| 2025002 | 250 ML sodium chloride 9 MG/ML Injectable Solution [JW NS] by JW |
| 1560751 | 50 ML glucose 500 MG/ML Prefilled Syringe |
| 46275280 | 50 ML magnesium sulfate 40 MG/ML Injection |
| 40752856 | Fentanyl Injectable Solution [Sublimaze] |
| 21050327 | Glucose Injectable Solution [Plasma-lyte] |
| 40744996 | Sugammadex 100 MG/ML Injectable Solution [Bridion] |
| 1127433 | acetaminophen 325 MG Oral Tablet |
| 1128524 | acetaminophen 650 MG Extended Release Oral Tablet [Tylenol] |
| 35872724 | acetaminophen Oral Tablet [Andufen] |
| 1113346 | aspirin 81 MG Chewable Tablet |
| 19073712 | aspirin 81 MG Delayed Release Oral Tablet |
| 1831072 | atorvastatin 40 MG Oral Tablet [Lipitor] Box of 84 by Upjohn Products |
| 19135374 | calcium chloride / potassium chloride 0.3 MG/ML / sodium chloride 6 MG/ML / sodium lactate 3.1 MG/ML Injectable Solution |
| 35830655 | cefazolin 1000 MG Injectable Solution [CEFAMEZIN] |
| 46287352 | cefepime 2000 MG Injection [Maxipime] |
| 42708658 | docusate sodium 100 MG Oral Capsule [Colace] |
| 44784779 | docusate sodium 100 MG Oral Capsule [DOK] |
| 40042753 | enoxaparin Injectable Solution [Lovenox] |
| 35603455 | fentanyl 0.05 MG/ML Injection [Sublimaze] |
| 19077513 | folic acid 1 MG Oral Tablet |
| 19016865 | glucagon (rDNA) 1 MG Injection [GlucaGen] |
| 1560525 | glucose 0.4 MG/MG Oral Gel |
| 1718370 | heparin Injectable Solution |
| 43011850 | heparin sodium, porcine 5000 UNT/ML Injectable Solution |
| 35603598 | hydromorphone Injection [Dilaudid] |
| 35854313 | iohexol 350 MG/ML Injectable Solution |
| 40048832 | ketorolac Injectable Solution [Toradol] |
| 40227056 | lidocaine hydrochloride 20 MG/ML Injectable Solution [Xylocaine] |

| Concept ID | Concept name |
|---|---|
| 46275281 | magnesium sulfate 40 MG/ML Injection |
| 1110410 | morphine |
| 19120263 | mupirocin 20 MG/ML [Bactroban] |
| 19005968 | ondansetron 2 MG/ML Injectable Solution [Zofran] |
| 19000634 | ondansetron 4 MG [Zofran] |
| 40232756 | oxycodone hydrochloride 5 MG Oral Tablet |
| 40232759 | oxycodone hydrochloride 5 MG Oral Tablet [Roxicodone] |
| 19080217 | pantoprazole 40 MG Delayed Release Oral Tablet |
| 40220390 | propofol 10 MG/ML Injection [Diprivan] |
| 42708029 | rocuronium bromide 10 MG/ML Injectable Solution [Zemuron] |
| 938330 | sennosides, USP 8.6 MG Oral Tablet |
| 19102651 | sennosides, USP 8.6 MG Oral Tablet [Senokot] |
| 19079524 | sodium chloride 9 MG/ML Injectable Solution |
| 19010309 | water |

*Note.* The table lists the 50 OMOP concept IDs used to derive first-24-hour medication count and presence features. Four aggregate treatment-exposure features were also used: `unique_drug_count_24h`, `repeat_drug_exposure_count_24h`, `time_to_first_drug_hours`, and `any_drug_24h`.

### eTable 4. Procedure-occurrence concepts

| Concept ID | Concept name |
|---|---|
| 4145308 | 12 lead ECG |
| 2147483056 | ASSISTANCE \| LEVEL OF ASSISTANCE \| LEVEL OF ASSISTANCE |
| 40483208 | Assessment of risk of venous thromboembolism |
| 2212090 | Basic metabolic panel |
| 4035541 | Blood cell count, automated |
| 4078442 | Catheter procedure |
| 40217302 | Clinical decision support mechanism under Medicare appropriate-use criteria |
| 4319466 | Complete blood count with white cell differential, automated |
| 3661905 | Comprehensive metabolic panel |
| 2211327 | Computed tomography, head or brain; without contrast material |
| 2514442 | Critical care evaluation and management; each additional 30 minutes |
| 2514441 | Critical care evaluation and management; first 30–74 minutes |
| 2313869 | Transthoracic echocardiography with spectral and color Doppler |
| 2313816 | Routine 12-lead ECG; interpretation and report only |
| 2313814 | Routine 12-lead ECG; with interpretation and report |
| 2147483387 | GASTROINTESTINAL \| ABDOMEN \| Abdomen INSPECTION |

| Concept ID | Concept name |
|---|---|
| 2212349 | Blood gases with directly measured oxygen saturation |
| 4149519 | Glucose measurement |
| 4144235 | Glucose measurement, blood |
| 2147483603 | HEENT \| Eyes/Vision \| Left Eye Assessment |
| 2147483590 | HEENT \| Eyes/Vision \| Right Eye Assessment |
| 2147483327 | HEENT \| NECK \| Neck Assessment |
| 2147483587 | HEENT \| THROAT \| Voice Assessment |
| 2147483323 | HEENT \| Throat/Buccal \| Throat Assessment |
| 4098135 | Hepatic function panel |
| 2514406 | Initial inpatient or observation care with high-complexity medical decision-making |
| 4213288 | Insertion of catheter into artery |
| 42536500 | Insertion of catheter into peripheral vein |
| 4049832 | Insertion of peripheral venous cannula into peripheral vein |
| 2147482992 | LINE CARE \| LINE SITE ASSESSMENT \| LINE SITE ASSESSMENT |
| 2147482991 | LINE CARE \| LINE STATUS \| LINE STATUS |
| 4213582 | Lactic acid measurement |
| 4243005 | Magnesium measurement |
| 46272910 | Measurement of ionized calcium in blood specimen |
| 4141149 | Open insertion of central venous catheter |
| 4021323 | Pain assessment |
| 4083171 | Pain management |
| 4017908 | Phosphate, inorganic measurement |
| 4163872 | Plain chest X-ray |
| 4245261 | Prothrombin time |
| 725068 | Radiologic examination, chest; single view |
| 2514409 | Subsequent inpatient or observation care with high-complexity medical decision-making |
| 2147482962 | Surgical Airway \| Airway \| Airway Measured From |
| 2147482961 | Surgical Airway \| Airway \| Airway Secured By |
| 4021291 | Troponin measurement |
| 2212605 | Troponin, quantitative |
| 2000000027 | Ultrasound \| Ultrasound \| US LYMPH NODE BIOPSY |
| 2212166 | Automated urinalysis with microscopy |
| 36713195 | X-ray of chest anteroposterior view |
| 0 | Unmapped procedure |

*Note*. The table lists the procedure concepts used to derive first-24-hour procedure presence and count features. Three aggregate procedure-burden features were also used: `unique_procedure_count_24h`, `procedure_count_total_24h`, and `any_procedure_24h`. Procedure missingness was not included as a selected aggregate feature.

eTable 6. Model preprocessing and hyperparameters used across all feature matrices

| Model | Preprocessing | Explicitly specified hyperparameters |
|---|---|---|
| Logistic regression | Median imputation; standardization using `StandardScaler` | `max_iter = 1000`; `random_state = 42` |
| Random forest | Median imputation | `n_estimators = 300`; `random_state = 42` |
| Gradient boosting | Median imputation | `random_state = 42` |
| LightGBM | Median imputation | `n_estimators = 500`; `learning_rate = 0.03`; `num_leaves = 31`; `max_depth = -1`; `subsample = 0.8`; `subsample_freq = 1`; `colsample_bytree = 0.8`; `random_state = 42`; `n_jobs = -1`; `verbose = -1` |

*Note.* All feature matrices used the same four model classes and the same hyperparameters. Feature matrices differed only in the predictor domains supplied to the model pipelines. Hyperparameters not explicitly specified in the analysis code used the corresponding software package defaults. Median imputation was performed within each model pipeline. Only logistic regression included feature standardization.

eTable 5. Leakage audit and final handling of potentially problematic variables

| Variable or domain reviewed | Potential leakage concern | Final handling |
|---|---|---|
| Death date | Directly defines the mortality outcome and would leak outcome information if used as a predictor | Used only to construct the 30-day mortality outcome; excluded from all predictor matrices |
| Visit end datetime | May encode length of stay, discharge timing, or post-baseline clinical course | Excluded from all predictor matrices |
| Post-24-hour measurements | Measurements after the first 24 hours occur outside the predefined prediction window | Excluded; measurement features were restricted to the first 24 hours after visit start |
| Post-24-hour medication exposures | Medication exposures after the prediction window may reflect later deterioration, treatment escalation, or outcome proximity | Excluded; medication features were restricted to the first 24 hours after visit start |
| Post-24-hour procedures | Procedures after the prediction window may reflect later clinical course or complications | Excluded; procedure features were restricted to the first 24 hours after visit start |
| Future visits after index visit | Future utilization may occur after the prediction time point and could encode subsequent clinical trajectory | Excluded; prior-visit features were computed using only visits before the index visit start time |
| Repeated visits from the same patient | Visit-level random splitting could place visits from the same patient in both training and testing folds | Patient-level cross-validation was used to prevent patient overlap across train and test folds |
| Duplicate visit rows | Duplicate records could inflate performance or duplicate outcome-predictor combinations | Visits were deduplicated by visit identifier before model development |
| Temporally invalid visits | Invalid visit timing could distort prediction-window construction or outcome timing | Visits with invalid temporal ordering were removed before final cohort construction |
| Outcome-window definition | Incorrect outcome timing could include deaths before visit start or outside the intended 30-day window | Mortality was defined using deaths occurring at or after 24 hours following visit start and within 30 days of the index acute-care visit. Visits with death occurring before 24 hours following visit start were excluded; visits shorter than 24 hours were retained and were not excluded solely on the basis of visit duration |

### Performance Metrics

This section reports cross-validation performance metrics for all feature matrices and model algorithms in CHoRUS and MIMIC-IV. Feature-domain terminology is standardized to Physiological Severity, Treatment Exposure, and Procedure Burden.

eTable 7. CHoRUS performance metrics for all feature matrices and algorithms.

| Feature matrix | Model | Fold | Accuracy | AUPRC | AUROC | Precision | Sensitivity | Specificity | F1 | Brier score | TN | TP | FN | FP |
|---|---|---|---|---|---|---|---|---|---|---|---|---|---|---|
| Baseline | Logistic Regression | 1 | 0.962 | 0.254 | 0.838 | 0.000 | 0.000 | 1.000 | 0.000 | 0.032 | 4361 | 0 | 174 | 0 |
| Baseline | Logistic Regression | 2 | 0.951 | 0.256 | 0.813 | 0.000 | 0.000 | 1.000 | 0.000 | 0.041 | 4187 | 0 | 215 | 2 |
| Baseline | Logistic Regression | 3 | 0.948 | 0.195 | 0.761 | 0.000 | 0.000 | 0.999 | 0.000 | 0.044 | 4140 | 0 | 221 | 4 |
| Baseline | Logistic Regression | 4 | 0.958 | 0.227 | 0.797 | 1.000 | 0.005 | 1.000 | 0.010 | 0.036 | 4367 | 1 | 191 | 0 |
| Baseline | Logistic Regression | 5 | 0.952 | 0.202 | 0.743 | 0.000 | 0.000 | 0.999 | 0.000 | 0.042 | 4030 | 0 | 202 | 3 |
| Baseline | Random Forest | 1 | 0.959 | 0.185 | 0.751 | 0.410 | 0.144 | 0.992 | 0.213 | 0.037 | 4325 | 25 | 149 | 36 |
| Baseline | Random Forest | 2 | 0.948 | 0.194 | 0.770 | 0.397 | 0.144 | 0.989 | 0.212 | 0.046 | 4142 | 31 | 184 | 47 |
| Baseline | Random Forest | 3 | 0.943 | 0.180 | 0.698 | 0.307 | 0.104 | 0.987 | 0.155 | 0.049 | 4092 | 23 | 198 | 52 |
| Baseline | Random Forest | 4 | 0.953 | 0.160 | 0.755 | 0.312 | 0.104 | 0.990 | 0.156 | 0.041 | 4323 | 20 | 172 | 44 |
| Baseline | Random Forest | 5 | 0.944 | 0.172 | 0.764 | 0.267 | 0.099 | 0.986 | 0.144 | 0.047 | 3978 | 20 | 182 | 55 |
| Baseline | Gradient Boosting | 1 | 0.960 | 0.248 | 0.845 | 0.318 | 0.040 | 0.997 | 0.071 | 0.032 | 4346 | 7 | 167 | 15 |
| Baseline | Gradient Boosting | 2 | 0.951 | 0.244 | 0.817 | 0.472 | 0.079 | 0.995 | 0.135 | 0.042 | 4170 | 17 | 198 | 19 |
| Baseline | Gradient Boosting | 3 | 0.948 | 0.241 | 0.808 | 0.444 | 0.072 | 0.995 | 0.125 | 0.044 | 4124 | 16 | 205 | 20 |
| Baseline | Gradient Boosting | 4 | 0.957 | 0.203 | 0.820 | 0.412 | 0.073 | 0.995 | 0.124 | 0.038 | 4347 | 14 | 178 | 20 |
| Baseline | Gradient Boosting | 5 | 0.952 | 0.211 | 0.795 | 0.458 | 0.054 | 0.997 | 0.097 | 0.042 | 4020 | 11 | 191 | 13 |
| Baseline | LightGBM | 1 | 0.959 | 0.227 | 0.828 | 0.303 | 0.057 | 0.995 | 0.097 | 0.034 | 4338 | 10 | 164 | 23 |
| Baseline | LightGBM | 2 | 0.949 | 0.225 | 0.807 | 0.408 | 0.093 | 0.993 | 0.152 | 0.043 | 4160 | 20 | 195 | 29 |
| Baseline | LightGBM | 3 | 0.948 | 0.216 | 0.775 | 0.410 | 0.072 | 0.994 | 0.123 | 0.045 | 4121 | 16 | 205 | 23 |
| Baseline | LightGBM | 4 | 0.957 | 0.227 | 0.810 | 0.469 | 0.078 | 0.996 | 0.134 | 0.037 | 4350 | 15 | 177 | 17 |
| Baseline | LightGBM | 5 | 0.949 | 0.177 | 0.779 | 0.324 | 0.059 | 0.994 | 0.100 | 0.044 | 4008 | 12 | 190 | 25 |
| Baseline + physiological severity | Logistic Regression | 1 | 0.964 | 0.351 | 0.857 | 0.576 | 0.195 | 0.994 | 0.292 | 0.030 | 4336 | 34 | 140 | 25 |
| Baseline + physiological severity | Logistic Regression | 2 | 0.955 | 0.354 | 0.829 | 0.655 | 0.167 | 0.995 | 0.267 | 0.038 | 4170 | 36 | 179 | 19 |
| Baseline + physiological severity | Logistic Regression | 3 | 0.952 | 0.313 | 0.822 | 0.622 | 0.127 | 0.996 | 0.211 | 0.041 | 4127 | 28 | 193 | 17 |
| Baseline + physiological severity | Logistic Regression | 4 | 0.959 | 0.300 | 0.836 | 0.568 | 0.109 | 0.996 | 0.183 | 0.034 | 4351 | 21 | 171 | 16 |
| Baseline + physiological severity | Logistic Regression | 5 | 0.954 | 0.277 | 0.802 | 0.585 | 0.119 | 0.996 | 0.198 | 0.040 | 4016 | 24 | 178 | 17 |
| Baseline + physiological severity | Random Forest | 1 | 0.964 | 0.392 | 0.847 | 0.594 | 0.218 | 0.994 | 0.319 | 0.029 | 4335 | 38 | 136 | 26 |
| Baseline + physiological severity | Random Forest | 2 | 0.958 | 0.339 | 0.822 | 0.719 | 0.214 | 0.996 | 0.330 | 0.039 | 4171 | 46 | 169 | 18 |
| Baseline + physiological severity | Random Forest | 3 | 0.950 | 0.313 | 0.790 | 0.515 | 0.158 | 0.992 | 0.242 | 0.041 | 4111 | 35 | 186 | 33 |
| Baseline + physiological severity | Random Forest | 4 | 0.960 | 0.351 | 0.832 | 0.580 | 0.208 | 0.993 | 0.307 | 0.034 | 4338 | 40 | 152 | 29 |
| Baseline + physiological severity | Random Forest | 5 | 0.954 | 0.353 | 0.849 | 0.571 | 0.158 | 0.994 | 0.248 | 0.037 | 4009 | 32 | 170 | 24 |
| Baseline + physiological severity | Gradient Boosting | 1 | 0.967 | 0.435 | 0.882 | 0.691 | 0.270 | 0.995 | 0.388 | 0.027 | 4340 | 47 | 127 | 21 |
| Baseline + physiological severity | Gradient Boosting | 2 | 0.958 | 0.415 | 0.858 | 0.729 | 0.237 | 0.995 | 0.358 | 0.036 | 4170 | 51 | 164 | 19 |
| Baseline + physiological severity | Gradient Boosting | 3 | 0.952 | 0.323 | 0.833 | 0.597 | 0.167 | 0.994 | 0.261 | 0.041 | 4119 | 37 | 184 | 25 |
| Baseline + physiological severity | Gradient Boosting | 4 | 0.960 | 0.354 | 0.874 | 0.571 | 0.229 | 0.992 | 0.327 | 0.033 | 4334 | 44 | 148 | 33 |
| Baseline + physiological severity | Gradient Boosting | 5 | 0.957 | 0.380 | 0.845 | 0.650 | 0.193 | 0.995 | 0.298 | 0.036 | 4012 | 39 | 163 | 21 |
| Baseline + physiological severity | LightGBM | 1 | 0.964 | 0.392 | 0.866 | 0.573 | 0.270 | 0.992 | 0.367 | 0.029 | 4326 | 47 | 127 | 35 |
| Baseline + physiological severity | LightGBM | 2 | 0.955 | 0.384 | 0.854 | 0.607 | 0.237 | 0.992 | 0.341 | 0.038 | 4156 | 51 | 164 | 33 |

Continued on next page

eTable 7. CHoRUS performance metrics for all feature matrices and algorithms. (continued)

| Feature matrix | Model | Fold | Accuracy | AUPRC | AUROC | Precision | Sensitivity | Specificity | F1 | Brier score | TN | TP | FN | FP |
|---|---|---|---|---|---|---|---|---|---|---|---|---|---|---|
| Baseline + physiological severity | LightGBM | 3 | 0.948 | 0.304 | 0.824 | 0.466 | 0.154 | 0.991 | 0.231 | 0.042 | 4105 | 34 | 187 | 39 |
| Baseline + physiological severity | LightGBM | 4 | 0.958 | 0.359 | 0.855 | 0.510 | 0.255 | 0.989 | 0.340 | 0.034 | 4320 | 49 | 143 | 47 |
| Baseline + physiological severity | LightGBM | 5 | 0.954 | 0.329 | 0.841 | 0.549 | 0.193 | 0.992 | 0.286 | 0.039 | 4001 | 39 | 163 | 32 |
| Baseline + treatment exposure | Logistic Regression | 1 | 0.960 | 0.254 | 0.847 | 0.360 | 0.052 | 0.996 | 0.090 | 0.032 | 4345 | 9 | 165 | 16 |
| Baseline + treatment exposure | Logistic Regression | 2 | 0.952 | 0.292 | 0.830 | 0.647 | 0.051 | 0.999 | 0.095 | 0.040 | 4183 | 11 | 204 | 6 |
| Baseline + treatment exposure | Logistic Regression | 3 | 0.950 | 0.234 | 0.785 | 0.565 | 0.059 | 0.998 | 0.107 | 0.043 | 4134 | 13 | 208 | 10 |
| Baseline + treatment exposure | Logistic Regression | 4 | 0.959 | 0.270 | 0.826 | 0.667 | 0.052 | 0.999 | 0.097 | 0.035 | 4362 | 10 | 182 | 5 |
| Baseline + treatment exposure | Logistic Regression | 5 | 0.952 | 0.250 | 0.774 | 0.500 | 0.030 | 0.999 | 0.056 | 0.040 | 4027 | 6 | 196 | 6 |
| Baseline + treatment exposure | Random Forest | 1 | 0.961 | 0.260 | 0.839 | 0.457 | 0.092 | 0.996 | 0.153 | 0.032 | 4342 | 16 | 158 | 19 |
| Baseline + treatment exposure | Random Forest | 2 | 0.952 | 0.249 | 0.803 | 0.549 | 0.130 | 0.995 | 0.211 | 0.042 | 4166 | 28 | 187 | 23 |
| Baseline + treatment exposure | Random Forest | 3 | 0.948 | 0.248 | 0.773 | 0.431 | 0.100 | 0.993 | 0.162 | 0.043 | 4115 | 22 | 199 | 29 |
| Baseline + treatment exposure | Random Forest | 4 | 0.957 | 0.237 | 0.816 | 0.417 | 0.078 | 0.995 | 0.132 | 0.037 | 4346 | 15 | 177 | 21 |
| Baseline + treatment exposure | Random Forest | 5 | 0.952 | 0.286 | 0.835 | 0.500 | 0.084 | 0.996 | 0.144 | 0.039 | 4016 | 17 | 185 | 17 |
| Baseline + treatment exposure | Gradient Boosting | 1 | 0.960 | 0.282 | 0.862 | 0.415 | 0.098 | 0.994 | 0.158 | 0.032 | 4337 | 17 | 157 | 24 |
| Baseline + treatment exposure | Gradient Boosting | 2 | 0.951 | 0.292 | 0.827 | 0.500 | 0.126 | 0.994 | 0.201 | 0.040 | 4162 | 27 | 188 | 27 |
| Baseline + treatment exposure | Gradient Boosting | 3 | 0.947 | 0.267 | 0.807 | 0.426 | 0.118 | 0.992 | 0.184 | 0.043 | 4109 | 26 | 195 | 35 |
| Baseline + treatment exposure | Gradient Boosting | 4 | 0.956 | 0.246 | 0.838 | 0.404 | 0.099 | 0.994 | 0.159 | 0.036 | 4339 | 19 | 173 | 28 |
| Baseline + treatment exposure | Gradient Boosting | 5 | 0.953 | 0.290 | 0.818 | 0.564 | 0.109 | 0.996 | 0.183 | 0.040 | 4016 | 22 | 180 | 17 |
| Baseline + treatment exposure | LightGBM | 1 | 0.961 | 0.271 | 0.841 | 0.432 | 0.092 | 0.995 | 0.152 | 0.032 | 4340 | 16 | 158 | 21 |
| Baseline + treatment exposure | LightGBM | 2 | 0.950 | 0.253 | 0.809 | 0.474 | 0.126 | 0.993 | 0.199 | 0.042 | 4159 | 27 | 188 | 30 |
| Baseline + treatment exposure | LightGBM | 3 | 0.949 | 0.257 | 0.809 | 0.473 | 0.118 | 0.993 | 0.188 | 0.044 | 4115 | 26 | 195 | 29 |
| Baseline + treatment exposure | LightGBM | 4 | 0.955 | 0.230 | 0.830 | 0.370 | 0.104 | 0.992 | 0.163 | 0.037 | 4333 | 20 | 172 | 34 |
| Baseline + treatment exposure | LightGBM | 5 | 0.951 | 0.271 | 0.834 | 0.452 | 0.094 | 0.994 | 0.156 | 0.040 | 4010 | 19 | 183 | 23 |
| Baseline + procedure burden | Logistic Regression | 1 | 0.967 | 0.403 | 0.865 | 0.745 | 0.218 | 0.997 | 0.338 | 0.029 | 4348 | 38 | 136 | 13 |
| Baseline + procedure burden | Logistic Regression | 2 | 0.953 | 0.326 | 0.823 | 0.609 | 0.130 | 0.996 | 0.215 | 0.039 | 4171 | 28 | 187 | 18 |
| Baseline + procedure burden | Logistic Regression | 3 | 0.951 | 0.283 | 0.786 | 0.578 | 0.118 | 0.995 | 0.195 | 0.042 | 4125 | 26 | 195 | 19 |
| Baseline + procedure burden | Logistic Regression | 4 | 0.958 | 0.287 | 0.832 | 0.508 | 0.161 | 0.993 | 0.245 | 0.035 | 4337 | 31 | 161 | 30 |
| Baseline + procedure burden | Logistic Regression | 5 | 0.957 | 0.315 | 0.792 | 0.723 | 0.168 | 0.997 | 0.273 | 0.039 | 4020 | 34 | 168 | 13 |
| Baseline + procedure burden | Random Forest | 1 | 0.963 | 0.320 | 0.825 | 0.562 | 0.207 | 0.994 | 0.303 | 0.031 | 4333 | 36 | 138 | 28 |
| Baseline + procedure burden | Random Forest | 2 | 0.956 | 0.289 | 0.802 | 0.656 | 0.195 | 0.995 | 0.301 | 0.041 | 4167 | 42 | 173 | 22 |
| Baseline + procedure burden | Random Forest | 3 | 0.951 | 0.305 | 0.775 | 0.561 | 0.167 | 0.993 | 0.258 | 0.042 | 4115 | 37 | 184 | 29 |
| Baseline + procedure burden | Random Forest | 4 | 0.959 | 0.302 | 0.813 | 0.537 | 0.188 | 0.993 | 0.278 | 0.036 | 4336 | 36 | 156 | 31 |
| Baseline + procedure burden | Random Forest | 5 | 0.953 | 0.295 | 0.827 | 0.529 | 0.134 | 0.994 | 0.213 | 0.040 | 4009 | 27 | 175 | 24 |
| Baseline + procedure burden | Gradient Boosting | 1 | 0.967 | 0.391 | 0.871 | 0.717 | 0.218 | 0.997 | 0.335 | 0.029 | 4346 | 38 | 136 | 15 |
| Baseline + procedure burden | Gradient Boosting | 2 | 0.953 | 0.330 | 0.836 | 0.557 | 0.158 | 0.994 | 0.246 | 0.039 | 4162 | 34 | 181 | 27 |
| Baseline + procedure burden | Gradient Boosting | 3 | 0.952 | 0.324 | 0.823 | 0.586 | 0.154 | 0.994 | 0.244 | 0.041 | 4120 | 34 | 187 | 24 |
| Baseline + procedure burden | Gradient Boosting | 4 | 0.959 | 0.333 | 0.851 | 0.521 | 0.198 | 0.992 | 0.287 | 0.034 | 4332 | 38 | 154 | 35 |

Continued on next page

eTable 7. CHoRUS performance metrics for all feature matrices and algorithms. (continued)

| Feature matrix | Model | Fold | Accuracy | AUPRC | AUROC | Precision | Sensitivity | Specificity | F1 | Brier score | TN | TP | FN | FP |
|---|---|---|---|---|---|---|---|---|---|---|---|---|---|---|
| Baseline + procedure burden | Gradient Boosting | 5 | 0.955 | 0.326 | 0.824 | 0.596 | 0.168 | 0.994 | 0.263 | 0.038 | 4010 | 34 | 168 | 23 |
| Baseline + procedure burden | LightGBM | 1 | 0.966 | 0.369 | 0.852 | 0.636 | 0.241 | 0.994 | 0.350 | 0.030 | 4337 | 42 | 132 | 24 |
| Baseline + procedure burden | LightGBM | 2 | 0.954 | 0.320 | 0.826 | 0.573 | 0.200 | 0.992 | 0.297 | 0.040 | 4157 | 43 | 172 | 32 |
| Baseline + procedure burden | LightGBM | 3 | 0.949 | 0.313 | 0.799 | 0.500 | 0.190 | 0.990 | 0.275 | 0.042 | 4102 | 42 | 179 | 42 |
| Baseline + procedure burden | LightGBM | 4 | 0.958 | 0.329 | 0.834 | 0.512 | 0.229 | 0.990 | 0.317 | 0.035 | 4325 | 44 | 148 | 42 |
| Baseline + procedure burden | LightGBM | 5 | 0.953 | 0.284 | 0.803 | 0.515 | 0.168 | 0.992 | 0.254 | 0.040 | 4001 | 34 | 168 | 32 |
| Baseline + physiological severity + treatment exposure | Logistic Regression | 1 | 0.962 | 0.363 | 0.867 | 0.525 | 0.184 | 0.993 | 0.272 | 0.029 | 4332 | 32 | 142 | 29 |
| Baseline + physiological severity + treatment exposure | Logistic Regression | 2 | 0.954 | 0.367 | 0.842 | 0.618 | 0.158 | 0.995 | 0.252 | 0.038 | 4168 | 34 | 181 | 21 |
| Baseline + physiological severity + treatment exposure | Logistic Regression | 3 | 0.951 | 0.302 | 0.833 | 0.547 | 0.131 | 0.994 | 0.212 | 0.042 | 4120 | 29 | 192 | 24 |
| Baseline + physiological severity + treatment exposure | Logistic Regression | 4 | 0.960 | 0.360 | 0.866 | 0.617 | 0.151 | 0.996 | 0.243 | 0.033 | 4349 | 29 | 163 | 18 |
| Baseline + physiological severity + treatment exposure | Logistic Regression | 5 | 0.954 | 0.306 | 0.817 | 0.574 | 0.134 | 0.995 | 0.217 | 0.039 | 4013 | 27 | 175 | 20 |
| Baseline + physiological severity + treatment exposure | Random Forest | 1 | 0.965 | 0.406 | 0.859 | 0.653 | 0.184 | 0.996 | 0.287 | 0.028 | 4344 | 32 | 142 | 17 |
| Baseline + physiological severity + treatment exposure | Random Forest | 2 | 0.957 | 0.354 | 0.833 | 0.714 | 0.209 | 0.996 | 0.324 | 0.038 | 4171 | 45 | 170 | 18 |
| Baseline + physiological severity + treatment exposure | Random Forest | 3 | 0.950 | 0.303 | 0.797 | 0.528 | 0.127 | 0.994 | 0.204 | 0.041 | 4119 | 28 | 193 | 25 |
| Baseline + physiological severity + treatment exposure | Random Forest | 4 | 0.962 | 0.375 | 0.846 | 0.656 | 0.208 | 0.995 | 0.316 | 0.033 | 4346 | 40 | 152 | 21 |
| Baseline + physiological severity + treatment exposure | Random Forest | 5 | 0.956 | 0.379 | 0.866 | 0.636 | 0.173 | 0.995 | 0.272 | 0.036 | 4013 | 35 | 167 | 20 |
| Baseline + physiological severity + treatment exposure | Gradient Boosting | 1 | 0.968 | 0.420 | 0.882 | 0.704 | 0.287 | 0.995 | 0.408 | 0.028 | 4340 | 50 | 124 | 21 |
| Baseline + physiological severity + treatment exposure | Gradient Boosting | 2 | 0.954 | 0.357 | 0.856 | 0.588 | 0.219 | 0.992 | 0.319 | 0.038 | 4156 | 47 | 168 | 33 |
| Baseline + physiological severity + treatment exposure | Gradient Boosting | 3 | 0.950 | 0.293 | 0.828 | 0.529 | 0.163 | 0.992 | 0.249 | 0.042 | 4112 | 36 | 185 | 32 |
| Baseline + physiological severity + treatment exposure | Gradient Boosting | 4 | 0.960 | 0.347 | 0.869 | 0.556 | 0.234 | 0.992 | 0.330 | 0.034 | 4331 | 45 | 147 | 36 |
| Baseline + physiological severity + treatment exposure | Gradient Boosting | 5 | 0.956 | 0.368 | 0.850 | 0.639 | 0.193 | 0.995 | 0.297 | 0.037 | 4011 | 39 | 163 | 22 |
| Baseline + physiological severity + treatment exposure | LightGBM | 1 | 0.965 | 0.433 | 0.867 | 0.616 | 0.259 | 0.994 | 0.364 | 0.028 | 4333 | 45 | 129 | 28 |
| Baseline + physiological severity + treatment exposure | LightGBM | 2 | 0.957 | 0.392 | 0.856 | 0.662 | 0.247 | 0.994 | 0.359 | 0.037 | 4162 | 53 | 162 | 27 |
| Baseline + physiological severity + treatment exposure | LightGBM | 3 | 0.949 | 0.298 | 0.839 | 0.482 | 0.181 | 0.990 | 0.263 | 0.042 | 4101 | 40 | 181 | 43 |
| Baseline + physiological severity + treatment exposure | LightGBM | 4 | 0.960 | 0.372 | 0.855 | 0.562 | 0.260 | 0.991 | 0.356 | 0.033 | 4328 | 50 | 142 | 39 |
| Baseline + physiological severity + treatment exposure | LightGBM | 5 | 0.957 | 0.368 | 0.851 | 0.641 | 0.203 | 0.994 | 0.308 | 0.037 | 4010 | 41 | 161 | 23 |
| Baseline + physiological severity + procedure burden | Logistic Regression | 1 | 0.966 | 0.391 | 0.872 | 0.643 | 0.259 | 0.994 | 0.369 | 0.029 | 4336 | 45 | 129 | 25 |

Continued on next page

eTable 7. CHoRUS performance metrics for all feature matrices and algorithms. (continued)

| Feature matrix | Model | Fold | Accuracy | AUPRC | AUROC | Precision | Sensitivity | Specificity | F1 | Brier score | TN | TP | FN | FP |
|---|---|---|---|---|---|---|---|---|---|---|---|---|---|---|
| Baseline + physiological severity + procedure burden | Logistic Regression | 2 | 0.954 | 0.354 | 0.837 | 0.597 | 0.172 | 0.994 | 0.267 | 0.039 | 4164 | 37 | 178 | 25 |
| Baseline + physiological severity + procedure burden | Logistic Regression | 3 | 0.952 | 0.317 | 0.839 | 0.611 | 0.149 | 0.995 | 0.240 | 0.041 | 4123 | 33 | 188 | 21 |
| Baseline + physiological severity + procedure burden | Logistic Regression | 4 | 0.960 | 0.334 | 0.866 | 0.574 | 0.182 | 0.994 | 0.277 | 0.034 | 4341 | 35 | 157 | 26 |
| Baseline + physiological severity + procedure burden | Logistic Regression | 5 | 0.955 | 0.301 | 0.819 | 0.611 | 0.163 | 0.995 | 0.258 | 0.039 | 4012 | 33 | 169 | 21 |
| Baseline + physiological severity + procedure burden | Random Forest | 1 | 0.965 | 0.403 | 0.844 | 0.630 | 0.195 | 0.995 | 0.298 | 0.029 | 4341 | 34 | 140 | 20 |
| Baseline + physiological severity + procedure burden | Random Forest | 2 | 0.956 | 0.354 | 0.827 | 0.677 | 0.195 | 0.995 | 0.303 | 0.038 | 4169 | 42 | 173 | 20 |
| Baseline + physiological severity + procedure burden | Random Forest | 3 | 0.950 | 0.317 | 0.799 | 0.516 | 0.145 | 0.993 | 0.226 | 0.041 | 4114 | 32 | 189 | 30 |
| Baseline + physiological severity + procedure burden | Random Forest | 4 | 0.962 | 0.376 | 0.835 | 0.643 | 0.234 | 0.994 | 0.344 | 0.033 | 4342 | 45 | 147 | 25 |
| Baseline + physiological severity + procedure burden | Random Forest | 5 | 0.954 | 0.360 | 0.856 | 0.571 | 0.158 | 0.994 | 0.248 | 0.037 | 4009 | 32 | 170 | 24 |
| Baseline + physiological severity + procedure burden | Gradient Boosting | 1 | 0.968 | 0.454 | 0.884 | 0.708 | 0.293 | 0.995 | 0.415 | 0.027 | 4340 | 51 | 123 | 21 |
| Baseline + physiological severity + procedure burden | Gradient Boosting | 2 | 0.958 | 0.404 | 0.853 | 0.710 | 0.228 | 0.995 | 0.345 | 0.037 | 4169 | 49 | 166 | 20 |
| Baseline + physiological severity + procedure burden | Gradient Boosting | 3 | 0.951 | 0.312 | 0.843 | 0.537 | 0.163 | 0.993 | 0.250 | 0.041 | 4113 | 36 | 185 | 31 |
| Baseline + physiological severity + procedure burden | Gradient Boosting | 4 | 0.961 | 0.380 | 0.878 | 0.581 | 0.260 | 0.992 | 0.360 | 0.032 | 4331 | 50 | 142 | 36 |
| Baseline + physiological severity + procedure burden | Gradient Boosting | 5 | 0.958 | 0.405 | 0.855 | 0.732 | 0.203 | 0.996 | 0.318 | 0.035 | 4018 | 41 | 161 | 15 |
| Baseline + physiological severity + procedure burden | LightGBM | 1 | 0.967 | 0.420 | 0.874 | 0.667 | 0.276 | 0.994 | 0.390 | 0.028 | 4337 | 48 | 126 | 24 |
| Baseline + physiological severity + procedure burden | LightGBM | 2 | 0.957 | 0.392 | 0.859 | 0.647 | 0.256 | 0.993 | 0.367 | 0.037 | 4159 | 55 | 160 | 30 |
| Baseline + physiological severity + procedure burden | LightGBM | 3 | 0.950 | 0.316 | 0.834 | 0.519 | 0.186 | 0.991 | 0.273 | 0.042 | 4106 | 41 | 180 | 38 |
| Baseline + physiological severity + procedure burden | LightGBM | 4 | 0.961 | 0.376 | 0.857 | 0.558 | 0.302 | 0.989 | 0.392 | 0.033 | 4321 | 58 | 134 | 46 |
| Baseline + physiological severity + procedure burden | LightGBM | 5 | 0.954 | 0.349 | 0.848 | 0.551 | 0.188 | 0.992 | 0.280 | 0.038 | 4002 | 38 | 164 | 31 |
| Baseline + treatment exposure + procedure burden | Logistic Regression | 1 | 0.966 | 0.386 | 0.869 | 0.694 | 0.195 | 0.997 | 0.305 | 0.029 | 4346 | 34 | 140 | 15 |
| Baseline + treatment exposure + procedure burden | Logistic Regression | 2 | 0.955 | 0.326 | 0.833 | 0.647 | 0.153 | 0.996 | 0.248 | 0.039 | 4171 | 33 | 182 | 18 |
| Baseline + treatment exposure + procedure burden | Logistic Regression | 3 | 0.952 | 0.281 | 0.792 | 0.593 | 0.145 | 0.995 | 0.233 | 0.042 | 4122 | 32 | 189 | 22 |
| Baseline + treatment exposure + procedure burden | Logistic Regression | 4 | 0.958 | 0.314 | 0.848 | 0.508 | 0.156 | 0.993 | 0.239 | 0.034 | 4338 | 30 | 162 | 29 |
| Baseline + treatment exposure + procedure burden | Logistic Regression | 5 | 0.957 | 0.335 | 0.805 | 0.696 | 0.158 | 0.997 | 0.258 | 0.038 | 4019 | 32 | 170 | 14 |

Continued on next page

eTable 7. CHoRUS performance metrics for all feature matrices and algorithms. (continued)

| Feature matrix | Model | Fold | Accuracy | AUPRC | AUROC | Precision | Sensitivity | Specificity | F1 | Brier score | TN | TP | FN | FP |
|---|---|---|---|---|---|---|---|---|---|---|---|---|---|---|
| Baseline + treatment exposure + procedure burden | Random Forest | 1 | 0.966 | 0.381 | 0.848 | 0.721 | 0.178 | 0.997 | 0.286 | 0.030 | 4349 | 31 | 143 | 12 |
| Baseline + treatment exposure + procedure burden | Random Forest | 2 | 0.955 | 0.303 | 0.824 | 0.667 | 0.158 | 0.996 | 0.256 | 0.040 | 4172 | 34 | 181 | 17 |
| Baseline + treatment exposure + procedure burden | Random Forest | 3 | 0.949 | 0.307 | 0.801 | 0.500 | 0.131 | 0.993 | 0.208 | 0.041 | 4115 | 29 | 192 | 29 |
| Baseline + treatment exposure + procedure burden | Random Forest | 4 | 0.960 | 0.327 | 0.837 | 0.604 | 0.151 | 0.996 | 0.242 | 0.034 | 4348 | 29 | 163 | 19 |
| Baseline + treatment exposure + procedure burden | Random Forest | 5 | 0.954 | 0.337 | 0.840 | 0.577 | 0.149 | 0.995 | 0.236 | 0.038 | 4011 | 30 | 172 | 22 |
| Baseline + treatment exposure + procedure burden | Gradient Boosting | 1 | 0.967 | 0.413 | 0.876 | 0.740 | 0.213 | 0.997 | 0.330 | 0.028 | 4348 | 37 | 137 | 13 |
| Baseline + treatment exposure + procedure burden | Gradient Boosting | 2 | 0.953 | 0.304 | 0.837 | 0.571 | 0.186 | 0.993 | 0.281 | 0.040 | 4159 | 40 | 175 | 30 |
| Baseline + treatment exposure + procedure burden | Gradient Boosting | 3 | 0.952 | 0.308 | 0.822 | 0.600 | 0.176 | 0.994 | 0.273 | 0.041 | 4118 | 39 | 182 | 26 |
| Baseline + treatment exposure + procedure burden | Gradient Boosting | 4 | 0.960 | 0.345 | 0.856 | 0.574 | 0.203 | 0.993 | 0.300 | 0.033 | 4338 | 39 | 153 | 29 |
| Baseline + treatment exposure + procedure burden | Gradient Boosting | 5 | 0.956 | 0.343 | 0.833 | 0.653 | 0.158 | 0.996 | 0.255 | 0.038 | 4016 | 32 | 170 | 17 |
| Baseline + treatment exposure + procedure burden | LightGBM | 1 | 0.966 | 0.401 | 0.857 | 0.661 | 0.236 | 0.995 | 0.347 | 0.028 | 4340 | 41 | 133 | 21 |
| Baseline + treatment exposure + procedure burden | LightGBM | 2 | 0.953 | 0.317 | 0.829 | 0.569 | 0.191 | 0.993 | 0.286 | 0.040 | 4158 | 41 | 174 | 31 |
| Baseline + treatment exposure + procedure burden | LightGBM | 3 | 0.949 | 0.317 | 0.824 | 0.480 | 0.163 | 0.991 | 0.243 | 0.042 | 4105 | 36 | 185 | 39 |
| Baseline + treatment exposure + procedure burden | LightGBM | 4 | 0.958 | 0.326 | 0.834 | 0.506 | 0.214 | 0.991 | 0.300 | 0.035 | 4327 | 41 | 151 | 40 |
| Baseline + treatment exposure + procedure burden | LightGBM | 5 | 0.955 | 0.319 | 0.830 | 0.578 | 0.183 | 0.993 | 0.278 | 0.039 | 4006 | 37 | 165 | 27 |
| Baseline + all clinical domains | Logistic Regression | 1 | 0.967 | 0.408 | 0.871 | 0.676 | 0.276 | 0.995 | 0.392 | 0.028 | 4338 | 48 | 126 | 23 |
| Baseline + all clinical domains | Logistic Regression | 2 | 0.953 | 0.367 | 0.846 | 0.571 | 0.186 | 0.993 | 0.281 | 0.038 | 4159 | 40 | 175 | 30 |
| Baseline + all clinical domains | Logistic Regression | 3 | 0.951 | 0.317 | 0.837 | 0.554 | 0.140 | 0.994 | 0.224 | 0.041 | 4119 | 31 | 190 | 25 |
| Baseline + all clinical domains | Logistic Regression | 4 | 0.961 | 0.364 | 0.874 | 0.594 | 0.198 | 0.994 | 0.297 | 0.033 | 4341 | 38 | 154 | 26 |
| Baseline + all clinical domains | Logistic Regression | 5 | 0.955 | 0.331 | 0.825 | 0.614 | 0.173 | 0.995 | 0.270 | 0.038 | 4011 | 35 | 167 | 22 |
| Baseline + all clinical domains | Random Forest | 1 | 0.967 | 0.437 | 0.863 | 0.709 | 0.224 | 0.996 | 0.341 | 0.028 | 4345 | 39 | 135 | 16 |
| Baseline + all clinical domains | Random Forest | 2 | 0.958 | 0.361 | 0.839 | 0.733 | 0.205 | 0.996 | 0.320 | 0.038 | 4173 | 44 | 171 | 16 |
| Baseline + all clinical domains | Random Forest | 3 | 0.950 | 0.307 | 0.795 | 0.533 | 0.145 | 0.993 | 0.228 | 0.041 | 4116 | 32 | 189 | 28 |
| Baseline + all clinical domains | Random Forest | 4 | 0.964 | 0.391 | 0.849 | 0.724 | 0.219 | 0.996 | 0.336 | 0.032 | 4351 | 42 | 150 | 16 |
| Baseline + all clinical domains | Random Forest | 5 | 0.955 | 0.383 | 0.864 | 0.611 | 0.163 | 0.995 | 0.258 | 0.036 | 4012 | 33 | 169 | 21 |
| Baseline + all clinical domains | Gradient Boosting | 1 | 0.969 | 0.444 | 0.885 | 0.758 | 0.287 | 0.996 | 0.417 | 0.027 | 4345 | 50 | 124 | 16 |
| Baseline + all clinical domains | Gradient Boosting | 2 | 0.958 | 0.398 | 0.853 | 0.703 | 0.242 | 0.995 | 0.360 | 0.037 | 4167 | 52 | 163 | 22 |
| Baseline + all clinical domains | Gradient Boosting | 3 | 0.948 | 0.284 | 0.830 | 0.463 | 0.167 | 0.990 | 0.246 | 0.043 | 4101 | 37 | 184 | 43 |

Continued on next page

eTable 7. CHoRUS performance metrics for all feature matrices and algorithms. (continued)

| Feature matrix | Model | Fold | Accuracy | AUPRC | AUROC | Precision | Sensitivity | Specificity | F1 | Brier score | TN | TP | FN | FP |
|---|---|---|---|---|---|---|---|---|---|---|---|---|---|---|
| Baseline + all clinical domains | Gradient Boosting | 4 | 0.959 | 0.373 | 0.872 | 0.543 | 0.229 | 0.992 | 0.322 | 0.033 | 4330 | 44 | 148 | 37 |
| Baseline + all clinical domains | Gradient Boosting | 5 | 0.958 | 0.409 | 0.855 | 0.719 | 0.203 | 0.996 | 0.317 | 0.035 | 4017 | 41 | 161 | 16 |
| Baseline + all clinical domains | LightGBM | 1 | 0.969 | 0.453 | 0.874 | 0.714 | 0.316 | 0.995 | 0.438 | 0.027 | 4339 | 55 | 119 | 22 |
| Baseline + all clinical domains | LightGBM | 2 | 0.957 | 0.387 | 0.853 | 0.643 | 0.251 | 0.993 | 0.361 | 0.037 | 4159 | 54 | 161 | 30 |
| Baseline + all clinical domains | LightGBM | 3 | 0.950 | 0.320 | 0.839 | 0.511 | 0.204 | 0.990 | 0.291 | 0.042 | 4101 | 45 | 176 | 43 |
| Baseline + all clinical domains | LightGBM | 4 | 0.960 | 0.369 | 0.856 | 0.542 | 0.271 | 0.990 | 0.361 | 0.033 | 4323 | 52 | 140 | 44 |
| Baseline + all clinical domains | LightGBM | 5 | 0.957 | 0.376 | 0.854 | 0.641 | 0.203 | 0.994 | 0.308 | 0.037 | 4010 | 41 | 161 | 23 |

*Notes:* AUPRC = area under the precision-recall curve; AUROC = area under the receiver operating characteristic curve; TN = true negatives; TP = true positives; FN = false negatives; FP = false positives. CHoRUS rows are validation-fold results from patient-grouped cross-validation outputs for each feature matrix and algorithm.

eTable 8. MIMIC-IV performance metrics for all feature matrices and algorithms.

| Feature matrix | Model | Fold | Accuracy | AUPRC | AUROC | Precision | Sensitivity | Specificity | F1 | Brier score | TN | TP | FN | FP |
|---|---|---|---|---|---|---|---|---|---|---|---|---|---|---|
| Baseline | Logistic Regression | 1 | 0.967 | 0.117 | 0.789 | 0.000 | 0.000 | 1.000 | 0.000 | 0.031 | 4368 | 0 | 151 | 0 |
| Baseline | Logistic Regression | 2 | 0.962 | 0.151 | 0.835 | 1.000 | 0.006 | 1.000 | 0.011 | 0.034 | 4584 | 1 | 179 | 0 |
| Baseline | Logistic Regression | 3 | 0.964 | 0.149 | 0.810 | 0.000 | 0.000 | 1.000 | 0.000 | 0.033 | 4260 | 0 | 160 | 0 |
| Baseline | Logistic Regression | 4 | 0.965 | 0.131 | 0.817 | 0.000 | 0.000 | 1.000 | 0.000 | 0.032 | 4335 | 0 | 156 | 0 |
| Baseline | Logistic Regression | 5 | 0.964 | 0.114 | 0.805 | 0.000 | 0.000 | 1.000 | 0.000 | 0.033 | 4633 | 0 | 172 | 1 |
| Baseline | Gradient Boosting | 1 | 0.966 | 0.112 | 0.784 | 0.286 | 0.013 | 0.999 | 0.025 | 0.032 | 4363 | 2 | 149 | 5 |
| Baseline | Gradient Boosting | 2 | 0.962 | 0.132 | 0.819 | 0.375 | 0.017 | 0.999 | 0.032 | 0.035 | 4579 | 3 | 177 | 5 |
| Baseline | Gradient Boosting | 3 | 0.963 | 0.111 | 0.810 | 0.000 | 0.000 | 0.999 | 0.000 | 0.034 | 4255 | 0 | 160 | 5 |
| Baseline | Gradient Boosting | 4 | 0.966 | 0.140 | 0.821 | 1.000 | 0.013 | 1.000 | 0.025 | 0.032 | 4335 | 2 | 154 | 0 |
| Baseline | Gradient Boosting | 5 | 0.964 | 0.116 | 0.807 | 0.333 | 0.006 | 1.000 | 0.011 | 0.033 | 4632 | 1 | 171 | 2 |
| Baseline | LightGBM | 1 | 0.964 | 0.090 | 0.772 | 0.188 | 0.020 | 0.997 | 0.036 | 0.033 | 4355 | 3 | 148 | 13 |
| Baseline | LightGBM | 2 | 0.955 | 0.108 | 0.808 | 0.051 | 0.011 | 0.992 | 0.018 | 0.038 | 4547 | 2 | 178 | 37 |
| Baseline | LightGBM | 3 | 0.963 | 0.098 | 0.791 | 0.000 | 0.000 | 0.999 | 0.000 | 0.035 | 4257 | 0 | 160 | 3 |
| Baseline | LightGBM | 4 | 0.966 | 0.136 | 0.802 | 0.750 | 0.019 | 1.000 | 0.037 | 0.032 | 4334 | 3 | 153 | 1 |
| Baseline | LightGBM | 5 | 0.963 | 0.100 | 0.789 | 0.231 | 0.017 | 0.998 | 0.032 | 0.035 | 4624 | 3 | 169 | 10 |
| Baseline | Random Forest | 1 | 0.966 | 0.089 | 0.768 | 0.000 | 0.000 | 0.999 | 0.000 | 0.033 | 4365 | 0 | 151 | 3 |
| Baseline | Random Forest | 2 | 0.962 | 0.111 | 0.780 | 0.000 | 0.000 | 1.000 | 0.000 | 0.036 | 4583 | 0 | 180 | 1 |
| Baseline | Random Forest | 3 | 0.964 | 0.095 | 0.777 | 0.400 | 0.013 | 0.999 | 0.024 | 0.035 | 4257 | 2 | 158 | 3 |
| Baseline | Random Forest | 4 | 0.965 | 0.104 | 0.790 | 0.000 | 0.000 | 1.000 | 0.000 | 0.033 | 4335 | 0 | 156 | 0 |
| Baseline | Random Forest | 5 | 0.963 | 0.084 | 0.755 | 0.167 | 0.006 | 0.999 | 0.011 | 0.036 | 4629 | 1 | 171 | 5 |
| Baseline + physiological severity | Random Forest | 1 | 0.968 | 0.239 | 0.815 | 1.000 | 0.040 | 1.000 | 0.076 | 0.030 | 4368 | 6 | 145 | 0 |
| Baseline + physiological severity | Random Forest | 2 | 0.962 | 0.286 | 0.877 | 0.667 | 0.011 | 1.000 | 0.022 | 0.031 | 4583 | 2 | 178 | 1 |
| Baseline + physiological severity | Random Forest | 3 | 0.964 | 0.255 | 0.869 | 0.750 | 0.019 | 1.000 | 0.037 | 0.031 | 4259 | 3 | 157 | 1 |
| Baseline + physiological severity | Random Forest | 4 | 0.965 | 0.223 | 0.885 | 0.500 | 0.013 | 1.000 | 0.025 | 0.030 | 4333 | 2 | 154 | 2 |
| Baseline + physiological severity | Random Forest | 5 | 0.964 | 0.206 | 0.850 | 0.500 | 0.023 | 0.999 | 0.044 | 0.031 | 4630 | 4 | 168 | 4 |
| Baseline + physiological severity | Gradient Boosting | 1 | 0.967 | 0.233 | 0.824 | 0.533 | 0.106 | 0.997 | 0.177 | 0.029 | 4354 | 16 | 135 | 14 |
| Baseline + physiological severity | Gradient Boosting | 2 | 0.963 | 0.284 | 0.887 | 0.586 | 0.094 | 0.997 | 0.163 | 0.031 | 4572 | 17 | 163 | 12 |
| Baseline + physiological severity | Gradient Boosting | 3 | 0.963 | 0.215 | 0.874 | 0.389 | 0.044 | 0.997 | 0.079 | 0.032 | 4249 | 7 | 153 | 11 |
| Baseline + physiological severity | Gradient Boosting | 4 | 0.965 | 0.236 | 0.885 | 0.480 | 0.077 | 0.997 | 0.133 | 0.030 | 4322 | 12 | 144 | 13 |

Continued on next page

eTable 8. MIMIC-IV performance metrics for all feature matrices and algorithms. (continued)

| Feature matrix | Model | Fold | Accuracy | AUPRC | AUROC | Precision | Sensitivity | Specificity | F1 | Brier score | TN | TP | FN | FP |
|---|---|---|---|---|---|---|---|---|---|---|---|---|---|---|
| Baseline + physiological severity | Gradient Boosting | 5 | 0.963 | 0.206 | 0.862 | 0.423 | 0.064 | 0.997 | 0.111 | 0.032 | 4619 | 11 | 161 | 15 |
| Baseline + physiological severity | Logistic Regression | 1 | 0.969 | 0.231 | 0.824 | 0.778 | 0.093 | 0.999 | 0.166 | 0.029 | 4364 | 14 | 137 | 4 |
| Baseline + physiological severity | Logistic Regression | 2 | 0.963 | 0.276 | 0.886 | 0.571 | 0.044 | 0.999 | 0.082 | 0.031 | 4578 | 8 | 172 | 6 |
| Baseline + physiological severity | Logistic Regression | 3 | 0.964 | 0.219 | 0.868 | 0.455 | 0.031 | 0.999 | 0.058 | 0.032 | 4254 | 5 | 155 | 6 |
| Baseline + physiological severity | Logistic Regression | 4 | 0.965 | 0.202 | 0.861 | 0.400 | 0.038 | 0.998 | 0.070 | 0.031 | 4326 | 6 | 150 | 9 |
| Baseline + physiological severity | Logistic Regression | 5 | 0.964 | 0.171 | 0.850 | 0.400 | 0.023 | 0.999 | 0.044 | 0.032 | 4628 | 4 | 168 | 6 |
| Baseline + physiological severity | LightGBM | 1 | 0.967 | 0.208 | 0.811 | 0.500 | 0.086 | 0.997 | 0.147 | 0.030 | 4355 | 13 | 138 | 13 |
| Baseline + physiological severity | LightGBM | 2 | 0.962 | 0.209 | 0.866 | 0.500 | 0.094 | 0.996 | 0.159 | 0.034 | 4567 | 17 | 163 | 17 |
| Baseline + physiological severity | LightGBM | 3 | 0.962 | 0.235 | 0.873 | 0.393 | 0.069 | 0.996 | 0.117 | 0.032 | 4243 | 11 | 149 | 17 |
| Baseline + physiological severity | LightGBM | 4 | 0.964 | 0.193 | 0.860 | 0.348 | 0.051 | 0.997 | 0.089 | 0.031 | 4320 | 8 | 148 | 15 |
| Baseline + physiological severity | LightGBM | 5 | 0.962 | 0.186 | 0.857 | 0.290 | 0.052 | 0.995 | 0.089 | 0.033 | 4612 | 9 | 163 | 22 |
| Baseline + treatment exposure | Logistic Regression | 1 | 0.967 | 0.184 | 0.823 | 0.500 | 0.053 | 0.998 | 0.096 | 0.030 | 4360 | 8 | 143 | 8 |
| Baseline + treatment exposure | Logistic Regression | 2 | 0.963 | 0.248 | 0.873 | 0.692 | 0.050 | 0.999 | 0.093 | 0.032 | 4580 | 9 | 171 | 4 |
| Baseline + treatment exposure | Logistic Regression | 3 | 0.964 | 0.259 | 0.858 | 0.615 | 0.050 | 0.999 | 0.092 | 0.030 | 4255 | 8 | 152 | 5 |
| Baseline + treatment exposure | Logistic Regression | 4 | 0.965 | 0.199 | 0.847 | 0.417 | 0.032 | 0.998 | 0.060 | 0.031 | 4328 | 5 | 151 | 7 |
| Baseline + treatment exposure | Logistic Regression | 5 | 0.964 | 0.200 | 0.838 | 0.500 | 0.070 | 0.997 | 0.122 | 0.032 | 4622 | 12 | 160 | 12 |
| Baseline + treatment exposure | Random Forest | 1 | 0.967 | 0.179 | 0.825 | 0.500 | 0.013 | 1.000 | 0.026 | 0.030 | 4366 | 2 | 149 | 2 |
| Baseline + treatment exposure | Random Forest | 2 | 0.963 | 0.227 | 0.847 | 0.714 | 0.028 | 1.000 | 0.053 | 0.033 | 4582 | 5 | 175 | 2 |
| Baseline + treatment exposure | Random Forest | 3 | 0.964 | 0.255 | 0.848 | 0.500 | 0.006 | 1.000 | 0.012 | 0.031 | 4259 | 1 | 159 | 1 |
| Baseline + treatment exposure | Random Forest | 4 | 0.966 | 0.203 | 0.847 | 1.000 | 0.013 | 1.000 | 0.025 | 0.030 | 4335 | 2 | 154 | 0 |
| Baseline + treatment exposure | Random Forest | 5 | 0.965 | 0.190 | 0.813 | 0.800 | 0.023 | 1.000 | 0.045 | 0.032 | 4633 | 4 | 168 | 1 |
| Baseline + treatment exposure | Gradient Boosting | 1 | 0.965 | 0.165 | 0.832 | 0.316 | 0.040 | 0.997 | 0.071 | 0.031 | 4355 | 6 | 145 | 13 |
| Baseline + treatment exposure | Gradient Boosting | 2 | 0.954 | 0.161 | 0.854 | 0.188 | 0.067 | 0.989 | 0.098 | 0.038 | 4532 | 12 | 168 | 52 |
| Baseline + treatment exposure | Gradient Boosting | 3 | 0.965 | 0.256 | 0.866 | 0.750 | 0.056 | 0.999 | 0.105 | 0.031 | 4257 | 9 | 151 | 3 |
| Baseline + treatment exposure | Gradient Boosting | 4 | 0.965 | 0.207 | 0.851 | 0.556 | 0.032 | 0.999 | 0.061 | 0.030 | 4331 | 5 | 151 | 4 |
| Baseline + treatment exposure | Gradient Boosting | 5 | 0.962 | 0.170 | 0.838 | 0.345 | 0.058 | 0.996 | 0.100 | 0.033 | 4615 | 10 | 162 | 19 |
| Baseline + treatment exposure | LightGBM | 1 | 0.965 | 0.165 | 0.807 | 0.320 | 0.053 | 0.996 | 0.091 | 0.031 | 4351 | 8 | 143 | 17 |
| Baseline + treatment exposure | LightGBM | 2 | 0.961 | 0.179 | 0.837 | 0.409 | 0.050 | 0.997 | 0.089 | 0.034 | 4571 | 9 | 171 | 13 |
| Baseline + treatment exposure | LightGBM | 3 | 0.965 | 0.203 | 0.840 | 0.700 | 0.044 | 0.999 | 0.082 | 0.032 | 4257 | 7 | 153 | 3 |
| Baseline + treatment exposure | LightGBM | 4 | 0.965 | 0.182 | 0.827 | 0.471 | 0.051 | 0.998 | 0.092 | 0.031 | 4326 | 8 | 148 | 9 |
| Baseline + treatment exposure | LightGBM | 5 | 0.962 | 0.170 | 0.833 | 0.355 | 0.064 | 0.996 | 0.108 | 0.033 | 4614 | 11 | 161 | 20 |
| Baseline + procedure burden | Logistic Regression | 1 | 0.966 | 0.181 | 0.823 | 0.429 | 0.060 | 0.997 | 0.105 | 0.030 | 4356 | 9 | 142 | 12 |
| Baseline + procedure burden | Logistic Regression | 2 | 0.962 | 0.210 | 0.861 | 0.500 | 0.039 | 0.998 | 0.072 | 0.033 | 4577 | 7 | 173 | 7 |
| Baseline + procedure burden | Logistic Regression | 3 | 0.965 | 0.236 | 0.839 | 0.733 | 0.069 | 0.999 | 0.126 | 0.031 | 4256 | 11 | 149 | 4 |

Continued on next page

eTable 8. MIMIC-IV performance metrics for all feature matrices and algorithms. (continued)

| Feature matrix | Model | Fold | Accuracy | AUPRC | AUROC | Precision | Sensitivity | Specificity | F1 | Brier score | TN | TP | FN | FP |
|---|---|---|---|---|---|---|---|---|---|---|---|---|---|---|
| Baseline + procedure burden | Logistic Regression | 4 | 0.963 | 0.175 | 0.838 | 0.200 | 0.019 | 0.997 | 0.035 | 0.031 | 4323 | 3 | 153 | 12 |
| Baseline + procedure burden | Logistic Regression | 5 | 0.964 | 0.176 | 0.829 | 0.444 | 0.047 | 0.998 | 0.084 | 0.032 | 4624 | 8 | 164 | 10 |
| Baseline + procedure burden | Gradient Boosting | 1 | 0.965 | 0.175 | 0.822 | 0.320 | 0.053 | 0.996 | 0.091 | 0.031 | 4351 | 8 | 143 | 17 |
| Baseline + procedure burden | Gradient Boosting | 2 | 0.955 | 0.160 | 0.841 | 0.179 | 0.056 | 0.990 | 0.085 | 0.038 | 4538 | 10 | 170 | 46 |
| Baseline + procedure burden | Gradient Boosting | 3 | 0.963 | 0.196 | 0.840 | 0.438 | 0.044 | 0.998 | 0.080 | 0.032 | 4251 | 7 | 153 | 9 |
| Baseline + procedure burden | Gradient Boosting | 4 | 0.964 | 0.177 | 0.846 | 0.273 | 0.019 | 0.998 | 0.036 | 0.031 | 4327 | 3 | 153 | 8 |
| Baseline + procedure burden | Gradient Boosting | 5 | 0.962 | 0.149 | 0.829 | 0.308 | 0.047 | 0.996 | 0.081 | 0.033 | 4616 | 8 | 164 | 18 |
| Baseline + procedure burden | Random Forest | 1 | 0.966 | 0.151 | 0.812 | 0.375 | 0.020 | 0.999 | 0.038 | 0.031 | 4363 | 3 | 148 | 5 |
| Baseline + procedure burden | Random Forest | 2 | 0.962 | 0.171 | 0.823 | 0.375 | 0.017 | 0.999 | 0.032 | 0.034 | 4579 | 3 | 177 | 5 |
| Baseline + procedure burden | Random Forest | 3 | 0.964 | 0.171 | 0.819 | 1.000 | 0.006 | 1.000 | 0.012 | 0.033 | 4260 | 1 | 159 | 0 |
| Baseline + procedure burden | Random Forest | 4 | 0.965 | 0.150 | 0.842 | 0.000 | 0.000 | 1.000 | 0.000 | 0.032 | 4334 | 0 | 156 | 1 |
| Baseline + procedure burden | Random Forest | 5 | 0.964 | 0.136 | 0.797 | 0.286 | 0.012 | 0.999 | 0.022 | 0.033 | 4629 | 2 | 170 | 5 |
| Baseline + procedure burden | LightGBM | 1 | 0.963 | 0.129 | 0.796 | 0.273 | 0.060 | 0.995 | 0.098 | 0.033 | 4344 | 9 | 142 | 24 |
| Baseline + procedure burden | LightGBM | 2 | 0.961 | 0.163 | 0.834 | 0.391 | 0.050 | 0.997 | 0.089 | 0.035 | 4570 | 9 | 171 | 14 |
| Baseline + procedure burden | LightGBM | 3 | 0.963 | 0.159 | 0.818 | 0.412 | 0.044 | 0.998 | 0.079 | 0.033 | 4250 | 7 | 153 | 10 |
| Baseline + procedure burden | LightGBM | 4 | 0.965 | 0.155 | 0.815 | 0.444 | 0.026 | 0.999 | 0.048 | 0.032 | 4330 | 4 | 152 | 5 |
| Baseline + procedure burden | LightGBM | 5 | 0.962 | 0.136 | 0.811 | 0.286 | 0.035 | 0.997 | 0.062 | 0.034 | 4619 | 6 | 166 | 15 |
| Baseline + physiological severity + treatment exposure | Random Forest | 1 | 0.967 | 0.232 | 0.829 | 0.600 | 0.020 | 1.000 | 0.038 | 0.029 | 4366 | 3 | 148 | 2 |
| Baseline + physiological severity + treatment exposure | Random Forest | 2 | 0.963 | 0.294 | 0.879 | 0.750 | 0.017 | 1.000 | 0.033 | 0.031 | 4583 | 3 | 177 | 1 |
| Baseline + physiological severity + treatment exposure | Random Forest | 3 | 0.964 | 0.268 | 0.872 | 0.800 | 0.025 | 1.000 | 0.048 | 0.030 | 4259 | 4 | 156 | 1 |
| Baseline + physiological severity + treatment exposure | Random Forest | 4 | 0.965 | 0.225 | 0.878 | 0.600 | 0.019 | 1.000 | 0.037 | 0.030 | 4333 | 3 | 153 | 2 |
| Baseline + physiological severity + treatment exposure | Random Forest | 5 | 0.965 | 0.215 | 0.851 | 0.714 | 0.029 | 1.000 | 0.056 | 0.031 | 4632 | 5 | 167 | 2 |
| Baseline + physiological severity + treatment exposure | Logistic Regression | 1 | 0.967 | 0.231 | 0.835 | 0.571 | 0.106 | 0.997 | 0.179 | 0.029 | 4356 | 16 | 135 | 12 |
| Baseline + physiological severity + treatment exposure | Logistic Regression | 2 | 0.963 | 0.301 | 0.893 | 0.609 | 0.078 | 0.998 | 0.138 | 0.031 | 4575 | 14 | 166 | 9 |
| Baseline + physiological severity + treatment exposure | Logistic Regression | 3 | 0.964 | 0.252 | 0.881 | 0.476 | 0.062 | 0.997 | 0.110 | 0.031 | 4249 | 10 | 150 | 11 |
| Baseline + physiological severity + treatment exposure | Logistic Regression | 4 | 0.964 | 0.216 | 0.868 | 0.316 | 0.038 | 0.997 | 0.069 | 0.030 | 4322 | 6 | 150 | 13 |
| Baseline + physiological severity + treatment exposure | Logistic Regression | 5 | 0.964 | 0.231 | 0.863 | 0.500 | 0.052 | 0.998 | 0.095 | 0.031 | 4625 | 9 | 163 | 9 |
| Baseline + physiological severity + treatment exposure | Gradient Boosting | 1 | 0.967 | 0.227 | 0.834 | 0.514 | 0.126 | 0.996 | 0.202 | 0.030 | 4350 | 19 | 132 | 18 |
| Baseline + physiological severity + treatment exposure | Gradient Boosting | 2 | 0.954 | 0.218 | 0.887 | 0.253 | 0.106 | 0.988 | 0.149 | 0.035 | 4528 | 19 | 161 | 56 |
| Baseline + physiological severity + treatment exposure | Gradient Boosting | 3 | 0.963 | 0.236 | 0.881 | 0.429 | 0.056 | 0.997 | 0.099 | 0.031 | 4248 | 9 | 151 | 12 |
| Baseline + physiological severity + treatment exposure | Gradient Boosting | 4 | 0.964 | 0.223 | 0.885 | 0.375 | 0.058 | 0.997 | 0.100 | 0.030 | 4320 | 9 | 147 | 15 |

Continued on next page

eTable 8. MIMIC-IV performance metrics for all feature matrices and algorithms. (continued)

| Feature matrix | Model | Fold | Accuracy | AUPRC | AUROC | Precision | Sensitivity | Specificity | F1 | Brier score | TN | TP | FN | FP |
|---|---|---|---|---|---|---|---|---|---|---|---|---|---|---|
| Baseline + physiological severity + treatment exposure | Gradient Boosting | 5 | 0.962 | 0.217 | 0.864 | 0.345 | 0.058 | 0.996 | 0.100 | 0.032 | 4615 | 10 | 162 | 19 |
| Baseline + physiological severity + treatment exposure | LightGBM | 1 | 0.967 | 0.219 | 0.821 | 0.520 | 0.086 | 0.997 | 0.148 | 0.030 | 4356 | 13 | 138 | 12 |
| Baseline + physiological severity + treatment exposure | LightGBM | 2 | 0.963 | 0.240 | 0.874 | 0.552 | 0.089 | 0.997 | 0.153 | 0.033 | 4571 | 16 | 164 | 13 |
| Baseline + physiological severity + treatment exposure | LightGBM | 3 | 0.963 | 0.248 | 0.879 | 0.400 | 0.050 | 0.997 | 0.089 | 0.031 | 4248 | 8 | 152 | 12 |
| Baseline + physiological severity + treatment exposure | LightGBM | 4 | 0.964 | 0.201 | 0.860 | 0.368 | 0.045 | 0.997 | 0.080 | 0.031 | 4323 | 7 | 149 | 12 |
| Baseline + physiological severity + treatment exposure | LightGBM | 5 | 0.962 | 0.209 | 0.863 | 0.273 | 0.035 | 0.997 | 0.062 | 0.032 | 4618 | 6 | 166 | 16 |
| Baseline + physiological severity + procedure burden | Random Forest | 1 | 0.968 | 0.236 | 0.823 | 1.000 | 0.033 | 1.000 | 0.064 | 0.029 | 4368 | 5 | 146 | 0 |
| Baseline + physiological severity + procedure burden | Random Forest | 2 | 0.962 | 0.284 | 0.884 | 0.333 | 0.006 | 1.000 | 0.011 | 0.031 | 4582 | 1 | 179 | 2 |
| Baseline + physiological severity + procedure burden | Random Forest | 3 | 0.964 | 0.258 | 0.867 | 0.600 | 0.019 | 1.000 | 0.036 | 0.031 | 4258 | 3 | 157 | 2 |
| Baseline + physiological severity + procedure burden | Random Forest | 4 | 0.965 | 0.219 | 0.885 | 0.500 | 0.013 | 1.000 | 0.025 | 0.030 | 4333 | 2 | 154 | 2 |
| Baseline + physiological severity + procedure burden | Random Forest | 5 | 0.963 | 0.211 | 0.856 | 0.300 | 0.017 | 0.998 | 0.033 | 0.031 | 4627 | 3 | 169 | 7 |
| Baseline + physiological severity + procedure burden | Logistic Regression | 1 | 0.967 | 0.243 | 0.841 | 0.529 | 0.119 | 0.996 | 0.195 | 0.029 | 4352 | 18 | 133 | 16 |
| Baseline + physiological severity + procedure burden | Logistic Regression | 2 | 0.962 | 0.283 | 0.895 | 0.520 | 0.072 | 0.997 | 0.127 | 0.031 | 4572 | 13 | 167 | 12 |
| Baseline + physiological severity + procedure burden | Logistic Regression | 3 | 0.965 | 0.253 | 0.870 | 0.583 | 0.087 | 0.998 | 0.152 | 0.031 | 4250 | 14 | 146 | 10 |
| Baseline + physiological severity + procedure burden | Logistic Regression | 4 | 0.964 | 0.204 | 0.867 | 0.364 | 0.051 | 0.997 | 0.090 | 0.031 | 4321 | 8 | 148 | 14 |
| Baseline + physiological severity + procedure burden | Logistic Regression | 5 | 0.964 | 0.210 | 0.862 | 0.412 | 0.041 | 0.998 | 0.074 | 0.031 | 4624 | 7 | 165 | 10 |
| Baseline + physiological severity + procedure burden | Gradient Boosting | 1 | 0.966 | 0.241 | 0.830 | 0.486 | 0.119 | 0.996 | 0.191 | 0.029 | 4349 | 18 | 133 | 19 |
| Baseline + physiological severity + procedure burden | Gradient Boosting | 2 | 0.962 | 0.283 | 0.886 | 0.500 | 0.078 | 0.997 | 0.135 | 0.031 | 4570 | 14 | 166 | 14 |
| Baseline + physiological severity + procedure burden | Gradient Boosting | 3 | 0.964 | 0.237 | 0.876 | 0.565 | 0.081 | 0.998 | 0.142 | 0.031 | 4250 | 13 | 147 | 10 |
| Baseline + physiological severity + procedure burden | Gradient Boosting | 4 | 0.963 | 0.221 | 0.887 | 0.343 | 0.077 | 0.995 | 0.126 | 0.031 | 4312 | 12 | 144 | 23 |
| Baseline + physiological severity + procedure burden | Gradient Boosting | 5 | 0.963 | 0.210 | 0.864 | 0.357 | 0.058 | 0.996 | 0.100 | 0.032 | 4616 | 10 | 162 | 18 |
| Baseline + physiological severity + procedure burden | LightGBM | 1 | 0.967 | 0.223 | 0.822 | 0.556 | 0.099 | 0.997 | 0.169 | 0.029 | 4356 | 15 | 136 | 12 |
| Baseline + physiological severity + procedure burden | LightGBM | 2 | 0.955 | 0.186 | 0.873 | 0.242 | 0.089 | 0.989 | 0.130 | 0.038 | 4534 | 16 | 164 | 50 |
| Baseline + physiological severity + procedure burden | LightGBM | 3 | 0.963 | 0.246 | 0.872 | 0.464 | 0.081 | 0.996 | 0.138 | 0.031 | 4245 | 13 | 147 | 15 |

Continued on next page

eTable 8. MIMIC-IV performance metrics for all feature matrices and algorithms. (continued)

| Feature matrix | Model | Fold | Accuracy | AUPRC | AUROC | Precision | Sensitivity | Specificity | F1 | Brier score | TN | TP | FN | FP |
|---|---|---|---|---|---|---|---|---|---|---|---|---|---|---|
| Baseline + physiological severity + procedure burden | LightGBM | 4 | 0.965 | 0.200 | 0.860 | 0.421 | 0.051 | 0.997 | 0.091 | 0.031 | 4324 | 8 | 148 | 11 |
| Baseline + physiological severity + procedure burden | LightGBM | 5 | 0.962 | 0.185 | 0.855 | 0.276 | 0.047 | 0.995 | 0.080 | 0.033 | 4613 | 8 | 164 | 21 |
| Baseline + treatment exposure + procedure burden | Logistic Regression | 1 | 0.966 | 0.206 | 0.831 | 0.444 | 0.079 | 0.997 | 0.135 | 0.030 | 4353 | 12 | 139 | 15 |
| Baseline + treatment exposure + procedure burden | Logistic Regression | 2 | 0.962 | 0.251 | 0.879 | 0.500 | 0.072 | 0.997 | 0.126 | 0.032 | 4571 | 13 | 167 | 13 |
| Baseline + treatment exposure + procedure burden | Logistic Regression | 3 | 0.966 | 0.283 | 0.864 | 0.778 | 0.087 | 0.999 | 0.157 | 0.030 | 4256 | 14 | 146 | 4 |
| Baseline + treatment exposure + procedure burden | Logistic Regression | 4 | 0.964 | 0.211 | 0.848 | 0.368 | 0.045 | 0.997 | 0.080 | 0.030 | 4323 | 7 | 149 | 12 |
| Baseline + treatment exposure + procedure burden | Logistic Regression | 5 | 0.964 | 0.212 | 0.841 | 0.452 | 0.081 | 0.996 | 0.138 | 0.031 | 4617 | 14 | 158 | 17 |
| Baseline + treatment exposure + procedure burden | Random Forest | 1 | 0.967 | 0.190 | 0.828 | 0.750 | 0.020 | 1.000 | 0.039 | 0.030 | 4367 | 3 | 148 | 1 |
| Baseline + treatment exposure + procedure burden | Random Forest | 2 | 0.962 | 0.229 | 0.855 | 0.667 | 0.011 | 1.000 | 0.022 | 0.033 | 4583 | 2 | 178 | 1 |
| Baseline + treatment exposure + procedure burden | Random Forest | 3 | 0.964 | 0.261 | 0.856 | 0.000 | 0.000 | 1.000 | 0.000 | 0.031 | 4260 | 0 | 160 | 0 |
| Baseline + treatment exposure + procedure burden | Random Forest | 4 | 0.965 | 0.196 | 0.848 | 0.500 | 0.006 | 1.000 | 0.013 | 0.031 | 4334 | 1 | 155 | 1 |
| Baseline + treatment exposure + procedure burden | Random Forest | 5 | 0.965 | 0.205 | 0.812 | 0.800 | 0.023 | 1.000 | 0.045 | 0.032 | 4633 | 4 | 168 | 1 |
| Baseline + treatment exposure + procedure burden | Gradient Boosting | 1 | 0.965 | 0.186 | 0.838 | 0.346 | 0.060 | 0.996 | 0.102 | 0.030 | 4351 | 9 | 142 | 17 |
| Baseline + treatment exposure + procedure burden | Gradient Boosting | 2 | 0.955 | 0.161 | 0.855 | 0.224 | 0.072 | 0.990 | 0.109 | 0.038 | 4539 | 13 | 167 | 45 |
| Baseline + treatment exposure + procedure burden | Gradient Boosting | 3 | 0.966 | 0.270 | 0.864 | 0.786 | 0.069 | 0.999 | 0.126 | 0.030 | 4257 | 11 | 149 | 3 |
| Baseline + treatment exposure + procedure burden | Gradient Boosting | 4 | 0.965 | 0.198 | 0.855 | 0.333 | 0.019 | 0.999 | 0.036 | 0.031 | 4329 | 3 | 153 | 6 |
| Baseline + treatment exposure + procedure burden | Gradient Boosting | 5 | 0.963 | 0.180 | 0.840 | 0.385 | 0.058 | 0.997 | 0.101 | 0.033 | 4618 | 10 | 162 | 16 |
| Baseline + treatment exposure + procedure burden | LightGBM | 1 | 0.966 | 0.167 | 0.812 | 0.458 | 0.073 | 0.997 | 0.126 | 0.031 | 4355 | 11 | 140 | 13 |
| Baseline + treatment exposure + procedure burden | LightGBM | 2 | 0.961 | 0.164 | 0.844 | 0.429 | 0.067 | 0.997 | 0.115 | 0.036 | 4568 | 12 | 168 | 16 |
| Baseline + treatment exposure + procedure burden | LightGBM | 3 | 0.965 | 0.238 | 0.855 | 0.667 | 0.050 | 0.999 | 0.093 | 0.031 | 4256 | 8 | 152 | 4 |
| Baseline + treatment exposure + procedure burden | LightGBM | 4 | 0.964 | 0.168 | 0.823 | 0.312 | 0.032 | 0.997 | 0.058 | 0.032 | 4324 | 5 | 151 | 11 |
| Baseline + treatment exposure + procedure burden | LightGBM | 5 | 0.963 | 0.172 | 0.827 | 0.393 | 0.064 | 0.996 | 0.110 | 0.033 | 4617 | 11 | 161 | 17 |
| Baseline + all clinical domains | Logistic Regression | 1 | 0.967 | 0.247 | 0.842 | 0.500 | 0.126 | 0.996 | 0.201 | 0.029 | 4349 | 19 | 132 | 19 |
| Baseline + all clinical domains | Logistic Regression | 2 | 0.962 | 0.303 | 0.897 | 0.516 | 0.089 | 0.997 | 0.152 | 0.031 | 4569 | 16 | 164 | 15 |

Continued on next page

eTable 8. MIMIC-IV performance metrics for all feature matrices and algorithms. (continued)

| Feature matrix | Model | Fold | Accuracy | AUPRC | AUROC | Precision | Sensitivity | Specificity | F1 | Brier score | TN | TP | FN | FP |
|---|---|---|---|---|---|---|---|---|---|---|---|---|---|---|
| Baseline + all clinical domains | Logistic Regression | 3 | 0.965 | 0.275 | 0.882 | 0.615 | 0.100 | 0.998 | 0.172 | 0.030 | 4250 | 16 | 144 | 10 |
| Baseline + all clinical domains | Logistic Regression | 4 | 0.963 | 0.215 | 0.870 | 0.273 | 0.038 | 0.996 | 0.067 | 0.030 | 4319 | 6 | 150 | 16 |
| Baseline + all clinical domains | Logistic Regression | 5 | 0.964 | 0.246 | 0.867 | 0.478 | 0.064 | 0.997 | 0.113 | 0.031 | 4622 | 11 | 161 | 12 |
| Baseline + all clinical domains | Random Forest | 1 | 0.967 | 0.234 | 0.826 | 0.750 | 0.020 | 1.000 | 0.039 | 0.029 | 4367 | 3 | 148 | 1 |
| Baseline + all clinical domains | Random Forest | 2 | 0.962 | 0.289 | 0.883 | 0.600 | 0.017 | 1.000 | 0.032 | 0.031 | 4582 | 3 | 177 | 2 |
| Baseline + all clinical domains | Random Forest | 3 | 0.964 | 0.274 | 0.873 | 0.800 | 0.025 | 1.000 | 0.048 | 0.030 | 4259 | 4 | 156 | 1 |
| Baseline + all clinical domains | Random Forest | 4 | 0.965 | 0.223 | 0.880 | 0.600 | 0.019 | 1.000 | 0.037 | 0.030 | 4333 | 3 | 153 | 2 |
| Baseline + all clinical domains | Random Forest | 5 | 0.964 | 0.225 | 0.855 | 0.571 | 0.023 | 0.999 | 0.045 | 0.031 | 4631 | 4 | 168 | 3 |
| Baseline + all clinical domains | Gradient Boosting | 1 | 0.967 | 0.232 | 0.837 | 0.526 | 0.132 | 0.996 | 0.212 | 0.029 | 4350 | 20 | 131 | 18 |
| Baseline + all clinical domains | Gradient Boosting | 2 | 0.954 | 0.214 | 0.886 | 0.247 | 0.106 | 0.987 | 0.148 | 0.035 | 4526 | 19 | 161 | 58 |
| Baseline + all clinical domains | Gradient Boosting | 3 | 0.964 | 0.236 | 0.881 | 0.522 | 0.075 | 0.997 | 0.131 | 0.031 | 4249 | 12 | 148 | 11 |
| Baseline + all clinical domains | Gradient Boosting | 4 | 0.964 | 0.227 | 0.887 | 0.375 | 0.058 | 0.997 | 0.100 | 0.030 | 4320 | 9 | 147 | 15 |
| Baseline + all clinical domains | Gradient Boosting | 5 | 0.963 | 0.212 | 0.863 | 0.360 | 0.052 | 0.997 | 0.091 | 0.032 | 4618 | 9 | 163 | 16 |
| Baseline + all clinical domains | LightGBM | 1 | 0.966 | 0.214 | 0.827 | 0.464 | 0.086 | 0.997 | 0.145 | 0.030 | 4353 | 13 | 138 | 15 |
| Baseline + all clinical domains | LightGBM | 2 | 0.955 | 0.195 | 0.874 | 0.275 | 0.122 | 0.987 | 0.169 | 0.036 | 4526 | 22 | 158 | 58 |
| Baseline + all clinical domains | LightGBM | 3 | 0.962 | 0.244 | 0.879 | 0.364 | 0.050 | 0.997 | 0.088 | 0.031 | 4246 | 8 | 152 | 14 |
| Baseline + all clinical domains | LightGBM | 4 | 0.964 | 0.186 | 0.859 | 0.294 | 0.032 | 0.997 | 0.058 | 0.031 | 4323 | 5 | 151 | 12 |
| Baseline + all clinical domains | LightGBM | 5 | 0.963 | 0.209 | 0.862 | 0.364 | 0.047 | 0.997 | 0.082 | 0.032 | 4620 | 8 | 164 | 14 |

*Notes:* AUPRC = area under the precision-recall curve; AUROC = area under the receiver operating characteristic curve; TN = true negatives; TP = true positives; FN = false negatives; FP = false positives. MIMIC-IV rows are validation-fold results from the saved patient-grouped five-fold cross-validation outputs for each feature matrix and algorithm.

### Confidence Intervals

This section reports best-model AUROC and AUPRC point estimates and 95% confidence intervals using a common structure for both datasets.

eTable 9. CHoRUS best-model AUPRC and AUROC confidence intervals.

| Feature matrix | Best algorithm | Mean AUPRC | AUPRC 95% CI lower | AUPRC 95% CI upper | Mean AUROC | AUROC 95% CI lower | AUROC 95% CI upper |
|---|---|---|---|---|---|---|---|
| Baseline | Gradient Boosting | 0.229 | 0.197 | 0.245 | 0.817 | 0.802 | 0.829 |
| Baseline + physiological severity | Gradient Boosting | 0.381 | 0.344 | 0.410 | 0.858 | 0.843 | 0.868 |
| Baseline + treatment exposure | Gradient Boosting | 0.275 | 0.241 | 0.301 | 0.830 | 0.814 | 0.841 |
| Baseline + procedure burden | Gradient Boosting | 0.341 | 0.301 | 0.363 | 0.841 | 0.824 | 0.850 |
| Baseline + physiological severity + treatment exposure | LightGBM | 0.372 | 0.332 | 0.400 | 0.854 | 0.840 | 0.865 |
| Baseline + physiological severity + procedure burden | Gradient Boosting | 0.391 | 0.352 | 0.416 | 0.863 | 0.846 | 0.871 |
| Baseline + treatment exposure + procedure burden | Gradient Boosting | 0.343 | 0.297 | 0.359 | 0.845 | 0.828 | 0.853 |
| Baseline + all clinical domains | Gradient Boosting | 0.382 | 0.338 | 0.402 | 0.859 | 0.841 | 0.867 |

*Notes:* Best algorithm was selected within each dataset and feature matrix. Confidence intervals are 95% intervals from the source sheets. CHoRUS point estimates are means across folds for the selected best-performing model in each feature matrix.

eTable 10. MIMIC-IV best-model AUPRC and AUROC confidence intervals.

| Feature matrix | Best algorithm | Mean AUPRC | AUPRC 95% CI lower | AUPRC 95% CI upper | Mean AUROC | AUROC 95% CI lower | AUROC 95% CI upper |
|---|---|---|---|---|---|---|---|
| Baseline | Logistic Regression | 0.133 | 0.112 | 0.145 | 0.811 | 0.797 | 0.825 |
| Baseline + physiological severity | Random Forest | 0.242 | 0.206 | 0.267 | 0.859 | 0.846 | 0.872 |
| Baseline + treatment exposure | Logistic Regression | 0.218 | 0.186 | 0.240 | 0.848 | 0.835 | 0.861 |
| Baseline + procedure burden | Logistic Regression | 0.196 | 0.165 | 0.214 | 0.838 | 0.823 | 0.851 |
| Baseline + physiological severity + treatment exposure | Random Forest | 0.247 | 0.213 | 0.274 | 0.862 | 0.849 | 0.874 |
| Baseline + physiological severity + procedure burden | Random Forest | 0.242 | 0.208 | 0.267 | 0.863 | 0.851 | 0.876 |
| Baseline + treatment exposure + procedure burden | Logistic Regression | 0.233 | 0.198 | 0.255 | 0.852 | 0.840 | 0.865 |
| Baseline + all clinical domains | Logistic Regression | 0.257 | 0.223 | 0.283 | 0.872 | 0.860 | 0.884 |

*Notes:* Best algorithm was selected within each dataset and feature matrix. Confidence intervals are 95% intervals from the source sheets. MIMIC-IV point estimates are means across folds for the selected best-performing model in each feature matrix.

### Best-Model Clinical-Domain Comparisons

eTable 11. Standalone clinical-domain performance in CHoRUS and MIMIC-IV using selected algorithms.

| Dataset | Feature matrix | Selected algorithm | Median AUPRC [Q1–Q3] | Change in AUPRC from baseline | Median AUROC [Q1–Q3] | Change in AUROC from baseline | Median Brier score [Q1–Q3] | Change in Brier score from baseline |
|---|---|---|---|---|---|---|---|---|
| CHoRUS | Baseline | Gradient Boosting | 0.241 [0.211–0.244] | 0.000 | 0.817 [0.808–0.820] | 0.000 | 0.042 [0.038–0.042] | 0.000 |
| | Baseline + physiological severity | Gradient Boosting | 0.380 [0.354–0.415] | +0.139 | 0.858 [0.845–0.874] | +0.041 | 0.036 [0.033–0.036] | -0.006 |
| | Baseline + treatment exposure | Gradient Boosting | 0.282 [0.267–0.290] | +0.041 | 0.827 [0.818–0.838] | +0.010 | 0.040 [0.036–0.040] | -0.002 |
| | Baseline + procedure burden | Gradient Boosting | 0.330 [0.326–0.333] | +0.089 | 0.836 [0.824–0.851] | +0.019 | 0.038 [0.034–0.039] | -0.004 |
| MIMIC-IV | Baseline | Logistic Regression | 0.131 [0.117–0.149] | 0.000 | 0.810 [0.805–0.817] | 0.000 | 0.033 [0.032–0.033] | 0.000 |
| | Baseline + physiological severity | Random Forest | 0.239 [0.223–0.255] | +0.108 | 0.869 [0.850–0.877] | +0.059 | 0.031 [0.030–0.031] | -0.002 |
| | Baseline + treatment exposure | Logistic Regression | 0.200 [0.199–0.248] | +0.069 | 0.847 [0.838–0.858] | +0.037 | 0.031 [0.030–0.032] | -0.002 |
| | Baseline + procedure burden | Logistic Regression | 0.181 [0.176–0.210] | +0.050 | 0.838 [0.829–0.839] | +0.028 | 0.031 [0.031–0.032] | -0.002 |

AUROC denotes area under the receiver operating characteristic curve; AUPRC denotes area under the precision-recall curve; Q1 denotes the 25th percentile; Q3 denotes the 75th percentile. Values are reported as median [25th percentile–75th percentile] across the five patient-grouped cross-validation folds for the selected algorithm within each feature matrix. Percentiles were calculated using the NumPy/pandas default linear percentile method; with five folds, Q1 and Q3 correspond to the second and fourth ordered fold values. Changes are calculated as the feature-matrix median minus the baseline median. Positive AUROC and AUPRC changes indicate higher discrimination; negative Brier-score changes indicate lower and better probabilistic error.

eTable 12. Complementary and all-domain performance in CHoRUS and MIMIC-IV using selected algorithms.

| Dataset | Feature matrix | Selected algorithm | Median AUPRC [Q1–Q3] | Change in AUPRC from baseline | Change in AUPRC from best standalone | Median AUROC [Q1–Q3] | Change in AUROC from baseline | Change in AUROC from best standalone | Median Brier score [Q1–Q3] | Change in Brier score from baseline | Change in Brier score from best standalone |
|---|---|---|---|---|---|---|---|---|---|---|---|
| CHoRUS | Baseline + physiological severity + treatment exposure | LightGBM | 0.372 [0.368–0.392] | +0.131 | -0.008 | 0.855 [0.851–0.856] | +0.038 | -0.003 | 0.037 [0.033–0.037] | -0.005 | +0.001 |
| | Baseline + physiological severity + procedure burden | Gradient Boosting | 0.404 [0.380–0.405] | +0.163 | +0.024 | 0.855 [0.853–0.878] | +0.038 | -0.003 | 0.035 [0.032–0.037] | -0.007 | -0.001 |
| | Baseline + treatment exposure + procedure burden | Gradient Boosting | 0.343 [0.308–0.345] | +0.102 | -0.037 | 0.837 [0.833–0.856] | +0.020 | -0.021 | 0.038 [0.033–0.040] | -0.004 | +0.002 |
| | Baseline + all clinical domains | Gradient Boosting | 0.398 [0.373–0.409] | +0.157 | +0.018 | 0.855 [0.853–0.872] | +0.038 | -0.003 | 0.035 [0.033–0.037] | -0.007 | -0.001 |
| MIMIC-IV | Baseline + physiological severity + treatment exposure | Random Forest | 0.232 [0.225–0.268] | +0.101 | -0.007 | 0.872 [0.851–0.878] | +0.062 | +0.003 | 0.030 [0.030–0.031] | -0.003 | -0.001 |
| | Baseline + physiological severity + procedure burden | Random Forest | 0.236 [0.219–0.258] | +0.105 | -0.003 | 0.867 [0.856–0.884] | +0.057 | -0.002 | 0.031 [0.030–0.031] | -0.002 | 0.000 |
| | Baseline + treatment exposure + procedure burden | Logistic Regression | 0.212 [0.211–0.251] | +0.081 | -0.027 | 0.848 [0.841–0.864] | +0.038 | -0.021 | 0.030 [0.030–0.031] | -0.003 | -0.001 |
| | Baseline + all clinical domains | Logistic Regression | 0.247 [0.246–0.275] | +0.116 | +0.008 | 0.870 [0.867–0.882] | +0.060 | +0.001 | 0.030 [0.030–0.031] | -0.003 | -0.001 |

The standalone comparator is Baseline + physiological severity for both datasets. AUROC denotes area under the receiver operating characteristic curve; AUPRC denotes area under the precision-recall curve; Q1 denotes the 25th percentile; Q3 denotes the 75th percentile. Values are reported as median [25th percentile–75th percentile] across the five patient-grouped cross-validation folds for the selected algorithm within each feature matrix. Percentiles were calculated using the NumPy/pandas default linear percentile method; with five folds, Q1 and Q3 correspond to the second and fourth ordered fold values. Changes are calculated as the complementary or all-domain model median minus the comparator median. Positive AUROC and AUPRC changes indicate higher discrimination; negative Brier-score changes indicate lower and better probabilistic error.

### Calibration Plots

Calibration plots use mean predicted probability on the x-axis and observed mortality rate on the y-axis. The dashed diagonal line indicates perfect calibration.

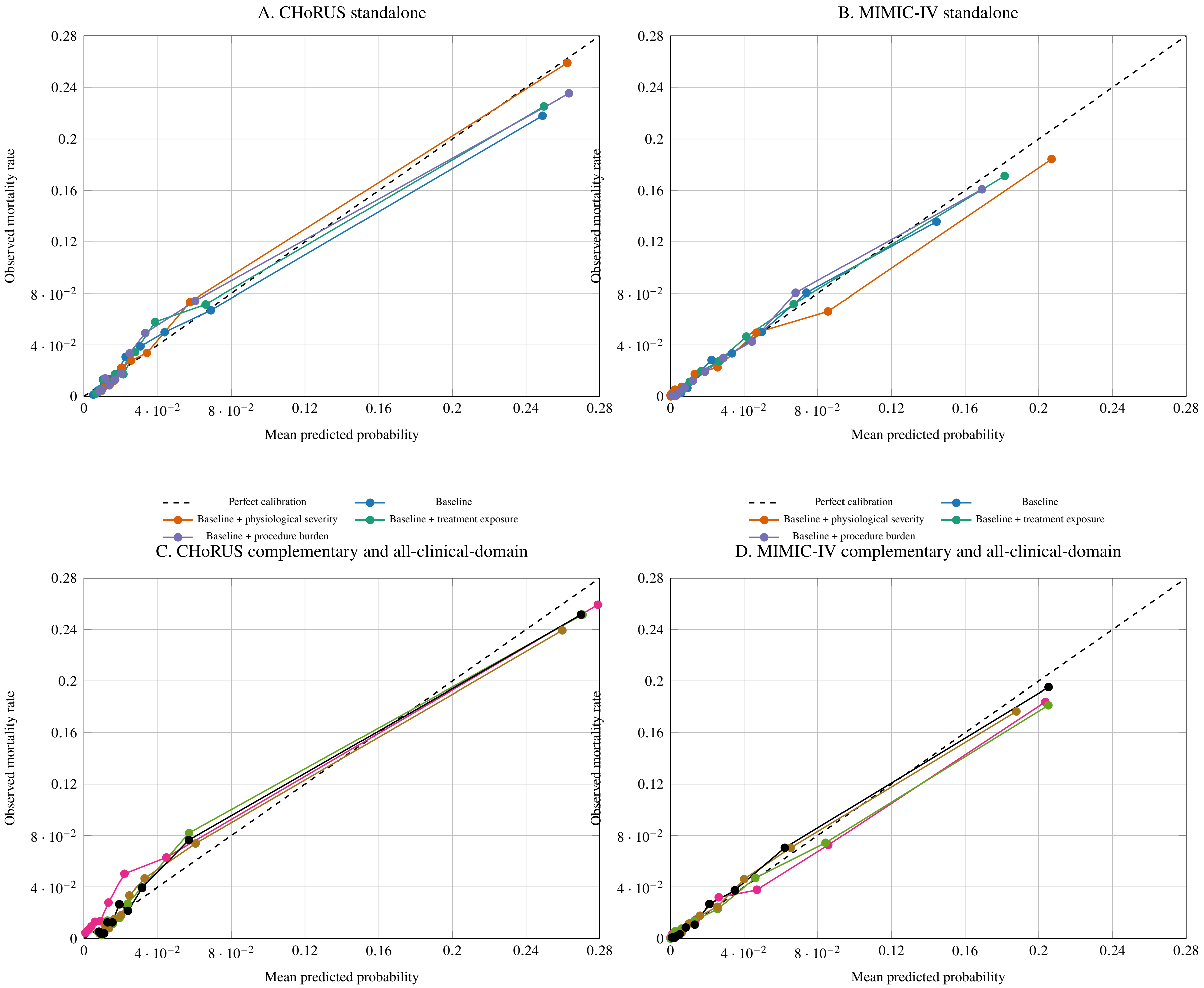
A. CHoRUS standalone
B. MIMIC-IV standalone
C. CHoRUS complementary and all-clinical-domain
D. MIMIC-IV complementary and all-clinical-domain
Observed mortality rate
Mean predicted probability
0
$4 \cdot 10^{-2}$
$8 \cdot 10^{-2}$
0.12
0.16
0.2
0.24
0.28
Perfect calibration
Baseline
Baseline + physiological severity
Baseline + treatment exposure
Baseline + procedure burden

### Decision-Curve Plots

Decision-curve plots use threshold probability on the x-axis and net benefit on the y-axis. Treat-all and treat-none reference strategies are shown for each dataset.

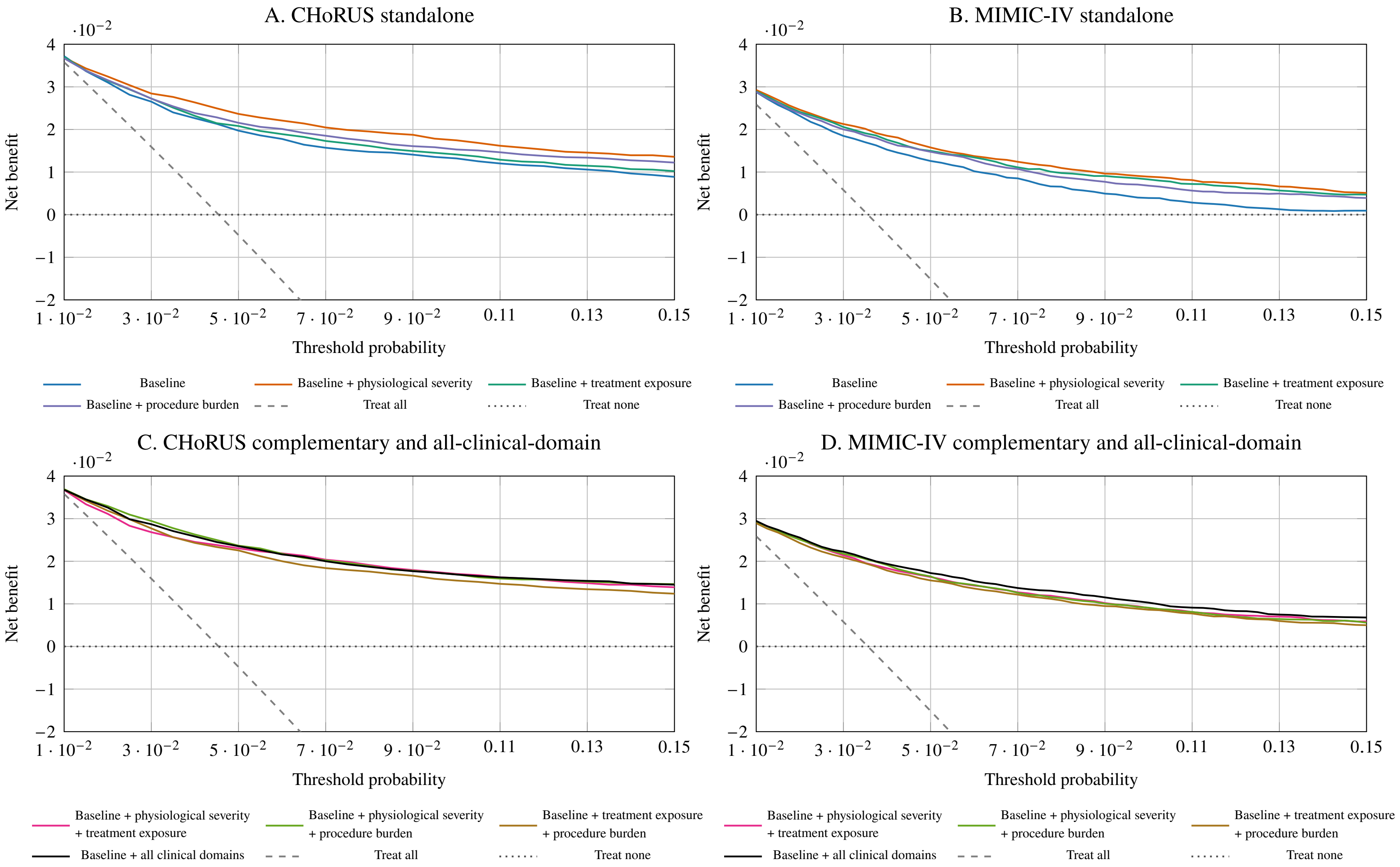

eFigure 2. Four-panel decision-curve plots generated from saved decision-curve coordinate outputs. Panel A shows CHoRUS standalone feature matrices; Panel B shows MIMIC-IV standalone feature matrices; Panel C shows CHoRUS complementary and all-clinical-domain feature matrices; Panel D shows MIMIC-IV complementary and all-clinical-domain feature matrices. Treat-all and treat-none reference strategies are shown in every panel.

### Sensitivity at 90% Specificity

eTable 13. CHoRUS sensitivity-at-90% specificity analysis.

| Feature matrix | Threshold | Specificity | Sensitivity | Alerts per 100 encounters | Deaths per 100 alerts | False alerts per 100 alerts | Alerts per death identified |
|---|---|---|---|---|---|---|---|
| Baseline | 0.080 | 0.900 | 0.507 | 11.847 | 19.442 | 80.558 | 5.143 |
| Baseline + physiological severity | 0.068 | 0.900 | 0.609 | 12.309 | 22.463 | 77.537 | 4.452 |
| Baseline + treatment exposure | 0.079 | 0.900 | 0.540 | 11.997 | 20.445 | 79.555 | 4.891 |
| Baseline + procedure burden | 0.073 | 0.900 | 0.562 | 12.096 | 21.100 | 78.900 | 4.739 |
| Baseline + physiological severity + treatment exposure | 0.055 | 0.900 | 0.613 | 12.327 | 22.577 | 77.423 | 4.429 |
| Baseline + physiological severity + procedure burden | 0.070 | 0.900 | 0.603 | 12.282 | 22.292 | 77.708 | 4.486 |
| Baseline + treatment exposure + procedure burden | 0.074 | 0.900 | 0.567 | 12.119 | 21.247 | 78.753 | 4.707 |
| Baseline + all clinical domains | 0.069 | 0.900 | 0.600 | 12.268 | 22.206 | 77.794 | 4.503 |

*Notes:* Thresholds were chosen to achieve approximately 90% specificity. Deaths per 100 alerts equals positive predictive value multiplied by 100; false alerts per 100 alerts equals 100 minus deaths per 100 alerts.

eTable 14. MIMIC-IV sensitivity-at-90% specificity analysis.

| Feature matrix | Threshold | Specificity | Sensitivity | Alerts per 100 encounters | Deaths per 100 alerts | False alerts per 100 alerts | Alerts per death identified |
|---|---|---|---|---|---|---|---|
| Baseline | 0.086 | 0.900 | 0.411 | 11.109 | 13.190 | 86.810 | 7.582 |
| Baseline + physiological severity | 0.104 | 0.900 | 0.557 | 11.626 | 17.053 | 82.947 | 5.864 |
| Baseline + treatment exposure | 0.080 | 0.900 | 0.510 | 11.461 | 15.857 | 84.143 | 6.306 |
| Baseline + procedure burden | 0.080 | 0.900 | 0.482 | 11.357 | 15.123 | 84.877 | 6.613 |
| Baseline + physiological severity + treatment exposure | 0.105 | 0.900 | 0.560 | 11.639 | 17.146 | 82.854 | 5.832 |
| Baseline + physiological severity + procedure burden | 0.104 | 0.900 | 0.553 | 11.613 | 16.960 | 83.040 | 5.896 |
| Baseline + treatment exposure + procedure burden | 0.078 | 0.900 | 0.542 | 11.574 | 16.679 | 83.321 | 5.995 |
| Baseline + all clinical domains | 0.076 | 0.900 | 0.591 | 11.748 | 17.913 | 82.087 | 5.583 |

*Notes:* Thresholds were chosen to achieve approximately 90% specificity. Deaths per 100 alerts equals positive predictive value multiplied by 100; false alerts per 100 alerts equals 100 minus deaths per 100 alerts.

### Top-10% Risk Analysis

eTable 15. CHoRUS highest-risk 10% analysis.

| Feature matrix | N encounters | N deaths | N alerts | Alerts per 100 encounters | Deaths captured (%) | Deaths captured 95% CI lower | Deaths captured 95% CI upper | Deaths per 100 alerts | Deaths per 100 alerts 95% CI lower | Deaths per 100 alerts 95% CI upper | False alerts per 100 alerts | Alerts per death identified |
|---|---|---|---|---|---|---|---|---|---|---|---|---|
| Baseline | 22098 | 1004 | 2210 | 10.001 | 48.008 | 44.073 | 52.116 | 21.810 | 19.739 | 24.041 | 78.190 | 4.585 |
| Baseline + physiological severity | 22098 | 1004 | 2210 | 10.001 | 56.972 | 53.110 | 61.240 | 25.882 | 23.405 | 28.432 | 74.118 | 3.864 |
| Baseline + treatment exposure | 22098 | 1004 | 2210 | 10.001 | 49.602 | 45.849 | 53.722 | 22.534 | 20.515 | 24.721 | 77.466 | 4.438 |
| Baseline + procedure burden | 22098 | 1004 | 2210 | 10.001 | 51.793 | 48.311 | 56.207 | 23.529 | 21.393 | 25.914 | 76.471 | 4.250 |
| Baseline + physiological severity + treatment exposure | 22098 | 1004 | 2210 | 10.001 | 57.072 | 53.299 | 61.079 | 25.928 | 23.604 | 28.185 | 74.072 | 3.857 |
| Baseline + physiological severity + procedure burden | 22098 | 1004 | 2210 | 10.001 | 55.378 | 51.571 | 59.566 | 25.158 | 22.817 | 27.502 | 74.842 | 3.975 |

Continued on next page

eTable 15. CHoRUS highest-risk 10% analysis. (continued)

| Feature matrix | N encounters | N deaths | N alerts | Alerts per 100 encounters | Deaths captured (%) | Deaths captured 95% CI lower | Deaths captured 95% CI upper | Deaths per 100 alerts | Deaths per 100 alerts 95% CI lower | Deaths per 100 alerts 95% CI upper | False alerts per 100 alerts | Alerts per death identified |
|---|---|---|---|---|---|---|---|---|---|---|---|---|
| Baseline + treatment exposure + procedure burden | 22098 | 1004 | 2210 | 10.001 | 52.689 | 49.056 | 56.853 | 23.937 | 21.839 | 26.251 | 76.063 | 4.178 |
| Baseline + all clinical domains | 22098 | 1004 | 2210 | 10.001 | 55.378 | 51.500 | 59.391 | 25.158 | 22.823 | 27.435 | 74.842 | 3.975 |

*Notes:* The top-risk analysis flags exactly the highest-risk 10% of encounters. Confidence intervals are reported from the CHoRUS source workbook.

eTable 16. MIMIC-IV highest-risk 10% analysis.

| Feature matrix | N encounters | N deaths | N alerts | Alerts per 100 encounters | Deaths captured (%) | Deaths captured 95% CI lower | Deaths captured 95% CI upper | Deaths per 100 alerts | Deaths per 100 alerts 95% CI lower | Deaths per 100 alerts 95% CI upper | False alerts per 100 alerts | Alerts per death identified |
|---|---|---|---|---|---|---|---|---|---|---|---|---|
| Baseline | 23000 | 819 | 2300 | 10.000 | 38.095 | 34.633 | 41.688 | 13.565 | 12.046 | 15.227 | 86.435 | 7.372 |
| Baseline + physiological severity | 23000 | 819 | 2300 | 10.000 | 51.770 | 48.371 | 55.462 | 18.435 | 16.666 | 20.322 | 81.565 | 5.425 |
| Baseline + treatment exposure | 23000 | 819 | 2300 | 10.000 | 48.107 | 44.728 | 51.786 | 17.130 | 15.443 | 18.896 | 82.870 | 5.838 |
| Baseline + procedure burden | 23000 | 819 | 2300 | 10.000 | 45.177 | 41.567 | 48.505 | 16.087 | 14.371 | 17.670 | 83.913 | 6.216 |
| Baseline + physiological severity + treatment exposure | 23000 | 819 | 2300 | 10.000 | 51.648 | 48.157 | 55.387 | 18.391 | 16.580 | 20.194 | 81.609 | 5.437 |
| Baseline + physiological severity + procedure burden | 23000 | 819 | 2300 | 10.000 | 50.916 | 47.406 | 54.674 | 18.130 | 16.423 | 19.966 | 81.870 | 5.516 |
| Baseline + treatment exposure + procedure burden | 23000 | 819 | 2300 | 10.000 | 49.573 | 46.055 | 53.199 | 17.652 | 15.957 | 19.485 | 82.348 | 5.665 |
| Baseline + all clinical domains | 23000 | 819 | 2300 | 10.000 | 54.823 | 51.432 | 58.529 | 19.522 | 17.786 | 21.348 | 80.478 | 5.122 |

*Notes:* The top-risk analysis flags exactly the highest-risk 10% of encounters. MIMIC-IV confidence intervals for fields absent from the source workbook were calculated from saved out-of-fold predictions using patient-level bootstrap resampling.

### Abbreviation Notes

AUROC denotes area under the receiver operating characteristic curve. AUPRC denotes area under the precision-recall curve. TP, TN, FP, and FN denote true positives, true negatives, false positives, and false negatives, respectively. Brier score is reported on the probability scale.

### SHAP Feature-Importance Tables

eTable 17. CHoRUS SHAP feature importance for Baseline.

| Feature matrix | feature | concept name | mean_absolute_shap |
|---|---|---|---|
| Baseline | prior_visit_count | Not Applicable | 0.025468 |
| Baseline | age_at_visit | Not Applicable | 0.014280 |
| Baseline | charlson_index | Not Applicable | 0.012629 |
| Baseline | visit_type_Emergency Room and Inpatient Visit | Not Applicable | 0.011471 |
| Baseline | prior_acute_visit_count | Not Applicable | 0.008380 |
| Baseline | visit_type_Observation Room | Not Applicable | 0.004032 |
| Baseline | has_prior_visit | Not Applicable | 0.003418 |
| Baseline | race_category_Other | Not Applicable | 0.002840 |
| Baseline | visit_type_Emergency Room - Hospital | Not Applicable | 0.002722 |
| Baseline | race_category_White | Not Applicable | 0.002700 |
| Baseline | visit_type_Hospital | Not Applicable | 0.002331 |
| Baseline | sex_category_Female | Not Applicable | 0.001393 |
| Baseline | race_category_Black | Not Applicable | 0.001001 |
| Baseline | visit_type_Inpatient Hospital | Not Applicable | 0.000714 |
| Baseline | sex_category_Male | Not Applicable | 0.000679 |

Continued on next page

eTable 17. CHoRUS SHAP feature importance for Baseline. (continued)

| Feature matrix | feature | concept name | mean_absolute_shap |
|---|---|---|---|
| Baseline | ethnicity_category_Unknown | Not Applicable | 0.000513 |
| Baseline | ethnicity_category_Hispanic | Not Applicable | 0.000373 |
| Baseline | ethnicity_category_Non-Hispanic | Not Applicable | 0.000281 |
| Baseline | race_category_Unknown | Not Applicable | 0.000119 |
| Baseline | race_category_Asian | Not Applicable | 0.000112 |
| Baseline | ethnicity_category_Other | Not Applicable | 0.000000 |

*Notes:* Rows, feature order, concept names, and mean absolute SHAP values are reproduced from the formatted CHoRUS SHAP workbook. SHAP values were generated from the best-performing algorithm selected for each feature matrix. Best-performing algorithms were selected by highest mean cross-validated AUPRC, with mean AUROC used as the tie-breaker. SHAP values are model-specific attributions, not causal effects; absolute SHAP magnitudes should not be compared directly across separately trained models or datasets.

eTable 18. CHoRUS SHAP feature importance for Baseline + physiological severity.

| Feature matrix | feature | concept name | mean_absolute_shap |
|---|---|---|---|
| Baseline + physiological severity | measurement_3037318_max_24h | Physical activity Braden scale — maximum, first 24 hours | 0.028827 |
| Baseline + physiological severity | measurement_3035206_mean_24h | Physical mobility Braden scale — mean, first 24 hours | 0.017330 |
| Baseline + physiological severity | prior_visit_count | Not applicable | 0.016571 |
| Baseline + physiological severity | measurement_4301868_mean_24h | Pulse rate — mean, first 24 hours | 0.014584 |
| Baseline + physiological severity | age_at_visit | Not applicable | 0.014496 |
| Baseline + physiological severity | measurement_3036098_max_24h | Sensory perception Braden scale — maximum, first 24 hours | 0.012971 |
| Baseline + physiological severity | charlson_index | Not applicable | 0.011618 |
| Baseline + physiological severity | measurement_3035816_mean_24h | Nutrition intake pattern Braden scale — mean, first 24 hours | 0.009690 |
| Baseline + physiological severity | prior_acute_visit_count | Not applicable | 0.009000 |
| Baseline + physiological severity | measurement_3024171_count_24h | Respiratory rate — measurement count, first 24 hours | 0.005450 |
| Baseline + physiological severity | visit_type_Emergency Room and Inpatient Visit | Not applicable | 0.005012 |
| Baseline + physiological severity | measurement_4020553_std_24h | Oxygen saturation measurement — standard deviation, first 24 hours | 0.004963 |
| Baseline + physiological severity | measurement_3037347_mean_24h | Friction and shear Braden scale — mean, first 24 hours | 0.004354 |
| Baseline + physiological severity | measurement_3004249_std_24h | Systolic blood pressure — standard deviation, first 24 hours | 0.003464 |
| Baseline + physiological severity | measurement_4301868_std_24h | Pulse rate — standard deviation, first 24 hours | 0.002900 |
| Baseline + physiological severity | measurement_3004249_mean_24h | Systolic blood pressure — mean, first 24 hours | 0.001989 |
| Baseline + physiological severity | measurement_3004249_min_24h | Systolic blood pressure — minimum, first 24 hours | 0.001901 |

Continued on next page

eTable 18. CHoRUS SHAP feature importance for Baseline + physiological severity. (continued)

| Feature matrix | feature | concept name | mean_absolute_shap |
|---|---|---|---|
| Baseline + physiological severity | race_category_Other | Not applicable | 0.001853 |
| Baseline + physiological severity | visit_type_Emergency Room - Hospital | Not applicable | 0.001821 |
| Baseline + physiological severity | has_prior_visit | Not applicable | 0.001735 |
| Baseline + physiological severity | measurement_4020553_count_24h | Oxygen saturation measurement — measurement count, first 24 hours | 0.001466 |
| Baseline + physiological severity | measurement_4301868_min_24h | Pulse rate — minimum, first 24 hours | 0.001346 |
| Baseline + physiological severity | measurement_4301868_count_24h | Pulse rate — measurement count, first 24 hours | 0.001338 |
| Baseline + physiological severity | measurement_4020553_mean_24h | Oxygen saturation measurement — mean, first 24 hours | 0.001325 |
| Baseline + physiological severity | race_category_White | Not applicable | 0.000808 |
| Baseline + physiological severity | measurement_4301868_max_24h | Pulse rate — maximum, first 24 hours | 0.000536 |
| Baseline + physiological severity | race_category_Black | Not applicable | 0.000378 |
| Baseline + physiological severity | visit_type_Observation Room | Not applicable | 0.000259 |
| Baseline + physiological severity | visit_type_Inpatient Hospital | Not applicable | 0.000143 |
| Baseline + physiological severity | measurement_4020553_min_24h | Oxygen saturation measurement — minimum, first 24 hours | 0.000071 |
| Baseline + physiological severity | race_category_Asian | Not applicable | 0.000068 |
| Baseline + physiological severity | sex_category_Male | Not applicable | 0.000001 |
| Baseline + physiological severity | measurement_4020553_missing_24h | Oxygen saturation measurement — missingness indicator, first 24 hours | 0.000000 |
| Baseline + physiological severity | visit_type_Hospital | Not applicable | 0.000000 |
| Baseline + physiological severity | measurement_4020553_max_24h | Oxygen saturation measurement — maximum, first 24 hours | 0.000000 |
| Baseline + physiological severity | ethnicity_category_Unknown | Not applicable | 0.000000 |
| Baseline + physiological severity | sex_category_Female | Not applicable | 0.000000 |
| Baseline + physiological severity | ethnicity_category_Other | Not applicable | 0.000000 |
| Baseline + physiological severity | ethnicity_category_Non-Hispanic | Not applicable | 0.000000 |
| Baseline + physiological severity | ethnicity_category_Hispanic | Not applicable | 0.000000 |
| Baseline + physiological severity | race_category_Unknown | Not applicable | 0.000000 |

Continued on next page

eTable 18. CHoRUS SHAP feature importance for Baseline + physiological severity. (continued)

| Feature matrix | feature | concept name | mean_absolute_shap |
|---|---|---|---|
| Baseline + physiological severity | measurement_4301868_missing_24h | Pulse rate — missingness indicator, first 24 hours | 0.000000 |

*Notes:* Rows, feature order, concept names, and mean absolute SHAP values are reproduced from the formatted CHoRUS SHAP workbook. SHAP values were generated from the best-performing algorithm selected for each feature matrix. Best-performing algorithms were selected by highest mean cross-validated AUPRC, with mean AUROC used as the tie-breaker. SHAP values are model-specific attributions, not causal effects; absolute SHAP magnitudes should not be compared directly across separately trained models or datasets.

eTable 19. CHoRUS SHAP feature importance for Baseline + treatment exposure.

| Feature matrix | feature | concept name | mean_absolute_shap |
|---|---|---|---|
| Baseline + treatment exposure | prior_visit_count | Not applicable | 0.025263 |
| Baseline + treatment exposure | age_at_visit | Not applicable | 0.018122 |
| Baseline + treatment exposure | charlson_index | Not applicable | 0.011761 |
| Baseline + treatment exposure | visit_type_Emergency Room and Inpatient Visit | Not applicable | 0.008249 |
| Baseline + treatment exposure | repeated_drug_exposure_count_24h | Aggregate treatment-exposure feature | 0.007640 |
| Baseline + treatment exposure | unique_drug_exposure_count_24h | Aggregate treatment-exposure feature | 0.005853 |
| Baseline + treatment exposure | drug_19079524_count24h | sodium chloride 9 MG/ML Injectable Solution | 0.004658 |
| Baseline + treatment exposure | drug_40220390_count24h | propofol 10 MG/ML Injection [Diprivan] | 0.004546 |
| Baseline + treatment exposure | time_to_first_drug_hours | Aggregate treatment-exposure feature | 0.004468 |
| Baseline + treatment exposure | has_prior_visit | Not applicable | 0.003638 |
| Baseline + treatment exposure | prior_acute_visit_count | Not applicable | 0.003187 |
| Baseline + treatment exposure | total_drug_exposure_count_24h | Aggregate treatment-exposure feature | 0.002530 |
| Baseline + treatment exposure | drug_40220388_count24h | 100 ML propofol 10 MG/ML Injection [Diprivan] | 0.002215 |
| Baseline + treatment exposure | race_category_White | Not applicable | 0.001658 |
| Baseline + treatment exposure | drug_43011850_count24h | heparin sodium, porcine 5000 UNT/ML Injectable Solution | 0.001283 |
| Baseline + treatment exposure | drug_43011850_exposed_24h | heparin sodium, porcine 5000 UNT/ML Injectable Solution | 0.000969 |
| Baseline + treatment exposure | race_category_Other | Not applicable | 0.000906 |
| Baseline + treatment exposure | drug_21050327_count24h | Glucose Injectable Solution [Plasma-lyte] | 0.000862 |
| Baseline + treatment exposure | drug_40221385_exposed_24h | 100 ML sodium chloride 9 MG/ML Injection | 0.000842 |
| Baseline + treatment exposure | visit_type_Observation Room | Not applicable | 0.000644 |
| Baseline + treatment exposure | race_category_Unknown | Not applicable | 0.000543 |
| Baseline + treatment exposure | drug_42708658_count24h | docusate sodium 100 MG Oral Capsule [Colace] | 0.000518 |
| Baseline + treatment exposure | drug_19102651_count24h | sennosides, USP 8.6 MG Oral Tablet [Senokot] | 0.000402 |
| Baseline + treatment exposure | drug_19135374_count24h | calcium chloride 0.2 MG/ML / potassium chloride 0.3 MG/ML / sodium chloride 6 MG/ML / sodium lactate 3.1 MG/ML Injectable Solution | 0.000345 |
| Baseline + treatment exposure | drug_1127433_count24h | acetaminophen 325 MG Oral Tablet | 0.000228 |
| Baseline + treatment exposure | visit_type_Emergency Room - Hospital | Not applicable | 0.000211 |
| Baseline + treatment exposure | drug_42479436_count24h | 1000 ML Sodium Chloride 9 MG/ML Injectable Solution | 0.000211 |
| Baseline + treatment exposure | drug_19135374_exposed_24h | calcium chloride 0.2 MG/ML / potassium chloride 0.3 MG/ML / sodium chloride 6 MG/ML / sodium lactate 3.1 MG/ML Injectable Solution | 0.000198 |
| Baseline + treatment exposure | visit_type_Hospital | Not applicable | 0.000196 |
| Baseline + treatment exposure | sex_category_Female | Not applicable | 0.000182 |
| Baseline + treatment exposure | drug_35605482_count24h | 2 ML ondansetron 2 MG/ML Injection | 0.000102 |

Continued on next page

eTable 19. CHoRUS SHAP feature importance for Baseline + treatment exposure. (continued)

| Feature matrix | feature | concept name | mean_absolute_shap |
|---|---|---|---|
| Baseline + treatment exposure | sex_category_Male | Not applicable | 0.000095 |
| Baseline + treatment exposure | drug_46275280_count24h | 50 ML magnesium sulfate 40 MG/ML Injection | 0.000025 |
| Baseline + treatment exposure | race_category_Asian | Not applicable | 0.000021 |
| Baseline + treatment exposure | drug_19127213_count24h | 10 ML sodium chloride 9 MG/ML Prefilled Syringe | 0.000006 |
| Baseline + treatment exposure | drug_40220357_exposed_24h | 1000 ML sodium chloride 9 MG/ML Injection | 0.000000 |
| Baseline + treatment exposure | visit_type_Inpatient Hospital | Not applicable | 0.000000 |
| Baseline + treatment exposure | ethnicity_category_Other | Not applicable | 0.000000 |
| Baseline + treatment exposure | ethnicity_category_Unknown | Not applicable | 0.000000 |
| Baseline + treatment exposure | ethnicity_category_Non-Hispanic | Not applicable | 0.000000 |
| Baseline + treatment exposure | race_category_Black | Not applicable | 0.000000 |
| Baseline + treatment exposure | ethnicity_category_Hispanic | Not applicable | 0.000000 |

*Notes:* Rows, feature order, concept names, and mean absolute SHAP values are reproduced from the formatted CHoRUS SHAP workbook. SHAP values were generated from the best-performing algorithm selected for each feature matrix. Best-performing algorithms were selected by highest mean cross-validated AUPRC, with mean AUROC used as the tie-breaker. SHAP values are model-specific attributions, not causal effects; absolute SHAP magnitudes should not be compared directly across separately trained models or datasets.

eTable 20. CHoRUS SHAP feature importance for Baseline + procedure burden.

| Feature matrix | feature | concept name | mean_absolute_shap |
|---|---|---|---|
| Baseline + procedure burden | age_at_visit | Not applicable | 0.015495 |
| Baseline + procedure burden | charlson_index | Not applicable | 0.012131 |
| Baseline + procedure burden | unique_procedure_count_24h | Aggregate procedure-burden feature | 0.011336 |
| Baseline + procedure burden | prior_visit_count | Not applicable | 0.009807 |
| Baseline + procedure burden | procedure_725068_count24h | Radiologic examination, chest; single view | 0.008408 |
| Baseline + procedure burden | procedure_2147482992_presence_24h | LINE CARE \| LINE SITE ASSESSMENT \| LINE SITE ASSESSMENT | 0.007905 |
| Baseline + procedure burden | procedure_count_total_24h | Aggregate procedure-burden feature | 0.006126 |
| Baseline + procedure burden | prior_acute_visit_count | Not applicable | 0.005105 |
| Baseline + procedure burden | procedure_2514441_presence_24h | Critical care evaluation and management; first 30–74 minutes | 0.004087 |
| Baseline + procedure burden | procedure_2313814_count24h | Routine 12-lead ECG; with interpretation and report | 0.003255 |
| Baseline + procedure burden | procedure_2147482991_count24h | LINE CARE \| LINE STATUS \| LINE STATUS | 0.002948 |
| Baseline + procedure burden | procedure_2313816_count24h | Routine 12-lead ECG; interpretation and report only | 0.002663 |
| Baseline + procedure burden | procedure_2147483387_count24h | GASTROINTESTINAL \| ABDOMEN \| Abdomen INSPECTION | 0.002651 |
| Baseline + procedure burden | procedure_2514441_count24h | Critical care evaluation and management; first 30–74 minutes | 0.002109 |
| Baseline + procedure burden | procedure_725068_presence_24h | Radiologic examination, chest; single view | 0.001631 |
| Baseline + procedure burden | procedure_2147482992_count24h | LINE CARE \| LINE SITE ASSESSMENT \| LINE SITE ASSESSMENT | 0.001509 |
| Baseline + procedure burden | visit_type_Emergency Room and Inpatient Visit | Not applicable | 0.001304 |
| Baseline + procedure burden | procedure_2147483056_count24h | ASSISTANCE \| LEVEL OF ASSISTANCE \| LEVEL OF ASSISTANCE | 0.000865 |
| Baseline + procedure burden | race_category_Other | Not applicable | 0.000778 |
| Baseline + procedure burden | has_prior_visit | Not applicable | 0.000691 |
| Baseline + procedure burden | procedure_2147483387_presence_24h | GASTROINTESTINAL \| ABDOMEN \| Abdomen INSPECTION | 0.000683 |
| Baseline + procedure burden | procedure_2147483603_presence_24h | HEENT \| Eyes/Vision \| Left Eye Assessment | 0.000679 |

Continued on next page

eTable 20. CHoRUS SHAP feature importance for Baseline + procedure burden. (continued)

| Feature matrix | feature | concept name | mean_absolute_shap |
|---|---|---|---|
| Baseline + procedure burden | procedure_2147482962_presence_24h | Surgical Airway \| Airway \| Airway Measured From | 0.000651 |
| Baseline + procedure burden | visit_type_Emergency Room - Hospital | Not applicable | 0.000407 |
| Baseline + procedure burden | sex_category_Female | Not applicable | 0.000360 |
| Baseline + procedure burden | sex_category_Male | Not applicable | 0.000344 |
| Baseline + procedure burden | ethnicity_category_Non-Hispanic | Not applicable | 0.000325 |
| Baseline + procedure burden | race_category_White | Not applicable | 0.000160 |
| Baseline + procedure burden | procedure_2313814_presence_24h | Routine 12-lead ECG; with interpretation and report | 0.000138 |
| Baseline + procedure burden | race_category_Unknown | Not applicable | 0.000113 |
| Baseline + procedure burden | race_category_Black | Not applicable | 0.000089 |
| Baseline + procedure burden | visit_type_Observation Room | Not applicable | 0.000047 |
| Baseline + procedure burden | race_category_Asian | Not applicable | 0.000037 |
| Baseline + procedure burden | ethnicity_category_Hispanic | Not applicable | 0.000029 |
| Baseline + procedure burden | procedure_2313816_presence_24h | Routine 12-lead ECG; interpretation and report only | 0.000016 |
| Baseline + procedure burden | ethnicity_category_Unknown | Not applicable | 0.000000 |
| Baseline + procedure burden | visit_type_Inpatient Hospital | Not applicable | 0.000000 |
| Baseline + procedure burden | procedure_2147483590_presence_24h | HEENT \| Eyes/Vision \| Right Eye Assessment | 0.000000 |
| Baseline + procedure burden | procedure_2147483056_presence_24h | ASSISTANCE \| LEVEL OF ASSISTANCE \| LEVEL OF ASSISTANCE | 0.000000 |
| Baseline + procedure burden | procedure_2147482991_presence_24h | LINE CARE \| LINE STATUS \| LINE STATUS | 0.000000 |
| Baseline + procedure burden | ethnicity_category_Other | Not applicable | 0.000000 |
| Baseline + procedure burden | visit_type_Hospital | Not applicable | 0.000000 |

*Notes:* Rows, feature order, concept names, and mean absolute SHAP values are reproduced from the formatted CHoRUS SHAP workbook. SHAP values were generated from the best-performing algorithm selected for each feature matrix. Best-performing algorithms were selected by highest mean cross-validated AUPRC, with mean AUROC used as the tie-breaker. SHAP values are model-specific attributions, not causal effects; absolute SHAP magnitudes should not be compared directly across separately trained models or datasets.

eTable 21. CHoRUS SHAP feature importance for Baseline + physiological severity + treatment exposure.

| Feature matrix | feature | concept name | mean_absolute_shap |
|---|---|---|---|
| Baseline + physiological severity + treatment exposure | age_at_visit | Not applicable | 0.014128 |
| Baseline + physiological severity + treatment exposure | measurement_3037318_max_24h | Physical activity Braden scale — maximum, first 24 hours | 0.011702 |
| Baseline + physiological severity + treatment exposure | measurement_3035206_mean_24h | Physical mobility Braden scale — mean, first 24 hours | 0.010944 |
| Baseline + physiological severity + treatment exposure | charlson_index | Not applicable | 0.010007 |
| Baseline + physiological severity + treatment exposure | measurement_3035816_mean_24h | Nutrition intake pattern Braden scale — mean, first 24 hours | 0.009816 |
| Baseline + physiological severity + treatment exposure | measurement_4301868_mean_24h | Pulse rate — mean, first 24 hours | 0.009499 |
| Baseline + physiological severity + treatment exposure | prior_visit_count | Not applicable | 0.009443 |
| Baseline + physiological severity + treatment exposure | measurement_3036098_max_24h | Sensory perception Braden scale — maximum, first 24 hours | 0.008094 |
| Baseline + physiological severity + treatment exposure | measurement_3024171_count_24h | Respiratory rate — measurement count, first 24 hours | 0.007988 |

Continued on next page

eTable 21. CHoRUS SHAP feature importance for Baseline + physiological severity + treatment exposure. (continued)

| Feature matrix | feature | concept name | mean_absolute_shap |
|---|---|---|---|
| Baseline + physiological severity + treatment exposure | measurement_3037347_mean_24h | Friction and shear Braden scale — mean, first 24 hours | 0.007198 |
| Baseline + physiological severity + treatment exposure | drug_19079524_count24h | sodium chloride 9 MG/ML Injectable Solution — exposure count, first 24 hours | 0.005337 |
| Baseline + physiological severity + treatment exposure | prior_acute_visit_count | Not applicable | 0.004750 |
| Baseline + physiological severity + treatment exposure | time_to_first_drug_hours | Aggregate treatment-exposure feature | 0.004044 |
| Baseline + physiological severity + treatment exposure | measurement_4020553_std_24h | Oxygen saturation measurement — standard deviation, first 24 hours | 0.003931 |
| Baseline + physiological severity + treatment exposure | total_drug_exposure_count_24h | Aggregate treatment-exposure feature | 0.003601 |
| Baseline + physiological severity + treatment exposure | repeated_drug_exposure_count_24h | Aggregate treatment-exposure feature | 0.003027 |
| Baseline + physiological severity + treatment exposure | measurement_3004249_min_24h | Systolic blood pressure — minimum, first 24 hours | 0.002978 |
| Baseline + physiological severity + treatment exposure | unique_drug_exposure_count_24h | Aggregate treatment-exposure feature | 0.002469 |
| Baseline + physiological severity + treatment exposure | measurement_3004249_std_24h | Systolic blood pressure — standard deviation, first 24 hours | 0.002359 |
| Baseline + physiological severity + treatment exposure | race_category_White | Not applicable | 0.001928 |
| Baseline + physiological severity + treatment exposure | race_category_Other | Not applicable | 0.001871 |
| Baseline + physiological severity + treatment exposure | drug_40220388_count24h | 100 ML propofol 10 MG/ML Injection [Diprivan] — exposure count, first 24 hours | 0.001861 |
| Baseline + physiological severity + treatment exposure | has_prior_visit | Not applicable | 0.001798 |
| Baseline + physiological severity + treatment exposure | visit_type_Emergency Room - Hospital | Not applicable | 0.001760 |
| Baseline + physiological severity + treatment exposure | measurement_4301868_std_24h | Pulse rate — standard deviation, first 24 hours | 0.001572 |
| Baseline + physiological severity + treatment exposure | drug_19102651_count24h | sennosides, USP 8.6 MG Oral Tablet [Senokot] — exposure count, first 24 hours | 0.001388 |
| Baseline + physiological severity + treatment exposure | measurement_3004249_mean_24h | Systolic blood pressure — mean, first 24 hours | 0.001375 |
| Baseline + physiological severity + treatment exposure | visit_type_Emergency Room and Inpatient Visit | Not applicable | 0.001302 |
| Baseline + physiological severity + treatment exposure | drug_42479436_count24h | 1000 ML Sodium Chloride 9 MG/ML Injectable Solution — exposure count, first 24 hours | 0.001235 |
| Baseline + physiological severity + treatment exposure | visit_type_Observation Room | Not applicable | 0.001185 |
| Baseline + physiological severity + treatment exposure | measurement_4301868_min_24h | Pulse rate — minimum, first 24 hours | 0.001175 |
| Baseline + physiological severity + treatment exposure | drug_46275280_count24h | 50 ML magnesium sulfate 40 MG/ML Injection — exposure count, first 24 hours | 0.001168 |
| Baseline + physiological severity + treatment exposure | sex_category_Female | Not applicable | 0.001068 |

Continued on next page

eTable 21. CHoRUS SHAP feature importance for Baseline + physiological severity + treatment exposure. (continued)

| Feature matrix | feature | concept name | mean_absolute_shap |
|---|---|---|---|
| Baseline + physiological severity + treatment exposure | visit_type_Hospital | Not applicable | 0.000994 |
| Baseline + physiological severity + treatment exposure | measurement_4020553_count_24h | Oxygen saturation measurement — measurement count, first 24 hours | 0.000946 |
| Baseline + physiological severity + treatment exposure | drug_19135374_count24h | calcium chloride 0.2 MG/ML / potassium chloride 0.3 MG/ML / sodium chloride 6 MG/ML / sodium lactate 3.1 MG/ML Injectable Solution — exposure count, first 24 hours | 0.000916 |
| Baseline + physiological severity + treatment exposure | measurement_4301868_count_24h | Pulse rate — measurement count, first 24 hours | 0.000868 |
| Baseline + physiological severity + treatment exposure | measurement_4020553_mean_24h | Oxygen saturation measurement — mean, first 24 hours | 0.000842 |
| Baseline + physiological severity + treatment exposure | drug_43011850_count24h | heparin sodium, porcine 5000 UNT/ML Injectable Solution — exposure count, first 24 hours | 0.000800 |
| Baseline + physiological severity + treatment exposure | drug_42708658_count24h | docusate sodium 100 MG Oral Capsule [Colace] — exposure count, first 24 hours | 0.000798 |
| Baseline + physiological severity + treatment exposure | measurement_4301868_max_24h | Pulse rate — maximum, first 24 hours | 0.000726 |
| Baseline + physiological severity + treatment exposure | visit_type_Inpatient Hospital | Not applicable | 0.000697 |
| Baseline + physiological severity + treatment exposure | drug_40220390_count24h | propofol 10 MG/ML Injection [Diprivan] — exposure count, first 24 hours | 0.000658 |
| Baseline + physiological severity + treatment exposure | race_category_Black | Not applicable | 0.000545 |
| Baseline + physiological severity + treatment exposure | drug_1127433_count24h | acetaminophen 325 MG Oral Tablet — exposure count, first 24 hours | 0.000526 |
| Baseline + physiological severity + treatment exposure | ethnicity_category_Hispanic | Not applicable | 0.000472 |
| Baseline + physiological severity + treatment exposure | measurement_4020553_min_24h | Oxygen saturation measurement — minimum, first 24 hours | 0.000458 |
| Baseline + physiological severity + treatment exposure | sex_category_Male | Not applicable | 0.000449 |
| Baseline + physiological severity + treatment exposure | drug_35605482_count24h | 2 ML ondansetron 2 MG/ML Injection — exposure count, first 24 hours | 0.000430 |
| Baseline + physiological severity + treatment exposure | drug_40221385_exposed_24h | 100 ML sodium chloride 9 MG/ML Injection — exposure indicator, first 24 hours | 0.000427 |
| Baseline + physiological severity + treatment exposure | drug_40220357_exposed_24h | 1000 ML sodium chloride 9 MG/ML Injection — exposure indicator, first 24 hours | 0.000393 |
| Baseline + physiological severity + treatment exposure | drug_19127213_count24h | 10 ML sodium chloride 9 MG/ML Prefilled Syringe — exposure count, first 24 hours | 0.000358 |
| Baseline + physiological severity + treatment exposure | drug_43011850_exposed_24h | heparin sodium, porcine 5000 UNT/ML Injectable Solution — exposure indicator, first 24 hours | 0.000356 |
| Baseline + physiological severity + treatment exposure | ethnicity_category_Non-Hispanic | Not applicable | 0.000352 |
| Baseline + physiological severity + treatment exposure | measurement_4020553_max_24h | Oxygen saturation measurement — maximum, first 24 hours | 0.000336 |
| Baseline + physiological severity + treatment exposure | drug_21050327_count24h | Glucose Injectable Solution [Plasma-lyte] — exposure count, first 24 hours | 0.000296 |

Continued on next page

eTable 21. CHoRUS SHAP feature importance for Baseline + physiological severity + treatment exposure. (continued)

| Feature matrix | feature | concept name | mean_absolute_shap |
|---|---|---|---|
| Baseline + physiological severity + treatment exposure | race_category_Unknown | Not applicable | 0.000136 |
| Baseline + physiological severity + treatment exposure | drug_19135374_exposed_24h | calcium chloride 0.2 MG/ML / potassium chloride 0.3 MG/ML / sodium chloride 6 MG/ML / sodium lactate 3.1 MG/ML Injectable Solution — exposure indicator, first 24 hours | 0.000124 |
| Baseline + physiological severity + treatment exposure | race_category_Asian | Not applicable | 0.000102 |
| Baseline + physiological severity + treatment exposure | ethnicity_category_Unknown | Not applicable | 0.000067 |
| Baseline + physiological severity + treatment exposure | measurement_4301868_missing_24h | Pulse rate — missingness indicator, first 24 hours | 0.000015 |
| Baseline + physiological severity + treatment exposure | ethnicity_category_Other | Not applicable | 0.000000 |
| Baseline + physiological severity + treatment exposure | measurement_4020553_missing_24h | Oxygen saturation measurement — missingness indicator, first 24 hours | 0.000000 |

*Notes:* Rows, feature order, concept names, and mean absolute SHAP values are reproduced from the formatted CHoRUS SHAP workbook. SHAP values were generated from the best-performing algorithm selected for each feature matrix. Best-performing algorithms were selected by highest mean cross-validated AUPRC, with mean AUROC used as the tie-breaker. SHAP values are model-specific attributions, not causal effects; absolute SHAP magnitudes should not be compared directly across separately trained models or datasets.

eTable 22. CHoRUS SHAP feature importance for Baseline + physiological severity + procedure burden.

| Feature matrix | feature | concept name | mean_absolute_shap |
|---|---|---|---|
| Baseline + physiological severity + procedure burden | measurement_3035206_mean_24h | Physical mobility Braden scale — mean, first 24 hours | 0.021187 |
| Baseline + physiological severity + procedure burden | measurement_3037318_max_24h | Physical activity Braden scale — maximum, first 24 hours | 0.019232 |
| Baseline + physiological severity + procedure burden | age_at_visit | Not applicable | 0.015691 |
| Baseline + physiological severity + procedure burden | charlson_index | Not applicable | 0.015478 |
| Baseline + physiological severity + procedure burden | measurement_4301868_mean_24h | Pulse rate — mean, first 24 hours | 0.012606 |
| Baseline + physiological severity + procedure burden | measurement_3036098_max_24h | Sensory perception Braden scale — maximum, first 24 hours | 0.010762 |
| Baseline + physiological severity + procedure burden | unique_procedure_count_24h | Aggregate procedure-burden feature | 0.010503 |
| Baseline + physiological severity + procedure burden | procedure_count_total_24h | Aggregate procedure-burden feature | 0.008953 |
| Baseline + physiological severity + procedure burden | measurement_3037347_mean_24h | Friction and shear Braden scale — mean, first 24 hours | 0.007770 |
| Baseline + physiological severity + procedure burden | prior_visit_count | Not applicable | 0.005777 |
| Baseline + physiological severity + procedure burden | measurement_3035816_mean_24h | Nutrition intake pattern Braden scale — mean, first 24 hours | 0.005725 |
| Baseline + physiological severity + procedure burden | measurement_4020553_std_24h | Oxygen saturation measurement — standard deviation, first 24 hours | 0.005126 |

Continued on next page

eTable 22. CHoRUS SHAP feature importance for Baseline + physiological severity + procedure burden. (continued)

| Feature matrix | feature | concept name | mean_absolute_shap |
|---|---|---|---|
| Baseline + physiological severity + procedure burden | procedure_725068_count24h | Radiologic examination, chest; single view — count, first 24 hours | 0.004478 |
| Baseline + physiological severity + procedure burden | procedure_2514441_count24h | Critical care evaluation and management; first 30–74 minutes — count, first 24 hours | 0.004247 |
| Baseline + physiological severity + procedure burden | prior_acute_visit_count | Not applicable | 0.003928 |
| Baseline + physiological severity + procedure burden | measurement_3004249_std_24h | Systolic blood pressure — standard deviation, first 24 hours | 0.003240 |
| Baseline + physiological severity + procedure burden | measurement_4301868_std_24h | Pulse rate — standard deviation, first 24 hours | 0.002665 |
| Baseline + physiological severity + procedure burden | has_prior_visit | Not applicable | 0.002231 |
| Baseline + physiological severity + procedure burden | measurement_4301868_min_24h | Pulse rate — minimum, first 24 hours | 0.001960 |
| Baseline + physiological severity + procedure burden | measurement_3004249_mean_24h | Systolic blood pressure — mean, first 24 hours | 0.001660 |
| Baseline + physiological severity + procedure burden | race_category_Other | Not applicable | 0.001338 |
| Baseline + physiological severity + procedure burden | measurement_3024171_count_24h | Respiratory rate — measurement count, first 24 hours | 0.001293 |
| Baseline + physiological severity + procedure burden | visit_type_Emergency Room - Hospital | Not applicable | 0.001097 |
| Baseline + physiological severity + procedure burden | measurement_4020553_count_24h | Oxygen saturation measurement — measurement count, first 24 hours | 0.001050 |
| Baseline + physiological severity + procedure burden | measurement_4301868_count_24h | Pulse rate — measurement count, first 24 hours | 0.000901 |
| Baseline + physiological severity + procedure burden | procedure_2514441_presence_24h | Critical care evaluation and management; first 30–74 minutes — presence indicator, first 24 hours | 0.000879 |
| Baseline + physiological severity + procedure burden | measurement_4020553_mean_24h | Oxygen saturation measurement — mean, first 24 hours | 0.000691 |
| Baseline + physiological severity + procedure burden | procedure_2147482991_count24h | LINE CARE \| LINE STATUS \| LINE STATUS — count, first 24 hours | 0.000533 |
| Baseline + physiological severity + procedure burden | procedure_2313816_count24h | Routine 12-lead ECG; interpretation and report only — count, first 24 hours | 0.000511 |
| Baseline + physiological severity + procedure burden | visit_type_Observation Room | Not applicable | 0.000448 |
| Baseline + physiological severity + procedure burden | procedure_2313814_count24h | Routine 12-lead ECG; with interpretation and report — count, first 24 hours | 0.000432 |
| Baseline + physiological severity + procedure burden | measurement_3004249_min_24h | Systolic blood pressure — minimum, first 24 hours | 0.000381 |
| Baseline + physiological severity + procedure burden | race_category_White | Not applicable | 0.000319 |
| Baseline + physiological severity + procedure burden | measurement_4301868_max_24h | Pulse rate — maximum, first 24 hours | 0.000292 |
| Baseline + physiological severity + procedure burden | procedure_2147482992_count24h | LINE CARE \| LINE SITE ASSESSMENT \| LINE SITE ASSESSMENT — count, first 24 hours | 0.000292 |
| Baseline + physiological severity + procedure burden | procedure_2147483056_count24h | ASSISTANCE \| LEVEL OF ASSISTANCE \| LEVEL OF ASSISTANCE — count, first 24 hours | 0.000259 |

Continued on next page

eTable 22. CHoRUS SHAP feature importance for Baseline + physiological severity + procedure burden. (continued)

| Feature matrix | feature | concept name | mean_absolute_shap |
|---|---|---|---|
| Baseline + physiological severity + procedure burden | sex_category_Male | Not applicable | 0.000197 |
| Baseline + physiological severity + procedure burden | race_category_Unknown | Not applicable | 0.000193 |
| Baseline + physiological severity + procedure burden | procedure_725068_presence_24h | Radiologic examination, chest; single view — presence indicator, first 24 hours | 0.000120 |
| Baseline + physiological severity + procedure burden | procedure_2147483603_presence_24h | HEENT \| Eyes/Vision \| Left Eye Assessment — presence indicator, first 24 hours | 0.000109 |
| Baseline + physiological severity + procedure burden | race_category_Black | Not applicable | 0.000083 |
| Baseline + physiological severity + procedure burden | procedure_2147483387_count24h | GASTROINTESTINAL \| ABDOMEN \| Abdomen INSPECTION — count, first 24 hours | 0.000083 |
| Baseline + physiological severity + procedure burden | measurement_4020553_min_24h | Oxygen saturation measurement — minimum, first 24 hours | 0.000046 |
| Baseline + physiological severity + procedure burden | sex_category_Female | Not applicable | 0.000011 |
| Baseline + physiological severity + procedure burden | ethnicity_category_Non-Hispanic | Not applicable | 0.000006 |
| Baseline + physiological severity + procedure burden | procedure_2147482962_presence_24h | Surgical Airway \| Airway \| Airway Measured From — presence indicator, first 24 hours | 0.000006 |
| Baseline + physiological severity + procedure burden | measurement_4020553_max_24h | Oxygen saturation measurement — maximum, first 24 hours | 0.000000 |
| Baseline + physiological severity + procedure burden | race_category_Asian | Not applicable | 0.000000 |
| Baseline + physiological severity + procedure burden | ethnicity_category_Other | Not applicable | 0.000000 |
| Baseline + physiological severity + procedure burden | ethnicity_category_Unknown | Not applicable | 0.000000 |
| Baseline + physiological severity + procedure burden | visit_type_Emergency Room and Inpatient Visit | Not applicable | 0.000000 |
| Baseline + physiological severity + procedure burden | measurement_4301868_missing_24h | Pulse rate — missingness indicator, first 24 hours | 0.000000 |
| Baseline + physiological severity + procedure burden | visit_type_Hospital | Not applicable | 0.000000 |
| Baseline + physiological severity + procedure burden | measurement_4020553_missing_24h | Oxygen saturation measurement — missingness indicator, first 24 hours | 0.000000 |
| Baseline + physiological severity + procedure burden | procedure_2147483056_presence_24h | ASSISTANCE \| LEVEL OF ASSISTANCE \| LEVEL OF ASSISTANCE — presence indicator, first 24 hours | 0.000000 |
| Baseline + physiological severity + procedure burden | ethnicity_category_Hispanic | Not applicable | 0.000000 |
| Baseline + physiological severity + procedure burden | procedure_2313816_presence_24h | Routine 12-lead ECG; interpretation and report only — presence indicator, first 24 hours | 0.000000 |
| Baseline + physiological severity + procedure burden | procedure_2147483387_presence_24h | GASTROINTESTINAL \| ABDOMEN \| Abdomen INSPECTION — presence indicator, first 24 hours | 0.000000 |
| Baseline + physiological severity + procedure burden | procedure_2147482992_presence_24h | LINE CARE \| LINE SITE ASSESSMENT \| LINE SITE ASSESSMENT — presence indicator, first 24 hours | 0.000000 |

Continued on next page

eTable 22. CHoRUS SHAP feature importance for Baseline + physiological severity + procedure burden. (continued)

| Feature matrix | feature | concept name | mean_absolute_shap |
|---|---|---|---|
| Baseline + physiological severity + procedure burden | procedure_2313814_presence_24h | Routine 12-lead ECG; with interpretation and report — presence indicator, first 24 hours | 0.000000 |
| Baseline + physiological severity + procedure burden | procedure_2147482991_presence_24h | LINE CARE \| LINE STATUS \| LINE STATUS — presence indicator, first 24 hours | 0.000000 |
| Baseline + physiological severity + procedure burden | visit_type_Inpatient Hospital | Not applicable | 0.000000 |
| Baseline + physiological severity + procedure burden | procedure_2147483590_presence_24h | HEENT \| Eyes/Vision \| Right Eye Assessment — presence indicator, first 24 hours | 0.000000 |

*Notes:* Rows, feature order, concept names, and mean absolute SHAP values are reproduced from the formatted CHoRUS SHAP workbook. SHAP values were generated from the best-performing algorithm selected for each feature matrix. Best-performing algorithms were selected by highest mean cross-validated AUPRC, with mean AUROC used as the tie-breaker. SHAP values are model-specific attributions, not causal effects; absolute SHAP magnitudes should not be compared directly across separately trained models or datasets.

eTable 23. CHoRUS SHAP feature importance for Baseline + treatment exposure + procedure burden.

| Feature matrix | feature | concept name | mean_absolute_shap |
|---|---|---|---|
| Baseline + treatment exposure + procedure burden | age_at_visit | Not applicable | 0.015972 |
| Baseline + treatment exposure + procedure burden | unique_procedure_count_24h | Unique procedure count, first 24 hours | 0.011095 |
| Baseline + treatment exposure + procedure burden | charlson_index | Not applicable | 0.010600 |
| Baseline + treatment exposure + procedure burden | prior_visit_count | Not applicable | 0.008756 |
| Baseline + treatment exposure + procedure burden | procedure_725068_count24h | Radiologic examination, chest; single view — count, first 24 hours | 0.008319 |
| Baseline + treatment exposure + procedure burden | procedure_2147482992_presence_24h | LINE CARE \| LINE SITE ASSESSMENT \| LINE SITE ASSESSMENT — presence indicator, first 24 hours | 0.005612 |
| Baseline + treatment exposure + procedure burden | procedure_count_total_24h | Total procedure count, first 24 hours | 0.004256 |
| Baseline + treatment exposure + procedure burden | procedure_2313814_count24h | Routine 12-lead ECG; with interpretation and report — count, first 24 hours | 0.003652 |
| Baseline + treatment exposure + procedure burden | prior_acute_visit_count | Not applicable | 0.003310 |
| Baseline + treatment exposure + procedure burden | procedure_2147482992_count24h | LINE CARE \| LINE SITE ASSESSMENT \| LINE SITE ASSESSMENT — count, first 24 hours | 0.003080 |
| Baseline + treatment exposure + procedure burden | procedure_2147482991_count24h | LINE CARE \| LINE STATUS \| LINE STATUS — count, first 24 hours | 0.003036 |
| Baseline + treatment exposure + procedure burden | procedure_2514441_count24h | Critical care evaluation and management; first 30–74 minutes — count, first 24 hours | 0.002772 |
| Baseline + treatment exposure + procedure burden | procedure_2313816_count24h | Routine 12-lead ECG; interpretation and report only — count, first 24 hours | 0.002735 |
| Baseline + treatment exposure + procedure burden | drug_19079524_count24h | sodium chloride 9 MG/ML Injectable Solution — exposure count, first 24 hours | 0.002353 |
| Baseline + treatment exposure + procedure burden | drug_40220388_count24h | 100 ML propofol 10 MG/ML Injection [Diprivan] — exposure count, first 24 hours | 0.002289 |
| Baseline + treatment exposure + procedure burden | drug_43011850_exposed_24h | heparin sodium, porcine 5000 UNT/ML Injectable Solution — exposure indicator, first 24 hours | 0.002192 |

Continued on next page

eTable 23. CHoRUS SHAP feature importance for Baseline + treatment exposure + procedure burden. (continued)

| Feature matrix | feature | concept name | mean_absolute_shap |
|---|---|---|---|
| Baseline + treatment exposure + procedure burden | has_prior_visit | Not applicable | 0.001944 |
| Baseline + treatment exposure + procedure burden | procedure_2147483387_presence_24h | GASTROINTESTINAL \| ABDOMEN \| Abdomen INSPECTION — presence indicator, first 24 hours | 0.001577 |
| Baseline + treatment exposure + procedure burden | time_to_first_drug_hours | Time to first drug exposure, hours | 0.001568 |
| Baseline + treatment exposure + procedure burden | drug_40220390_count24h | propofol 10 MG/ML Injection [Diprivan] — exposure count, first 24 hours | 0.001481 |
| Baseline + treatment exposure + procedure burden | procedure_2147483387_count24h | GASTROINTESTINAL \| ABDOMEN \| Abdomen INSPECTION — count, first 24 hours | 0.001446 |
| Baseline + treatment exposure + procedure burden | procedure_2514441_presence_24h | Critical care evaluation and management; first 30–74 minutes — presence indicator, first 24 hours | 0.001226 |
| Baseline + treatment exposure + procedure burden | drug_43011850_count24h | heparin sodium, porcine 5000 UNT/ML Injectable Solution — exposure count, first 24 hours | 0.001092 |
| Baseline + treatment exposure + procedure burden | repeated_drug_exposure_count_24h | Repeated drug-exposure count, first 24 hours | 0.001081 |
| Baseline + treatment exposure + procedure burden | procedure_2147482962_presence_24h | Surgical Airway \| Airway \| Airway Measured From — presence indicator, first 24 hours | 0.000735 |
| Baseline + treatment exposure + procedure burden | procedure_725068_presence_24h | Radiologic examination, chest; single view — presence indicator, first 24 hours | 0.000715 |
| Baseline + treatment exposure + procedure burden | unique_drug_exposure_count_24h | Unique drug-exposure count, first 24 hours | 0.000710 |
| Baseline + treatment exposure + procedure burden | total_drug_exposure_count_24h | Total drug-exposure count, first 24 hours | 0.000680 |
| Baseline + treatment exposure + procedure burden | race_category_Other | Not applicable | 0.000545 |
| Baseline + treatment exposure + procedure burden | procedure_2147483603_presence_24h | HEENT \| Eyes/Vision \| Left Eye Assessment — presence indicator, first 24 hours | 0.000520 |
| Baseline + treatment exposure + procedure burden | procedure_2147483056_count24h | ASSISTANCE \| LEVEL OF ASSISTANCE \| LEVEL OF ASSISTANCE — count, first 24 hours | 0.000361 |
| Baseline + treatment exposure + procedure burden | drug_21050327_count24h | Glucose Injectable Solution [Plasma-lyte] — exposure count, first 24 hours | 0.000357 |
| Baseline + treatment exposure + procedure burden | drug_19135374_count24h | calcium chloride 0.2 MG/ML / potassium chloride 0.3 MG/ML / sodium chloride 6 MG/ML / sodium lactate 3.1 MG/ML Injectable Solution — exposure count, first 24 hours | 0.000308 |
| Baseline + treatment exposure + procedure burden | drug_1127433_count24h | acetaminophen 325 MG Oral Tablet — exposure count, first 24 hours | 0.000306 |
| Baseline + treatment exposure + procedure burden | drug_35605482_count24h | 2 ML ondansetron 2 MG/ML Injection — exposure count, first 24 hours | 0.000252 |
| Baseline + treatment exposure + procedure burden | drug_19102651_count24h | sennosides, USP 8.6 MG Oral Tablet [Senokot] — exposure count, first 24 hours | 0.000225 |
| Baseline + treatment exposure + procedure burden | sex_category_Female | Not applicable | 0.000214 |
| Baseline + treatment exposure + procedure burden | visit_type_Hospital | Not applicable | 0.000187 |
| Baseline + treatment exposure + procedure burden | drug_42708658_count24h | docusate sodium 100 MG Oral Capsule [Colace] — exposure count, first 24 hours | 0.000182 |

Continued on next page

eTable 23. CHoRUS SHAP feature importance for Baseline + treatment exposure + procedure burden. (continued)

| Feature matrix | feature | concept name | mean_absolute_shap |
|---|---|---|---|
| Baseline + treatment exposure + procedure burden | sex_category_Male | Not applicable | 0.000174 |
| Baseline + treatment exposure + procedure burden | visit_type_Observation Room | Not applicable | 0.000149 |
| Baseline + treatment exposure + procedure burden | drug_42479436_count24h | 1000 ML Sodium Chloride 9 MG/ML Injectable Solution — exposure count, first 24 hours | 0.000133 |
| Baseline + treatment exposure + procedure burden | ethnicity_category_Hispanic | Not applicable | 0.000123 |
| Baseline + treatment exposure + procedure burden | race_category_White | Not applicable | 0.000117 |
| Baseline + treatment exposure + procedure burden | ethnicity_category_Non-Hispanic | Not applicable | 0.000099 |
| Baseline + treatment exposure + procedure burden | drug_40220357_exposed_24h | 1000 ML sodium chloride 9 MG/ML Injection — exposure indicator, first 24 hours | 0.000067 |
| Baseline + treatment exposure + procedure burden | race_category_Unknown | Not applicable | 0.000040 |
| Baseline + treatment exposure + procedure burden | visit_type_Inpatient Hospital | Not applicable | 0.000035 |
| Baseline + treatment exposure + procedure burden | drug_46275280_count24h | 50 ML magnesium sulfate 40 MG/ML Injection — exposure count, first 24 hours | 0.000026 |
| Baseline + treatment exposure + procedure burden | procedure_2313814_presence_24h | Routine 12-lead ECG; with interpretation and report — presence indicator, first 24 hours | 0.000019 |
| Baseline + treatment exposure + procedure burden | race_category_Asian | Not applicable | 0.000019 |
| Baseline + treatment exposure + procedure burden | drug_19127213_count24h | 10 ML sodium chloride 9 MG/ML Prefilled Syringe — exposure count, first 24 hours | 0.000010 |
| Baseline + treatment exposure + procedure burden | procedure_2147483590_presence_24h | HEENT \| Eyes/Vision \| Right Eye Assessment — presence indicator, first 24 hours | 0.000000 |
| Baseline + treatment exposure + procedure burden | procedure_2147483056_presence_24h | ASSISTANCE \| LEVEL OF ASSISTANCE \| LEVEL OF ASSISTANCE — presence indicator, first 24 hours | 0.000000 |
| Baseline + treatment exposure + procedure burden | procedure_2313816_presence_24h | Routine 12-lead ECG; interpretation and report only — presence indicator, first 24 hours | 0.000000 |
| Baseline + treatment exposure + procedure burden | drug_19135374_exposed_24h | calcium chloride 0.2 MG/ML / potassium chloride 0.3 MG/ML / sodium chloride 6 MG/ML / sodium lactate 3.1 MG/ML Injectable Solution — exposure indicator, first 24 hours | 0.000000 |
| Baseline + treatment exposure + procedure burden | race_category_Black | Not applicable | 0.000000 |
| Baseline + treatment exposure + procedure burden | ethnicity_category_Unknown | Not applicable | 0.000000 |
| Baseline + treatment exposure + procedure burden | ethnicity_category_Other | Not applicable | 0.000000 |
| Baseline + treatment exposure + procedure burden | drug_40221385_exposed_24h | 100 ML sodium chloride 9 MG/ML Injection — exposure indicator, first 24 hours | 0.000000 |
| Baseline + treatment exposure + procedure burden | procedure_2147482991_presence_24h | LINE CARE \| LINE STATUS \| LINE STATUS — presence indicator, first 24 hours | 0.000000 |

Continued on next page

eTable 23. CHoRUS SHAP feature importance for Baseline + treatment exposure + procedure burden. (continued)

| Feature matrix | feature | concept name | mean_absolute_shap |
|---|---|---|---|
| Baseline + treatment exposure + procedure burden | visit_type_Emergency Room - Hospital | Not applicable | 0.000000 |
| Baseline + treatment exposure + procedure burden | visit_type_Emergency Room and Inpatient Visit | Not applicable | 0.000000 |

*Notes:* Rows, feature order, concept names, and mean absolute SHAP values are reproduced from the formatted CHoRUS SHAP workbook. SHAP values were generated from the best-performing algorithm selected for each feature matrix. Best-performing algorithms were selected by highest mean cross-validated AUPRC, with mean AUROC used as the tie-breaker. SHAP values are model-specific attributions, not causal effects; absolute SHAP magnitudes should not be compared directly across separately trained models or datasets.

eTable 24. CHoRUS SHAP feature importance for Baseline + all clinical domains.

| Feature matrix | feature | concept name | mean_absolute_shap |
|---|---|---|---|
| Baseline + all clinical domains | age_at_visit | Not applicable | 0.014816 |
| Baseline + all clinical domains | measurement_3035206_mean_24h | Physical mobility Braden scale — mean, first 24 hours | 0.014011 |
| Baseline + all clinical domains | measurement_4301868_mean_24h | Pulse rate — mean, first 24 hours | 0.012437 |
| Baseline + all clinical domains | charlson_index | Not applicable | 0.011623 |
| Baseline + all clinical domains | unique_procedure_count_24h | Unique procedure count, first 24 hours | 0.009811 |
| Baseline + all clinical domains | procedure_count_total_24h | Total procedure count, first 24 hours | 0.007586 |
| Baseline + all clinical domains | measurement_3037347_mean_24h | Friction and shear Braden scale — mean, first 24 hours | 0.006524 |
| Baseline + all clinical domains | measurement_3037318_max_24h | Physical activity Braden scale — maximum, first 24 hours | 0.006506 |
| Baseline + all clinical domains | prior_visit_count | Not applicable | 0.006369 |
| Baseline + all clinical domains | measurement_3035816_mean_24h | Nutrition intake pattern Braden scale — mean, first 24 hours | 0.006291 |
| Baseline + all clinical domains | measurement_3036098_max_24h | Sensory perception Braden scale — maximum, first 24 hours | 0.005521 |
| Baseline + all clinical domains | measurement_4020553_std_24h | Oxygen saturation measurement — standard deviation, first 24 hours | 0.005324 |
| Baseline + all clinical domains | drug_19079524_count24h | sodium chloride 9 MG/ML Injectable Solution — exposure count, first 24 hours | 0.003738 |
| Baseline + all clinical domains | prior_acute_visit_count | Not applicable | 0.003594 |
| Baseline + all clinical domains | procedure_725068_count24h | Radiologic examination, chest; single view — count, first 24 hours | 0.003415 |
| Baseline + all clinical domains | time_to_first_drug_hours | Time to first drug exposure, hours | 0.003003 |
| Baseline + all clinical domains | procedure_2514441_count24h | Critical care evaluation and management; first 30–74 minutes — count, first 24 hours | 0.002798 |
| Baseline + all clinical domains | measurement_3004249_mean_24h | Systolic blood pressure — mean, first 24 hours | 0.002796 |
| Baseline + all clinical domains | unique_drug_exposure_count_24h | Unique drug-exposure count, first 24 hours | 0.002129 |
| Baseline + all clinical domains | has_prior_visit | Not applicable | 0.002042 |
| Baseline + all clinical domains | drug_43011850_exposed_24h | heparin sodium, porcine 5000 UNT/ML Injectable Solution — exposure indicator, first 24 hours | 0.001640 |
| Baseline + all clinical domains | measurement_4301868_min_24h | Pulse rate — minimum, first 24 hours | 0.001528 |
| Baseline + all clinical domains | visit_type_Emergency Room - Hospital | Not applicable | 0.001503 |
| Baseline + all clinical domains | measurement_3004249_std_24h | Systolic blood pressure — standard deviation, first 24 hours | 0.001468 |

Continued on next page

eTable 24. CHoRUS SHAP feature importance for Baseline + all clinical domains. (continued)

| Feature matrix | feature | concept name | mean_absolute_shap |
|---|---|---|---|
| Baseline + all clinical domains | procedure_2514441_presence_24h | Critical care evaluation and management; first 30–74 minutes — presence indicator, first 24 hours | 0.001297 |
| Baseline + all clinical domains | measurement_4301868_std_24h | Pulse rate — standard deviation, first 24 hours | 0.001145 |
| Baseline + all clinical domains | race_category_Other | Not applicable | 0.001088 |
| Baseline + all clinical domains | measurement_4301868_count_24h | Pulse rate — measurement count, first 24 hours | 0.001087 |
| Baseline + all clinical domains | measurement_4020553_count_24h | Oxygen saturation measurement — measurement count, first 24 hours | 0.001067 |
| Baseline + all clinical domains | measurement_3024171_count_24h | Respiratory rate — measurement count, first 24 hours | 0.000960 |
| Baseline + all clinical domains | repeated_drug_exposure_count_24h | Repeated drug-exposure count, first 24 hours | 0.000841 |
| Baseline + all clinical domains | total_drug_exposure_count_24h | Total drug-exposure count, first 24 hours | 0.000791 |
| Baseline + all clinical domains | measurement_4020553_mean_24h | Oxygen saturation measurement — mean, first 24 hours | 0.000630 |
| Baseline + all clinical domains | drug_40220388_count24h | 100 ML propofol 10 MG/ML Injection [Diprivan] — exposure count, first 24 hours | 0.000629 |
| Baseline + all clinical domains | drug_35605482_count24h | 2 ML ondansetron 2 MG/ML Injection — exposure count, first 24 hours | 0.000560 |
| Baseline + all clinical domains | procedure_2147482991_count24h | LINE CARE \| LINE STATUS \| LINE STATUS — count, first 24 hours | 0.000449 |
| Baseline + all clinical domains | drug_19102651_count24h | sennosides, USP 8.6 MG Oral Tablet [Senokot] — exposure count, first 24 hours | 0.000335 |
| Baseline + all clinical domains | measurement_3004249_min_24h | Systolic blood pressure — minimum, first 24 hours | 0.000333 |
| Baseline + all clinical domains | drug_46275280_count24h | 50 ML magnesium sulfate 40 MG/ML Injection — exposure count, first 24 hours | 0.000327 |
| Baseline + all clinical domains | drug_40220390_count24h | propofol 10 MG/ML Injection [Diprivan] — exposure count, first 24 hours | 0.000305 |
| Baseline + all clinical domains | procedure_2147482992_count24h | LINE CARE \| LINE SITE ASSESSMENT \| LINE SITE ASSESSMENT — count, first 24 hours | 0.000292 |
| Baseline + all clinical domains | measurement_4301868_max_24h | Pulse rate — maximum, first 24 hours | 0.000242 |
| Baseline + all clinical domains | visit_type_Inpatient Hospital | Not applicable | 0.000222 |
| Baseline + all clinical domains | visit_type_Observation Room | Not applicable | 0.000219 |
| Baseline + all clinical domains | drug_40221385_exposed_24h | 100 ML sodium chloride 9 MG/ML Injection — exposure indicator, first 24 hours | 0.000185 |
| Baseline + all clinical domains | procedure_2147482962_presence_24h | Surgical Airway \| Airway \| Airway Measured From — presence indicator, first 24 hours | 0.000166 |
| Baseline + all clinical domains | drug_40220357_exposed_24h | 1000 ML sodium chloride 9 MG/ML Injection — exposure indicator, first 24 hours | 0.000149 |
| Baseline + all clinical domains | procedure_2147483056_count24h | ASSISTANCE \| LEVEL OF ASSISTANCE \| LEVEL OF ASSISTANCE — count, first 24 hours | 0.000110 |
| Baseline + all clinical domains | race_category_White | Not applicable | 0.000108 |
| Baseline + all clinical domains | measurement_4020553_min_24h | Oxygen saturation measurement — minimum, first 24 hours | 0.000102 |
| Baseline + all clinical domains | procedure_2147483603_presence_24h | HEENT \| Eyes/Vision \| Left Eye Assessment — presence indicator, first 24 hours | 0.000076 |
| Baseline + all clinical domains | drug_19135374_count24h | calcium chloride 0.2 MG/ML / potassium chloride 0.3 MG/ML / sodium chloride 6 MG/ML / sodium lactate 3.1 MG/ML Injectable Solution — exposure count, first 24 hours | 0.000063 |

Continued on next page

eTable 24. CHoRUS SHAP feature importance for Baseline + all clinical domains. (continued)

| Feature matrix | feature | concept name | mean_absolute_shap |
|---|---|---|---|
| Baseline + all clinical domains | race_category_Asian | Not applicable | 0.000040 |
| Baseline + all clinical domains | drug_1127433_count24h | acetaminophen 325 MG Oral Tablet — exposure count, first 24 hours | 0.000040 |
| Baseline + all clinical domains | drug_21050327_count24h | Glucose Injectable Solution [Plasma-lyte] — exposure count, first 24 hours | 0.000035 |
| Baseline + all clinical domains | sex_category_Male | Not applicable | 0.000012 |
| Baseline + all clinical domains | procedure_2313816_count24h | Routine 12-lead ECG; interpretation and report only — count, first 24 hours | 0.000010 |
| Baseline + all clinical domains | drug_43011850_count24h | heparin sodium, porcine 5000 UNT/ML Injectable Solution — exposure count, first 24 hours | 0.000009 |
| Baseline + all clinical domains | drug_42479436_count24h | 1000 ML Sodium Chloride 9 MG/ML Injectable Solution — exposure count, first 24 hours | 0.000007 |
| Baseline + all clinical domains | procedure_2147483387_count24h | GASTROINTESTINAL \| ABDOMEN \| Abdomen INSPECTION — count, first 24 hours | 0.000001 |
| Baseline + all clinical domains | procedure_2147483590_presence_24h | HEENT \| Eyes/Vision \| Right Eye Assessment — presence indicator, first 24 hours | 0.000000 |
| Baseline + all clinical domains | drug_19135374_exposed_24h | calcium chloride 0.2 MG/ML / potassium chloride 0.3 MG/ML / sodium chloride 6 MG/ML / sodium lactate 3.1 MG/ML Injectable Solution — exposure indicator, first 24 hours | 0.000000 |
| Baseline + all clinical domains | drug_19127213_count24h | 10 ML sodium chloride 9 MG/ML Prefilled Syringe — exposure count, first 24 hours | 0.000000 |
| Baseline + all clinical domains | measurement_4301868_missing_24h | Pulse rate — missingness indicator, first 24 hours | 0.000000 |
| Baseline + all clinical domains | measurement_4020553_max_24h | Oxygen saturation measurement — maximum, first 24 hours | 0.000000 |
| Baseline + all clinical domains | measurement_4020553_missing_24h | Oxygen saturation measurement — missingness indicator, first 24 hours | 0.000000 |
| Baseline + all clinical domains | sex_category_Female | Not applicable | 0.000000 |
| Baseline + all clinical domains | race_category_Black | Not applicable | 0.000000 |
| Baseline + all clinical domains | race_category_Unknown | Not applicable | 0.000000 |
| Baseline + all clinical domains | ethnicity_category_Hispanic | Not applicable | 0.000000 |
| Baseline + all clinical domains | ethnicity_category_Non-Hispanic | Not applicable | 0.000000 |
| Baseline + all clinical domains | ethnicity_category_Unknown | Not applicable | 0.000000 |
| Baseline + all clinical domains | ethnicity_category_Other | Not applicable | 0.000000 |
| Baseline + all clinical domains | visit_type_Emergency Room and Inpatient Visit | Not applicable | 0.000000 |
| Baseline + all clinical domains | visit_type_Hospital | Not applicable | 0.000000 |
| Baseline + all clinical domains | procedure_2147482991_presence_24h | LINE CARE \| LINE STATUS \| LINE STATUS — presence indicator, first 24 hours | 0.000000 |
| Baseline + all clinical domains | procedure_2313814_count24h | Routine 12-lead ECG; with interpretation and report — count, first 24 hours | 0.000000 |
| Baseline + all clinical domains | procedure_2313814_presence_24h | Routine 12-lead ECG; with interpretation and report — presence indicator, first 24 hours | 0.000000 |
| Baseline + all clinical domains | procedure_2147482992_presence_24h | LINE CARE \| LINE SITE ASSESSMENT \| LINE SITE ASSESSMENT — presence indicator, first 24 hours | 0.000000 |
| Baseline + all clinical domains | procedure_2147483387_presence_24h | GASTROINTESTINAL \| ABDOMEN \| Abdomen INSPECTION — presence indicator, first 24 hours | 0.000000 |

Continued on next page

eTable 24. CHoRUS SHAP feature importance for Baseline + all clinical domains. (continued)

| Feature matrix | feature | concept name | mean_absolute_shap |
|---|---|---|---|
| Baseline + all clinical domains | procedure_725068_presence_24h | Radiologic examination, chest; single view — presence indicator, first 24 hours | 0.000000 |
| Baseline + all clinical domains | procedure_2313816_presence_24h | Routine 12-lead ECG; interpretation and report only — presence indicator, first 24 hours | 0.000000 |
| Baseline + all clinical domains | procedure_2147483056_presence_24h | ASSISTANCE \| LEVEL OF ASSISTANCE \| LEVEL OF ASSISTANCE — presence indicator, first 24 hours | 0.000000 |
| Baseline + all clinical domains | drug_42708658_count24h | docusate sodium 100 MG Oral Capsule [Colace] — exposure count, first 24 hours | 0.000000 |

*Notes:* Rows, feature order, concept names, and mean absolute SHAP values are reproduced from the formatted CHoRUS SHAP workbook. SHAP values were generated from the best-performing algorithm selected for each feature matrix. Best-performing algorithms were selected by highest mean cross-validated AUPRC, with mean AUROC used as the tie-breaker. SHAP values are model-specific attributions, not causal effects; absolute SHAP magnitudes should not be compared directly across separately trained models or datasets.

eTable 25. MIMIC-IV SHAP feature importance for Baseline.

| Model | Rank or source order | Feature domain | Original feature name | Readable concept or feature name | Mean absolute SHAP |
|---|---|---|---|---|---|
| LogisticRegression | 1 | | `age_at_visit` | Age at visit | 0.020071 |
| LogisticRegression | 2 | | `visit_type` | Visit type | 0.015933 |
| LogisticRegression | 3 | | `charlson_index` | Charlson Comorbidity Index | 0.015100 |
| LogisticRegression | 4 | | `prior_visit_count` | Prior visit count | 0.010238 |
| LogisticRegression | 5 | | `prior_acute_visit_count` | Prior acute-care visit count | 0.007333 |
| LogisticRegression | 6 | | sex | Sex | 0.004526 |
| LogisticRegression | 7 | | ethnicity | Ethnicity | 0.004509 |
| LogisticRegression | 8 | | `has_prior_visit` | Has prior visit | 0.003386 |
| LogisticRegression | 9 | | race | Race | 0.003181 |

*Notes:* Rows are reproduced from the saved MIMIC-IV SHAP summary output. When a rank column was absent in the source file, source row order is shown. SHAP values were generated from the best-performing algorithm selected for each feature matrix. Best-performing algorithms were selected by highest mean cross-validated AUPRC, with mean AUROC used as the tie-breaker. SHAP values are model-specific attributions, not causal effects; absolute SHAP magnitudes should not be compared directly across separately trained models or datasets.

eTable 26. MIMIC-IV SHAP feature importance for Baseline + physiological severity.

| Model | Rank or source order | Feature domain | Original feature name | Readable concept or feature name | Mean absolute SHAP |
|---|---|---|---|---|---|
| RandomForest | 1 | baseline | `age_at_visit` | Age at visit | 0.013207 |
| RandomForest | 2 | baseline | `charlson_index` | Charlson Comorbidity Index | 0.007225 |
| RandomForest | 3 | baseline | `visit_type` | Visit type | 0.005841 |
| RandomForest | 4 | physiological severity | `lab_51277_rdw_mean` | RDW - mean, first 24 hours | 0.005062 |
| RandomForest | 5 | physiological severity | `lab_51222_hemoglobin_max` | Hemoglobin - maximum, first 24 hours | 0.005010 |
| RandomForest | 6 | physiological severity | `lab_51006_urea_nitrogen_max` | Urea Nitrogen - maximum, first 24 hours | 0.004930 |
| RandomForest | 7 | physiological severity | `lab_51301_white_blood_cells_max` | White Blood Cells - maximum, first 24 hours | 0.004853 |
| RandomForest | 8 | physiological severity | `lab_51277_rdw_max` | RDW - maximum, first 24 hours | 0.004586 |
| RandomForest | 9 | physiological severity | `lab_50983_sodium_max` | Sodium - maximum, first 24 hours | 0.004390 |

Continued on next page

eTable 26. MIMIC-IV SHAP feature importance for Baseline + physiological severity. (continued)

| Model | Rank or source order | Feature domain | Original feature name | Readable concept or feature name | Mean absolute SHAP |
|---|---|---|---|---|---|
| RandomForest | 10 | physiological severity | `lab_51006_urea_nitrogen_mean` | Urea Nitrogen - mean, first 24 hours | 0.004163 |
| RandomForest | 11 | physiological severity | `lab_50983_sodium_mean` | Sodium - mean, first 24 hours | 0.004150 |
| RandomForest | 12 | physiological severity | `lab_50813_lactate_mean` | Lactate - mean, first 24 hours | 0.003841 |
| RandomForest | 13 | physiological severity | `lab_51274_pt_mean` | PT - mean, first 24 hours | 0.003769 |
| RandomForest | 14 | physiological severity | `lab_51006_urea_nitrogen_min` | Urea Nitrogen - minimum, first 24 hours | 0.003207 |
| RandomForest | 15 | physiological severity | `lab_50912_creatinine_mean` | Creatinine - mean, first 24 hours | 0.003176 |
| RandomForest | 16 | physiological severity | `lab_50983_sodium_min` | Sodium - minimum, first 24 hours | 0.003152 |
| RandomForest | 17 | physiological severity | `lab_51277_rdw_min` | RDW - minimum, first 24 hours | 0.002951 |
| RandomForest | 18 | physiological severity | `lab_51237_inr_pt_mean` | INR(PT) - mean, first 24 hours | 0.002886 |
| RandomForest | 19 | physiological severity | `lab_51274_pt_max` | PT - maximum, first 24 hours | 0.002809 |
| RandomForest | 20 | physiological severity | `lab_51237_inr_pt_max` | INR(PT) - maximum, first 24 hours | 0.002791 |
| RandomForest | 21 | physiological severity | `lab_50954_lactate_dehydrogenase_ld_min` | Lactate Dehydrogenase (LD) - minimum, first 24 hours | 0.002714 |
| RandomForest | 22 | physiological severity | `lab_50893_calcium_total_max` | Calcium, Total - maximum, first 24 hours | 0.002585 |
| RandomForest | 23 | physiological severity | `lab_50813_lactate_min` | Lactate - minimum, first 24 hours | 0.002545 |
| RandomForest | 24 | baseline | race | Race | 0.002521 |
| RandomForest | 25 | physiological severity | `lab_50882_bicarbonate_mean` | Bicarbonate - mean, first 24 hours | 0.002452 |
| RandomForest | 26 | physiological severity | `lab_50818_pco2_mean` | pCO2 - mean, first 24 hours | 0.002425 |
| RandomForest | 27 | physiological severity | `lab_50882_bicarbonate_std` | Bicarbonate - standard deviation, first 24 hours | 0.002335 |
| RandomForest | 28 | physiological severity | `lab_50893_calcium_total_mean` | Calcium, Total - mean, first 24 hours | 0.002233 |
| RandomForest | 29 | physiological severity | `lab_50893_calcium_total_min` | Calcium, Total - minimum, first 24 hours | 0.002204 |
| RandomForest | 30 | physiological severity | `lab_50813_lactate_max` | Lactate - maximum, first 24 hours | 0.002187 |
| RandomForest | 31 | physiological severity | `lab_50820_ph_min` | pH - minimum, first 24 hours | 0.002119 |
| RandomForest | 32 | baseline | sex | Sex | 0.002065 |
| RandomForest | 33 | physiological severity | `lab_50818_pco2_min` | pCO2 - minimum, first 24 hours | 0.002029 |
| RandomForest | 34 | physiological severity | `lab_50983_sodium_count` | Sodium - count, first 24 hours | 0.002022 |

Continued on next page

eTable 26. MIMIC-IV SHAP feature importance for Baseline + physiological severity. (continued)

| Model | Rank or source order | Feature domain | Original feature name | Readable concept or feature name | Mean absolute SHAP |
|---|---|---|---|---|---|
| RandomForest | 35 | baseline | ethnicity | Ethnicity | 0.001992 |
| RandomForest | 36 | baseline | prior_visit_count | Prior visit count | 0.001784 |
| RandomForest | 37 | physiological severity | lab_50820_ph_mean | pH - mean, first 24 hours | 0.001736 |
| RandomForest | 38 | physiological severity | lab_50820_ph_std | pH - standard deviation, first 24 hours | 0.001735 |
| RandomForest | 39 | physiological severity | lab_50818_pco2_std | pCO2 - standard deviation, first 24 hours | 0.001671 |
| RandomForest | 40 | physiological severity | lab_50931_glucose_count | Glucose - count, first 24 hours | 0.001628 |
| RandomForest | 41 | physiological severity | lab_50820_ph_max | pH - maximum, first 24 hours | 0.001611 |
| RandomForest | 42 | physiological severity | lab_50983_sodium_std | Sodium - standard deviation, first 24 hours | 0.001526 |
| RandomForest | 43 | physiological severity | lab_50818_pco2_max | pCO2 - maximum, first 24 hours | 0.001488 |
| RandomForest | 44 | physiological severity | lab_50970_phosphate_count | Phosphate - count, first 24 hours | 0.001410 |
| RandomForest | 45 | baseline | prior_acute_visit_count | Prior acute-care visit count | 0.001389 |
| RandomForest | 46 | physiological severity | lab_50893_calcium_total_count | Calcium, Total - count, first 24 hours | 0.001361 |
| RandomForest | 47 | physiological severity | lab_50821_po2_min | pO2 - minimum, first 24 hours | 0.001282 |
| RandomForest | 48 | physiological severity | lab_50804_calculated_total_co2_mean | Calculated Total CO2 - mean, first 24 hours | 0.001252 |
| RandomForest | 49 | physiological severity | lab_50971_potassium_count | Potassium - count, first 24 hours | 0.001215 |
| RandomForest | 50 | physiological severity | lab_50804_calculated_total_co2_std | Calculated Total CO2 - standard deviation, first 24 hours | 0.001209 |
| RandomForest | 51 | physiological severity | lab_50821_po2_mean | pO2 - mean, first 24 hours | 0.001030 |
| RandomForest | 52 | physiological severity | lab_50821_po2_std | pO2 - standard deviation, first 24 hours | 0.000912 |
| RandomForest | 53 | physiological severity | lab_50902_chloride_count | Chloride - count, first 24 hours | 0.000906 |
| RandomForest | 54 | physiological severity | lab_50821_po2_max | pO2 - maximum, first 24 hours | 0.000733 |
| RandomForest | 55 | baseline | has_prior_visit | Has prior visit | 0.000552 |

*Notes:* Rows are reproduced from the saved MIMIC-IV SHAP summary output. When a rank column was absent in the source file, source row order is shown. SHAP values were generated from the best-performing algorithm selected for each feature matrix. Best-performing algorithms were selected by highest mean cross-validated AUPRC, with mean AUROC used as the tie-breaker. SHAP values are model-specific attributions, not causal effects; absolute SHAP magnitudes should not be compared directly across separately trained models or datasets.

eTable 27. MIMIC-IV SHAP feature importance for Baseline + treatment exposure.

| Model | Rank or source order | Feature domain | Original feature name | Readable concept or feature name | Mean absolute SHAP |
|---|---|---|---|---|---|
| LogisticRegression | 1 | baseline | visit_type | Visit type | 0.021299 |
| LogisticRegression | 2 | baseline | sex | Sex | 0.020440 |

Continued on next page

eTable 27. MIMIC-IV SHAP feature importance for Baseline + treatment exposure. (continued)

| Model | Rank or source order | Feature domain | Original feature name | Readable concept or feature name | Mean absolute SHAP |
|---|---|---|---|---|---|
| LogisticRegression | 3 | baseline | age_at_visit | Age at visit | 0.019454 |
| LogisticRegression | 4 | baseline | prior_visit_count | Prior visit count | 0.015862 |
| LogisticRegression | 5 | baseline | charlson_index | Charlson Comorbidity Index | 0.013065 |
| LogisticRegression | 6 | baseline | prior_acute_visit_count | Prior acute-care visit count | 0.012645 |
| LogisticRegression | 7 | treatment exposure | drug_gsn_060304_4b8ed8c2a4_count_24h | Heparin Flush (10 units/ml) - count, first 24 hours | 0.006687 |
| LogisticRegression | 8 | baseline | ethnicity | Ethnicity | 0.006137 |
| LogisticRegression | 9 | treatment exposure | drug_formulary_vial_b4c8f642b1_exposed_24h | Vial - exposure indicator, first 24 hours | 0.006039 |
| LogisticRegression | 10 | baseline | race | Race | 0.005324 |
| LogisticRegression | 11 | treatment exposure | drug_gsn_001210_ba3dc43c8d_exposed_24h | 0.9% Sodium Chloride - exposure indicator, first 24 hours | 0.004117 |
| LogisticRegression | 12 | treatment exposure | repeat_drug_exposure_count_24h | Repeat drug exposure count, first 24 hours | 0.004106 |
| LogisticRegression | 13 | treatment exposure | drug_gsn_001210_ba3dc43c8d_count_24h | 0.9% Sodium Chloride - count, first 24 hours | 0.002849 |
| LogisticRegression | 14 | treatment exposure | drug_formulary_fentsoln50_7775dcade8_exposed_24h | Soln - exposure indicator, first 24 hours | 0.002511 |
| LogisticRegression | 15 | treatment exposure | drug_formulary_vancobase_228466e611_exposed_24h | Iso-Osmotic Dextrose - exposure indicator, first 24 hours | 0.002415 |
| LogisticRegression | 16 | treatment exposure | drug_gsn_001972_b5f0a0eff8_count_24h | 5% Dextrose - count, first 24 hours | 0.002400 |
| LogisticRegression | 17 | treatment exposure | drug_gsn_043952_0602de6ed2_exposed_24h | Vancomycin - exposure indicator, first 24 hours | 0.002368 |
| LogisticRegression | 18 | treatment exposure | drug_gsn_048287_21c090e9e3_exposed_24h | Fentanyl Citrate - exposure indicator, first 24 hours | 0.001986 |
| LogisticRegression | 19 | treatment exposure | drug_gsn_057959_a518f33e58_exposed_24h | Chlorhexidine Gluconate 0.12% Oral Rinse - exposure indicator, first 24 hours | 0.001971 |
| LogisticRegression | 20 | baseline | has_prior_visit | Has prior visit | 0.001944 |
| LogisticRegression | 21 | treatment exposure | drug_formulary_vial_b4c8f642b1_count_24h | Vial - count, first 24 hours | 0.001715 |
| LogisticRegression | 22 | treatment exposure | drug_gsn_057959_a518f33e58_count_24h | Chlorhexidine Gluconate 0.12% Oral Rinse - count, first 24 hours | 0.001692 |
| LogisticRegression | 23 | treatment exposure | any_drug_24h | Any drug exposure, first 24 hours | 0.001515 |
| LogisticRegression | 24 | treatment exposure | drug_formulary_fentsoln50_7775dcade8_count_24h | Soln - count, first 24 hours | 0.001460 |
| LogisticRegression | 25 | treatment exposure | unique_drug_count_24h | Unique drug count, first 24 hours | 0.001450 |
| LogisticRegression | 26 | treatment exposure | drug_gsn_016796_f69d8264fe_exposed_24h | Propofol - exposure indicator, first 24 hours | 0.001349 |
| LogisticRegression | 27 | treatment exposure | drug_gsn_048287_21c090e9e3_count_24h | Fentanyl Citrate - count, first 24 hours | 0.001148 |
| LogisticRegression | 28 | treatment exposure | time_to_first_drug_hours | Time to first drug exposure, hours | 0.000885 |
| LogisticRegression | 29 | treatment exposure | drug_gsn_043952_0602de6ed2_count_24h | Vancomycin - count, first 24 hours | 0.000873 |
| LogisticRegression | 30 | treatment exposure | drug_formulary_vancobase_228466e611_count_24h | Iso-Osmotic Dextrose - count, first 24 hours | 0.000859 |
| LogisticRegression | 31 | treatment exposure | drug_gsn_016796_f69d8264fe_count_24h | Propofol - count, first 24 hours | 0.000653 |

Continued on next page

eTable 27. MIMIC-IV SHAP feature importance for Baseline + treatment exposure. (continued)

| Model | Rank or source order | Feature domain | Original feature name | Readable concept or feature name | Mean absolute SHAP |
|---|---|---|---|---|---|
| LogisticRegression | 32 | treatment exposure | drug_gsn_001972_b5f0a0eff8_exposed_24h | 5% Dextrose - exposure indicator, first 24 hours | 0.000344 |
| LogisticRegression | 33 | treatment exposure | drug_gsn_066419_c17fadb0e8_count_24h | NORepinephrine - count, first 24 hours | 0.000033 |
| LogisticRegression | 34 | treatment exposure | drug_formulary_norebasens_c31a0d47cc_count_24h | 0.9% Sodium Chloride - count, first 24 hours | 0.000031 |

*Notes:* Rows are reproduced from the saved MIMIC-IV SHAP summary output. When a rank column was absent in the source file, source row order is shown. SHAP values were generated from the best-performing algorithm selected for each feature matrix. Best-performing algorithms were selected by highest mean cross-validated AUPRC, with mean AUROC used as the tie-breaker. SHAP values are model-specific attributions, not causal effects; absolute SHAP magnitudes should not be compared directly across separately trained models or datasets.

eTable 28. MIMIC-IV SHAP feature importance for Baseline + procedure burden.

| Model | Rank or source order | Feature domain | Original feature name | Readable concept or feature name | Mean absolute SHAP |
|---|---|---|---|---|---|
| LogisticRegression | 1 | baseline | visit_type | Visit type | 0.025748 |
| LogisticRegression | 2 | baseline | sex | Sex | 0.022931 |
| LogisticRegression | 3 | baseline | age_at_visit | Age at visit | 0.019281 |
| LogisticRegression | 4 | baseline | charlson_index | Charlson Comorbidity Index | 0.013757 |
| LogisticRegression | 5 | procedure burden | unique_procedure_count_24h | Unique procedure count, first 24 hours | 0.009617 |
| LogisticRegression | 6 | baseline | prior_visit_count | Prior visit count | 0.008759 |
| LogisticRegression | 7 | baseline | race | Race | 0.007384 |
| LogisticRegression | 8 | baseline | ethnicity | Ethnicity | 0.006865 |
| LogisticRegression | 9 | baseline | prior_acute_visit_count | Prior acute-care visit count | 0.006157 |
| LogisticRegression | 10 | procedure burden | procedure_count_total_24h | Total procedure count, first 24 hours | 0.005309 |
| LogisticRegression | 11 | baseline | has_prior_visit | Has prior visit | 0.002571 |
| LogisticRegression | 12 | procedure burden | procedure_icd9_9671_5bce41582d_presence_24h | ICD-9 procedure 9671 - presence indicator, first 24 hours | 0.001431 |
| LogisticRegression | 13 | procedure burden | procedure_icd10_5a1955z_6a2b52611f_count_24h | ICD-10-PCS procedure 5A1955Z - count, first 24 hours | 0.001429 |
| LogisticRegression | 14 | procedure burden | procedure_icd10_5a1955z_6a2b52611f_presence_24h | ICD-10-PCS procedure 5A1955Z - presence indicator, first 24 hours | 0.001363 |
| LogisticRegression | 15 | procedure burden | any_procedure_24h | Any procedure recorded, first 24 hours | 0.001282 |
| LogisticRegression | 16 | procedure burden | procedure_icd10_5a1945z_f9e7fcf8fd_count_24h | ICD-10-PCS procedure 5A1945Z - count, first 24 hours | 0.000894 |
| LogisticRegression | 17 | procedure burden | procedure_icd10_5a1945z_f9e7fcf8fd_presence_24h | ICD-10-PCS procedure 5A1945Z - presence indicator, first 24 hours | 0.000888 |
| LogisticRegression | 18 | procedure burden | procedure_icd10_02hv33z_8e9f454673_presence_24h | ICD-10-PCS procedure 02HV33Z - presence indicator, first 24 hours | 0.000807 |
| LogisticRegression | 19 | procedure burden | procedure_icd10_02hv33z_8e9f454673_count_24h | ICD-10-PCS procedure 02HV33Z - count, first 24 hours | 0.000734 |
| LogisticRegression | 20 | procedure burden | procedure_icd10_3e0g76z_fa1c50513e_count_24h | ICD-10-PCS procedure 3E0G76Z - count, first 24 hours | 0.000507 |
| LogisticRegression | 21 | procedure burden | procedure_icd10_3e0g76z_fa1c50513e_presence_24h | ICD-10-PCS procedure 3E0G76Z - presence indicator, first 24 hours | 0.000506 |
| LogisticRegression | 22 | procedure burden | procedure_icd9_9672_93b639be5d_presence_24h | ICD-9 procedure 9672 - presence indicator, first 24 hours | 0.000471 |
| LogisticRegression | 23 | procedure burden | procedure_icd9_9672_93b639be5d_count_24h | ICD-9 procedure 9672 - count, first 24 hours | 0.000467 |

Continued on next page

eTable 28. MIMIC-IV SHAP feature importance for Baseline + procedure burden. (continued)

| Model | Rank or source order | Feature domain | Original feature name | Readable concept or feature name | Mean absolute SHAP |
|---|---|---|---|---|---|
| LogisticRegression | 24 | procedure burden | `procedure_icd9_3893_49b890a736_presence_24h` | ICD-9 procedure 3893 - presence indicator, first 24 hours | 0.000365 |
| LogisticRegression | 25 | procedure burden | `procedure_icd9_3893_49b890a736_count_24h` | ICD-9 procedure 3893 - count, first 24 hours | 0.000187 |
| LogisticRegression | 26 | procedure burden | `procedure_icd9_9671_5bce41582d_count_24h` | ICD-9 procedure 9671 - count, first 24 hours | 0.000151 |
| LogisticRegression | 27 | procedure burden | `procedure_icd10_0bh17ez_3d5ba254ee_presence_24h` | ICD-10-PCS procedure 0BH17EZ - presence indicator, first 24 hours | 0.000082 |
| LogisticRegression | 28 | procedure burden | `procedure_icd10_0bh17ez_3d5ba254ee_count_24h` | ICD-10-PCS procedure 0BH17EZ - count, first 24 hours | 0.000080 |
| LogisticRegression | 29 | procedure burden | `procedure_icd9_3891_71dd24c736_presence_24h` | ICD-9 procedure 3891 - presence indicator, first 24 hours | 0.000027 |
| LogisticRegression | 30 | procedure burden | `procedure_icd9_3891_71dd24c736_count_24h` | ICD-9 procedure 3891 - count, first 24 hours | 0.000018 |
| LogisticRegression | 31 | procedure burden | `procedure_icd9_9604_93039b475e_count_24h` | ICD-9 procedure 9604 - count, first 24 hours | 0.000008 |
| LogisticRegression | 32 | procedure burden | `procedure_icd9_9604_93039b475e_presence_24h` | ICD-9 procedure 9604 - presence indicator, first 24 hours | 0.000007 |

*Notes:* Rows are reproduced from the saved MIMIC-IV SHAP summary output. When a rank column was absent in the source file, source row order is shown. SHAP values were generated from the best-performing algorithm selected for each feature matrix. Best-performing algorithms were selected by highest mean cross-validated AUPRC, with mean AUROC used as the tie-breaker. SHAP values are model-specific attributions, not causal effects; absolute SHAP magnitudes should not be compared directly across separately trained models or datasets.

eTable 29. MIMIC-IV SHAP feature importance for Baseline + physiological severity + treatment exposure.

| Model | Rank or source order | Feature domain | Original feature name | Readable concept or feature name | Mean absolute SHAP |
|---|---|---|---|---|---|
| RandomForest | 1 | baseline | `age_at_visit` | Age at visit | 0.013410 |
| RandomForest | 2 | baseline | `charlson_index` | Charlson Comorbidity Index | 0.006110 |
| RandomForest | 3 | physiological severity | `lab_51006_urea_nitrogen_max` | Urea Nitrogen - maximum, first 24 hours | 0.004353 |
| RandomForest | 4 | physiological severity | `lab_51277_rdw_max` | RDW - maximum, first 24 hours | 0.004244 |
| RandomForest | 5 | physiological severity | `lab_50983_sodium_min` | Sodium - minimum, first 24 hours | 0.004222 |
| RandomForest | 6 | physiological severity | `lab_51006_urea_nitrogen_mean` | Urea Nitrogen - mean, first 24 hours | 0.004150 |
| RandomForest | 7 | baseline | `visit_type` | Visit type | 0.004133 |
| RandomForest | 8 | physiological severity | `lab_51277_rdw_mean` | RDW - mean, first 24 hours | 0.003717 |
| RandomForest | 9 | physiological severity | `lab_51301_white_blood_cells_max` | White Blood Cells - maximum, first 24 hours | 0.003468 |
| RandomForest | 10 | physiological severity | `lab_50983_sodium_mean` | Sodium - mean, first 24 hours | 0.003459 |
| RandomForest | 11 | physiological severity | `lab_51222_hemoglobin_max` | Hemoglobin - maximum, first 24 hours | 0.002961 |
| RandomForest | 12 | treatment exposure | `unique_drug_count_24h` | Unique drug count, first 24 hours | 0.002875 |
| RandomForest | 13 | physiological severity | `lab_51237_inr_pt_mean` | INR(PT) - mean, first 24 hours | 0.002853 |

Continued on next page

eTable 29. MIMIC-IV SHAP feature importance for Baseline + physiological severity + treatment exposure. (continued)

| Model | Rank or source order | Feature domain | Original feature name | Readable concept or feature name | Mean absolute SHAP |
|---|---|---|---|---|---|
| RandomForest | 14 | physiological severity | lab_51274_pt_mean | PT - mean, first 24 hours | 0.002751 |
| RandomForest | 15 | physiological severity | lab_51277_rdw_min | RDW - minimum, first 24 hours | 0.002707 |
| RandomForest | 16 | physiological severity | lab_51237_inr_pt_max | INR(PT) - maximum, first 24 hours | 0.002551 |
| RandomForest | 17 | physiological severity | lab_51006_urea_nitrogen_min | Urea Nitrogen - minimum, first 24 hours | 0.002523 |
| RandomForest | 18 | physiological severity | lab_50954_lactate_dehydrogenase_ld_min | Lactate Dehydrogenase (LD) - minimum, first 24 hours | 0.002464 |
| RandomForest | 19 | physiological severity | lab_50912_creatinine_mean | Creatinine - mean, first 24 hours | 0.002408 |
| RandomForest | 20 | physiological severity | lab_50882_bicarbonate_mean | Bicarbonate - mean, first 24 hours | 0.002369 |
| RandomForest | 21 | treatment exposure | drug_gsn_001210_ba3dc43c8d_count_24h | 0.9% Sodium Chloride - count, first 24 hours | 0.002324 |
| RandomForest | 22 | treatment exposure | repeat_drug_exposure_count_24h | Repeat drug exposure count, first 24 hours | 0.002209 |
| RandomForest | 23 | physiological severity | lab_51274_pt_max | PT - maximum, first 24 hours | 0.002155 |
| RandomForest | 24 | physiological severity | lab_50893_calcium_total_min | Calcium, Total - minimum, first 24 hours | 0.002150 |
| RandomForest | 25 | physiological severity | lab_50813_lactate_min | Lactate - minimum, first 24 hours | 0.002118 |
| RandomForest | 26 | treatment exposure | time_to_first_drug_hours | Time to first drug exposure, hours | 0.002056 |
| RandomForest | 27 | physiological severity | lab_50983_sodium_max | Sodium - maximum, first 24 hours | 0.001985 |
| RandomForest | 28 | baseline | race | Race | 0.001918 |
| RandomForest | 29 | physiological severity | lab_50813_lactate_max | Lactate - maximum, first 24 hours | 0.001757 |
| RandomForest | 30 | physiological severity | lab_50813_lactate_mean | Lactate - mean, first 24 hours | 0.001753 |
| RandomForest | 31 | physiological severity | lab_50893_calcium_total_mean | Calcium, Total - mean, first 24 hours | 0.001695 |
| RandomForest | 32 | treatment exposure | drug_gsn_060304_4b8ed8c2a4_count_24h | Heparin Flush (10 units/ml) - count, first 24 hours | 0.001634 |
| RandomForest | 33 | physiological severity | lab_50983_sodium_count | Sodium - count, first 24 hours | 0.001558 |
| RandomForest | 34 | baseline | ethnicity | Ethnicity | 0.001501 |
| RandomForest | 35 | physiological severity | lab_50931_glucose_count | Glucose - count, first 24 hours | 0.001482 |
| RandomForest | 36 | physiological severity | lab_50821_po2_min | pO2 - minimum, first 24 hours | 0.001477 |
| RandomForest | 37 | physiological severity | lab_50893_calcium_total_max | Calcium, Total - maximum, first 24 hours | 0.001476 |
| RandomForest | 38 | baseline | sex | Sex | 0.001451 |
| RandomForest | 39 | physiological severity | lab_50882_bicarbonate_std | Bicarbonate - standard deviation, first 24 hours | 0.001334 |
| RandomForest | 40 | baseline | prior_visit_count | Prior visit count | 0.001222 |

Continued on next page

eTable 29. MIMIC-IV SHAP feature importance for Baseline + physiological severity + treatment exposure. (continued)

| Model | Rank or source order | Feature domain | Original feature name | Readable concept or feature name | Mean absolute SHAP |
|---|---|---|---|---|---|
| RandomForest | 41 | physiological severity | lab_50821_po2_mean | pO2 - mean, first 24 hours | 0.001199 |
| RandomForest | 42 | treatment exposure | drug_gsn_001210_ba3dc43c8d_exposed_24h | 0.9% Sodium Chloride - exposure indicator, first 24 hours | 0.001089 |
| RandomForest | 43 | physiological severity | lab_50820_ph_mean | pH - mean, first 24 hours | 0.001037 |
| RandomForest | 44 | physiological severity | lab_50983_sodium_std | Sodium - standard deviation, first 24 hours | 0.001007 |
| RandomForest | 45 | physiological severity | lab_50804_calculated_total_co2_std | Calculated Total CO2 - standard deviation, first 24 hours | 0.000980 |
| RandomForest | 46 | baseline | prior_acute_visit_count | Prior acute-care visit count | 0.000970 |
| RandomForest | 47 | physiological severity | lab_50820_ph_min | pH - minimum, first 24 hours | 0.000962 |
| RandomForest | 48 | physiological severity | lab_50818_pco2_std | pCO2 - standard deviation, first 24 hours | 0.000937 |
| RandomForest | 49 | treatment exposure | drug_gsn_001972_b5f0a0eff8_count_24h | 5% Dextrose - count, first 24 hours | 0.000935 |
| RandomForest | 50 | physiological severity | lab_50971_potassium_count | Potassium - count, first 24 hours | 0.000922 |
| RandomForest | 51 | physiological severity | lab_50821_po2_max | pO2 - maximum, first 24 hours | 0.000918 |
| RandomForest | 52 | physiological severity | lab_50820_ph_max | pH - maximum, first 24 hours | 0.000905 |
| RandomForest | 53 | physiological severity | lab_50893_calcium_total_count | Calcium, Total - count, first 24 hours | 0.000886 |
| RandomForest | 54 | physiological severity | lab_50818_pco2_max | pCO2 - maximum, first 24 hours | 0.000795 |
| RandomForest | 55 | physiological severity | lab_50818_pco2_mean | pCO2 - mean, first 24 hours | 0.000788 |
| RandomForest | 56 | treatment exposure | drug_formulary_norebasens_c31a0d47cc_count_24h | 0.9% Sodium Chloride - count, first 24 hours | 0.000787 |
| RandomForest | 57 | physiological severity | lab_50970_phosphate_count | Phosphate - count, first 24 hours | 0.000776 |
| RandomForest | 58 | physiological severity | lab_50902_chloride_count | Chloride - count, first 24 hours | 0.000766 |
| RandomForest | 59 | treatment exposure | drug_formulary_vial_b4c8f642b1_exposed_24h | Vial - exposure indicator, first 24 hours | 0.000723 |
| RandomForest | 60 | physiological severity | lab_50818_pco2_min | pCO2 - minimum, first 24 hours | 0.000655 |
| RandomForest | 61 | treatment exposure | drug_formulary_vancobase_228466e611_count_24h | Iso-Osmotic Dextrose - count, first 24 hours | 0.000608 |
| RandomForest | 62 | physiological severity | lab_50804_calculated_total_co2_mean | Calculated Total CO2 - mean, first 24 hours | 0.000601 |
| RandomForest | 63 | treatment exposure | drug_gsn_066419_c17fadb0e8_count_24h | NORepinephrine - count, first 24 hours | 0.000599 |
| RandomForest | 64 | treatment exposure | drug_gsn_043952_0602de6ed2_count_24h | Vancomycin - count, first 24 hours | 0.000570 |

Continued on next page

eTable 29. MIMIC-IV SHAP feature importance for Baseline + physiological severity + treatment exposure. (continued)

| Model | Rank or source order | Feature domain | Original feature name | Readable concept or feature name | Mean absolute SHAP |
|---|---|---|---|---|---|
| RandomForest | 65 | treatment exposure | `drug_formulary_vial_b4c8f642b1_count_24h` | Vial - count, first 24 hours | 0.000562 |
| RandomForest | 66 | treatment exposure | `drug_formulary_vancobase_228466e611_exposed_24h` | Iso-Osmotic Dextrose - exposure indicator, first 24 hours | 0.000473 |
| RandomForest | 67 | treatment exposure | `drug_gsn_043952_0602de6ed2_exposed_24h` | Vancomycin - exposure indicator, first 24 hours | 0.000457 |
| RandomForest | 68 | treatment exposure | `any_drug_24h` | Any drug exposure, first 24 hours | 0.000427 |
| RandomForest | 69 | baseline | `has_prior_visit` | Has prior visit | 0.000423 |
| RandomForest | 70 | physiological severity | `lab_50820_ph_std` | pH - standard deviation, first 24 hours | 0.000377 |
| RandomForest | 71 | treatment exposure | `drug_gsn_001972_b5f0a0eff8_exposed_24h` | 5% Dextrose - exposure indicator, first 24 hours | 0.000375 |
| RandomForest | 72 | treatment exposure | `drug_gsn_016796_f69d8264fe_count_24h` | Propofol - count, first 24 hours | 0.000319 |
| RandomForest | 73 | treatment exposure | `drug_gsn_016796_f69d8264fe_exposed_24h` | Propofol - exposure indicator, first 24 hours | 0.000295 |
| RandomForest | 74 | physiological severity | `lab_50821_po2_std` | pO2 - standard deviation, first 24 hours | 0.000278 |
| RandomForest | 75 | treatment exposure | `drug_gsn_057959_a518f33e58_exposed_24h` | Chlorhexidine Gluconate 0.12% Oral Rinse - exposure indicator, first 24 hours | 0.000157 |
| RandomForest | 76 | treatment exposure | `drug_gsn_048287_21c090e9e3_count_24h` | Fentanyl Citrate - count, first 24 hours | 0.000138 |
| RandomForest | 77 | treatment exposure | `drug_gsn_057959_a518f33e58_count_24h` | Chlorhexidine Gluconate 0.12% Oral Rinse - count, first 24 hours | 0.000121 |
| RandomForest | 78 | treatment exposure | `drug_formulary_fentsoln50_7775dcade8_count_24h` | Soln - count, first 24 hours | 0.000117 |
| RandomForest | 79 | treatment exposure | `drug_gsn_048287_21c090e9e3_exposed_24h` | Fentanyl Citrate - exposure indicator, first 24 hours | 0.000111 |
| RandomForest | 80 | treatment exposure | `drug_formulary_fentsoln50_7775dcade8_exposed_24h` | Soln - exposure indicator, first 24 hours | 0.000092 |

*Notes:* Rows are reproduced from the saved MIMIC-IV SHAP summary output. When a rank column was absent in the source file, source row order is shown. SHAP values were generated from the best-performing algorithm selected for each feature matrix. Best-performing algorithms were selected by highest mean cross-validated AUPRC, with mean AUROC used as the tie-breaker. SHAP values are model-specific attributions, not causal effects; absolute SHAP magnitudes should not be compared directly across separately trained models or datasets.

eTable 30. MIMIC-IV SHAP feature importance for Baseline + physiological severity + procedure burden.

| Model | Rank or source order | Feature domain | Original feature name | Readable concept or feature name | Mean absolute SHAP |
|---|---|---|---|---|---|
| RandomForest | 1 | baseline | `age_at_visit` | Age at visit | 0.014204 |
| RandomForest | 2 | baseline | `charlson_index` | Charlson Comorbidity Index | 0.006329 |
| RandomForest | 3 | baseline | `visit_type` | Visit type | 0.005573 |
| RandomForest | 4 | physiological severity | `lab_50983_sodium_min` | Sodium - minimum, first 24 hours | 0.005020 |
| RandomForest | 5 | physiological severity | `lab_51277_rdw_max` | RDW - maximum, first 24 hours | 0.004590 |
| RandomForest | 6 | physiological severity | `lab_51222_hemoglobin_max` | Hemoglobin - maximum, first 24 hours | 0.004547 |
| RandomForest | 7 | physiological severity | `lab_51006_urea_nitrogen_max` | Urea Nitrogen - maximum, first 24 hours | 0.004324 |

Continued on next page

eTable 30. MIMIC-IV SHAP feature importance for Baseline + physiological severity + procedure burden. (continued)

| Model | Rank or source order | Feature domain | Original feature name | Readable concept or feature name | Mean absolute SHAP |
|---|---|---|---|---|---|
| RandomForest | 8 | physiological severity | lab_51277_rdw_mean | RDW - mean, first 24 hours | 0.004264 |
| RandomForest | 9 | physiological severity | lab_51301_white_blood_cells_max | White Blood Cells - maximum, first 24 hours | 0.004249 |
| RandomForest | 10 | physiological severity | lab_50983_sodium_mean | Sodium - mean, first 24 hours | 0.003959 |
| RandomForest | 11 | physiological severity | lab_51006_urea_nitrogen_mean | Urea Nitrogen - mean, first 24 hours | 0.003917 |
| RandomForest | 12 | physiological severity | lab_50893_calcium_total_min | Calcium, Total - minimum, first 24 hours | 0.003165 |
| RandomForest | 13 | physiological severity | lab_51006_urea_nitrogen_min | Urea Nitrogen - minimum, first 24 hours | 0.003160 |
| RandomForest | 14 | physiological severity | lab_51277_rdw_min | RDW - minimum, first 24 hours | 0.003113 |
| RandomForest | 15 | physiological severity | lab_50912_creatinine_mean | Creatinine - mean, first 24 hours | 0.003109 |
| RandomForest | 16 | physiological severity | lab_51274_pt_mean | PT - mean, first 24 hours | 0.002901 |
| RandomForest | 17 | physiological severity | lab_51237_inr_pt_mean | INR(PT) - mean, first 24 hours | 0.002803 |
| RandomForest | 18 | physiological severity | lab_51237_inr_pt_max | INR(PT) - maximum, first 24 hours | 0.002546 |
| RandomForest | 19 | physiological severity | lab_50882_bicarbonate_mean | Bicarbonate - mean, first 24 hours | 0.002484 |
| RandomForest | 20 | physiological severity | lab_50893_calcium_total_mean | Calcium, Total - mean, first 24 hours | 0.002410 |
| RandomForest | 21 | baseline | race | Race | 0.002338 |
| RandomForest | 22 | physiological severity | lab_50954_lactate_dehydrogenase_ld_min | Lactate Dehydrogenase (LD) - minimum, first 24 hours | 0.002331 |
| RandomForest | 23 | physiological severity | lab_50813_lactate_min | Lactate - minimum, first 24 hours | 0.002319 |
| RandomForest | 24 | physiological severity | lab_50983_sodium_max | Sodium - maximum, first 24 hours | 0.002177 |
| RandomForest | 25 | physiological severity | lab_50893_calcium_total_max | Calcium, Total - maximum, first 24 hours | 0.002153 |
| RandomForest | 26 | physiological severity | lab_50813_lactate_max | Lactate - maximum, first 24 hours | 0.002070 |
| RandomForest | 27 | baseline | sex | Sex | 0.002063 |
| RandomForest | 28 | physiological severity | lab_50813_lactate_mean | Lactate - mean, first 24 hours | 0.002012 |
| RandomForest | 29 | physiological severity | lab_51274_pt_max | PT - maximum, first 24 hours | 0.001922 |
| RandomForest | 30 | physiological severity | lab_50983_sodium_count | Sodium - count, first 24 hours | 0.001826 |
| RandomForest | 31 | baseline | prior_visit_count | Prior visit count | 0.001509 |
| RandomForest | 32 | physiological severity | lab_50931_glucose_count | Glucose - count, first 24 hours | 0.001502 |
| RandomForest | 33 | baseline | ethnicity | Ethnicity | 0.001498 |

Continued on next page

eTable 30. MIMIC-IV SHAP feature importance for Baseline + physiological severity + procedure burden. (continued)

| Model | Rank or source order | Feature domain | Original feature name | Readable concept or feature name | Mean absolute SHAP |
|---|---|---|---|---|---|
| RandomForest | 34 | physiological severity | lab_50818_pco2_mean | pCO2 - mean, first 24 hours | 0.001447 |
| RandomForest | 35 | physiological severity | lab_50882_bicarbonate_std | Bicarbonate - standard deviation, first 24 hours | 0.001365 |
| RandomForest | 36 | baseline | prior_acute_visit_count | Prior acute-care visit count | 0.001322 |
| RandomForest | 37 | physiological severity | lab_50893_calcium_total_count | Calcium, Total - count, first 24 hours | 0.001283 |
| RandomForest | 38 | physiological severity | lab_50821_po2_mean | pO2 - mean, first 24 hours | 0.001260 |
| RandomForest | 39 | physiological severity | lab_50820_ph_mean | pH - mean, first 24 hours | 0.001212 |
| RandomForest | 40 | physiological severity | lab_50821_po2_min | pO2 - minimum, first 24 hours | 0.001131 |
| RandomForest | 41 | physiological severity | lab_50804_calculated_total_co2_std | Calculated Total CO2 - standard deviation, first 24 hours | 0.001125 |
| RandomForest | 42 | physiological severity | lab_50983_sodium_std | Sodium - standard deviation, first 24 hours | 0.001090 |
| RandomForest | 43 | physiological severity | lab_50971_potassium_count | Potassium - count, first 24 hours | 0.001053 |
| RandomForest | 44 | physiological severity | lab_50820_ph_max | pH - maximum, first 24 hours | 0.001040 |
| RandomForest | 45 | physiological severity | lab_50818_pco2_max | pCO2 - maximum, first 24 hours | 0.001029 |
| RandomForest | 46 | physiological severity | lab_50820_ph_min | pH - minimum, first 24 hours | 0.001028 |
| RandomForest | 47 | physiological severity | lab_50821_po2_max | pO2 - maximum, first 24 hours | 0.001003 |
| RandomForest | 48 | physiological severity | lab_50818_pco2_std | pCO2 - standard deviation, first 24 hours | 0.000961 |
| RandomForest | 49 | physiological severity | lab_50970_phosphate_count | Phosphate - count, first 24 hours | 0.000898 |
| RandomForest | 50 | physiological severity | lab_50818_pco2_min | pCO2 - minimum, first 24 hours | 0.000847 |
| RandomForest | 51 | physiological severity | lab_50804_calculated_total_co2_mean | Calculated Total CO2 - mean, first 24 hours | 0.000764 |
| RandomForest | 52 | physiological severity | lab_50902_chloride_count | Chloride - count, first 24 hours | 0.000763 |
| RandomForest | 53 | procedure burden | unique_procedure_count_24h | Unique procedure count, first 24 hours | 0.000752 |
| RandomForest | 54 | procedure burden | procedure_count_total_24h | Total procedure count, first 24 hours | 0.000741 |
| RandomForest | 55 | physiological severity | lab_50820_ph_std | pH - standard deviation, first 24 hours | 0.000667 |
| RandomForest | 56 | procedure burden | any_procedure_24h | Any procedure recorded, first 24 hours | 0.000563 |
| RandomForest | 57 | baseline | has_prior_visit | Has prior visit | 0.000535 |
| RandomForest | 58 | procedure burden | procedure_icd9_9672_93b639be5d_presence_24h | ICD-9 procedure 9672 - presence indicator, first 24 hours | 0.000439 |
| RandomForest | 59 | procedure burden | procedure_icd10_5a1955z_6a2b52611f_presence_24h | ICD-10-PCS procedure 5A1955Z - presence indicator, first 24 hours | 0.000438 |

Continued on next page

eTable 30. MIMIC-IV SHAP feature importance for Baseline + physiological severity + procedure burden. (continued)

| Model | Rank or source order | Feature domain | Original feature name | Readable concept or feature name | Mean absolute SHAP |
|---|---|---|---|---|---|
| RandomForest | 60 | procedure burden | `procedure_icd9_9672_93b639be5d_count_24h` | ICD-9 procedure 9672 - count, first 24 hours | 0.000401 |
| RandomForest | 61 | procedure burden | `procedure_icd10_5a1955z_6a2b52611f_count_24h` | ICD-10-PCS procedure 5A1955Z - count, first 24 hours | 0.000362 |
| RandomForest | 62 | physiological severity | `lab_50821_po2_std` | pO2 - standard deviation, first 24 hours | 0.000360 |
| RandomForest | 63 | procedure burden | `procedure_icd10_0bh17ez_3d5ba254ee_count_24h` | ICD-10-PCS procedure 0BH17EZ - count, first 24 hours | 0.000246 |
| RandomForest | 64 | procedure burden | `procedure_icd9_3893_49b890a736_presence_24h` | ICD-9 procedure 3893 - presence indicator, first 24 hours | 0.000236 |
| RandomForest | 65 | procedure burden | `procedure_icd9_3893_49b890a736_count_24h` | ICD-9 procedure 3893 - count, first 24 hours | 0.000233 |
| RandomForest | 66 | procedure burden | `procedure_icd10_0bh17ez_3d5ba254ee_presence_24h` | ICD-10-PCS procedure 0BH17EZ - presence indicator, first 24 hours | 0.000231 |
| RandomForest | 67 | procedure burden | `procedure_icd10_5a1945z_f9e7fcf8fd_count_24h` | ICD-10-PCS procedure 5A1945Z - count, first 24 hours | 0.000224 |
| RandomForest | 68 | procedure burden | `procedure_icd9_9604_93039b475e_count_24h` | ICD-9 procedure 9604 - count, first 24 hours | 0.000209 |
| RandomForest | 69 | procedure burden | `procedure_icd10_5a1945z_f9e7fcf8fd_presence_24h` | ICD-10-PCS procedure 5A1945Z - presence indicator, first 24 hours | 0.000203 |
| RandomForest | 70 | procedure burden | `procedure_icd9_9604_93039b475e_presence_24h` | ICD-9 procedure 9604 - presence indicator, first 24 hours | 0.000201 |
| RandomForest | 71 | procedure burden | `procedure_icd9_9671_5bce41582d_count_24h` | ICD-9 procedure 9671 - count, first 24 hours | 0.000165 |
| RandomForest | 72 | procedure burden | `procedure_icd9_9671_5bce41582d_presence_24h` | ICD-9 procedure 9671 - presence indicator, first 24 hours | 0.000148 |
| RandomForest | 73 | procedure burden | `procedure_icd10_3e0g76z_fa1c50513e_presence_24h` | ICD-10-PCS procedure 3E0G76Z - presence indicator, first 24 hours | 0.000068 |
| RandomForest | 74 | procedure burden | `procedure_icd10_3e0g76z_fa1c50513e_count_24h` | ICD-10-PCS procedure 3E0G76Z - count, first 24 hours | 0.000063 |
| RandomForest | 75 | procedure burden | `procedure_icd10_02hv33z_8e9f454673_count_24h` | ICD-10-PCS procedure 02HV33Z - count, first 24 hours | 0.000056 |
| RandomForest | 76 | procedure burden | `procedure_icd10_02hv33z_8e9f454673_presence_24h` | ICD-10-PCS procedure 02HV33Z - presence indicator, first 24 hours | 0.000043 |
| RandomForest | 77 | procedure burden | `procedure_icd9_3891_71dd24c736_count_24h` | ICD-9 procedure 3891 - count, first 24 hours | 0.000000 |
| RandomForest | 78 | procedure burden | `procedure_icd9_3891_71dd24c736_presence_24h` | ICD-9 procedure 3891 - presence indicator, first 24 hours | 0.000000 |

*Notes:* Rows are reproduced from the saved MIMIC-IV SHAP summary output. When a rank column was absent in the source file, source row order is shown. SHAP values were generated from the best-performing algorithm selected for each feature matrix. Best-performing algorithms were selected by highest mean cross-validated AUPRC, with mean AUROC used as the tie-breaker. SHAP values are model-specific attributions, not causal effects; absolute SHAP magnitudes should not be compared directly across separately trained models or datasets.

eTable 31. MIMIC-IV SHAP feature importance for Baseline + treatment exposure + procedure burden.

| Model | Rank or source order | Feature domain | Original feature name | Readable concept or feature name | Mean absolute SHAP |
|---|---|---|---|---|---|
| LogisticRegression | 1 | baseline | `visit_type` | Visit type | 0.023100 |
| LogisticRegression | 2 | baseline | sex | Sex | 0.022444 |
| LogisticRegression | 3 | baseline | `age_at_visit` | Age at visit | 0.022182 |

Continued on next page

eTable 31. MIMIC-IV SHAP feature importance for Baseline + treatment exposure + procedure burden. (continued)

| Model | Rank or source order | Feature domain | Original feature name | Readable concept or feature name | Mean absolute SHAP |
|---|---|---|---|---|---|
| LogisticRegression | 4 | baseline | charlson_index | Charlson Comorbidity Index | 0.012359 |
| LogisticRegression | 5 | baseline | prior_visit_count | Prior visit count | 0.011010 |
| LogisticRegression | 6 | procedure burden | unique_procedure_count_24h | Unique procedure count, first 24 hours | 0.010681 |
| LogisticRegression | 7 | baseline | prior_acute_visit_count | Prior acute-care visit count | 0.008699 |
| LogisticRegression | 8 | treatment exposure | drug_formulary_vial_b4c8f642b1_exposed_24h | Vial - exposure indicator, first 24 hours | 0.006636 |
| LogisticRegression | 9 | baseline | race | Race | 0.006382 |
| LogisticRegression | 10 | baseline | ethnicity | Ethnicity | 0.006332 |
| LogisticRegression | 11 | procedure burden | procedure_count_total_24h | Total procedure count, first 24 hours | 0.005929 |
| LogisticRegression | 12 | treatment exposure | drug_formulary_fentsoln50_7775dcade8_exposed_24h | Soln - exposure indicator, first 24 hours | 0.005374 |
| LogisticRegression | 13 | treatment exposure | drug_gsn_001210_ba3dc43c8d_exposed_24h | 0.9% Sodium Chloride - exposure indicator, first 24 hours | 0.005146 |
| LogisticRegression | 14 | treatment exposure | repeat_drug_exposure_count_24h | Repeat drug exposure count, first 24 hours | 0.004462 |
| LogisticRegression | 15 | treatment exposure | drug_gsn_060304_4b8ed8c2a4_count_24h | Heparin Flush (10 units/ml) - count, first 24 hours | 0.003690 |
| LogisticRegression | 16 | treatment exposure | drug_gsn_001972_b5f0a0eff8_count_24h | 5% Dextrose - count, first 24 hours | 0.003594 |
| LogisticRegression | 17 | treatment exposure | drug_gsn_057959_a518f33e58_exposed_24h | Chlorhexidine Gluconate 0.12% Oral Rinse - exposure indicator, first 24 hours | 0.003557 |
| LogisticRegression | 18 | procedure burden | procedure_icd10_02hv33z_8e9f454673_count_24h | ICD-10-PCS procedure 02HV33Z - count, first 24 hours | 0.003016 |
| LogisticRegression | 19 | treatment exposure | drug_gsn_001210_ba3dc43c8d_count_24h | 0.9% Sodium Chloride - count, first 24 hours | 0.002971 |
| LogisticRegression | 20 | procedure burden | procedure_icd10_02hv33z_8e9f454673_presence_24h | ICD-10-PCS procedure 02HV33Z - presence indicator, first 24 hours | 0.002780 |
| LogisticRegression | 21 | treatment exposure | drug_gsn_043952_0602de6ed2_exposed_24h | Vancomycin - exposure indicator, first 24 hours | 0.002761 |
| LogisticRegression | 22 | treatment exposure | drug_formulary_vancobase_228466e611_exposed_24h | Iso-Osmotic Dextrose - exposure indicator, first 24 hours | 0.002760 |
| LogisticRegression | 23 | treatment exposure | drug_formulary_fentsoln50_7775dcade8_count_24h | Soln - count, first 24 hours | 0.002636 |
| LogisticRegression | 24 | baseline | has_prior_visit | Has prior visit | 0.002608 |
| LogisticRegression | 25 | procedure burden | procedure_icd9_3893_49b890a736_presence_24h | ICD-9 procedure 3893 - presence indicator, first 24 hours | 0.002451 |
| LogisticRegression | 26 | treatment exposure | unique_drug_count_24h | Unique drug count, first 24 hours | 0.002301 |
| LogisticRegression | 27 | procedure burden | procedure_icd9_3893_49b890a736_count_24h | ICD-9 procedure 3893 - count, first 24 hours | 0.002022 |
| LogisticRegression | 28 | treatment exposure | drug_formulary_vial_b4c8f642b1_count_24h | Vial - count, first 24 hours | 0.001777 |
| LogisticRegression | 29 | treatment exposure | drug_gsn_057959_a518f33e58_count_24h | Chlorhexidine Gluconate 0.12% Oral Rinse - count, first 24 hours | 0.001767 |
| LogisticRegression | 30 | treatment exposure | drug_gsn_048287_21c090e9e3_exposed_24h | Fentanyl Citrate - exposure indicator, first 24 hours | 0.001621 |
| LogisticRegression | 31 | treatment exposure | any_drug_24h | Any drug exposure, first 24 hours | 0.001545 |
| LogisticRegression | 32 | treatment exposure | drug_formulary_vancobase_228466e611_count_24h | Iso-Osmotic Dextrose - count, first 24 hours | 0.001505 |

Continued on next page

eTable 31. MIMIC-IV SHAP feature importance for Baseline + treatment exposure + procedure burden. (continued)

| Model | Rank or source order | Feature domain | Original feature name | Readable concept or feature name | Mean absolute SHAP |
|---|---|---|---|---|---|
| LogisticRegression | 33 | treatment exposure | drug_gsn_043952_0602de6ed2_count_24h | Vancomycin - count, first 24 hours | 0.001502 |
| LogisticRegression | 34 | procedure burden | procedure_icd9_9672_93b639be5d_presence_24h | ICD-9 procedure 9672 - presence indicator, first 24 hours | 0.001408 |
| LogisticRegression | 35 | procedure burden | procedure_icd9_9672_93b639be5d_count_24h | ICD-9 procedure 9672 - count, first 24 hours | 0.001282 |
| LogisticRegression | 36 | treatment exposure | time_to_first_drug_hours | Time to first drug exposure, hours | 0.001115 |
| LogisticRegression | 37 | procedure burden | any_procedure_24h | Any procedure recorded, first 24 hours | 0.001015 |
| LogisticRegression | 38 | procedure burden | procedure_icd10_5a1945z_f9e7fcf8fd_count_24h | ICD-10-PCS procedure 5A1945Z - count, first 24 hours | 0.000953 |
| LogisticRegression | 39 | procedure burden | procedure_icd9_9671_5bce41582d_presence_24h | ICD-9 procedure 9671 - presence indicator, first 24 hours | 0.000943 |
| LogisticRegression | 40 | procedure burden | procedure_icd10_5a1945z_f9e7fcf8fd_presence_24h | ICD-10-PCS procedure 5A1945Z - presence indicator, first 24 hours | 0.000913 |
| LogisticRegression | 41 | treatment exposure | drug_gsn_016796_f69d8264fe_exposed_24h | Propofol - exposure indicator, first 24 hours | 0.000908 |
| LogisticRegression | 42 | treatment exposure | drug_gsn_048287_21c090e9e3_count_24h | Fentanyl Citrate - count, first 24 hours | 0.000748 |
| LogisticRegression | 43 | treatment exposure | drug_gsn_016796_f69d8264fe_count_24h | Propofol - count, first 24 hours | 0.000607 |
| LogisticRegression | 44 | procedure burden | procedure_icd9_9671_5bce41582d_count_24h | ICD-9 procedure 9671 - count, first 24 hours | 0.000546 |
| LogisticRegression | 45 | treatment exposure | drug_gsn_001972_b5f0a0eff8_exposed_24h | 5% Dextrose - exposure indicator, first 24 hours | 0.000508 |
| LogisticRegression | 46 | procedure burden | procedure_icd10_5a1955z_6a2b52611f_presence_24h | ICD-10-PCS procedure 5A1955Z - presence indicator, first 24 hours | 0.000488 |
| LogisticRegression | 47 | procedure burden | procedure_icd10_5a1955z_6a2b52611f_count_24h | ICD-10-PCS procedure 5A1955Z - count, first 24 hours | 0.000485 |
| LogisticRegression | 48 | procedure burden | procedure_icd9_9604_93039b475e_presence_24h | ICD-9 procedure 9604 - presence indicator, first 24 hours | 0.000336 |
| LogisticRegression | 49 | procedure burden | procedure_icd9_9604_93039b475e_count_24h | ICD-9 procedure 9604 - count, first 24 hours | 0.000313 |
| LogisticRegression | 50 | treatment exposure | drug_gsn_066419_c17fadb0e8_count_24h | NORepinephrine - count, first 24 hours | 0.000306 |
| LogisticRegression | 51 | treatment exposure | drug_formulary_norebasens_c31a0d47cc_count_24h | 0.9% Sodium Chloride - count, first 24 hours | 0.000285 |
| LogisticRegression | 52 | procedure burden | procedure_icd10_3e0g76z_fa1c50513e_count_24h | ICD-10-PCS procedure 3E0G76Z - count, first 24 hours | 0.000097 |
| LogisticRegression | 53 | procedure burden | procedure_icd10_3e0g76z_fa1c50513e_presence_24h | ICD-10-PCS procedure 3E0G76Z - presence indicator, first 24 hours | 0.000090 |
| LogisticRegression | 54 | procedure burden | procedure_icd10_0bh17ez_3d5ba254ee_count_24h | ICD-10-PCS procedure 0BH17EZ - count, first 24 hours | 0.000068 |
| LogisticRegression | 55 | procedure burden | procedure_icd10_0bh17ez_3d5ba254ee_presence_24h | ICD-10-PCS procedure 0BH17EZ - presence indicator, first 24 hours | 0.000066 |
| LogisticRegression | 56 | procedure burden | procedure_icd9_3891_71dd24c736_count_24h | ICD-9 procedure 3891 - count, first 24 hours | 0.000000 |

| | | | | | |
|---|---|---|---|---|---|
| LogisticRegression | 57 | procedure burden | procedure_icd9_3891_71dd24c736_presence_24h | ICD-9 procedure 3891 - presence indicator, first 24 hours | 0.000000 |

*Notes:* Rows are reproduced from the saved MIMIC-IV SHAP summary output. When a rank column was absent in the source file, source row order is shown. SHAP values were generated from the best-performing algorithm selected for each feature matrix. Best-performing algorithms were selected by highest mean cross-validated AUPRC, with mean AUROC used as the tie-breaker. SHAP values are model-specific attributions, not causal effects; absolute SHAP magnitudes should not be compared directly across separately trained models or datasets.

eTable 32. MIMIC-IV SHAP feature importance for Baseline + all clinical domains.

| Model | Rank or source order | Feature domain | Original feature name | Readable concept or feature name | Mean absolute SHAP |
|---|---|---|---|---|---|
| LogisticRegression | 1 | physiological severity | lab_50983_sodium_max | Sodium - maximum, first 24 hours | 0.035457 |
| LogisticRegression | 2 | physiological severity | lab_51006_urea_nitrogen_max | Urea Nitrogen - maximum, first 24 hours | 0.024442 |
| LogisticRegression | 3 | baseline | sex | Sex | 0.023446 |
| LogisticRegression | 4 | baseline | age_at_visit | Age at visit | 0.022351 |
| LogisticRegression | 5 | physiological severity | lab_51006_urea_nitrogen_mean | Urea Nitrogen - mean, first 24 hours | 0.019890 |
| LogisticRegression | 6 | physiological severity | lab_50818_pco2_mean | pCO2 - mean, first 24 hours | 0.019217 |
| LogisticRegression | 7 | baseline | visit_type | Visit type | 0.018549 |
| LogisticRegression | 8 | physiological severity | lab_50983_sodium_mean | Sodium - mean, first 24 hours | 0.016997 |
| LogisticRegression | 9 | baseline | prior_visit_count | Prior visit count | 0.011907 |
| LogisticRegression | 10 | baseline | charlson_index | Charlson Comorbidity Index | 0.011381 |
| LogisticRegression | 11 | physiological severity | lab_51006_urea_nitrogen_min | Urea Nitrogen - minimum, first 24 hours | 0.011148 |
| LogisticRegression | 12 | physiological severity | lab_51277_rdw_min | RDW - minimum, first 24 hours | 0.010985 |
| LogisticRegression | 13 | physiological severity | lab_50971_potassium_count | Potassium - count, first 24 hours | 0.009514 |
| LogisticRegression | 14 | baseline | ethnicity | Ethnicity | 0.009170 |
| LogisticRegression | 15 | baseline | prior_acute_visit_count | Prior acute-care visit count | 0.008726 |
| LogisticRegression | 16 | physiological severity | lab_50931_glucose_count | Glucose - count, first 24 hours | 0.007943 |
| LogisticRegression | 17 | baseline | race | Race | 0.007526 |
| LogisticRegression | 18 | procedure burden | unique_procedure_count_24h | Unique procedure count, first 24 hours | 0.006845 |
| LogisticRegression | 19 | physiological severity | lab_51277_rdw_max | RDW - maximum, first 24 hours | 0.006431 |
| LogisticRegression | 20 | physiological severity | lab_50818_pco2_min | pCO2 - minimum, first 24 hours | 0.006352 |
| LogisticRegression | 21 | treatment exposure | drug_formulary_fentsoln50_7775dcade8_exposed_24h | Soln - exposure indicator, first 24 hours | 0.006300 |
| LogisticRegression | 22 | physiological severity | lab_51277_rdw_mean | RDW - mean, first 24 hours | 0.005765 |
| LogisticRegression | 23 | physiological severity | lab_50820_ph_mean | pH - mean, first 24 hours | 0.005684 |
| LogisticRegression | 24 | physiological severity | lab_50983_sodium_count | Sodium - count, first 24 hours | 0.005486 |
| LogisticRegression | 25 | procedure burden | procedure_icd10_02hv33z_8e9f454673_presence_24h | ICD-10-PCS procedure 02HV33Z - presence indicator, first 24 hours | 0.005260 |

Continued on next page

eTable 32. MIMIC-IV SHAP feature importance for Baseline + all clinical domains. (continued)

| Model | Rank or source order | Feature domain | Original feature name | Readable concept or feature name | Mean absolute SHAP |
|---|---|---|---|---|---|
| LogisticRegression | 26 | treatment exposure | `drug_gsn_001210_ba3dc43c8d_exposed_24h` | 0.9% Sodium Chloride - exposure indicator, first 24 hours | 0.005079 |
| LogisticRegression | 27 | treatment exposure | `drug_formulary_vial_b4c8f642b1_exposed_24h` | Vial - exposure indicator, first 24 hours | 0.004816 |
| LogisticRegression | 28 | physiological severity | `lab_50818_pco2_max` | pCO2 - maximum, first 24 hours | 0.004708 |
| LogisticRegression | 29 | physiological severity | `lab_50912_creatinine_mean` | Creatinine - mean, first 24 hours | 0.004464 |
| LogisticRegression | 30 | treatment exposure | `drug_formulary_fentsoln50_7775dcade8_count_24h` | Soln - count, first 24 hours | 0.004438 |
| LogisticRegression | 31 | treatment exposure | `drug_gsn_060304_4b8ed8c2a4_count_24h` | Heparin Flush (10 units/ml) - count, first 24 hours | 0.004388 |
| LogisticRegression | 32 | procedure burden | `procedure_icd10_02hv33z_8e9f454673_count_24h` | ICD-10-PCS procedure 02HV33Z - count, first 24 hours | 0.003973 |
| LogisticRegression | 33 | physiological severity | `lab_50820_ph_max` | pH - maximum, first 24 hours | 0.003634 |
| LogisticRegression | 34 | physiological severity | `lab_50893_calcium_total_min` | Calcium, Total - minimum, first 24 hours | 0.003587 |
| LogisticRegression | 35 | procedure burden | `procedure_count_total_24h` | Total procedure count, first 24 hours | 0.003546 |
| LogisticRegression | 36 | baseline | `has_prior_visit` | Has prior visit | 0.003289 |
| LogisticRegression | 37 | treatment exposure | `repeat_drug_exposure_count_24h` | Repeat drug exposure count, first 24 hours | 0.003192 |
| LogisticRegression | 38 | treatment exposure | `drug_formulary_vancobase_228466e611_exposed_24h` | Iso-Osmotic Dextrose - exposure indicator, first 24 hours | 0.003144 |
| LogisticRegression | 39 | physiological severity | `lab_50882_bicarbonate_mean` | Bicarbonate - mean, first 24 hours | 0.003142 |
| LogisticRegression | 40 | treatment exposure | `drug_gsn_043952_0602de6ed2_exposed_24h` | Vancomycin - exposure indicator, first 24 hours | 0.003092 |
| LogisticRegression | 41 | physiological severity | `lab_50820_ph_min` | pH - minimum, first 24 hours | 0.003089 |
| LogisticRegression | 42 | treatment exposure | `drug_gsn_057959_a518f33e58_exposed_24h` | Chlorhexidine Gluconate 0.12% Oral Rinse - exposure indicator, first 24 hours | 0.002984 |
| LogisticRegression | 43 | physiological severity | `lab_51237_inr_pt_max` | INR(PT) - maximum, first 24 hours | 0.002960 |
| LogisticRegression | 44 | treatment exposure | `drug_gsn_001972_b5f0a0eff8_count_24h` | 5% Dextrose - count, first 24 hours | 0.002878 |
| LogisticRegression | 45 | procedure burden | `procedure_icd9_3893_49b890a736_presence_24h` | ICD-9 procedure 3893 - presence indicator, first 24 hours | 0.002823 |
| LogisticRegression | 46 | physiological severity | `lab_50821_po2_mean` | pO2 - mean, first 24 hours | 0.002332 |
| LogisticRegression | 47 | treatment exposure | `drug_gsn_001210_ba3dc43c8d_count_24h` | 0.9% Sodium Chloride - count, first 24 hours | 0.002295 |
| LogisticRegression | 48 | treatment exposure | `drug_gsn_048287_21c090e9e3_exposed_24h` | Fentanyl Citrate - exposure indicator, first 24 hours | 0.002290 |
| LogisticRegression | 49 | procedure burden | `procedure_icd9_3893_49b890a736_count_24h` | ICD-9 procedure 3893 - count, first 24 hours | 0.002228 |
| LogisticRegression | 50 | physiological severity | `lab_50893_calcium_total_mean` | Calcium, Total - mean, first 24 hours | 0.002205 |

Continued on next page

eTable 32. MIMIC-IV SHAP feature importance for Baseline + all clinical domains. (continued)

| Model | Rank or source order | Feature domain | Original feature name | Readable concept or feature name | Mean absolute SHAP |
|---|---|---|---|---|---|
| LogisticRegression | 51 | physiological severity | lab_50893_calcium_total_count | Calcium, Total - count, first 24 hours | 0.002152 |
| LogisticRegression | 52 | physiological severity | lab_50970_phosphate_count | Phosphate - count, first 24 hours | 0.002127 |
| LogisticRegression | 53 | physiological severity | lab_50813_lactate_min | Lactate - minimum, first 24 hours | 0.001948 |
| LogisticRegression | 54 | physiological severity | lab_50813_lactate_mean | Lactate - mean, first 24 hours | 0.001915 |
| LogisticRegression | 55 | physiological severity | lab_50983_sodium_min | Sodium - minimum, first 24 hours | 0.001826 |
| LogisticRegression | 56 | treatment exposure | any_drug_24h | Any drug exposure, first 24 hours | 0.001734 |
| LogisticRegression | 57 | treatment exposure | drug_gsn_043952_0602de6ed2_count_24h | Vancomycin - count, first 24 hours | 0.001697 |
| LogisticRegression | 58 | physiological severity | lab_51222_hemoglobin_max | Hemoglobin - maximum, first 24 hours | 0.001613 |
| LogisticRegression | 59 | physiological severity | lab_50983_sodium_std | Sodium - standard deviation, first 24 hours | 0.001607 |
| LogisticRegression | 60 | treatment exposure | drug_formulary_vancobase_228466e611_count_24h | Iso-Osmotic Dextrose - count, first 24 hours | 0.001595 |
| LogisticRegression | 61 | physiological severity | lab_50902_chloride_count | Chloride - count, first 24 hours | 0.001537 |
| LogisticRegression | 62 | treatment exposure | drug_gsn_057959_a518f33e58_count_24h | Chlorhexidine Gluconate 0.12% Oral Rinse - count, first 24 hours | 0.001456 |
| LogisticRegression | 63 | physiological severity | lab_51237_inr_pt_mean | INR(PT) - mean, first 24 hours | 0.001439 |
| LogisticRegression | 64 | procedure burden | procedure_icd9_9671_5bce41582d_presence_24h | ICD-9 procedure 9671 - presence indicator, first 24 hours | 0.001420 |
| LogisticRegression | 65 | physiological severity | lab_50821_po2_std | pO2 - standard deviation, first 24 hours | 0.001392 |
| LogisticRegression | 66 | physiological severity | lab_50804_calculated_total_co2_mean | Calculated Total CO2 - mean, first 24 hours | 0.001342 |
| LogisticRegression | 67 | treatment exposure | drug_gsn_048287_21c090e9e3_count_24h | Fentanyl Citrate - count, first 24 hours | 0.001292 |
| LogisticRegression | 68 | procedure burden | procedure_icd10_5a1945z_f9e7fcf8fd_count_24h | ICD-10-PCS procedure 5A1945Z - count, first 24 hours | 0.001238 |
| LogisticRegression | 69 | treatment exposure | drug_formulary_vial_b4c8f642b1_count_24h | Vial - count, first 24 hours | 0.001215 |
| LogisticRegression | 70 | procedure burden | procedure_icd10_5a1955z_6a2b52611f_presence_24h | ICD-10-PCS procedure 5A1955Z - presence indicator, first 24 hours | 0.001214 |
| LogisticRegression | 71 | physiological severity | lab_50821_po2_max | pO2 - maximum, first 24 hours | 0.001201 |
| LogisticRegression | 72 | physiological severity | lab_51274_pt_mean | PT - mean, first 24 hours | 0.001165 |
| LogisticRegression | 73 | treatment exposure | drug_gsn_016796_f69d8264fe_exposed_24h | Propofol - exposure indicator, first 24 hours | 0.001146 |
| LogisticRegression | 74 | procedure burden | procedure_icd10_5a1945z_f9e7fcf8fd_presence_24h | ICD-10-PCS procedure 5A1945Z - presence indicator, first 24 hours | 0.001135 |

Continued on next page

eTable 32. MIMIC-IV SHAP feature importance for Baseline + all clinical domains. (continued)

| Model | Rank or source order | Feature domain | Original feature name | Readable concept or feature name | Mean absolute SHAP |
|---|---|---|---|---|---|
| LogisticRegression | 75 | physiological severity | `lab_50820_ph_std` | pH - standard deviation, first 24 hours | 0.001123 |
| LogisticRegression | 76 | physiological severity | `lab_50813_lactate_max` | Lactate - maximum, first 24 hours | 0.001087 |
| LogisticRegression | 77 | physiological severity | `lab_50882_bicarbonate_std` | Bicarbonate - standard deviation, first 24 hours | 0.001073 |
| LogisticRegression | 78 | procedure burden | `procedure_icd10_5a1955z_6a2b52611f_count_24h` | ICD-10-PCS procedure 5A1955Z - count, first 24 hours | 0.001026 |
| LogisticRegression | 79 | procedure burden | `any_procedure_24h` | Any procedure recorded, first 24 hours | 0.000956 |
| LogisticRegression | 80 | physiological severity | `lab_50804_calculated_total_co2_std` | Calculated Total CO2 - standard deviation, first 24 hours | 0.000956 |
| LogisticRegression | 81 | physiological severity | `lab_51274_pt_max` | PT - maximum, first 24 hours | 0.000943 |
| LogisticRegression | 82 | treatment exposure | `drug_gsn_001972_b5f0a0eff8_exposed_24h` | 5% Dextrose - exposure indicator, first 24 hours | 0.000927 |
| LogisticRegression | 83 | physiological severity | `lab_51301_white_blood_cells_max` | White Blood Cells - maximum, first 24 hours | 0.000775 |
| LogisticRegression | 84 | treatment exposure | `time_to_first_drug_hours` | Time to first drug exposure, hours | 0.000753 |
| LogisticRegression | 85 | procedure burden | `procedure_icd9_9671_5bce41582d_count_24h` | ICD-9 procedure 9671 - count, first 24 hours | 0.000706 |
| LogisticRegression | 86 | physiological severity | `lab_50893_calcium_total_max` | Calcium, Total - maximum, first 24 hours | 0.000659 |
| LogisticRegression | 87 | procedure burden | `procedure_icd9_9604_93039b475e_presence_24h` | ICD-9 procedure 9604 - presence indicator, first 24 hours | 0.000637 |
| LogisticRegression | 88 | physiological severity | `lab_50821_po2_min` | pO2 - minimum, first 24 hours | 0.000610 |
| LogisticRegression | 89 | physiological severity | `lab_50818_pco2_std` | pCO2 - standard deviation, first 24 hours | 0.000598 |
| LogisticRegression | 90 | treatment exposure | `unique_drug_count_24h` | Unique drug count, first 24 hours | 0.000550 |
| LogisticRegression | 91 | treatment exposure | `drug_gsn_016796_f69d8264fe_count_24h` | Propofol - count, first 24 hours | 0.000533 |
| LogisticRegression | 92 | procedure burden | `procedure_icd10_3e0g76z_fa1c50513e_count_24h` | ICD-10-PCS procedure 3E0G76Z - count, first 24 hours | 0.000528 |
| LogisticRegression | 93 | procedure burden | `procedure_icd10_3e0g76z_fa1c50513e_presence_24h` | ICD-10-PCS procedure 3E0G76Z - presence indicator, first 24 hours | 0.000513 |
| LogisticRegression | 94 | procedure burden | `procedure_icd9_9604_93039b475e_count_24h` | ICD-9 procedure 9604 - count, first 24 hours | 0.000506 |
| LogisticRegression | 95 | procedure burden | `procedure_icd9_9672_93b639be5d_presence_24h` | ICD-9 procedure 9672 - presence indicator, first 24 hours | 0.000290 |
| LogisticRegression | 96 | physiological severity | `lab_50954_lactate_dehydrogenase_ld_min` | Lactate Dehydrogenase (LD) - minimum, first 24 hours | 0.000289 |
| LogisticRegression | 97 | procedure burden | `procedure_icd9_9672_93b639be5d_count_24h` | ICD-9 procedure 9672 - count, first 24 hours | 0.000266 |
| LogisticRegression | 98 | treatment exposure | `drug_formulary_norebasens_c31a0d47cc_count_24h` | 0.9% Sodium Chloride - count, first 24 hours | 0.000073 |
| LogisticRegression | 99 | treatment exposure | `drug_gsn_066419_c17fadb0e8_count_24h` | NORepinephrine - count, first 24 hours | 0.000070 |

Continued on next page

eTable 32. MIMIC-IV SHAP feature importance for Baseline + all clinical domains. (continued)

| Model | Rank or source order | Feature domain | Original feature name | Readable concept or feature name | Mean absolute SHAP |
|---|---|---|---|---|---|
| LogisticRegression | 100 | procedure burden | `procedure_icd9_3891_71dd24c736_presence_24h` | ICD-9 procedure 3891 - presence indicator, first 24 hours | 0.000065 |
| LogisticRegression | 101 | procedure burden | `procedure_icd9_3891_71dd24c736_count_24h` | ICD-9 procedure 3891 - count, first 24 hours | 0.000058 |
| LogisticRegression | 102 | procedure burden | `procedure_icd10_0bh17ez_3d5ba254ee_presence_24h` | ICD-10-PCS procedure 0BH17EZ - presence indicator, first 24 hours | 0.000039 |
| LogisticRegression | 103 | procedure burden | `procedure_icd10_0bh17ez_3d5ba254ee_count_24h` | ICD-10-PCS procedure 0BH17EZ - count, first 24 hours | 0.000037 |

*Notes:* Rows are reproduced from the saved MIMIC-IV SHAP summary output. When a rank column was absent in the source file, source row order is shown. SHAP values were generated from the best-performing algorithm selected for each feature matrix. Best-performing algorithms were selected by highest mean cross-validated AUPRC, with mean AUROC used as the tie-breaker. SHAP values are model-specific attributions, not causal effects; absolute SHAP magnitudes should not be compared directly across separately trained models or datasets.

### Features Selected per Fold

The CHoRUS baseline analysis used the fixed 21-feature baseline list shown above. The saved MIMIC-IV analysis outputs show a fixed set of nine raw baseline predictors in every fold; categorical baseline predictors are one-hot encoded inside the model pipelines. All baseline demographic and vulnerability variables were used in every fold. Broad category labels are shown for categorical MIMIC-IV predictors, so entries such as `sex`, `race`, `ethnicity`, and `visit_type` summarize encoded categorical predictors rather than listing every one-hot indicator.

eTable 33. CHoRUS features selected per fold.

| Feature Matrix | Fold | Feature | Concept name |
|---|---|---|---|
| Baseline + physiological severity | 1 | measurement_4301868_missing_24h | Pulse rate — missingness indicator, first 24 hours |
| Baseline + physiological severity | 1 | measurement_4020553_missing_24h | Oxygen saturation measurement — missingness indicator, first 24 hours |
| Baseline + physiological severity | 1 | measurement_3004249_min_24h | Systolic blood pressure — minimum, first 24 hours |
| Baseline + physiological severity | 1 | measurement_4301868_count_24h | Pulse rate — measurement count, first 24 hours |
| Baseline + physiological severity | 1 | measurement_4301868_std_24h | Pulse rate — standard deviation, first 24 hours |
| Baseline + physiological severity | 1 | measurement_4020553_count_24h | Oxygen saturation measurement — measurement count, first 24 hours |
| Baseline + physiological severity | 1 | measurement_4020553_mean_24h | Oxygen saturation measurement — mean, first 24 hours |
| Baseline + physiological severity | 1 | measurement_4020553_min_24h | Oxygen saturation measurement — minimum, first 24 hours |
| Baseline + physiological severity | 1 | measurement_4020553_max_24h | Oxygen saturation measurement — maximum, first 24 hours |
| Baseline + physiological severity | 1 | measurement_4020553_std_24h | Oxygen saturation measurement — standard deviation, first 24 hours |
| Baseline + physiological severity | 1 | measurement_3035816_mean_24h | Nutrition intake pattern Braden scale — mean, first 24 hours |
| Baseline + physiological severity | 1 | measurement_4301868_min_24h | Pulse rate — minimum, first 24 hours |
| Baseline + physiological severity | 1 | measurement_4301868_mean_24h | Pulse rate — mean, first 24 hours |
| Baseline + physiological severity | 1 | measurement_3004249_std_24h | Systolic blood pressure — standard deviation, first 24 hours |
| Baseline + physiological severity | 1 | measurement_3035206_min_24h | Physical mobility Braden scale — minimum, first 24 hours |
| Baseline + physiological severity | 1 | measurement_3004249_mean_24h | Systolic blood pressure — mean, first 24 hours |
| Baseline + physiological severity | 1 | measurement_3024171_count_24h | Respiratory rate — measurement count, first 24 hours |
| Baseline + physiological severity | 1 | measurement_3036098_mean_24h | Sensory perception Braden scale — mean, first 24 hours |
| Baseline + physiological severity | 1 | measurement_3037347_max_24h | Friction and shear Braden scale — maximum, first 24 hours |
| Baseline + physiological severity | 1 | measurement_4301868_max_24h | Pulse rate — maximum, first 24 hours |
| Baseline + physiological severity | 1 | measurement_3035206_mean_24h | Physical mobility Braden scale — mean, first 24 hours |

Continued on next page

eTable 33. CHoRUS features selected per fold. (continued)

| Feature Matrix | Fold | Feature | Concept name |
|---|---|---|---|
| Baseline + physiological severity | 2 | measurement_4301868_missing_24h | Pulse rate — missingness indicator, first 24 hours |
| Baseline + physiological severity | 2 | measurement_4020553_max_24h | Oxygen saturation measurement — maximum, first 24 hours |
| Baseline + physiological severity | 2 | measurement_4020553_std_24h | Oxygen saturation measurement — standard deviation, first 24 hours |
| Baseline + physiological severity | 2 | measurement_3024171_count_24h | Respiratory rate — measurement count, first 24 hours |
| Baseline + physiological severity | 2 | measurement_4020553_missing_24h | Oxygen saturation measurement — missingness indicator, first 24 hours |
| Baseline + physiological severity | 2 | measurement_4301868_count_24h | Pulse rate — measurement count, first 24 hours |
| Baseline + physiological severity | 2 | measurement_4020553_mean_24h | Oxygen saturation measurement — mean, first 24 hours |
| Baseline + physiological severity | 2 | measurement_3004249_min_24h | Systolic blood pressure — minimum, first 24 hours |
| Baseline + physiological severity | 2 | measurement_3004249_std_24h | Systolic blood pressure — standard deviation, first 24 hours |
| Baseline + physiological severity | 2 | measurement_4020553_min_24h | Oxygen saturation measurement — minimum, first 24 hours |
| Baseline + physiological severity | 2 | measurement_4301868_std_24h | Pulse rate — standard deviation, first 24 hours |
| Baseline + physiological severity | 2 | measurement_4020553_count_24h | Oxygen saturation measurement — measurement count, first 24 hours |
| Baseline + physiological severity | 2 | measurement_4301868_max_24h | Pulse rate — maximum, first 24 hours |
| Baseline + physiological severity | 2 | measurement_3004249_mean_24h | Systolic blood pressure — mean, first 24 hours |
| Baseline + physiological severity | 2 | measurement_3024171_max_24h | Respiratory rate — maximum, first 24 hours |
| Baseline + physiological severity | 2 | measurement_4301868_mean_24h | Pulse rate — mean, first 24 hours |
| Baseline + physiological severity | 2 | measurement_4301868_min_24h | Pulse rate — minimum, first 24 hours |
| Baseline + physiological severity | 2 | measurement_3036098_mean_24h | Sensory perception Braden scale — mean, first 24 hours |
| Baseline + physiological severity | 2 | measurement_3024171_missing_24h | Respiratory rate — missingness indicator, first 24 hours |
| Baseline + physiological severity | 2 | measurement_3037318_max_24h | Physical activity Braden scale — maximum, first 24 hours |
| Baseline + physiological severity | 2 | measurement_3037347_mean_24h | Friction and shear Braden scale — mean, first 24 hours |
| Baseline + physiological severity | 3 | measurement_4301868_missing_24h | Pulse rate — missingness indicator, first 24 hours |
| Baseline + physiological severity | 3 | measurement_4020553_missing_24h | Oxygen saturation measurement — missingness indicator, first 24 hours |
| Baseline + physiological severity | 3 | measurement_4020553_count_24h | Oxygen saturation measurement — measurement count, first 24 hours |
| Baseline + physiological severity | 3 | measurement_4301868_std_24h | Pulse rate — standard deviation, first 24 hours |
| Baseline + physiological severity | 3 | measurement_4020553_max_24h | Oxygen saturation measurement — maximum, first 24 hours |
| Baseline + physiological severity | 3 | measurement_4020553_std_24h | Oxygen saturation measurement — standard deviation, first 24 hours |
| Baseline + physiological severity | 3 | measurement_3004249_std_24h | Systolic blood pressure — standard deviation, first 24 hours |
| Baseline + physiological severity | 3 | measurement_4301868_max_24h | Pulse rate — maximum, first 24 hours |
| Baseline + physiological severity | 3 | measurement_3024171_count_24h | Respiratory rate — measurement count, first 24 hours |
| Baseline + physiological severity | 3 | measurement_3004249_min_24h | Systolic blood pressure — minimum, first 24 hours |
| Baseline + physiological severity | 3 | measurement_4020553_min_24h | Oxygen saturation measurement — minimum, first 24 hours |
| Baseline + physiological severity | 3 | measurement_3036098_min_24h | Sensory perception Braden scale — minimum, first 24 hours |
| Baseline + physiological severity | 3 | measurement_4301868_count_24h | Pulse rate — measurement count, first 24 hours |
| Baseline + physiological severity | 3 | measurement_3035206_mean_24h | Physical mobility Braden scale — mean, first 24 hours |
| Baseline + physiological severity | 3 | measurement_4301868_min_24h | Pulse rate — minimum, first 24 hours |
| Baseline + physiological severity | 3 | measurement_3037347_mean_24h | Friction and shear Braden scale — mean, first 24 hours |
| Baseline + physiological severity | 3 | measurement_4301868_mean_24h | Pulse rate — mean, first 24 hours |
| Baseline + physiological severity | 3 | measurement_3004249_mean_24h | Systolic blood pressure — mean, first 24 hours |
| Baseline + physiological severity | 3 | measurement_3035206_min_24h | Physical mobility Braden scale — minimum, first 24 hours |
| Baseline + physiological severity | 3 | measurement_3035816_max_24h | Nutrition intake pattern Braden scale — maximum, first 24 hours |

Continued on next page

eTable 33. CHoRUS features selected per fold. (continued)

| Feature Matrix | Fold | Feature | Concept name |
|---|---|---|---|
| Baseline + physiological severity | 3 | measurement_3024171_max_24h | Respiratory rate — maximum, first 24 hours |
| Baseline + physiological severity | 4 | measurement_4020553_missing_24h | Oxygen saturation measurement — missingness indicator, first 24 hours |
| Baseline + physiological severity | 4 | measurement_4301868_missing_24h | Pulse rate — missingness indicator, first 24 hours |
| Baseline + physiological severity | 4 | measurement_4020553_min_24h | Oxygen saturation measurement — minimum, first 24 hours |
| Baseline + physiological severity | 4 | measurement_4020553_max_24h | Oxygen saturation measurement — maximum, first 24 hours |
| Baseline + physiological severity | 4 | measurement_4301868_count_24h | Pulse rate — measurement count, first 24 hours |
| Baseline + physiological severity | 4 | measurement_4301868_mean_24h | Pulse rate — mean, first 24 hours |
| Baseline + physiological severity | 4 | measurement_4020553_count_24h | Oxygen saturation measurement — measurement count, first 24 hours |
| Baseline + physiological severity | 4 | measurement_4020553_std_24h | Oxygen saturation measurement — standard deviation, first 24 hours |
| Baseline + physiological severity | 4 | measurement_4301868_max_24h | Pulse rate — maximum, first 24 hours |
| Baseline + physiological severity | 4 | measurement_3004249_min_24h | Systolic blood pressure — minimum, first 24 hours |
| Baseline + physiological severity | 4 | measurement_4301868_std_24h | Pulse rate — standard deviation, first 24 hours |
| Baseline + physiological severity | 4 | measurement_4301868_min_24h | Pulse rate — minimum, first 24 hours |
| Baseline + physiological severity | 4 | measurement_3024171_count_24h | Respiratory rate — measurement count, first 24 hours |
| Baseline + physiological severity | 4 | measurement_4020553_mean_24h | Oxygen saturation measurement — mean, first 24 hours |
| Baseline + physiological severity | 4 | measurement_3004249_std_24h | Systolic blood pressure — standard deviation, first 24 hours |
| Baseline + physiological severity | 4 | measurement_3004249_mean_24h | Systolic blood pressure — mean, first 24 hours |
| Baseline + physiological severity | 4 | measurement_3024171_max_24h | Respiratory rate — maximum, first 24 hours |
| Baseline + physiological severity | 4 | measurement_3036098_mean_24h | Sensory perception Braden scale — mean, first 24 hours |
| Baseline + physiological severity | 4 | measurement_3035206_mean_24h | Physical mobility Braden scale — mean, first 24 hours |
| Baseline + physiological severity | 4 | measurement_3024171_missing_24h | Respiratory rate — missingness indicator, first 24 hours |
| Baseline + physiological severity | 4 | measurement_3024171_mean_24h | Respiratory rate — mean, first 24 hours |
| Baseline + physiological severity | 5 | measurement_4301868_missing_24h | Pulse rate — missingness indicator, first 24 hours |
| Baseline + physiological severity | 5 | measurement_3004249_min_24h | Systolic blood pressure — minimum, first 24 hours |
| Baseline + physiological severity | 5 | measurement_4020553_missing_24h | Oxygen saturation measurement — missingness indicator, first 24 hours |
| Baseline + physiological severity | 5 | measurement_4301868_count_24h | Pulse rate — measurement count, first 24 hours |
| Baseline + physiological severity | 5 | measurement_4301868_max_24h | Pulse rate — maximum, first 24 hours |
| Baseline + physiological severity | 5 | measurement_3024171_count_24h | Respiratory rate — measurement count, first 24 hours |
| Baseline + physiological severity | 5 | measurement_4020553_mean_24h | Oxygen saturation measurement — mean, first 24 hours |
| Baseline + physiological severity | 5 | measurement_4301868_std_24h | Pulse rate — standard deviation, first 24 hours |
| Baseline + physiological severity | 5 | measurement_4301868_min_24h | Pulse rate — minimum, first 24 hours |
| Baseline + physiological severity | 5 | measurement_4020553_min_24h | Oxygen saturation measurement — minimum, first 24 hours |
| Baseline + physiological severity | 5 | measurement_4020553_count_24h | Oxygen saturation measurement — measurement count, first 24 hours |
| Baseline + physiological severity | 5 | measurement_4020553_std_24h | Oxygen saturation measurement — standard deviation, first 24 hours |
| Baseline + physiological severity | 5 | measurement_4020553_max_24h | Oxygen saturation measurement — maximum, first 24 hours |
| Baseline + physiological severity | 5 | measurement_4301868_mean_24h | Pulse rate — mean, first 24 hours |
| Baseline + physiological severity | 5 | measurement_3004249_mean_24h | Systolic blood pressure — mean, first 24 hours |
| Baseline + physiological severity | 5 | measurement_3037318_max_24h | Physical activity Braden scale — maximum, first 24 hours |
| Baseline + physiological severity | 5 | measurement_3036098_max_24h | Sensory perception Braden scale — maximum, first 24 hours |
| Baseline + physiological severity | 5 | measurement_3035206_mean_24h | Physical mobility Braden scale — mean, first 24 hours |
| Baseline + physiological severity | 5 | measurement_3037347_mean_24h | Friction and shear Braden scale — mean, first 24 hours |

Continued on next page

eTable 33. CHoRUS features selected per fold. (continued)

| Feature Matrix | Fold | Feature | Concept name |
|---|---|---|---|
| Baseline + physiological severity | 5 | measurement_3004249_std_24h | Systolic blood pressure — standard deviation, first 24 hours |
| Baseline + physiological severity | 5 | measurement_3035816_mean_24h | Nutrition intake pattern Braden scale — mean, first 24 hours |
| Baseline + treatment exposure | 1 | total_drug_exposure_count_24h | Total drug-exposure count, first 24 hours |
| Baseline + treatment exposure | 1 | repeated_drug_exposure_count_24h | Repeated drug-exposure count, first 24 hours |
| Baseline + treatment exposure | 1 | drug_43011850_count24h | heparin sodium, porcine 5000 UNT/ML Injectable Solution — exposure count, first 24 hours |
| Baseline + treatment exposure | 1 | drug_19135374_count24h | calcium chloride 0.2 MG/ML / potassium chloride 0.3 MG/ML / sodium chloride 6 MG/ML / sodium lactate 3.1 MG/ML Injectable Solution — exposure count, first 24 hours |
| Baseline + treatment exposure | 1 | drug_40221385_count24h | 100 ML sodium chloride 9 MG/ML Injection — exposure count, first 24 hours |
| Baseline + treatment exposure | 1 | unique_drug_exposure_count_24h | Unique drug-exposure count, first 24 hours |
| Baseline + treatment exposure | 1 | time_to_first_drug_hours | Time to first drug exposure, hours |
| Baseline + treatment exposure | 1 | drug_43011850_exposed_24h | heparin sodium, porcine 5000 UNT/ML Injectable Solution — exposure indicator, first 24 hours |
| Baseline + treatment exposure | 1 | drug_19135374_exposed_24h | calcium chloride 0.2 MG/ML / potassium chloride 0.3 MG/ML / sodium chloride 6 MG/ML / sodium lactate 3.1 MG/ML Injectable Solution — exposure indicator, first 24 hours |
| Baseline + treatment exposure | 1 | drug_40048832_count24h | ketorolac Injectable Solution [Toradol] — exposure count, first 24 hours |
| Baseline + treatment exposure | 1 | drug_46275280_count24h | 50 ML magnesium sulfate 40 MG/ML Injection — exposure count, first 24 hours |
| Baseline + treatment exposure | 1 | drug_35605482_count24h | 2 ML ondansetron 2 MG/ML Injection — exposure count, first 24 hours |
| Baseline + treatment exposure | 1 | drug_19079524_count24h | sodium chloride 9 MG/ML Injectable Solution — exposure count, first 24 hours |
| Baseline + treatment exposure | 1 | drug_1560751_count24h | 50 ML glucose 500 MG/ML Prefilled Syringe — exposure count, first 24 hours |
| Baseline + treatment exposure | 1 | drug_40221385_exposed_24h | 100 ML sodium chloride 9 MG/ML Injection — exposure indicator, first 24 hours |
| Baseline + treatment exposure | 1 | drug_40220357_exposed_24h | 1000 ML sodium chloride 9 MG/ML Injection — exposure indicator, first 24 hours |
| Baseline + treatment exposure | 1 | drug_2025002_count24h | 250 ML sodium chloride 9 MG/ML Injectable Solution [JW NS] by JW — exposure count, first 24 hours |
| Baseline + treatment exposure | 1 | drug_35830655_count24h | cefazolin 1000 MG Injectable Solution [CEFAMEZIN] — exposure count, first 24 hours |
| Baseline + treatment exposure | 1 | drug_35854313_count24h | iohexol 350 MG/ML Injectable Solution — exposure count, first 24 hours |
| Baseline + treatment exposure | 1 | drug_46275280_exposed_24h | 50 ML magnesium sulfate 40 MG/ML Injection — exposure indicator, first 24 hours |
| Baseline + treatment exposure | 1 | drug_40220388_count24h | 100 ML propofol 10 MG/ML Injection [Diprivan] — exposure count, first 24 hours |
| Baseline + treatment exposure | 2 | total_drug_exposure_count_24h | Total drug-exposure count, first 24 hours |
| Baseline + treatment exposure | 2 | repeated_drug_exposure_count_24h | Repeated drug-exposure count, first 24 hours |
| Baseline + treatment exposure | 2 | drug_19135374_count24h | calcium chloride 0.2 MG/ML / potassium chloride 0.3 MG/ML / sodium chloride 6 MG/ML / sodium lactate 3.1 MG/ML Injectable Solution — exposure count, first 24 hours |
| Baseline + treatment exposure | 2 | time_to_first_drug_hours | Time to first drug exposure, hours |

Continued on next page

eTable 33. CHoRUS features selected per fold. (continued)

| Feature Matrix | Fold | Feature | Concept name |
|---|---|---|---|
| Baseline + treatment exposure | 2 | unique_drug_exposure_count_24h | Unique drug-exposure count, first 24 hours |
| Baseline + treatment exposure | 2 | drug_43011850_count24h | heparin sodium, porcine 5000 UNT/ML Injectable Solution — exposure count, first 24 hours |
| Baseline + treatment exposure | 2 | drug_43011850_exposed_24h | heparin sodium, porcine 5000 UNT/ML Injectable Solution — exposure indicator, first 24 hours |
| Baseline + treatment exposure | 2 | drug_46275280_count24h | 50 ML magnesium sulfate 40 MG/ML Injection — exposure count, first 24 hours |
| Baseline + treatment exposure | 2 | drug_40744996_count24h | Sugammadex 100 MG/ML Injectable Solution [Bridion] — exposure count, first 24 hours |
| Baseline + treatment exposure | 2 | drug_40220357_exposed_24h | 1000 ML sodium chloride 9 MG/ML Injection — exposure indicator, first 24 hours |
| Baseline + treatment exposure | 2 | drug_40221385_exposed_24h | 100 ML sodium chloride 9 MG/ML Injection — exposure indicator, first 24 hours |
| Baseline + treatment exposure | 2 | drug_19135374_exposed_24h | calcium chloride 0.2 MG/ML / potassium chloride 0.3 MG/ML / sodium chloride 6 MG/ML / sodium lactate 3.1 MG/ML Injectable Solution — exposure indicator, first 24 hours |
| Baseline + treatment exposure | 2 | drug_40220388_count24h | 100 ML propofol 10 MG/ML Injection [Diprivan] — exposure count, first 24 hours |
| Baseline + treatment exposure | 2 | drug_19079322_exposed_24h | 100 ML potassium chloride 0.1 MEQ/ML Injection — exposure indicator, first 24 hours |
| Baseline + treatment exposure | 2 | drug_42479436_count24h | 1000 ML Sodium Chloride 9 MG/ML Injectable Solution — exposure count, first 24 hours |
| Baseline + treatment exposure | 2 | drug_1560751_count24h | 50 ML glucose 500 MG/ML Prefilled Syringe — exposure count, first 24 hours |
| Baseline + treatment exposure | 2 | drug_19005968_count24h | ondansetron 2 MG/ML Injectable Solution [Zofran] — exposure count, first 24 hours |
| Baseline + treatment exposure | 2 | drug_1110410_count24h | morphine — exposure count, first 24 hours |
| Baseline + treatment exposure | 2 | drug_35605482_count24h | 2 ML ondansetron 2 MG/ML Injection — exposure count, first 24 hours |
| Baseline + treatment exposure | 2 | drug_35854313_count24h | iohexol 350 MG/ML Injectable Solution — exposure count, first 24 hours |
| Baseline + treatment exposure | 2 | drug_35830655_count_24h | cefazolin 1000 MG Injectable Solution [CEFAMEZIN] — exposure count, first 24 hours |
| Baseline + treatment exposure | 3 | total_drug_exposure_count_24h | Total drug-exposure count, first 24 hours |
| Baseline + treatment exposure | 3 | repeated_drug_exposure_count_24h | Repeated drug-exposure count, first 24 hours |
| Baseline + treatment exposure | 3 | time_to_first_drug_hours | Time to first drug exposure, hours |
| Baseline + treatment exposure | 3 | unique_drug_exposure_count_24h | Unique drug-exposure count, first 24 hours |
| Baseline + treatment exposure | 3 | drug_43011850_count24h | heparin sodium, porcine 5000 UNT/ML Injectable Solution — exposure count, first 24 hours |
| Baseline + treatment exposure | 3 | drug_19135374_count24h | calcium chloride 0.2 MG/ML / potassium chloride 0.3 MG/ML / sodium chloride 6 MG/ML / sodium lactate 3.1 MG/ML Injectable Solution — exposure count, first 24 hours |
| Baseline + treatment exposure | 3 | drug_43011850_exposed_24h | heparin sodium, porcine 5000 UNT/ML Injectable Solution — exposure indicator, first 24 hours |
| Baseline + treatment exposure | 3 | drug_19135374_exposed_24h | calcium chloride 0.2 MG/ML / potassium chloride 0.3 MG/ML / sodium chloride 6 MG/ML / sodium lactate 3.1 MG/ML Injectable Solution — exposure indicator, first 24 hours |

Continued on next page

eTable 33. CHoRUS features selected per fold. (continued)

| Feature Matrix | Fold | Feature | Concept name |
|---|---|---|---|
| Baseline + treatment exposure | 3 | drug_40048832_count24h | ketorolac Injectable Solution [Toradol] — exposure count, first 24 hours |
| Baseline + treatment exposure | 3 | drug_40221385_count24h | 100 ML sodium chloride 9 MG/ML Injection — exposure count, first 24 hours |
| Baseline + treatment exposure | 3 | drug_40221385_exposed_24h | 100 ML sodium chloride 9 MG/ML Injection — exposure indicator, first 24 hours |
| Baseline + treatment exposure | 3 | drug_40220357_exposed_24h | 1000 ML sodium chloride 9 MG/ML Injection — exposure indicator, first 24 hours |
| Baseline + treatment exposure | 3 | drug_40220390_count24h | propofol 10 MG/ML Injection [Diprivan] — exposure count, first 24 hours |
| Baseline + treatment exposure | 3 | drug_46275280_count24h | 50 ML magnesium sulfate 40 MG/ML Injection — exposure count, first 24 hours |
| Baseline + treatment exposure | 3 | drug_40042753_count24h | enoxaparin Injectable Solution [Lovenox] — exposure count, first 24 hours |
| Baseline + treatment exposure | 3 | drug_40220357_count24h | 1000 ML sodium chloride 9 MG/ML Injection — exposure count, first 24 hours |
| Baseline + treatment exposure | 3 | drug_35854313_count24h | iohexol 350 MG/ML Injectable Solution — exposure count, first 24 hours |
| Baseline + treatment exposure | 3 | drug_19073712_count24h | aspirin 81 MG Delayed Release Oral Tablet — exposure count, first 24 hours |
| Baseline + treatment exposure | 3 | drug_35782395_count24h | 10 ML Phenylephrine 0.1 MG/ML Injectable Solution — exposure count, first 24 hours |
| Baseline + treatment exposure | 3 | drug_40220388_exposed_24h | 100 ML propofol 10 MG/ML Injection [Diprivan] — exposure indicator, first 24 hours |
| Baseline + treatment exposure | 3 | drug_19127213_count24h | 10 ML sodium chloride 9 MG/ML Prefilled Syringe — exposure count, first 24 hours |
| Baseline + treatment exposure | 4 | total_drug_exposure_count_24h | Total drug-exposure count, first 24 hours |
| Baseline + treatment exposure | 4 | repeated_drug_exposure_count_24h | Repeated drug-exposure count, first 24 hours |
| Baseline + treatment exposure | 4 | drug_43011850_count24h | heparin sodium, porcine 5000 UNT/ML Injectable Solution — exposure count, first 24 hours |
| Baseline + treatment exposure | 4 | drug_19135374_count24h | calcium chloride 0.2 MG/ML / potassium chloride 0.3 MG/ML / sodium chloride 6 MG/ML / sodium lactate 3.1 MG/ML Injectable Solution — exposure count, first 24 hours |
| Baseline + treatment exposure | 4 | unique_drug_exposure_count_24h | Unique drug-exposure count, first 24 hours |
| Baseline + treatment exposure | 4 | time_to_first_drug_hours | Time to first drug exposure, hours |
| Baseline + treatment exposure | 4 | drug_46275280_count24h | 50 ML magnesium sulfate 40 MG/ML Injection — exposure count, first 24 hours |
| Baseline + treatment exposure | 4 | drug_19135374_exposed_24h | calcium chloride 0.2 MG/ML / potassium chloride 0.3 MG/ML / sodium chloride 6 MG/ML / sodium lactate 3.1 MG/ML Injectable Solution — exposure indicator, first 24 hours |
| Baseline + treatment exposure | 4 | drug_43011850_exposed_24h | heparin sodium, porcine 5000 UNT/ML Injectable Solution — exposure indicator, first 24 hours |
| Baseline + treatment exposure | 4 | drug_1128524_count24h | acetaminophen 650 MG Extended Release Oral Tablet [Tylenol] — exposure count, first 24 hours |
| Baseline + treatment exposure | 4 | drug_19000634_count24h | ondansetron 4 MG [Zofran] — exposure count, first 24 hours |
| Baseline + treatment exposure | 4 | drug_40042753_count24h | enoxaparin Injectable Solution [Lovenox] — exposure count, first 24 hours |

Continued on next page

eTable 33. CHoRUS features selected per fold. (continued)

| Feature Matrix | Fold | Feature | Concept name |
|---|---|---|---|
| Baseline + treatment exposure | 4 | drug_40220357_count24h | 1000 ML sodium chloride 9 MG/ML Injection — exposure count, first 24 hours |
| Baseline + treatment exposure | 4 | drug_40221385_count24h | 100 ML sodium chloride 9 MG/ML Injection — exposure count, first 24 hours |
| Baseline + treatment exposure | 4 | drug_40220357_exposed_24h | 1000 ML sodium chloride 9 MG/ML Injection — exposure indicator, first 24 hours |
| Baseline + treatment exposure | 4 | drug_40221385_exposed_24h | 100 ML sodium chloride 9 MG/ML Injection — exposure indicator, first 24 hours |
| Baseline + treatment exposure | 4 | drug_1560751_count24h | 50 ML glucose 500 MG/ML Prefilled Syringe — exposure count, first 24 hours |
| Baseline + treatment exposure | 4 | drug_1718370_count24h | heparin Injectable Solution — exposure count, first 24 hours |
| Baseline + treatment exposure | 4 | drug_19102651_count24h | sennosides, USP 8.6 MG Oral Tablet [Senokot] — exposure count, first 24 hours |
| Baseline + treatment exposure | 4 | drug_19077513_count24h | folic acid 1 MG Oral Tablet — exposure count, first 24 hours |
| Baseline + treatment exposure | 4 | drug_19079322_exposed_24h | 100 ML potassium chloride 0.1 MEQ/ML Injection — exposure indicator, first 24 hours |
| Baseline + treatment exposure | 5 | total_drug_exposure_count_24h | Total drug-exposure count, first 24 hours |
| Baseline + treatment exposure | 5 | repeated_drug_exposure_count_24h | Repeated drug-exposure count, first 24 hours |
| Baseline + treatment exposure | 5 | drug_43011850_count24h | heparin sodium, porcine 5000 UNT/ML Injectable Solution — exposure count, first 24 hours |
| Baseline + treatment exposure | 5 | time_to_first_drug_hours | Time to first drug exposure, hours |
| Baseline + treatment exposure | 5 | unique_drug_exposure_count_24h | Unique drug-exposure count, first 24 hours |
| Baseline + treatment exposure | 5 | drug_19135374_count24h | calcium chloride 0.2 MG/ML / potassium chloride 0.3 MG/ML / sodium chloride 6 MG/ML / sodium lactate 3.1 MG/ML Injectable Solution — exposure count, first 24 hours |
| Baseline + treatment exposure | 5 | drug_43011850_exposed_24h | heparin sodium, porcine 5000 UNT/ML Injectable Solution — exposure indicator, first 24 hours |
| Baseline + treatment exposure | 5 | drug_40220388_count24h | 100 ML propofol 10 MG/ML Injection [Diprivan] — exposure count, first 24 hours |
| Baseline + treatment exposure | 5 | drug_46275280_count24h | 50 ML magnesium sulfate 40 MG/ML Injection — exposure count, first 24 hours |
| Baseline + treatment exposure | 5 | drug_19135374_exposed_24h | calcium chloride 0.2 MG/ML / potassium chloride 0.3 MG/ML / sodium chloride 6 MG/ML / sodium lactate 3.1 MG/ML Injectable Solution — exposure indicator, first 24 hours |
| Baseline + treatment exposure | 5 | drug_42479436_count24h | 1000 ML Sodium Chloride 9 MG/ML Injectable Solution — exposure count, first 24 hours |
| Baseline + treatment exposure | 5 | drug_40221385_exposed_24h | 100 ML sodium chloride 9 MG/ML Injection — exposure indicator, first 24 hours |
| Baseline + treatment exposure | 5 | drug_35605482_count24h | 2 ML ondansetron 2 MG/ML Injection — exposure count, first 24 hours |
| Baseline + treatment exposure | 5 | drug_1127433_count24h | acetaminophen 325 MG Oral Tablet — exposure count, first 24 hours |
| Baseline + treatment exposure | 5 | drug_19102651_count24h | sennosides, USP 8.6 MG Oral Tablet [Senokot] — exposure count, first 24 hours |
| Baseline + treatment exposure | 5 | drug_40220357_exposed_24h | 1000 ML sodium chloride 9 MG/ML Injection — exposure indicator, first 24 hours |
| Baseline + treatment exposure | 5 | drug_19079524_count24h | sodium chloride 9 MG/ML Injectable Solution — exposure count, first 24 hours |

Continued on next page

eTable 33. CHoRUS features selected per fold. (continued)

| Feature Matrix | Fold | Feature | Concept name |
|---|---|---|---|
| Baseline + treatment exposure | 5 | drug_40220390_count24h | propofol 10 MG/ML Injection [Diprivan] — exposure count, first 24 hours |
| Baseline + treatment exposure | 5 | drug_19127213_count24h | 10 ML sodium chloride 9 MG/ML Prefilled Syringe — exposure count, first 24 hours |
| Baseline + treatment exposure | 5 | drug_21050327_count24h | Glucose Injectable Solution [Plasma-lyte] — exposure count, first 24 hours |
| Baseline + treatment exposure | 5 | drug_42708658_count24h | docusate sodium 100 MG Oral Capsule [Colace] — exposure count, first 24 hours |
| Baseline + procedure burden | 1 | unique_procedure_count_24h | Unique procedure count, first 24 hours |
| Baseline + procedure burden | 1 | procedure_count_total_24h | Total procedure count, first 24 hours |
| Baseline + procedure burden | 1 | procedure_2147482991_presence_24h | LINE CARE \| LINE STATUS \| LINE STATUS — presence indicator, first 24 hours |
| Baseline + procedure burden | 1 | procedure_725068_count24h | Radiologic examination, chest; single view — count, first 24 hours |
| Baseline + procedure burden | 1 | procedure_2147482992_presence_24h | LINE CARE \| LINE SITE ASSESSMENT \| LINE SITE ASSESSMENT — presence indicator, first 24 hours |
| Baseline + procedure burden | 1 | procedure_2147482992_count24h | LINE CARE \| LINE SITE ASSESSMENT \| LINE SITE ASSESSMENT — count, first 24 hours |
| Baseline + procedure burden | 1 | procedure_2147482991_count24h | LINE CARE \| LINE STATUS \| LINE STATUS — count, first 24 hours |
| Baseline + procedure burden | 1 | procedure_2147483387_count24h | GASTROINTESTINAL \| ABDOMEN \| Abdomen INSPECTION — count, first 24 hours |
| Baseline + procedure burden | 1 | procedure_2313814_count24h | Routine 12-lead ECG; with interpretation and report — count, first 24 hours |
| Baseline + procedure burden | 1 | procedure_2313814_presence_24h | Routine 12-lead ECG; with interpretation and report — presence indicator, first 24 hours |
| Baseline + procedure burden | 1 | procedure_2514442_count24h | Critical care evaluation and management; each additional 30 minutes — count, first 24 hours |
| Baseline + procedure burden | 1 | procedure_2514441_count24h | Critical care evaluation and management; first 30–74 minutes — count, first 24 hours |
| Baseline + procedure burden | 1 | procedure_2147483387_presence_24h | GASTROINTESTINAL \| ABDOMEN \| Abdomen INSPECTION — presence indicator, first 24 hours |
| Baseline + procedure burden | 1 | procedure_2514442_presence_24h | Critical care evaluation and management; each additional 30 minutes — presence indicator, first 24 hours |
| Baseline + procedure burden | 1 | procedure_725068_presence_24h | Radiologic examination, chest; single view — presence indicator, first 24 hours |
| Baseline + procedure burden | 1 | procedure_2313816_count24h | Routine 12-lead ECG; interpretation and report only — count, first 24 hours |
| Baseline + procedure burden | 1 | procedure_2514441_presence_24h | Critical care evaluation and management; first 30–74 minutes — presence indicator, first 24 hours |
| Baseline + procedure burden | 1 | procedure_2147483587_count24h | HEENT \| THROAT \| Voice Assessment — count, first 24 hours |
| Baseline + procedure burden | 1 | procedure_2147483056_count24h | ASSISTANCE \| LEVEL OF ASSISTANCE \| LEVEL OF ASSISTANCE — count, first 24 hours |
| Baseline + procedure burden | 1 | procedure_2313816_presence_24h | Routine 12-lead ECG; interpretation and report only — presence indicator, first 24 hours |
| Baseline + procedure burden | 1 | procedure_2147483590_presence_24h | HEENT \| Eyes/Vision \| Right Eye Assessment — presence indicator, first 24 hours |
| Baseline + procedure burden | 2 | unique_procedure_count_24h | Unique procedure count, first 24 hours |

Continued on next page

eTable 33. CHoRUS features selected per fold. (continued)

| Feature Matrix | Fold | Feature | Concept name |
|---|---|---|---|
| Baseline + procedure burden | 2 | procedure_count_total_24h | Total procedure count, first 24 hours |
| Baseline + procedure burden | 2 | procedure_725068_count24h | Radiologic examination, chest; single view — count, first 24 hours |
| Baseline + procedure burden | 2 | procedure_2147482992_count24h | LINE CARE \| LINE SITE ASSESSMENT \| LINE SITE ASSESSMENT — count, first 24 hours |
| Baseline + procedure burden | 2 | procedure_2147482992_presence_24h | LINE CARE \| LINE SITE ASSESSMENT \| LINE SITE ASSESSMENT — presence indicator, first 24 hours |
| Baseline + procedure burden | 2 | procedure_2147482991_count24h | LINE CARE \| LINE STATUS \| LINE STATUS — count, first 24 hours |
| Baseline + procedure burden | 2 | procedure_2313814_count24h | Routine 12-lead ECG; with interpretation and report — count, first 24 hours |
| Baseline + procedure burden | 2 | procedure_2313814_presence_24h | Routine 12-lead ECG; with interpretation and report — presence indicator, first 24 hours |
| Baseline + procedure burden | 2 | procedure_2147482991_presence_24h | LINE CARE \| LINE STATUS \| LINE STATUS — presence indicator, first 24 hours |
| Baseline + procedure burden | 2 | procedure_725068_presence_24h | Radiologic examination, chest; single view — presence indicator, first 24 hours |
| Baseline + procedure burden | 2 | procedure_2514441_count24h | Critical care evaluation and management; first 30–74 minutes — count, first 24 hours |
| Baseline + procedure burden | 2 | procedure_2147483387_presence_24h | GASTROINTESTINAL \| ABDOMEN \| Abdomen INSPECTION — presence indicator, first 24 hours |
| Baseline + procedure burden | 2 | procedure_2147483056_presence_24h | ASSISTANCE \| LEVEL OF ASSISTANCE \| LEVEL OF ASSISTANCE — presence indicator, first 24 hours |
| Baseline + procedure burden | 2 | procedure_2313816_count24h | Routine 12-lead ECG; interpretation and report only — count, first 24 hours |
| Baseline + procedure burden | 2 | procedure_2147483387_count24h | GASTROINTESTINAL \| ABDOMEN \| Abdomen INSPECTION — count, first 24 hours |
| Baseline + procedure burden | 2 | procedure_2147483056_count24h | ASSISTANCE \| LEVEL OF ASSISTANCE \| LEVEL OF ASSISTANCE — count, first 24 hours |
| Baseline + procedure burden | 2 | procedure_2514441_presence_24h | Critical care evaluation and management; first 30–74 minutes — presence indicator, first 24 hours |
| Baseline + procedure burden | 2 | procedure_2147483590_count24h | HEENT \| Eyes/Vision \| Right Eye Assessment — count, first 24 hours |
| Baseline + procedure burden | 2 | procedure_2147483603_count24h | HEENT \| Eyes/Vision \| Left Eye Assessment — count, first 24 hours |
| Baseline + procedure burden | 2 | procedure_2147483590_presence_24h | HEENT \| Eyes/Vision \| Right Eye Assessment — presence indicator, first 24 hours |
| Baseline + procedure burden | 2 | procedure_2147483603_presence_24h | HEENT \| Eyes/Vision \| Left Eye Assessment — presence indicator, first 24 hours |
| Baseline + procedure burden | 3 | unique_procedure_count_24h | Unique procedure count, first 24 hours |
| Baseline + procedure burden | 3 | procedure_count_total_24h | Total procedure count, first 24 hours |
| Baseline + procedure burden | 3 | procedure_2147482992_count24h | LINE CARE \| LINE SITE ASSESSMENT \| LINE SITE ASSESSMENT — count, first 24 hours |
| Baseline + procedure burden | 3 | procedure_725068_count24h | Radiologic examination, chest; single view — count, first 24 hours |
| Baseline + procedure burden | 3 | procedure_2147482992_presence_24h | LINE CARE \| LINE SITE ASSESSMENT \| LINE SITE ASSESSMENT — presence indicator, first 24 hours |

Continued on next page

eTable 33. CHoRUS features selected per fold. (continued)

| Feature Matrix | Fold | Feature | Concept name |
|---|---|---|---|
| Baseline + procedure burden | 3 | procedure_2147482991_count24h | LINE CARE \| LINE STATUS \| LINE STATUS — count, first 24 hours |
| Baseline + procedure burden | 3 | procedure_2147482991_presence_24h | LINE CARE \| LINE STATUS \| LINE STATUS — presence indicator, first 24 hours |
| Baseline + procedure burden | 3 | procedure_2313814_presence_24h | Routine 12-lead ECG; with interpretation and report — presence indicator, first 24 hours |
| Baseline + procedure burden | 3 | procedure_2514441_count24h | Critical care evaluation and management; first 30–74 minutes — count, first 24 hours |
| Baseline + procedure burden | 3 | procedure_2147483387_presence_24h | GASTROINTESTINAL \| ABDOMEN \| Abdomen INSPECTION — presence indicator, first 24 hours |
| Baseline + procedure burden | 3 | procedure_2313814_count24h | Routine 12-lead ECG; with interpretation and report — count, first 24 hours |
| Baseline + procedure burden | 3 | procedure_2147483387_count24h | GASTROINTESTINAL \| ABDOMEN \| Abdomen INSPECTION — count, first 24 hours |
| Baseline + procedure burden | 3 | procedure_725068_presence_24h | Radiologic examination, chest; single view — presence indicator, first 24 hours |
| Baseline + procedure burden | 3 | procedure_2313816_count24h | Routine 12-lead ECG; interpretation and report only — count, first 24 hours |
| Baseline + procedure burden | 3 | procedure_2514441_presence_24h | Critical care evaluation and management; first 30–74 minutes — presence indicator, first 24 hours |
| Baseline + procedure burden | 3 | procedure_2514442_count24h | Critical care evaluation and management; each additional 30 minutes — count, first 24 hours |
| Baseline + procedure burden | 3 | procedure_2147483056_count24h | ASSISTANCE \| LEVEL OF ASSISTANCE \| LEVEL OF ASSISTANCE — count, first 24 hours |
| Baseline + procedure burden | 3 | procedure_2147483056_presence_24h | ASSISTANCE \| LEVEL OF ASSISTANCE \| LEVEL OF ASSISTANCE — presence indicator, first 24 hours |
| Baseline + procedure burden | 3 | procedure_2147483590_count24h | HEENT \| Eyes/Vision \| Right Eye Assessment — count, first 24 hours |
| Baseline + procedure burden | 3 | procedure_2147483590_presence_24h | HEENT \| Eyes/Vision \| Right Eye Assessment — presence indicator, first 24 hours |
| Baseline + procedure burden | 3 | procedure_2147483603_presence_24h | HEENT \| Eyes/Vision \| Left Eye Assessment — presence indicator, first 24 hours |
| Baseline + procedure burden | 4 | unique_procedure_count_24h | Unique procedure count, first 24 hours |
| Baseline + procedure burden | 4 | procedure_count_total_24h | Total procedure count, first 24 hours |
| Baseline + procedure burden | 4 | procedure_725068_count24h | Radiologic examination, chest; single view — count, first 24 hours |
| Baseline + procedure burden | 4 | procedure_2147482991_presence_24h | LINE CARE \| LINE STATUS \| LINE STATUS — presence indicator, first 24 hours |
| Baseline + procedure burden | 4 | procedure_2147482992_count24h | LINE CARE \| LINE SITE ASSESSMENT \| LINE SITE ASSESSMENT — count, first 24 hours |
| Baseline + procedure burden | 4 | procedure_2147482992_presence_24h | LINE CARE \| LINE SITE ASSESSMENT \| LINE SITE ASSESSMENT — presence indicator, first 24 hours |
| Baseline + procedure burden | 4 | procedure_2313814_presence_24h | Routine 12-lead ECG; with interpretation and report — presence indicator, first 24 hours |
| Baseline + procedure burden | 4 | procedure_2147482991_count24h | LINE CARE \| LINE STATUS \| LINE STATUS — count, first 24 hours |
| Baseline + procedure burden | 4 | procedure_2313814_count24h | Routine 12-lead ECG; with interpretation and report — count, first 24 hours |

Continued on next page

eTable 33. CHoRUS features selected per fold. (continued)

| Feature Matrix | Fold | Feature | Concept name |
|---|---|---|---|
| Baseline + procedure burden | 4 | procedure_2514441_count24h | Critical care evaluation and management; first 30–74 minutes — count, first 24 hours |
| Baseline + procedure burden | 4 | procedure_2147483387_presence_24h | GASTROINTESTINAL \| ABDOMEN \| Abdomen INSPECTION — presence indicator, first 24 hours |
| Baseline + procedure burden | 4 | procedure_2147483387_count24h | GASTROINTESTINAL \| ABDOMEN \| Abdomen INSPECTION — count, first 24 hours |
| Baseline + procedure burden | 4 | procedure_2313816_count24h | Routine 12-lead ECG; interpretation and report only — count, first 24 hours |
| Baseline + procedure burden | 4 | procedure_725068_presence_24h | Radiologic examination, chest; single view — presence indicator, first 24 hours |
| Baseline + procedure burden | 4 | procedure_2147483056_count24h | ASSISTANCE \| LEVEL OF ASSISTANCE \| LEVEL OF ASSISTANCE — count, first 24 hours |
| Baseline + procedure burden | 4 | procedure_2514441_presence_24h | Critical care evaluation and management; first 30–74 minutes — presence indicator, first 24 hours |
| Baseline + procedure burden | 4 | procedure_36713195_count24h | X-ray of chest anteroposterior view — count, first 24 hours |
| Baseline + procedure burden | 4 | procedure_2147483056_presence_24h | ASSISTANCE \| LEVEL OF ASSISTANCE \| LEVEL OF ASSISTANCE — presence indicator, first 24 hours |
| Baseline + procedure burden | 4 | procedure_2147482961_presence_24h | Surgical Airway \| Airway \| Airway Secured By — presence indicator, first 24 hours |
| Baseline + procedure burden | 4 | procedure_2313816_presence_24h | Routine 12-lead ECG; interpretation and report only — presence indicator, first 24 hours |
| Baseline + procedure burden | 4 | procedure_2147483603_presence_24h | HEENT \| Eyes/Vision \| Left Eye Assessment — presence indicator, first 24 hours |
| Baseline + procedure burden | 5 | unique_procedure_count_24h | Unique procedure count, first 24 hours |
| Baseline + procedure burden | 5 | procedure_count_total_24h | Total procedure count, first 24 hours |
| Baseline + procedure burden | 5 | procedure_2147482992_count24h | LINE CARE \| LINE SITE ASSESSMENT \| LINE SITE ASSESSMENT — count, first 24 hours |
| Baseline + procedure burden | 5 | procedure_725068_count24h | Radiologic examination, chest; single view — count, first 24 hours |
| Baseline + procedure burden | 5 | procedure_2147482991_presence_24h | LINE CARE \| LINE STATUS \| LINE STATUS — presence indicator, first 24 hours |
| Baseline + procedure burden | 5 | procedure_2313814_count24h | Routine 12-lead ECG; with interpretation and report — count, first 24 hours |
| Baseline + procedure burden | 5 | procedure_2313814_presence_24h | Routine 12-lead ECG; with interpretation and report — presence indicator, first 24 hours |
| Baseline + procedure burden | 5 | procedure_2147482991_count24h | LINE CARE \| LINE STATUS \| LINE STATUS — count, first 24 hours |
| Baseline + procedure burden | 5 | procedure_2147482992_presence_24h | LINE CARE \| LINE SITE ASSESSMENT \| LINE SITE ASSESSMENT — presence indicator, first 24 hours |
| Baseline + procedure burden | 5 | procedure_2147483387_presence_24h | GASTROINTESTINAL \| ABDOMEN \| Abdomen INSPECTION — presence indicator, first 24 hours |
| Baseline + procedure burden | 5 | procedure_2514441_count24h | Critical care evaluation and management; first 30–74 minutes — count, first 24 hours |
| Baseline + procedure burden | 5 | procedure_2313816_count24h | Routine 12-lead ECG; interpretation and report only — count, first 24 hours |
| Baseline + procedure burden | 5 | procedure_2147483387_count24h | GASTROINTESTINAL \| ABDOMEN \| Abdomen INSPECTION — count, first 24 hours |

Continued on next page

eTable 33. CHoRUS features selected per fold. (continued)

| Feature Matrix | Fold | Feature | Concept name |
|---|---|---|---|
| Baseline + procedure burden | 5 | procedure_2514441_presence_24h | Critical care evaluation and management; first 30–74 minutes — presence indicator, first 24 hours |
| Baseline + procedure burden | 5 | procedure_725068_presence_24h | Radiologic examination, chest; single view — presence indicator, first 24 hours |
| Baseline + procedure burden | 5 | procedure_2313816_presence_24h | Routine 12-lead ECG; interpretation and report only — presence indicator, first 24 hours |
| Baseline + procedure burden | 5 | procedure_2147483590_presence_24h | HEENT \| Eyes/Vision \| Right Eye Assessment — presence indicator, first 24 hours |
| Baseline + procedure burden | 5 | procedure_2147483056_presence_24h | ASSISTANCE \| LEVEL OF ASSISTANCE \| LEVEL OF ASSISTANCE — presence indicator, first 24 hours |
| Baseline + procedure burden | 5 | procedure_2147483603_presence_24h | HEENT \| Eyes/Vision \| Left Eye Assessment — presence indicator, first 24 hours |
| Baseline + procedure burden | 5 | procedure_2147482962_presence_24h | Surgical Airway \| Airway \| Airway Measured From — presence indicator, first 24 hours |
| Baseline + procedure burden | 5 | procedure_2147483056_count24h | ASSISTANCE \| LEVEL OF ASSISTANCE \| LEVEL OF ASSISTANCE — count, first 24 hours |

*Notes:* The source workbook structure, fold order, and feature order within each fold are preserved. Blank source cells are shown as blank.

eTable 34. MIMIC-IV standalone features selected per fold.

| Feature matrix | Fold | Feature | Concept name |
|---|---|---|---|
| Baseline | 1 | age_at_visit | Age at visit |
| Baseline | 1 | sex | Sex |
| Baseline | 1 | race | Race |
| Baseline | 1 | ethnicity | Ethnicity |
| Baseline | 1 | visit_type | Visit type |
| Baseline | 1 | has_prior_visit | Has prior visit |
| Baseline | 1 | prior_visit_count | Prior visit count |
| Baseline | 1 | prior_acute_visit_count | Prior acute-care visit count |
| Baseline | 1 | charlson_index | Charlson Comorbidity Index |
| Baseline | 2 | age_at_visit | Age at visit |
| Baseline | 2 | sex | Sex |
| Baseline | 2 | race | Race |
| Baseline | 2 | ethnicity | Ethnicity |
| Baseline | 2 | visit_type | Visit type |
| Baseline | 2 | has_prior_visit | Has prior visit |
| Baseline | 2 | prior_visit_count | Prior visit count |
| Baseline | 2 | prior_acute_visit_count | Prior acute-care visit count |
| Baseline | 2 | charlson_index | Charlson Comorbidity Index |
| Baseline | 3 | age_at_visit | Age at visit |
| Baseline | 3 | sex | Sex |
| Baseline | 3 | race | Race |
| Baseline | 3 | ethnicity | Ethnicity |
| Baseline | 3 | visit_type | Visit type |
| Baseline | 3 | has_prior_visit | Has prior visit |
| Baseline | 3 | prior_visit_count | Prior visit count |
| Baseline | 3 | prior_acute_visit_count | Prior acute-care visit count |

Continued on next page

eTable 34. MIMIC-IV standalone features selected per fold. (continued)

| Feature matrix | Fold | Feature | Concept name |
|---|---|---|---|
| Baseline | 3 | charlson_index | Charlson Comorbidity Index |
| Baseline | 4 | age_at_visit | Age at visit |
| Baseline | 4 | sex | Sex |
| Baseline | 4 | race | Race |
| Baseline | 4 | ethnicity | Ethnicity |
| Baseline | 4 | visit_type | Visit type |
| Baseline | 4 | has_prior_visit | Has prior visit |
| Baseline | 4 | prior_visit_count | Prior visit count |
| Baseline | 4 | prior_acute_visit_count | Prior acute-care visit count |
| Baseline | 4 | charlson_index | Charlson Comorbidity Index |
| Baseline | 5 | age_at_visit | Age at visit |
| Baseline | 5 | sex | Sex |
| Baseline | 5 | race | Race |
| Baseline | 5 | ethnicity | Ethnicity |
| Baseline | 5 | visit_type | Visit type |
| Baseline | 5 | has_prior_visit | Has prior visit |
| Baseline | 5 | prior_visit_count | Prior visit count |
| Baseline | 5 | prior_acute_visit_count | Prior acute-care visit count |
| Baseline | 5 | charlson_index | Charlson Comorbidity Index |
| Baseline + physiological severity | 1 | lab_50882_bicarbonate_mean | Bicarbonate - mean, first 24 hours |
| Baseline + physiological severity | 1 | lab_50813_lactate_max | Lactate - maximum, first 24 hours |
| Baseline + physiological severity | 1 | lab_51277_rdw_max | RDW - maximum, first 24 hours |
| Baseline + physiological severity | 1 | lab_50818_pco2_std | pCO2 - standard deviation, first 24 hours |
| Baseline + physiological severity | 1 | lab_50813_lactate_min | Lactate - minimum, first 24 hours |
| Baseline + physiological severity | 1 | lab_50818_pco2_min | pCO2 - minimum, first 24 hours |
| Baseline + physiological severity | 1 | lab_50820_ph_mean | pH - mean, first 24 hours |
| Baseline + physiological severity | 1 | lab_51006_urea_nitrogen_max | Urea Nitrogen - maximum, first 24 hours |
| Baseline + physiological severity | 1 | lab_50820_ph_min | pH - minimum, first 24 hours |
| Baseline + physiological severity | 1 | lab_50820_ph_max | pH - maximum, first 24 hours |
| Baseline + physiological severity | 1 | lab_50893_calcium_total_mean | Calcium, Total - mean, first 24 hours |
| Baseline + physiological severity | 1 | lab_50820_ph_std | pH - standard deviation, first 24 hours |
| Baseline + physiological severity | 1 | lab_50804_calculated_total_co2_mean | Calculated Total CO2 - mean, first 24 hours |
| Baseline + physiological severity | 1 | lab_50983_sodium_min | Sodium - minimum, first 24 hours |
| Baseline + physiological severity | 1 | lab_50893_calcium_total_min | Calcium, Total - minimum, first 24 hours |
| Baseline + physiological severity | 1 | lab_50983_sodium_mean | Sodium - mean, first 24 hours |
| Baseline + physiological severity | 1 | lab_50983_sodium_count | Sodium - count, first 24 hours |
| Baseline + physiological severity | 1 | lab_51277_rdw_min | RDW - minimum, first 24 hours |
| Baseline + physiological severity | 1 | lab_50912_creatinine_mean | Creatinine - mean, first 24 hours |
| Baseline + physiological severity | 1 | lab_51277_rdw_mean | RDW - mean, first 24 hours |
| Baseline + physiological severity | 1 | lab_50818_pco2_mean | pCO2 - mean, first 24 hours |
| Baseline + physiological severity | 2 | lab_50820_ph_mean | pH - mean, first 24 hours |
| Baseline + physiological severity | 2 | lab_50804_calculated_total_co2_std | Calculated Total CO2 - standard deviation, first 24 hours |
| Baseline + physiological severity | 2 | lab_50983_sodium_max | Sodium - maximum, first 24 hours |
| Baseline + physiological severity | 2 | lab_50983_sodium_mean | Sodium - mean, first 24 hours |
| Baseline + physiological severity | 2 | lab_51301_white_blood_cells_max | White Blood Cells - maximum, first 24 hours |
| Baseline + physiological severity | 2 | lab_50818_pco2_std | pCO2 - standard deviation, first 24 hours |

Continued on next page

eTable 34. MIMIC-IV standalone features selected per fold. (continued)

| Feature matrix | Fold | Feature | Concept name |
|---|---|---|---|
| Baseline + physiological severity | 2 | lab_51006_urea_nitrogen_mean | Urea Nitrogen - mean, first 24 hours |
| Baseline + physiological severity | 2 | lab_51006_urea_nitrogen_min | Urea Nitrogen - minimum, first 24 hours |
| Baseline + physiological severity | 2 | lab_51277_rdw_max | RDW - maximum, first 24 hours |
| Baseline + physiological severity | 2 | lab_50820_ph_max | pH - maximum, first 24 hours |
| Baseline + physiological severity | 2 | lab_50804_calculated_total_co2_mean | Calculated Total CO2 - mean, first 24 hours |
| Baseline + physiological severity | 2 | lab_50821_po2_mean | pO2 - mean, first 24 hours |
| Baseline + physiological severity | 2 | lab_50820_ph_min | pH - minimum, first 24 hours |
| Baseline + physiological severity | 2 | lab_50983_sodium_count | Sodium - count, first 24 hours |
| Baseline + physiological severity | 2 | lab_50813_lactate_min | Lactate - minimum, first 24 hours |
| Baseline + physiological severity | 2 | lab_50893_calcium_total_mean | Calcium, Total - mean, first 24 hours |
| Baseline + physiological severity | 2 | lab_51274_pt_mean | PT - mean, first 24 hours |
| Baseline + physiological severity | 2 | lab_50983_sodium_std | Sodium - standard deviation, first 24 hours |
| Baseline + physiological severity | 2 | lab_50813_lactate_mean | Lactate - mean, first 24 hours |
| Baseline + physiological severity | 2 | lab_50818_pco2_max | pCO2 - maximum, first 24 hours |
| Baseline + physiological severity | 2 | lab_50882_bicarbonate_mean | Bicarbonate - mean, first 24 hours |
| Baseline + physiological severity | 3 | lab_51274_pt_mean | PT - mean, first 24 hours |
| Baseline + physiological severity | 3 | lab_51277_rdw_max | RDW - maximum, first 24 hours |
| Baseline + physiological severity | 3 | lab_50821_po2_max | pO2 - maximum, first 24 hours |
| Baseline + physiological severity | 3 | lab_50983_sodium_mean | Sodium - mean, first 24 hours |
| Baseline + physiological severity | 3 | lab_50821_po2_min | pO2 - minimum, first 24 hours |
| Baseline + physiological severity | 3 | lab_50820_ph_min | pH - minimum, first 24 hours |
| Baseline + physiological severity | 3 | lab_50820_ph_max | pH - maximum, first 24 hours |
| Baseline + physiological severity | 3 | lab_50821_po2_std | pO2 - standard deviation, first 24 hours |
| Baseline + physiological severity | 3 | lab_51237_inr_pt_mean | INR(PT) - mean, first 24 hours |
| Baseline + physiological severity | 3 | lab_50820_ph_mean | pH - mean, first 24 hours |
| Baseline + physiological severity | 3 | lab_50813_lactate_min | Lactate - minimum, first 24 hours |
| Baseline + physiological severity | 3 | lab_50813_lactate_max | Lactate - maximum, first 24 hours |
| Baseline + physiological severity | 3 | lab_50818_pco2_std | pCO2 - standard deviation, first 24 hours |
| Baseline + physiological severity | 3 | lab_51237_inr_pt_max | INR(PT) - maximum, first 24 hours |
| Baseline + physiological severity | 3 | lab_50954_lactate_dehydrogenase_ld_min | Lactate Dehydrogenase (LD) - minimum, first 24 hours |
| Baseline + physiological severity | 3 | lab_50882_bicarbonate_std | Bicarbonate - standard deviation, first 24 hours |
| Baseline + physiological severity | 3 | lab_50818_pco2_max | pCO2 - maximum, first 24 hours |
| Baseline + physiological severity | 3 | lab_50893_calcium_total_max | Calcium, Total - maximum, first 24 hours |
| Baseline + physiological severity | 3 | lab_50983_sodium_count | Sodium - count, first 24 hours |
| Baseline + physiological severity | 3 | lab_51277_rdw_mean | RDW - mean, first 24 hours |
| Baseline + physiological severity | 3 | lab_51006_urea_nitrogen_mean | Urea Nitrogen - mean, first 24 hours |
| Baseline + physiological severity | 4 | lab_51277_rdw_min | RDW - minimum, first 24 hours |
| Baseline + physiological severity | 4 | lab_50820_ph_mean | pH - mean, first 24 hours |
| Baseline + physiological severity | 4 | lab_51006_urea_nitrogen_mean | Urea Nitrogen - mean, first 24 hours |
| Baseline + physiological severity | 4 | lab_50818_pco2_std | pCO2 - standard deviation, first 24 hours |
| Baseline + physiological severity | 4 | lab_50813_lactate_min | Lactate - minimum, first 24 hours |
| Baseline + physiological severity | 4 | lab_50813_lactate_max | Lactate - maximum, first 24 hours |
| Baseline + physiological severity | 4 | lab_50821_po2_mean | pO2 - mean, first 24 hours |
| Baseline + physiological severity | 4 | lab_51274_pt_max | PT - maximum, first 24 hours |
| Baseline + physiological severity | 4 | lab_50821_po2_min | pO2 - minimum, first 24 hours |
| Baseline + physiological severity | 4 | lab_50983_sodium_mean | Sodium - mean, first 24 hours |

Continued on next page

eTable 34. MIMIC-IV standalone features selected per fold. (continued)

| Feature matrix | Fold | Feature | Concept name |
|---|---|---|---|
| Baseline + physiological severity | 4 | lab_51277_rdw_mean | RDW - mean, first 24 hours |
| Baseline + physiological severity | 4 | lab_50983_sodium_max | Sodium - maximum, first 24 hours |
| Baseline + physiological severity | 4 | lab_50820_ph_max | pH - maximum, first 24 hours |
| Baseline + physiological severity | 4 | lab_50983_sodium_count | Sodium - count, first 24 hours |
| Baseline + physiological severity | 4 | lab_50821_po2_max | pO2 - maximum, first 24 hours |
| Baseline + physiological severity | 4 | lab_51301_white_blood_cells_max | White Blood Cells - maximum, first 24 hours |
| Baseline + physiological severity | 4 | lab_51006_urea_nitrogen_max | Urea Nitrogen - maximum, first 24 hours |
| Baseline + physiological severity | 4 | lab_51237_inr_pt_mean | INR(PT) - mean, first 24 hours |
| Baseline + physiological severity | 4 | lab_50983_sodium_std | Sodium - standard deviation, first 24 hours |
| Baseline + physiological severity | 4 | lab_50804_calculated_total_co2_mean | Calculated Total CO2 - mean, first 24 hours |
| Baseline + physiological severity | 4 | lab_50818_pco2_max | pCO2 - maximum, first 24 hours |
| Baseline + physiological severity | 5 | lab_51277_rdw_max | RDW - maximum, first 24 hours |
| Baseline + physiological severity | 5 | lab_51006_urea_nitrogen_mean | Urea Nitrogen - mean, first 24 hours |
| Baseline + physiological severity | 5 | lab_50983_sodium_mean | Sodium - mean, first 24 hours |
| Baseline + physiological severity | 5 | lab_50820_ph_min | pH - minimum, first 24 hours |
| Baseline + physiological severity | 5 | lab_50813_lactate_min | Lactate - minimum, first 24 hours |
| Baseline + physiological severity | 5 | lab_51222_hemoglobin_max | Hemoglobin - maximum, first 24 hours |
| Baseline + physiological severity | 5 | lab_51277_rdw_min | RDW - minimum, first 24 hours |
| Baseline + physiological severity | 5 | lab_50820_ph_mean | pH - mean, first 24 hours |
| Baseline + physiological severity | 5 | lab_50804_calculated_total_co2_mean | Calculated Total CO2 - mean, first 24 hours |
| Baseline + physiological severity | 5 | lab_50983_sodium_count | Sodium - count, first 24 hours |
| Baseline + physiological severity | 5 | lab_51277_rdw_mean | RDW - mean, first 24 hours |
| Baseline + physiological severity | 5 | lab_50821_po2_max | pO2 - maximum, first 24 hours |
| Baseline + physiological severity | 5 | lab_50818_pco2_min | pCO2 - minimum, first 24 hours |
| Baseline + physiological severity | 5 | lab_51006_urea_nitrogen_max | Urea Nitrogen - maximum, first 24 hours |
| Baseline + physiological severity | 5 | lab_50893_calcium_total_count | Calcium, Total - count, first 24 hours |
| Baseline + physiological severity | 5 | lab_50818_pco2_std | pCO2 - standard deviation, first 24 hours |
| Baseline + physiological severity | 5 | lab_50970_phosphate_count | Phosphate - count, first 24 hours |
| Baseline + physiological severity | 5 | lab_50902_chloride_count | Chloride - count, first 24 hours |
| Baseline + physiological severity | 5 | lab_50971_potassium_count | Potassium - count, first 24 hours |
| Baseline + physiological severity | 5 | lab_50931_glucose_count | Glucose - count, first 24 hours |
| Baseline + physiological severity | 5 | lab_50813_lactate_max | Lactate - maximum, first 24 hours |
| Baseline + treatment exposure | 1 | unique_drug_count_24h | Unique drug count, first 24 hours |
| Baseline + treatment exposure | 1 | repeat_drug_exposure_count_24h | Repeat drug exposure count, first 24 hours |
| Baseline + treatment exposure | 1 | drug_gsn_001210_ba3dc43c8d_count_24h | 0.9% Sodium Chloride - count, first 24 hours |
| Baseline + treatment exposure | 1 | drug_gsn_001210_ba3dc43c8d_exposed_24h | 0.9% Sodium Chloride - exposure indicator, first 24 hours |
| Baseline + treatment exposure | 1 | time_to_first_drug_hours | Time to first drug exposure, hours |
| Baseline + treatment exposure | 1 | drug_gsn_001972_b5f0a0eff8_count_24h | 5% Dextrose - count, first 24 hours |
| Baseline + treatment exposure | 1 | any_drug_24h | Any drug exposure, first 24 hours |
| Baseline + treatment exposure | 1 | drug_formulary_vancobase_228466e611_count_24h | Iso-Osmotic Dextrose - count, first 24 hours |
| Baseline + treatment exposure | 1 | drug_gsn_043952_0602de6ed2_count_24h | Vancomycin - count, first 24 hours |
| Baseline + treatment exposure | 1 | drug_formulary_fentsoln50_7775dcade8_count_24h | Soln - count, first 24 hours |
| Baseline + treatment exposure | 1 | drug_formulary_vancobase_228466e611_exposed_24h | Iso-Osmotic Dextrose - exposure indicator, first 24 hours |
| Baseline + treatment exposure | 1 | drug_gsn_043952_0602de6ed2_exposed_24h | Vancomycin - exposure indicator, first 24 hours |
| Baseline + treatment exposure | 1 | drug_gsn_048287_21c090e9e3_count_24h | Fentanyl Citrate - count, first 24 hours |
| Baseline + treatment exposure | 1 | drug_formulary_fentsoln50_7775dcade8_exposed_24h | Soln - exposure indicator, first 24 hours |

Continued on next page

eTable 34. MIMIC-IV standalone features selected per fold. (continued)

| Feature matrix | Fold | Feature | Concept name |
|---|---|---|---|
| Baseline + treatment exposure | 1 | drug_gsn_001972_b5f0a0eff8_exposed_24h | 5% Dextrose - exposure indicator, first 24 hours |
| Baseline + treatment exposure | 1 | drug_gsn_057959_a518f33e58_count_24h | Chlorhexidine Gluconate 0.12% Oral Rinse - count, first 24 hours |
| Baseline + treatment exposure | 1 | drug_gsn_048287_21c090e9e3_exposed_24h | Fentanyl Citrate - exposure indicator, first 24 hours |
| Baseline + treatment exposure | 1 | drug_gsn_057959_a518f33e58_exposed_24h | Chlorhexidine Gluconate 0.12% Oral Rinse - exposure indicator, first 24 hours |
| Baseline + treatment exposure | 1 | drug_gsn_016796_f69d8264fe_count_24h | Propofol - count, first 24 hours |
| Baseline + treatment exposure | 1 | drug_gsn_016796_f69d8264fe_exposed_24h | Propofol - exposure indicator, first 24 hours |
| Baseline + treatment exposure | 1 | drug_gsn_060304_4b8ed8c2a4_count_24h | Heparin Flush (10 units/ml) - count, first 24 hours |
| Baseline + treatment exposure | 2 | unique_drug_count_24h | Unique drug count, first 24 hours |
| Baseline + treatment exposure | 2 | repeat_drug_exposure_count_24h | Repeat drug exposure count, first 24 hours |
| Baseline + treatment exposure | 2 | drug_gsn_001210_ba3dc43c8d_count_24h | 0.9% Sodium Chloride - count, first 24 hours |
| Baseline + treatment exposure | 2 | drug_gsn_001972_b5f0a0eff8_count_24h | 5% Dextrose - count, first 24 hours |
| Baseline + treatment exposure | 2 | drug_gsn_001210_ba3dc43c8d_exposed_24h | 0.9% Sodium Chloride - exposure indicator, first 24 hours |
| Baseline + treatment exposure | 2 | drug_gsn_048287_21c090e9e3_count_24h | Fentanyl Citrate - count, first 24 hours |
| Baseline + treatment exposure | 2 | drug_gsn_048287_21c090e9e3_exposed_24h | Fentanyl Citrate - exposure indicator, first 24 hours |
| Baseline + treatment exposure | 2 | any_drug_24h | Any drug exposure, first 24 hours |
| Baseline + treatment exposure | 2 | time_to_first_drug_hours | Time to first drug exposure, hours |
| Baseline + treatment exposure | 2 | drug_formulary_fentsoln50_7775dcade8_count_24h | Soln - count, first 24 hours |
| Baseline + treatment exposure | 2 | drug_formulary_vancobase_228466e611_count_24h | Iso-Osmotic Dextrose - count, first 24 hours |
| Baseline + treatment exposure | 2 | drug_gsn_043952_0602de6ed2_count_24h | Vancomycin - count, first 24 hours |
| Baseline + treatment exposure | 2 | drug_gsn_057959_a518f33e58_count_24h | Chlorhexidine Gluconate 0.12% Oral Rinse - count, first 24 hours |
| Baseline + treatment exposure | 2 | drug_formulary_fentsoln50_7775dcade8_exposed_24h | Soln - exposure indicator, first 24 hours |
| Baseline + treatment exposure | 2 | drug_gsn_016796_f69d8264fe_count_24h | Propofol - count, first 24 hours |
| Baseline + treatment exposure | 2 | drug_gsn_016796_f69d8264fe_exposed_24h | Propofol - exposure indicator, first 24 hours |
| Baseline + treatment exposure | 2 | drug_formulary_vancobase_228466e611_exposed_24h | Iso-Osmotic Dextrose - exposure indicator, first 24 hours |
| Baseline + treatment exposure | 2 | drug_gsn_043952_0602de6ed2_exposed_24h | Vancomycin - exposure indicator, first 24 hours |
| Baseline + treatment exposure | 2 | drug_gsn_057959_a518f33e58_exposed_24h | Chlorhexidine Gluconate 0.12% Oral Rinse - exposure indicator, first 24 hours |
| Baseline + treatment exposure | 2 | drug_formulary_vial_b4c8f642b1_count_24h | Vial - count, first 24 hours |
| Baseline + treatment exposure | 2 | drug_gsn_001972_b5f0a0eff8_exposed_24h | 5% Dextrose - exposure indicator, first 24 hours |
| Baseline + treatment exposure | 3 | unique_drug_count_24h | Unique drug count, first 24 hours |
| Baseline + treatment exposure | 3 | repeat_drug_exposure_count_24h | Repeat drug exposure count, first 24 hours |
| Baseline + treatment exposure | 3 | drug_gsn_001210_ba3dc43c8d_count_24h | 0.9% Sodium Chloride - count, first 24 hours |
| Baseline + treatment exposure | 3 | drug_gsn_001210_ba3dc43c8d_exposed_24h | 0.9% Sodium Chloride - exposure indicator, first 24 hours |
| Baseline + treatment exposure | 3 | drug_gsn_001972_b5f0a0eff8_count_24h | 5% Dextrose - count, first 24 hours |
| Baseline + treatment exposure | 3 | any_drug_24h | Any drug exposure, first 24 hours |
| Baseline + treatment exposure | 3 | drug_gsn_048287_21c090e9e3_count_24h | Fentanyl Citrate - count, first 24 hours |
| Baseline + treatment exposure | 3 | drug_formulary_vancobase_228466e611_count_24h | Iso-Osmotic Dextrose - count, first 24 hours |
| Baseline + treatment exposure | 3 | drug_gsn_043952_0602de6ed2_count_24h | Vancomycin - count, first 24 hours |
| Baseline + treatment exposure | 3 | drug_formulary_vancobase_228466e611_exposed_24h | Iso-Osmotic Dextrose - exposure indicator, first 24 hours |
| Baseline + treatment exposure | 3 | drug_gsn_043952_0602de6ed2_exposed_24h | Vancomycin - exposure indicator, first 24 hours |
| Baseline + treatment exposure | 3 | drug_formulary_fentsoln50_7775dcade8_count_24h | Soln - count, first 24 hours |
| Baseline + treatment exposure | 3 | drug_gsn_048287_21c090e9e3_exposed_24h | Fentanyl Citrate - exposure indicator, first 24 hours |
| Baseline + treatment exposure | 3 | time_to_first_drug_hours | Time to first drug exposure, hours |
| Baseline + treatment exposure | 3 | drug_formulary_fentsoln50_7775dcade8_exposed_24h | Soln - exposure indicator, first 24 hours |
| Baseline + treatment exposure | 3 | drug_gsn_016796_f69d8264fe_count_24h | Propofol - count, first 24 hours |
| Baseline + treatment exposure | 3 | drug_gsn_057959_a518f33e58_count_24h | Chlorhexidine Gluconate 0.12% Oral Rinse - count, first 24 hours |

Continued on next page

eTable 34. MIMIC-IV standalone features selected per fold. (continued)

| Feature matrix | Fold | Feature | Concept name |
|---|---|---|---|
| Baseline + treatment exposure | 3 | drug_gsn_016796_f69d8264fe_exposed_24h | Propofol - exposure indicator, first 24 hours |
| Baseline + treatment exposure | 3 | drug_formulary_vial_b4c8f642b1_count_24h | Vial - count, first 24 hours |
| Baseline + treatment exposure | 3 | drug_gsn_001972_b5f0a0eff8_exposed_24h | 5% Dextrose - exposure indicator, first 24 hours |
| Baseline + treatment exposure | 3 | drug_gsn_057959_a518f33e58_exposed_24h | Chlorhexidine Gluconate 0.12% Oral Rinse - exposure indicator, first 24 hours |
| Baseline + treatment exposure | 4 | repeat_drug_exposure_count_24h | Repeat drug exposure count, first 24 hours |
| Baseline + treatment exposure | 4 | unique_drug_count_24h | Unique drug count, first 24 hours |
| Baseline + treatment exposure | 4 | drug_gsn_001210_ba3dc43c8d_count_24h | 0.9% Sodium Chloride - count, first 24 hours |
| Baseline + treatment exposure | 4 | drug_gsn_001210_ba3dc43c8d_exposed_24h | 0.9% Sodium Chloride - exposure indicator, first 24 hours |
| Baseline + treatment exposure | 4 | drug_gsn_001972_b5f0a0eff8_count_24h | 5% Dextrose - count, first 24 hours |
| Baseline + treatment exposure | 4 | time_to_first_drug_hours | Time to first drug exposure, hours |
| Baseline + treatment exposure | 4 | drug_formulary_vancobase_228466e611_count_24h | Iso-Osmotic Dextrose - count, first 24 hours |
| Baseline + treatment exposure | 4 | drug_gsn_043952_0602de6ed2_count_24h | Vancomycin - count, first 24 hours |
| Baseline + treatment exposure | 4 | drug_gsn_001972_b5f0a0eff8_exposed_24h | 5% Dextrose - exposure indicator, first 24 hours |
| Baseline + treatment exposure | 4 | drug_formulary_vancobase_228466e611_exposed_24h | Iso-Osmotic Dextrose - exposure indicator, first 24 hours |
| Baseline + treatment exposure | 4 | drug_gsn_043952_0602de6ed2_exposed_24h | Vancomycin - exposure indicator, first 24 hours |
| Baseline + treatment exposure | 4 | any_drug_24h | Any drug exposure, first 24 hours |
| Baseline + treatment exposure | 4 | drug_formulary_fentsoln50_7775dcade8_count_24h | Soln - count, first 24 hours |
| Baseline + treatment exposure | 4 | drug_gsn_016796_f69d8264fe_count_24h | Propofol - count, first 24 hours |
| Baseline + treatment exposure | 4 | drug_gsn_048287_21c090e9e3_count_24h | Fentanyl Citrate - count, first 24 hours |
| Baseline + treatment exposure | 4 | drug_gsn_016796_f69d8264fe_exposed_24h | Propofol - exposure indicator, first 24 hours |
| Baseline + treatment exposure | 4 | drug_formulary_fentsoln50_7775dcade8_exposed_24h | Soln - exposure indicator, first 24 hours |
| Baseline + treatment exposure | 4 | drug_formulary_vial_b4c8f642b1_count_24h | Vial - count, first 24 hours |
| Baseline + treatment exposure | 4 | drug_gsn_048287_21c090e9e3_exposed_24h | Fentanyl Citrate - exposure indicator, first 24 hours |
| Baseline + treatment exposure | 4 | drug_formulary_norebasens_c31a0d47cc_count_24h | 0.9% Sodium Chloride - count, first 24 hours |
| Baseline + treatment exposure | 4 | drug_gsn_066419_c17fadb0e8_count_24h | NORepinephrine - count, first 24 hours |
| Baseline + treatment exposure | 5 | repeat_drug_exposure_count_24h | Repeat drug exposure count, first 24 hours |
| Baseline + treatment exposure | 5 | unique_drug_count_24h | Unique drug count, first 24 hours |
| Baseline + treatment exposure | 5 | drug_gsn_001210_ba3dc43c8d_count_24h | 0.9% Sodium Chloride - count, first 24 hours |
| Baseline + treatment exposure | 5 | drug_gsn_001210_ba3dc43c8d_exposed_24h | 0.9% Sodium Chloride - exposure indicator, first 24 hours |
| Baseline + treatment exposure | 5 | drug_gsn_001972_b5f0a0eff8_count_24h | 5% Dextrose - count, first 24 hours |
| Baseline + treatment exposure | 5 | drug_gsn_001972_b5f0a0eff8_exposed_24h | 5% Dextrose - exposure indicator, first 24 hours |
| Baseline + treatment exposure | 5 | any_drug_24h | Any drug exposure, first 24 hours |
| Baseline + treatment exposure | 5 | drug_formulary_vancobase_228466e611_count_24h | Iso-Osmotic Dextrose - count, first 24 hours |
| Baseline + treatment exposure | 5 | drug_gsn_043952_0602de6ed2_count_24h | Vancomycin - count, first 24 hours |
| Baseline + treatment exposure | 5 | drug_formulary_fentsoln50_7775dcade8_count_24h | Soln - count, first 24 hours |
| Baseline + treatment exposure | 5 | drug_formulary_vancobase_228466e611_exposed_24h | Iso-Osmotic Dextrose - exposure indicator, first 24 hours |
| Baseline + treatment exposure | 5 | drug_gsn_043952_0602de6ed2_exposed_24h | Vancomycin - exposure indicator, first 24 hours |
| Baseline + treatment exposure | 5 | drug_gsn_048287_21c090e9e3_count_24h | Fentanyl Citrate - count, first 24 hours |
| Baseline + treatment exposure | 5 | drug_gsn_057959_a518f33e58_count_24h | Chlorhexidine Gluconate 0.12% Oral Rinse - count, first 24 hours |
| Baseline + treatment exposure | 5 | drug_formulary_fentsoln50_7775dcade8_exposed_24h | Soln - exposure indicator, first 24 hours |
| Baseline + treatment exposure | 5 | drug_formulary_vial_b4c8f642b1_count_24h | Vial - count, first 24 hours |
| Baseline + treatment exposure | 5 | drug_gsn_048287_21c090e9e3_exposed_24h | Fentanyl Citrate - exposure indicator, first 24 hours |
| Baseline + treatment exposure | 5 | time_to_first_drug_hours | Time to first drug exposure, hours |
| Baseline + treatment exposure | 5 | drug_gsn_057959_a518f33e58_exposed_24h | Chlorhexidine Gluconate 0.12% Oral Rinse - exposure indicator, first 24 hours |
| Baseline + treatment exposure | 5 | drug_formulary_vial_b4c8f642b1_exposed_24h | Vial - exposure indicator, first 24 hours |

Continued on next page

eTable 34. MIMIC-IV standalone features selected per fold. (continued)

| Feature matrix | Fold | Feature | Concept name |
|---|---|---|---|
| Baseline + treatment exposure | 5 | drug_gsn_016796_f69d8264fe_count_24h | Propofol - count, first 24 hours |
| Baseline + procedure burden | 1 | procedure_icd10_5a1955z_6a2b52611f_count_24h | ICD-10-PCS procedure 5A1955Z - count, first 24 hours |
| Baseline + procedure burden | 1 | procedure_icd10_5a1955z_6a2b52611f_presence_24h | ICD-10-PCS procedure 5A1955Z - presence indicator, first 24 hours |
| Baseline + procedure burden | 1 | procedure_icd9_9672_93b639be5d_count_24h | ICD-9 procedure 9672 - count, first 24 hours |
| Baseline + procedure burden | 1 | procedure_icd9_9672_93b639be5d_presence_24h | ICD-9 procedure 9672 - presence indicator, first 24 hours |
| Baseline + procedure burden | 1 | procedure_icd10_5a1945z_f9e7fcf8fd_count_24h | ICD-10-PCS procedure 5A1945Z - count, first 24 hours |
| Baseline + procedure burden | 1 | procedure_icd10_5a1945z_f9e7fcf8fd_presence_24h | ICD-10-PCS procedure 5A1945Z - presence indicator, first 24 hours |
| Baseline + procedure burden | 1 | unique_procedure_count_24h | Unique procedure count, first 24 hours |
| Baseline + procedure burden | 1 | procedure_count_total_24h | Total procedure count, first 24 hours |
| Baseline + procedure burden | 1 | procedure_icd10_3e0g76z_fa1c50513e_count_24h | ICD-10-PCS procedure 3E0G76Z - count, first 24 hours |
| Baseline + procedure burden | 1 | procedure_icd10_3e0g76z_fa1c50513e_presence_24h | ICD-10-PCS procedure 3E0G76Z - presence indicator, first 24 hours |
| Baseline + procedure burden | 1 | procedure_icd10_0bh17ez_3d5ba254ee_count_24h | ICD-10-PCS procedure 0BH17EZ - count, first 24 hours |
| Baseline + procedure burden | 1 | procedure_icd10_0bh17ez_3d5ba254ee_presence_24h | ICD-10-PCS procedure 0BH17EZ - presence indicator, first 24 hours |
| Baseline + procedure burden | 1 | procedure_icd9_3893_49b890a736_count_24h | ICD-9 procedure 3893 - count, first 24 hours |
| Baseline + procedure burden | 1 | procedure_icd9_3893_49b890a736_presence_24h | ICD-9 procedure 3893 - presence indicator, first 24 hours |
| Baseline + procedure burden | 1 | procedure_icd9_9604_93039b475e_count_24h | ICD-9 procedure 9604 - count, first 24 hours |
| Baseline + procedure burden | 1 | procedure_icd9_9604_93039b475e_presence_24h | ICD-9 procedure 9604 - presence indicator, first 24 hours |
| Baseline + procedure burden | 1 | procedure_icd9_9671_5bce41582d_count_24h | ICD-9 procedure 9671 - count, first 24 hours |
| Baseline + procedure burden | 1 | procedure_icd9_9671_5bce41582d_presence_24h | ICD-9 procedure 9671 - presence indicator, first 24 hours |
| Baseline + procedure burden | 1 | procedure_icd10_02hv33z_8e9f454673_count_24h | ICD-10-PCS procedure 02HV33Z - count, first 24 hours |
| Baseline + procedure burden | 1 | procedure_icd10_02hv33z_8e9f454673_presence_24h | ICD-10-PCS procedure 02HV33Z - presence indicator, first 24 hours |
| Baseline + procedure burden | 1 | any_procedure_24h | Any procedure recorded, first 24 hours |
| Baseline + procedure burden | 2 | procedure_icd10_5a1955z_6a2b52611f_count_24h | ICD-10-PCS procedure 5A1955Z - count, first 24 hours |
| Baseline + procedure burden | 2 | procedure_icd10_5a1955z_6a2b52611f_presence_24h | ICD-10-PCS procedure 5A1955Z - presence indicator, first 24 hours |
| Baseline + procedure burden | 2 | procedure_icd10_5a1945z_f9e7fcf8fd_count_24h | ICD-10-PCS procedure 5A1945Z - count, first 24 hours |
| Baseline + procedure burden | 2 | procedure_icd10_5a1945z_f9e7fcf8fd_presence_24h | ICD-10-PCS procedure 5A1945Z - presence indicator, first 24 hours |
| Baseline + procedure burden | 2 | procedure_icd9_9672_93b639be5d_count_24h | ICD-9 procedure 9672 - count, first 24 hours |
| Baseline + procedure burden | 2 | procedure_icd9_9672_93b639be5d_presence_24h | ICD-9 procedure 9672 - presence indicator, first 24 hours |
| Baseline + procedure burden | 2 | procedure_icd10_0bh17ez_3d5ba254ee_count_24h | ICD-10-PCS procedure 0BH17EZ - count, first 24 hours |
| Baseline + procedure burden | 2 | procedure_icd10_0bh17ez_3d5ba254ee_presence_24h | ICD-10-PCS procedure 0BH17EZ - presence indicator, first 24 hours |
| Baseline + procedure burden | 2 | procedure_icd10_3e0g76z_fa1c50513e_count_24h | ICD-10-PCS procedure 3E0G76Z - count, first 24 hours |
| Baseline + procedure burden | 2 | procedure_icd10_3e0g76z_fa1c50513e_presence_24h | ICD-10-PCS procedure 3E0G76Z - presence indicator, first 24 hours |
| Baseline + procedure burden | 2 | unique_procedure_count_24h | Unique procedure count, first 24 hours |
| Baseline + procedure burden | 2 | procedure_count_total_24h | Total procedure count, first 24 hours |
| Baseline + procedure burden | 2 | procedure_icd10_02hv33z_8e9f454673_count_24h | ICD-10-PCS procedure 02HV33Z - count, first 24 hours |
| Baseline + procedure burden | 2 | procedure_icd10_02hv33z_8e9f454673_presence_24h | ICD-10-PCS procedure 02HV33Z - presence indicator, first 24 hours |
| Baseline + procedure burden | 2 | procedure_icd9_3893_49b890a736_count_24h | ICD-9 procedure 3893 - count, first 24 hours |

Continued on next page

eTable 34. MIMIC-IV standalone features selected per fold. (continued)

| Feature matrix | Fold | Feature | Concept name |
|---|---|---|---|
| Baseline + procedure burden | 2 | procedure_icd9_3893_49b890a736_presence_24h | ICD-9 procedure 3893 - presence indicator, first 24 hours |
| Baseline + procedure burden | 2 | any_procedure_24h | Any procedure recorded, first 24 hours |
| Baseline + procedure burden | 2 | procedure_icd9_9604_93039b475e_count_24h | ICD-9 procedure 9604 - count, first 24 hours |
| Baseline + procedure burden | 2 | procedure_icd9_9604_93039b475e_presence_24h | ICD-9 procedure 9604 - presence indicator, first 24 hours |
| Baseline + procedure burden | 2 | procedure_icd9_9671_5bce41582d_count_24h | ICD-9 procedure 9671 - count, first 24 hours |
| Baseline + procedure burden | 2 | procedure_icd9_9671_5bce41582d_presence_24h | ICD-9 procedure 9671 - presence indicator, first 24 hours |
| Baseline + procedure burden | 3 | procedure_icd10_5a1955z_6a2b52611f_count_24h | ICD-10-PCS procedure 5A1955Z - count, first 24 hours |
| Baseline + procedure burden | 3 | procedure_icd10_5a1955z_6a2b52611f_presence_24h | ICD-10-PCS procedure 5A1955Z - presence indicator, first 24 hours |
| Baseline + procedure burden | 3 | procedure_icd9_9672_93b639be5d_count_24h | ICD-9 procedure 9672 - count, first 24 hours |
| Baseline + procedure burden | 3 | procedure_icd9_9672_93b639be5d_presence_24h | ICD-9 procedure 9672 - presence indicator, first 24 hours |
| Baseline + procedure burden | 3 | procedure_icd10_3e0g76z_fa1c50513e_count_24h | ICD-10-PCS procedure 3E0G76Z - count, first 24 hours |
| Baseline + procedure burden | 3 | procedure_icd10_3e0g76z_fa1c50513e_presence_24h | ICD-10-PCS procedure 3E0G76Z - presence indicator, first 24 hours |
| Baseline + procedure burden | 3 | unique_procedure_count_24h | Unique procedure count, first 24 hours |
| Baseline + procedure burden | 3 | procedure_icd10_5a1945z_f9e7fcf8fd_count_24h | ICD-10-PCS procedure 5A1945Z - count, first 24 hours |
| Baseline + procedure burden | 3 | procedure_icd10_5a1945z_f9e7fcf8fd_presence_24h | ICD-10-PCS procedure 5A1945Z - presence indicator, first 24 hours |
| Baseline + procedure burden | 3 | procedure_count_total_24h | Total procedure count, first 24 hours |
| Baseline + procedure burden | 3 | procedure_icd10_0bh17ez_3d5ba254ee_count_24h | ICD-10-PCS procedure 0BH17EZ - count, first 24 hours |
| Baseline + procedure burden | 3 | procedure_icd10_0bh17ez_3d5ba254ee_presence_24h | ICD-10-PCS procedure 0BH17EZ - presence indicator, first 24 hours |
| Baseline + procedure burden | 3 | procedure_icd9_3893_49b890a736_count_24h | ICD-9 procedure 3893 - count, first 24 hours |
| Baseline + procedure burden | 3 | procedure_icd9_3893_49b890a736_presence_24h | ICD-9 procedure 3893 - presence indicator, first 24 hours |
| Baseline + procedure burden | 3 | any_procedure_24h | Any procedure recorded, first 24 hours |
| Baseline + procedure burden | 3 | procedure_icd9_9604_93039b475e_count_24h | ICD-9 procedure 9604 - count, first 24 hours |
| Baseline + procedure burden | 3 | procedure_icd9_9604_93039b475e_presence_24h | ICD-9 procedure 9604 - presence indicator, first 24 hours |
| Baseline + procedure burden | 3 | procedure_icd10_02hv33z_8e9f454673_count_24h | ICD-10-PCS procedure 02HV33Z - count, first 24 hours |
| Baseline + procedure burden | 3 | procedure_icd10_02hv33z_8e9f454673_presence_24h | ICD-10-PCS procedure 02HV33Z - presence indicator, first 24 hours |
| Baseline + procedure burden | 3 | procedure_icd9_9671_5bce41582d_count_24h | ICD-9 procedure 9671 - count, first 24 hours |
| Baseline + procedure burden | 3 | procedure_icd9_9671_5bce41582d_presence_24h | ICD-9 procedure 9671 - presence indicator, first 24 hours |
| Baseline + procedure burden | 4 | procedure_icd10_5a1955z_6a2b52611f_count_24h | ICD-10-PCS procedure 5A1955Z - count, first 24 hours |
| Baseline + procedure burden | 4 | procedure_icd10_5a1955z_6a2b52611f_presence_24h | ICD-10-PCS procedure 5A1955Z - presence indicator, first 24 hours |
| Baseline + procedure burden | 4 | procedure_icd9_9672_93b639be5d_count_24h | ICD-9 procedure 9672 - count, first 24 hours |
| Baseline + procedure burden | 4 | procedure_icd9_9672_93b639be5d_presence_24h | ICD-9 procedure 9672 - presence indicator, first 24 hours |
| Baseline + procedure burden | 4 | unique_procedure_count_24h | Unique procedure count, first 24 hours |
| Baseline + procedure burden | 4 | procedure_count_total_24h | Total procedure count, first 24 hours |
| Baseline + procedure burden | 4 | procedure_icd10_5a1945z_f9e7fcf8fd_count_24h | ICD-10-PCS procedure 5A1945Z - count, first 24 hours |
| Baseline + procedure burden | 4 | procedure_icd10_5a1945z_f9e7fcf8fd_presence_24h | ICD-10-PCS procedure 5A1945Z - presence indicator, first 24 hours |
| Baseline + procedure burden | 4 | procedure_icd9_3893_49b890a736_count_24h | ICD-9 procedure 3893 - count, first 24 hours |
| Baseline + procedure burden | 4 | procedure_icd9_3893_49b890a736_presence_24h | ICD-9 procedure 3893 - presence indicator, first 24 hours |
| Baseline + procedure burden | 4 | procedure_icd10_3e0g76z_fa1c50513e_count_24h | ICD-10-PCS procedure 3E0G76Z - count, first 24 hours |
| Baseline + procedure burden | 4 | procedure_icd10_3e0g76z_fa1c50513e_presence_24h | ICD-10-PCS procedure 3E0G76Z - presence indicator, first 24 hours |

Continued on next page

eTable 34. MIMIC-IV standalone features selected per fold. (continued)

| Feature matrix | Fold | Feature | Concept name |
|---|---|---|---|
| Baseline + procedure burden | 4 | procedure_icd10_02hv33z_8e9f454673_count_24h | ICD-10-PCS procedure 02HV33Z - count, first 24 hours |
| Baseline + procedure burden | 4 | procedure_icd10_02hv33z_8e9f454673_presence_24h | ICD-10-PCS procedure 02HV33Z - presence indicator, first 24 hours |
| Baseline + procedure burden | 4 | procedure_icd10_0bh17ez_3d5ba254ee_count_24h | ICD-10-PCS procedure 0BH17EZ - count, first 24 hours |
| Baseline + procedure burden | 4 | procedure_icd10_0bh17ez_3d5ba254ee_presence_24h | ICD-10-PCS procedure 0BH17EZ - presence indicator, first 24 hours |
| Baseline + procedure burden | 4 | procedure_icd9_9671_5bce41582d_count_24h | ICD-9 procedure 9671 - count, first 24 hours |
| Baseline + procedure burden | 4 | procedure_icd9_9671_5bce41582d_presence_24h | ICD-9 procedure 9671 - presence indicator, first 24 hours |
| Baseline + procedure burden | 4 | any_procedure_24h | Any procedure recorded, first 24 hours |
| Baseline + procedure burden | 4 | procedure_icd9_3891_71dd24c736_count_24h | ICD-9 procedure 3891 - count, first 24 hours |
| Baseline + procedure burden | 4 | procedure_icd9_3891_71dd24c736_presence_24h | ICD-9 procedure 3891 - presence indicator, first 24 hours |
| Baseline + procedure burden | 5 | procedure_icd10_5a1955z_6a2b52611f_count_24h | ICD-10-PCS procedure 5A1955Z - count, first 24 hours |
| Baseline + procedure burden | 5 | procedure_icd10_5a1955z_6a2b52611f_presence_24h | ICD-10-PCS procedure 5A1955Z - presence indicator, first 24 hours |
| Baseline + procedure burden | 5 | procedure_icd9_9672_93b639be5d_count_24h | ICD-9 procedure 9672 - count, first 24 hours |
| Baseline + procedure burden | 5 | procedure_icd9_9672_93b639be5d_presence_24h | ICD-9 procedure 9672 - presence indicator, first 24 hours |
| Baseline + procedure burden | 5 | procedure_icd9_3893_49b890a736_count_24h | ICD-9 procedure 3893 - count, first 24 hours |
| Baseline + procedure burden | 5 | procedure_icd9_3893_49b890a736_presence_24h | ICD-9 procedure 3893 - presence indicator, first 24 hours |
| Baseline + procedure burden | 5 | unique_procedure_count_24h | Unique procedure count, first 24 hours |
| Baseline + procedure burden | 5 | procedure_count_total_24h | Total procedure count, first 24 hours |
| Baseline + procedure burden | 5 | procedure_icd10_3e0g76z_fa1c50513e_count_24h | ICD-10-PCS procedure 3E0G76Z - count, first 24 hours |
| Baseline + procedure burden | 5 | procedure_icd10_3e0g76z_fa1c50513e_presence_24h | ICD-10-PCS procedure 3E0G76Z - presence indicator, first 24 hours |
| Baseline + procedure burden | 5 | procedure_icd10_5a1945z_f9e7fcf8fd_count_24h | ICD-10-PCS procedure 5A1945Z - count, first 24 hours |
| Baseline + procedure burden | 5 | procedure_icd10_5a1945z_f9e7fcf8fd_presence_24h | ICD-10-PCS procedure 5A1945Z - presence indicator, first 24 hours |
| Baseline + procedure burden | 5 | any_procedure_24h | Any procedure recorded, first 24 hours |
| Baseline + procedure burden | 5 | procedure_icd10_0bh17ez_3d5ba254ee_count_24h | ICD-10-PCS procedure 0BH17EZ - count, first 24 hours |
| Baseline + procedure burden | 5 | procedure_icd10_0bh17ez_3d5ba254ee_presence_24h | ICD-10-PCS procedure 0BH17EZ - presence indicator, first 24 hours |
| Baseline + procedure burden | 5 | procedure_icd9_9671_5bce41582d_count_24h | ICD-9 procedure 9671 - count, first 24 hours |
| Baseline + procedure burden | 5 | procedure_icd9_9671_5bce41582d_presence_24h | ICD-9 procedure 9671 - presence indicator, first 24 hours |
| Baseline + procedure burden | 5 | procedure_icd10_02hv33z_8e9f454673_count_24h | ICD-10-PCS procedure 02HV33Z - count, first 24 hours |
| Baseline + procedure burden | 5 | procedure_icd10_02hv33z_8e9f454673_presence_24h | ICD-10-PCS procedure 02HV33Z - presence indicator, first 24 hours |
| Baseline + procedure burden | 5 | procedure_icd9_3891_71dd24c736_count_24h | ICD-9 procedure 3891 - count, first 24 hours |
| Baseline + procedure burden | 5 | procedure_icd9_3891_71dd24c736_presence_24h | ICD-9 procedure 3891 - presence indicator, first 24 hours |

*Notes:* MIMIC-IV baseline predictors are raw model-input variables repeated by fold. Categorical variables are encoded inside the MIMIC-IV model pipelines; broad labels are shown here for readability and do not imply that only one encoded category variable was used.

Cohort demographic characteristics

| Dataset | Demographic | Demographic category | No. of patients/visits |
|---|---|---|---|
| CHoRUS | Age | Under 18 years | 3,203 |
| | | 18–59 years | 9,662 |
| | | 60 years or older | 9,228 |
| | | Unknown | 5 |
| | Sex | Male | 3,217 |
| | | Female | 2,675 |
| | Race | Black | 736 |
| | | White | 4,177 |
| | | Asian | 148 |
| | | Other or Unknown | 831 |
| | Ethnicity | Hispanic | 447 |
| | | Non-Hispanic | 5354 |
| | | Other or Unknown | 91 |
| MIMIC-IV | Age | Under 18 years | 0 |
| | | 18–59 years | 11,130 |
| | | 60 years or older | 11,870 |
| | | Unknown | 0 |
| | Sex | Male | 4,693 |
| | | Female | 5,313 |
| | | Other or unknown | 0 |
| | Race | Black | 1,241 |
| | | White | 6,631 |
| | | Asian | 465 |
| | | Other or unknown | 1,669 |
| | Ethnicity | Hispanic | 521 |
| | | Non-Hispanic | 8,811 |
| | | Other or unknown | 674 |

*Note.* Sex, race, and ethnicity are reported as numbers of unique patients. Age is reported at the visit level because a patient's age may differ across encounters. The visit type is reported at the visit level. The age and visit categories therefore sum to the total number of eligible visits, whereas the sex, race, and ethnicity categories sum to the total number of unique patients.
